# De novo Powered Air-Purifying Respirator Design and Fabrication for Pandemic Response

**DOI:** 10.1101/2021.03.25.21252076

**Authors:** Akshay Kothakonda, Lyla Atta, Deborah Plana, Ferrous Ward, Chris Davis, Avilash Cramer, Robert Moran, Jacob Freake, Enze Tian, Ofer Mazor, Pavel Gorelik, Christopher Van, Christopher Hansen, Helen Yang, Michael S. Sinha, Ju Li, Sherry H. Yu, Nicole R. LeBoeuf, Peter K. Sorger

## Abstract

The rapid spread of COVID-19 and disruption of normal supply chains resulted in severe shortages of personal protective equipment (PPE), particularly devices with few suppliers such as powered air-purifying respirators (PAPRs). A scarcity of information describing design and performance criteria represents a substantial barrier to new approaches to address these shortages. We sought to apply open-source product development to PAPRs to enable alternative sources of supply and further innovation. We describe the design, prototyping, validation, and user testing of locally manufactured, modular, PAPR components, including filter cartridges and blower units, developed by the Greater Boston Pandemic Fabrication Team (PanFab). Two designs, one with a fully custom-made filter and blower unit housing, and the other with commercially available variants (the “Custom” and “Commercial” designs respectively) were developed. Engineering performance of the prototypes was measured and safety validated using NIOSH-equivalent tests on apparatus available under pandemic conditions, at university laboratories. Feedback on designs was obtained from four individuals, including two clinicians working in an ambulatory clinical setting and two research technical staff for whom PAPR use is a standard part of occupational PPE. Respondents rated the PanFab Custom PAPR a 4 to 5 on a 5 Likert-scale 1) as compared to current PPE options, 2) for the sense of security with use in a clinical setting, and 3) for comfort. The three other versions of the designs (with a commercial blower unit, filter, or both) performed favorably, with survey responses consisting of scores ranging from 3-5. Engineering testing and clinical feedback demonstrate that the PanFab designs represents favorable alternative PAPRs in terms of user comfort, mobility, and sense of security. A nonrestrictive license promotes innovation in respiratory protection for current and future medical emergencies.

## INTRODUCTION

The rapid and global spread of COVID-19 led to a large increase in demand for personal protective equipment (PPE) for healthcare workers as well as significant disruption of supply chain and distribution networks for these products. As a consequence, the availability of high-quality respiratory protection has been problematic, causing healthcare institutions to reuse normally disposable filtering facepiece respirators (FFRs; N95-type masks) and turn to non-traditional devices as substititues^1–5^. Non-traditional supply chains commonly involve community-based collaborations among independent engineers, scientists, hobbyists, and volunteers in partnership with healthcare and academic institutions^6–10^. While multiple non-traditional designs for simple PPE products such as face shields have emerged, and multiple commercial and non-traditional technologies and methods have been developed to decontaminate and reuse N95-type masks ^11,12^, few alternative sources of supply exist for more complex products. This is particularly true of powered air-purifying respirators (PAPRs), which can not only be worn by individuals unable to fit N95-style masks, but are also more comfortable in many settings and may provide a higher level of respiratory protection. Ongoing efforts to increase the supply of PAPRs have largely involved large manufacturing companies who have been provided with government support (e.g. the 3M Ford Limited-Use Public Health Emergency PAPR)^13,14^.

PAPRs typically cover the entire head with a loose-fitting headpiece or hood and provide a continuous supply of filtered air to a user from a blower worn on a belt or backpack. Like N95 masks, PAPRs are used in both healthcare and industrial settings but under non-pandemic conditions, healthcare use of PAPRs is typically limited to situations in which a healthcare worker (HCW) is unable to wear a disposable N95 mask yet must care for a patient with a suspected or confirmed airborne infection, such as tuberculosis^15^. Common reasons for being unable to wear an N95 mask are the presence of facial hair and poor mask fit for individuals with small or narrow faces; the latter is often revealed by qualitative fit tests routinely performed on HCWs.^16^ It is estimated that ∼10% of HCWs fail fit testing,^17^ and for these individuals PAPRs are the best, and in some instances, the only alternative. N95 masks are often reused in pandemic conditions due to supply shortages, leading to concerns about further loss of fit after multiple don-doff cycles ^18^. In addition, HCWs report that PAPRs are more comfortable than masks in situations in which continuous respiratory protection is required for many hours, especially for those who have respiratory symptoms under normal circumstances or who work in hot conditions. It has also been observed that many healthcare workers who must wear N95-style masks day after day (i.e. during a sustained pandemic) experience painful abrasions^19,20^. Attempts to mitigate this discomfort by using creams, tapes or loosening the straps that hold masks against a user’s face degrade respiratory protection^21^. A recent systematic review of the available literature has identified a need to rigorously investigate all of these issues.^15^

Commercial PAPRs certified by NIOSH offer higher filtration efficiency as compared to N95 masks (99.97% vs. 95%)^15,22^ and have assigned protection factors between 2.5 and 100 fold, substantially higher than those of N95-style masks^23^. PAPRs are also better suited to periods of very high demand: whereas N95 masks are designed for one-time use, commercially available PAPRs are designed to be sterilized for reuse^24^. Feedback from healthcare institutions following the 2009 H1N1 pandemic confirmed that HCWs with underlying respiratory conditions and individuals who must perform physically demanding tasks prefer PAPRs over masks; PAPRs are also broadly perceived to provide superior respiratory protection as compared to masks, which increases HCW confidence in high-risk settings^17^. However, the acquisition costs of commercial PAPRs are ∼1,000-fold higher than N95 masks. PAPR filters, which must be changed regularly are also relatively expensive^17^. PAPRs are therefore in short supply in most US healthcare institutions, which typically seek the lowest cost approach to respiratory protection for HCWs. Despite calls by multiple Federal panels over a period of two decades to promote innovation in respiratory protection^10^ there has been little concrete response. This is a setting in which open-source product development (OSPD)^25^ has the potential to make a substantial contribution.

PAPRs are composed of three primary functional components: the filter cartridge, the blower unit, and the facepiece, which is connected to the blower via a flexible hose; additional components, such as a low flow rate alarm, enhance user safety and usability (**Figure 1**). The blower unit and its associated power and control systems are enclosed inside an air-tight housing. This housing couples to filter cartridges and a hose. The blower unit pulls room air through one or multiple high-efficiency particulate air/ high-efficiency particulate absorbing (HEPA) filter cartridges, thereby removing aerosols and small particles. The blower pushes the filtered air into the facepiece (also known as a hood) through the hose, where it is either breathed in by the user or (in case of a loose-fitting facepiece) escapes through gaps between the facepiece material and the user’s body. The presence of positive pressure in the facepiece ensures that unfiltered outside air does not enter the facepiece and is not inhaled by the user.

**Figure 1:**
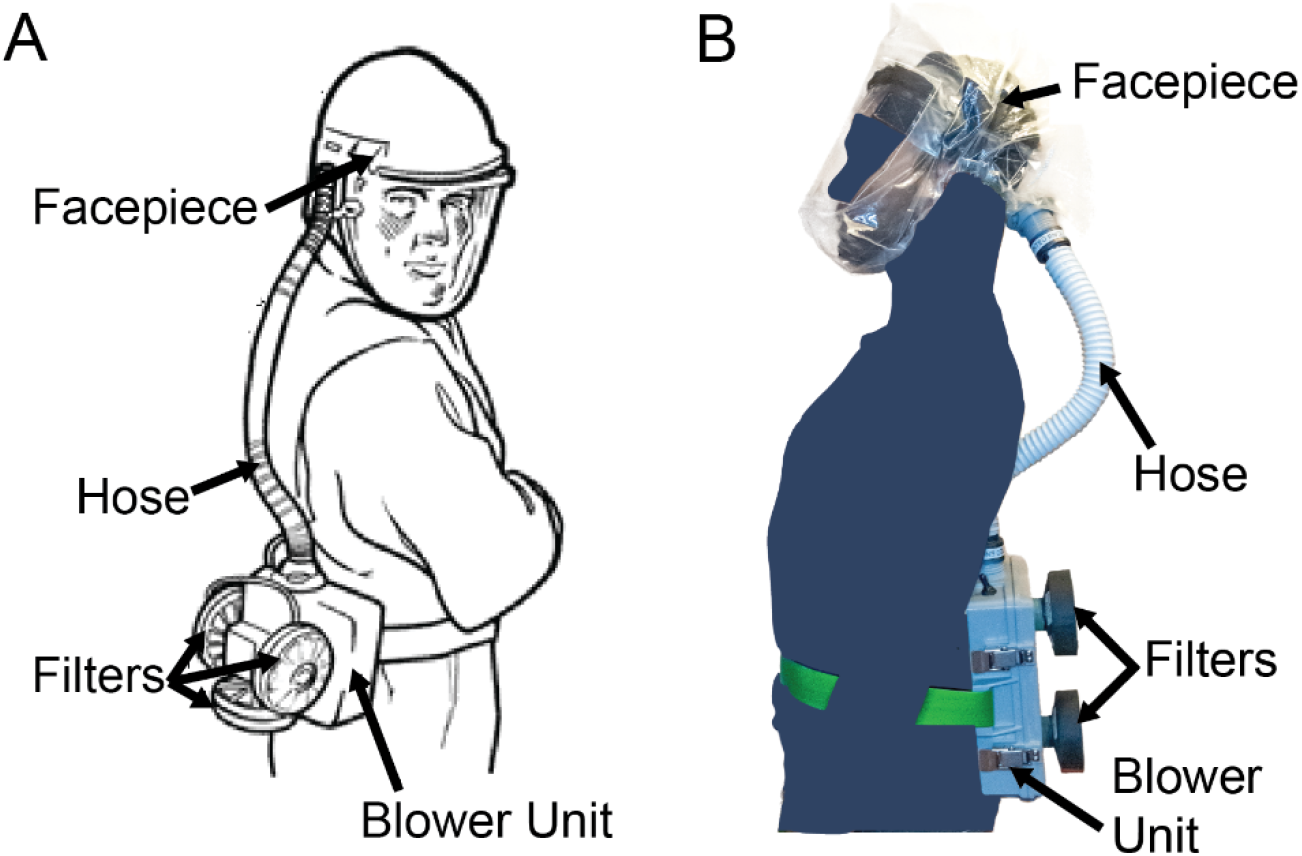
PAPR components. **A)** Diagram of PAPR components, adapted from OSHA.gov^26^. **B)** PanFab PAPR described in this work.

The current shortage of PAPRs likely reflects the complexity of these devices, which have multiple components, each requiring significant expertise to design, engineer and test. Resources detailing the design criteria for PAPRs used in healthcare settings are scarce because most designs are proprietary, making it challenging for new or local manufacturers to help address shortages. Additionally, the regulatory approval process for PAPRs via the U.S. Food and Drug Administration (FDA) and National Institutes for Occupational Safety and Health (NIOSH) is significantly more complex than for simpler devices such as face shields. In the U.S., PAPRs are regulated by the Occupational Safety and Health Agency (OSHA) under the Respiratory Protection standard (29 CFR 1910.134). This requires that PAPRs be approved by NIOSH but does not require 510(k) premarket notification or clearance by the FDA^27,28^. One challenge is that NIOSH testing standards are highly prescriptive, typically describing the precise instruments to be used in a test. The physical or engineering principles underlying these tests are not always obvious. The prescriptive approach may be appropriate under normal circumstances when it is important to maintain quality standards in the face of cost pressure, but is problematic in emergency conditions in which the approved testing apparatus is in short supply. In the current work we therefore rely on “NIOSH-equivalent” testing to assess performance.

We sought to create public domain PAPR designs with non-restrictive licensing that could help to address current shortages in respiratory protection. We also sought to harness the power of open source product development to address a broader problem in supply chains disruption caused by disease pandemics or other healthcare emergencies, as well as long-standing problems in the supply of medical products in resource-limited environments, developing nations for example. After consulting with clinicians and infection control specialists at Harvard Medical School-affiliated hospitals, we focused our efforts on designing filter cartridges and blower units (consisting of a housing, blower, battery, flow control system, and flow control alarm), the two PAPR components most commonly in shortage. NIOSH standard testing procedures (STP)^29^, which specify the testing requirements needed for NIOSH approval of PAPRs, provided performance specifications for the filter cartridge and blower unit components (**Supplementary Material 1**).

In this paper, we describe the design, validation, and user testing of modular PAPR components- the filter cartridges and the blower units, developed by the Greater Boston Pandemic Fabrication Team (PanFab) ^30^. These components are intended to provide alternatives to standard commercially available PAPR components that can be locally manufactured in times of severe PAPR shortage. For both the filter cartridge and the blower unit components, we describe a “PanFab Custom Design” and a “PanFab Commercial Design” to accommodate different scenarios with respect to shortages of materials. The custom design has less reliance on commercial products and supply chains and can be fabricated in large part using additive manufacturing (3D-printing) methods for low volume production or injection molding for high volume needs^31^. The commercial design relies on commercially available parts made for other products and requires fewer custom fabrication steps, facilitating rapid introduction of new, locally fabricated units. The PanFab PAPR components are modular and interchangeable: any combination of components can be used together and with traditional PAPR components from leading suppliers. For example, the PanFab Custom Filter can be used with the PanFab Commercial Blower Unit and vice-versa. The PanFab PAPR components are also compatible with the widely used ILC Dover Sentinel XL PAPR facepiece^32^ and filters. The blower can also be adapted to other commercially available PAPR facepieces by fabricating a slightly modified hose-to-facepiece connector. Under the provisions of a Creative Commons Attribution-Share Alike 4.0 International Public License, other entities are free to use components of the PanFab PAPR, by itself, in their own designs, or to further innovate.

Initial prototype testing was conducted at academic laboratories using equipment and supplies that were available during the COVID-19 pandemic. User feedback on the functionality and comfort of the design was then obtained at a major US academic medical center from four participants: two healthcare providers and two research technicians who used PAPRs regularly as part of standard PPE prior to the pandemic. As mentioned above, performance testing was conducted using alternative procedures and with fewer samples than is required for NIOSH certification. An additional limitation is that PAPR certification, like certification of most medical products, requires a manufacturing process controlled by a quality management system (e.g. one to ISO 9001 standards). Achieving this is only possible in a commercial setting, and we are therefore collaborating with an industrial partner to create a design amenable to large-scale manufacturing and certified to NIOSH standards. Other users of the PanFab PAPR must perform their own testing and confirm that fabricated products meet the requirements of FDA Emergency Use Authorizations and similar regulatory guidance. We return to this issue in the discussion.

## METHODS

### Prototype development

NIOSH requirements (NIOSH STP CVB-APR-STP-0081) specify that PAPRs have a minimum filtration efficiency of 99.97% for NaCl aerosols (this corresponds to the N100 class of PAPRs). To reduce the power required to drive air through filters, they should also have as little pressure drop as possible at the minimum required flow rate of 170 liters per minute (lpm; NIOSH STP RCT-APR-STP-0012). For the PanFab Commercial PAPR, we selected a commercially available HEPA filter that is used in consumer vacuum cleaners and is widely available; we speculated that supply of these filters is unlikely to be significantly affected by disruption of medical device supply chains caused by the COVID-19 pandemic. For the PanFab Custom PAPR, a custom-designed filter cartridge was designed to be lighter in weight and have a lower form factor.

Design of the blower units focused on meeting the required flow rate of at ≥170 lpm and overcoming pressure drops caused by the filters and tubing in the air flow path at this flow rate. In addition, blower units needed to be operational for at least 1 hour, comfortable to wear, sterilizable, airtight, and relatively silent (NIOSH STP RCT-APR-STP-0030). For a full list of design requirements identified and NIOSH testing requirements, refer to **Tables 1** and **2** respectively. **Supplementary Material 2** provides a full discussion of design methodology and resulting prototype components.

**Table 1:**
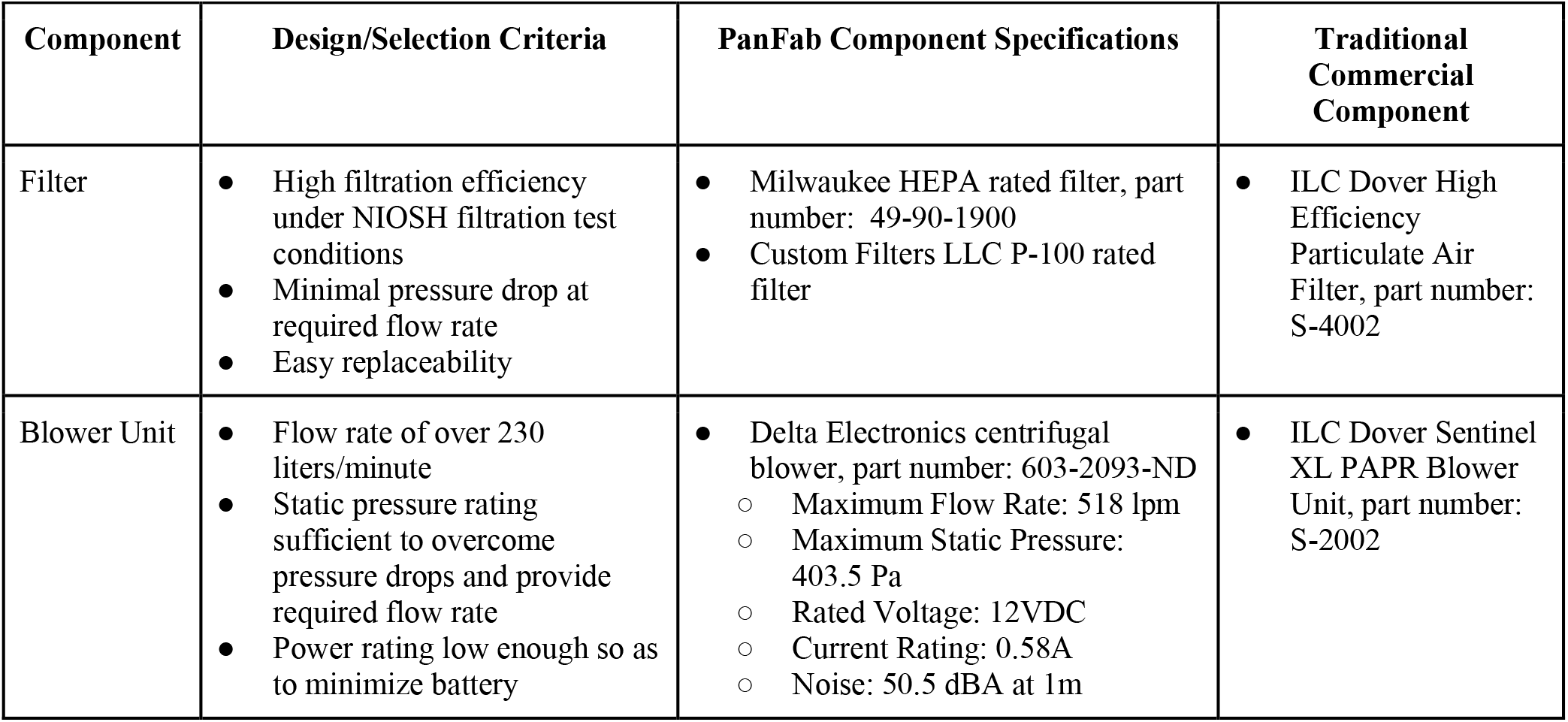
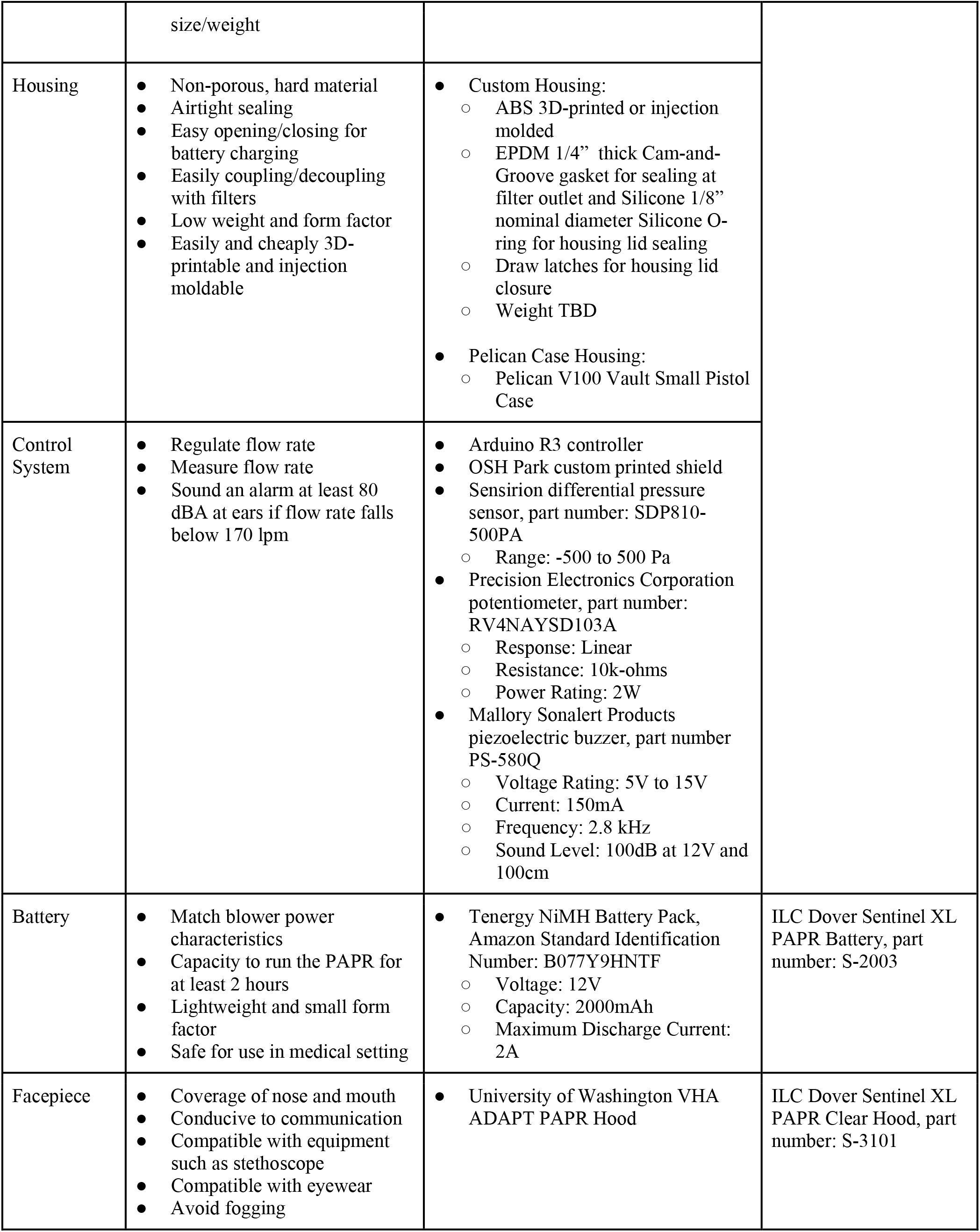
PanFab PAPR design components, selection criteria, specifications, and commercial components.

**Table 2:** PanFab PAPR component validation test type, regulatory guidance, and alternative test used due to shortages experienced during COVID-19. Full STP available as **Supplementary Material 1**.

| Test Type | Relevant NIOSH STP | Result of NIOSH-alternative test |
| --- | --- | --- |
| Filtration efficiency | Procedure No. CVB-APR-STP-0081 Determination of Particulate Filter Efficiency Level Against Solid Particulates (PAPR 100-N) | Milwaukee Filters:<br>99.99%, 100% at 300 nm and 230 lpm |
|  |  | Custom Filter:<br>99.99% at 300 nm and 230 lpm |
|  | Procedure No. TEB-APR-STP-0001 Determination of Particulate Filter Penetration (PAPR) Test | Milwaukee Filters:<br>99.18% and 99.58% at 170 lpm |
|  |  | Custom Filter:<br>99.98% at 170 lpm |
| Flow rate | Procedure No. RCT-APR-STP-0012 Determination of Air Flow For Powered Air-Purifying Respirators | 240 liter/min at 70% blower duty cycle |
| Qualitative fit | Procedure No. RCT-APR-STP-0067<br>Particulate Respirator Qualitative Fit Test Utilizing Saccharin or Bitrex Solutions | Pass, n=1 |
| Noise level | Procedure No. RCT-APR-STP-0030: Determination of Noise Level Test, Power Air-Purifying Respirator With Hoods Hoods or Helmets | 58.1 to 59.2 dBA at full battery charge and maximum blower speed |
| Low flow rate alarm | Procedure No. CVB-APR-STP-0085 Determination of Low Flow Warning Device Sound Level<br>Procedure No. CVB-APR-STP-0088 Determination of Low Flow Warning Device Activation | Between 82.95 and 84.7 dBA at 230 lpm flow rate |
| Audibility test | Procedure No. CVB-APR-STP-0089 Determination of Communication Performance Test For Speech Conveyance And Intelligibility | Pass, n=1 |

### Prototype testing

NIOSH has developed several STPs for testing the safety and functionality of PAPR components^29^. Third-party testing in commercial laboratories offers testing of PAPRs to these STPs and represents an established route for demonstrating compliance with NIOSH standards. These tests are not only expensive, but high demand for testing accompanying the COVID-19 pandemic also made many of these test options either unavailable or considerably delayed. As an alternative, we devised an equivalent testing apparatus to those specified in the corresponding NIOSH STPs. The tests were carried out across several university laboratories on the filters, blowers, power systems, control and warning systems, and the seals between components. The use of these “NIOSH equivalent” tests allowed us to make progress on PAPR design and testing but does not obviate the need for testing to NIOSH STPs prior to use in clinical settings.

For prototype testing, we used a loose-fitting facepiece known as the VHA ADAPT PAPR Hood, developed by the Center for Limb Loss and MoBility (CLiMB) at the University of Washington^33^. This was used in place of standard commercial PAPR facepieces such as the ILC Dover Sentinel XL PAPR facepiece, for which supply was limited. However, the PanFab PAPR is compatible with ILC Dover facepieces should they become readily available.

#### Filter testing

NIOSH performs two distinct filtration efficiency tests; a full loading test and an instantaneous (abbreviated) test, the latter of which estimates the lowest filtration efficiency expected at the start of a filter’s service life. NIOSH performs the loading filtration efficiency test of N100 class of PAPR filters using 75 nm NaCl aerosols (NIOSH STP CVB-APR-STP-0081). To test the filtration efficiency for the filters we selected, we modified a previously described university-based apparatus^34^ that was originally used to assess the filtration efficiency of N95-type FFRs (**Figure 2**). Due to the unavailability of NaCl aerosol generators, we used KCl instead. A Handheld Particle Counter (TSI 9306-V2 AeroTrak, TEquipment, Long Branch, NJ) measured filtration efficiency, a Collison Nebulizer (MRE 6-Jet, BGI Inc., Waltham, MA) generated aqueous particle streams, and a differential pressure gauge (purchased from McMaster-Carr, part number 4125K21) measured the pressure drop across the filter.

**Figure 2:**
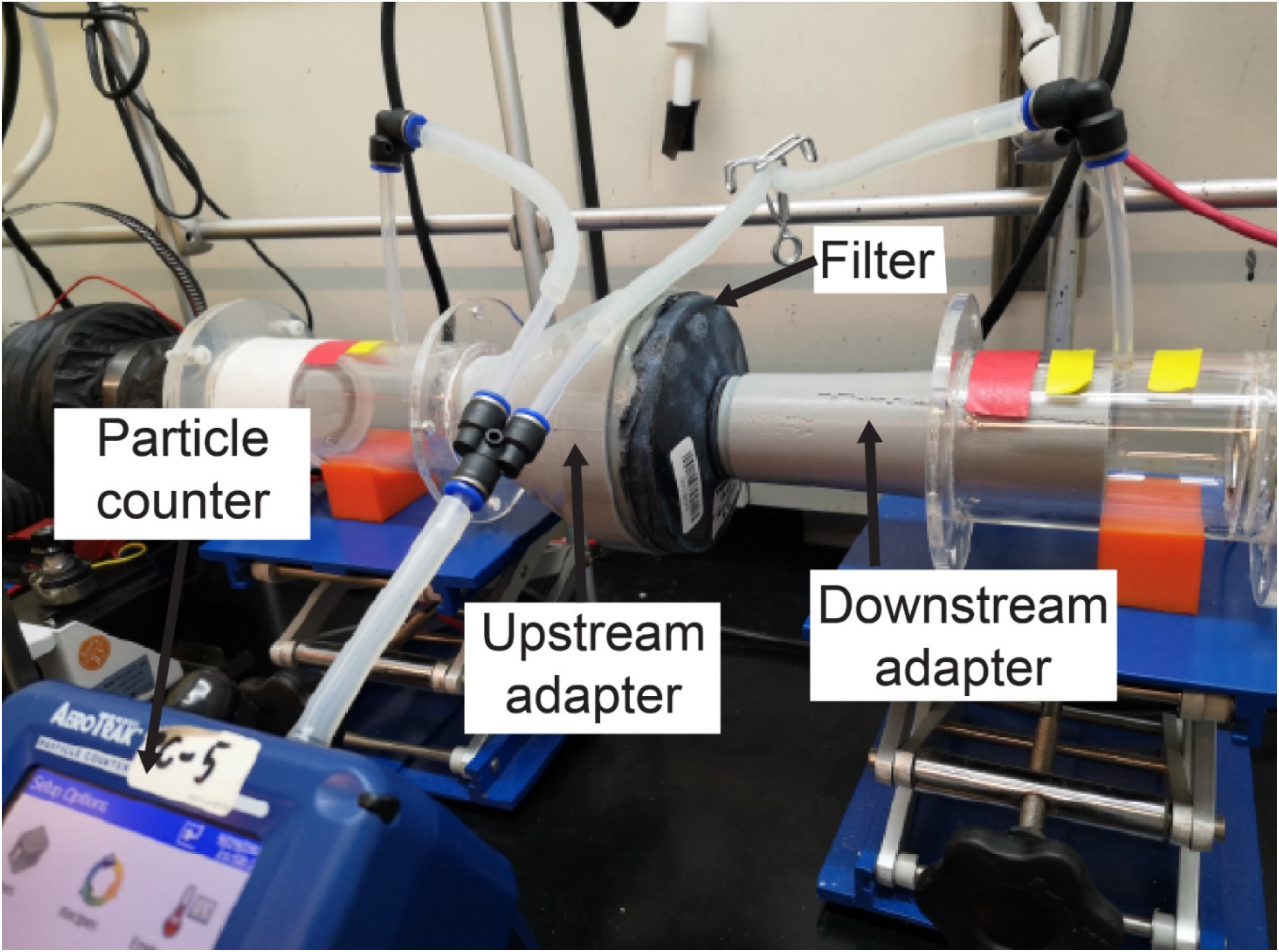
Loading filtration test setup, with filter cartridge in line with KCl-containing air stream. Other components of the apparatus has been previously described^34^.

Our apparatus had a lower measurable limit of 300 nm particle size, which we expect to result in a more conservative estimate of filtration efficiency than particle sizes specified in NIOSH STPs.^35^ Filter cartridges were placed within the apparatus in line with the flow of the KCl-containing particle stream. Special 3D-printed adapters, sealed to the cartridges, were tightly coupled to upstream and downstream air ducts, ensuring no leakage. The KCl concentration was measured upstream and downstream of the cartridges and used to estimate filtration efficiency.

The US Code of Federal Regulations (42 CFR § 84.175) specifies that PAPR performance is tested with a dioctyl phthalate aerosol (NIOSH STP TEB-APR-STP-0001; note that “DOP” is used as an abbreviation both for “dispersed oil particulate” and “dioctyl phthalate”). This is an instantaneous filtration test, in which, a filter cartridge is challenged with a dioctyl phthalate-containing aerosol for approximately 10 seconds. In lieu of dioctyl phthalate, which is a suspected carcinogen^36^, we used an oil aerosol containing polyalphaolefin (PAO), which is representative of the most widely used class of synthetic lubricants and an accepted substitute for dioctyl phthalate.^37^ Filtration efficiency testing was then performed as described above.

#### Air-flow testing

NIOSH requires a minimum flow rate of 170 lpm for PAPRs that have loose fitting facepieces, ^38^ based on a test described in NIOSH STP RCT-APR-STP-0012. To measure flow rate, the STP connects a vacuum chamber, evacuated with a vacuum pump, to a running PAPR blower unit, and uses a dry test meter to measure flow rate. In the absence of this setup, we used an impeller type anemometer (Vernier Software and Technology, Beaverton, OR) connected to the air inlet at the facepiece, with the neck opening sealed with duct tape (**Figure 3A**). An adapter was 3D-printed to couple the facepiece inlet to the anemometer flow area, such that all the flow into the facepiece passed through the cross-sectional area of the anemometer inlet. Flow rate was calculated by multiplying the air velocity recorded on the anemometer with the cross-sectional area. Additionally, we used a Vernier Gas Pressure Sensor placed inside the facepiece to measure the positive pressure created in the facepiece. While not as precise as the procedure described in NIOSH STP RCT-APR-STP-0012, we expect the test we performed to provide a close approximation to the flow rate, using equipment available to us in a university laboratory.

**Figure 3:**
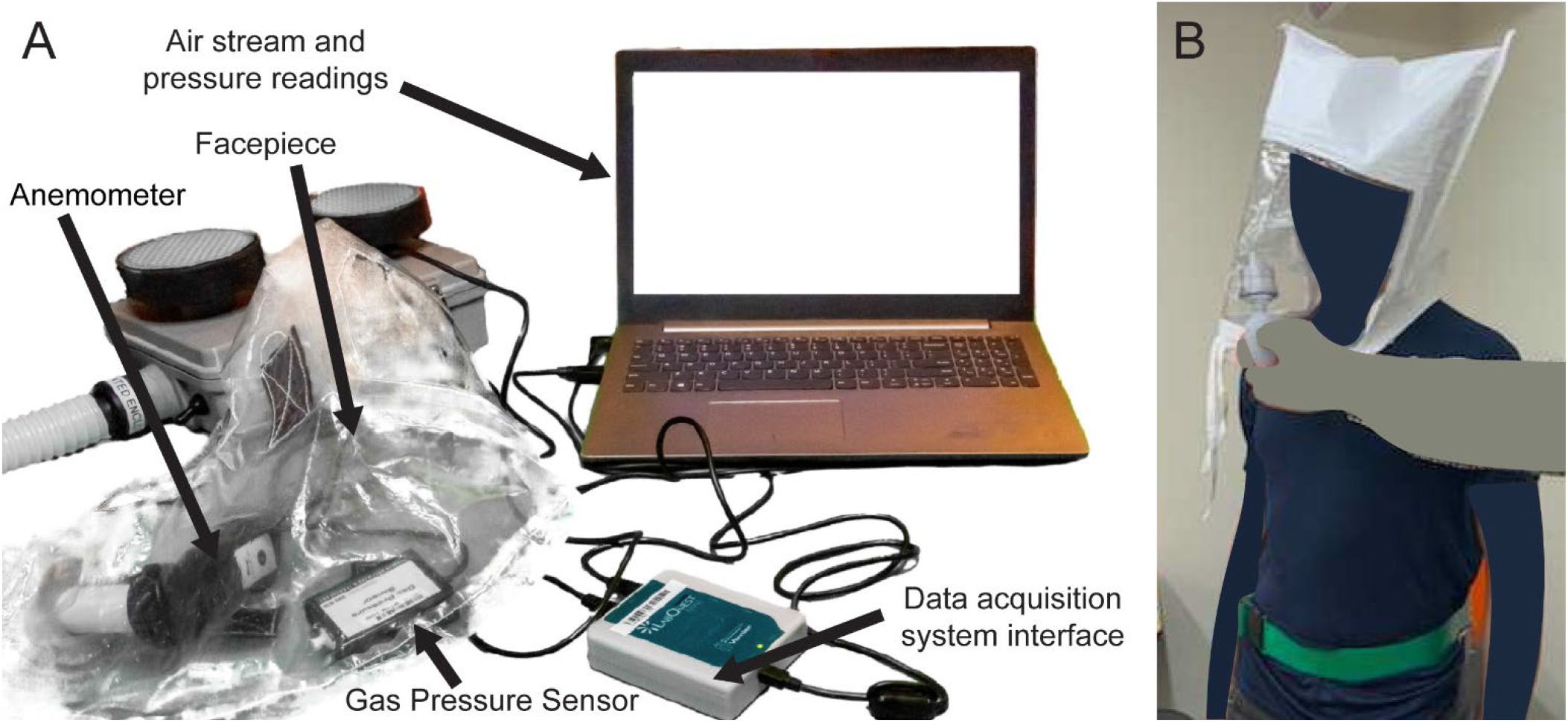
**A)** Test setup to measure flow rate and the positive pressure inside the facepiece, using Vernier Anemometer and Gas Pressure Sensor. PAPR facepiece contains anemometer and gas pressure sensor. Duck tape covers the neck opening of the facepiece for testing. **B)** Bitrex Fit Test setup.

#### Facepiece fit testing

To determine if unfiltered air can enter the facepiece, we conducted a Bitrex qualitative fit test as described in NIOSH STP RCT-APR-STP-0067^39^ on one test subject (**Figure 3B**). This test evaluates whether flow rate into the facepiece is sufficient to prevent unfiltered ambient air from reaching the user; the unfiltered ambient air contains an aqueous aerosol of denatonium benzoate (a bitter chemical) and subjects are asked if they can taste it during the test. Under the NIOSH Interim Final Rule for PAPR testing, the fit test is to be performed using corn oil, as per NIOSH STP CVB-APR-STP-0010. However, Bitrex fit testing is commonly used in hospital settings and allowed according to pre-pandemic NIOSH testing requirements.

#### Auditory, communication, and low flow rate alarm testing

NIOSH STP RCT-APR-STP-0030 requires the noise level at each ear, with the blower unit running at maximum flow, not exceed 80 A-weighted decibels (dBA). We used a Vernier SLM-BTA Sound Level Meter to measure sound level. We also tested ease of communication while wearing each PanFab PAPR following NIOSH STP CVB-APR-STP-0089 with one subject. The subject was tasked with speaking and listening to a set of words. Ease of communication was evaluated by counting the number of words correctly transcribed in each task, normalized by baseline performance without the PAPR.

NIOSH STP CVB-APR-STP-0085 requires that PAPRs have an alarm to alert users when air flow rate falls below the minimum level of 170 lpm. Auditory alarms are required to be louder than 80 dBA. We again used a Vernier SLM-BTA Sound Level Meter to measure the sound level of the low flow rate alarm built into the PanFab blower units while triggering the alarm by manually restricting the air flow at the facepiece inlet.

## RESULTS

PanFab Custom and Commercial Designs were developed for both filter cartridges and blower units **(Figure 4)**. For a full description of the filter and blower unit designs, refer to **Table 1** and **Supplementary Material 2**. A Milwaukee Tool (Brookfield, WI) HEPA-rated vacuum cleaner filter (part number 49-90-1900) was selected as the PanFab Commercial Filter Cartridge. A custom 3D-printed adapter converts the outlet of the Milwaukee filter to standard NATO 40-millimeter threaded connection, allowing it to be used with the PanFab and other commercial blower units. With slight modification of the adapters, other commercial HEPA-rated vacuum cleaner filters could be used as alternatives. A custom 3D-printed filter cover protects the filter fabric. The Custom variant of the PanFab filter cartridge was designed in collaboration with Custom Filters LLC (Aurora, IL) to have the necessary P-100 rating while remaining small and light. Two filters were used in each variant of the PAPR as opposed to one, so as to minimize the pressure drop for a given flow rate.

**Figure 4:**
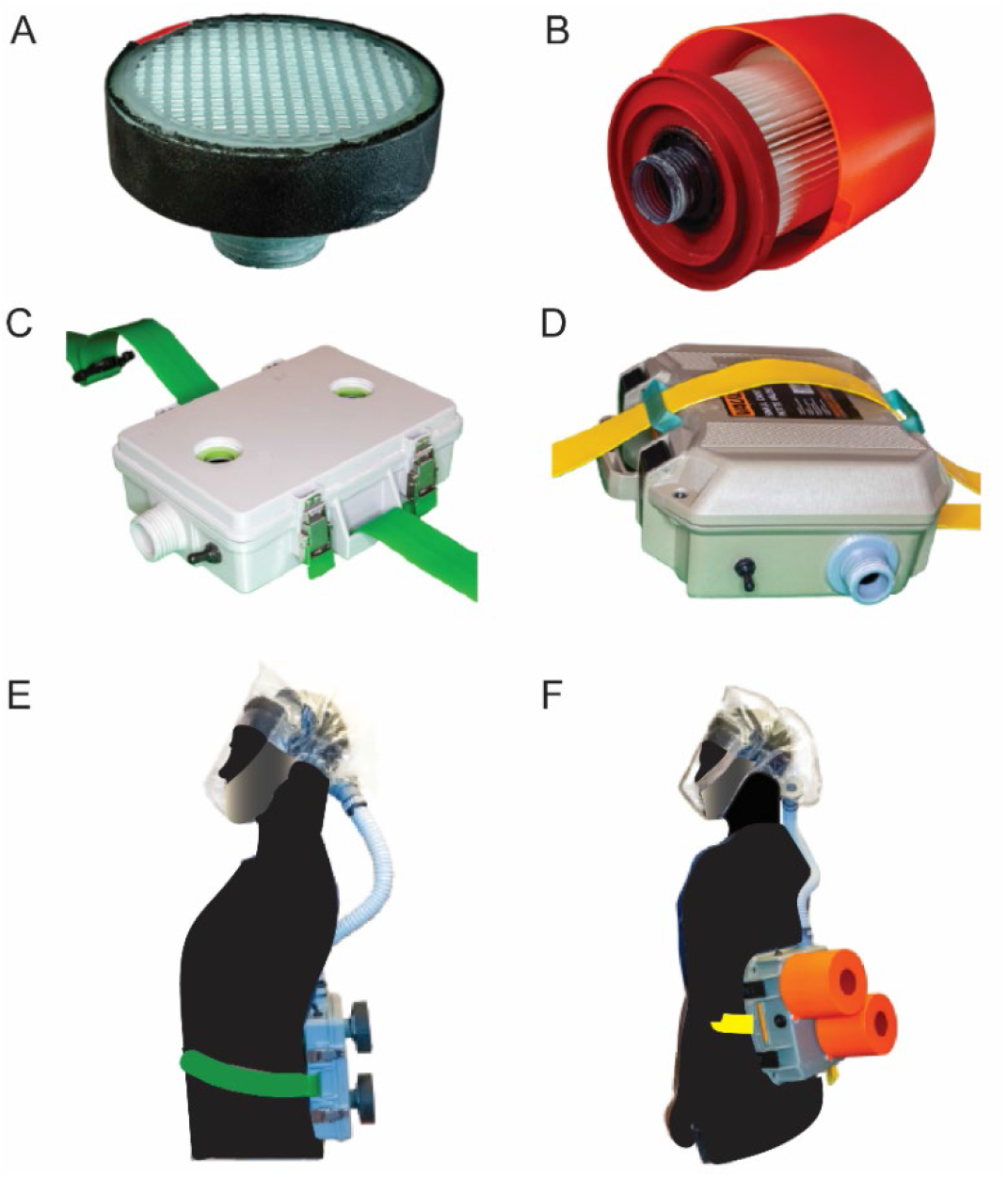
PanFab PAPR components. **A)** PanFab Custom Filter Cartridge. **B)** PanFab Commercial Filter Cartridge. **C)** PanFab Custom blower units. **D)** PanFab Commercial blower units. **E)** PanFab Custom Design (Custom Filter Cartridge plus Custom blower units). **F)** PanFab Commercial Design (Commercial Filter Cartridge plus Commercial blower units).

A centrifugal blower (Delta Electronics, Neihu, Taiwan, Part Number BFB1012HD-04D4L) generates a 230 lpm flow rate, higher than the required 170 lpm, and a 12-volt (V) NiMH battery pack (Tenergy, Fremont, CA, Amazon Standard Identification Number: B077Y9HNTF) was used to power it; this battery pack was sufficient for ∼4 hours of continuous use. The battery can be charged in various ways, including with solar power, as long as 12 Voltage Direct Current (VDC) < 1.8 amperes (Amp) can be supplied through an electrical connector compatible with that used in the battery. A wide variety of 12V NiMH and lithium ion battery packs are available and could be used as substitutes following performance testing,

Control circuitry was based on a standard Arduino R3 board (Arduino LLC, Boston, MA) with custom-fabricated shield (OSH Park, Portland, OR). Discrete components connected to the shield included a 10 kiloohm potentiometer (Precision Electronics Corporation, North York, ON, Canada, part number RV4NAYSD103A), differential pressure sensor (Sensirion AG, Staefa, Switzerland, part number SDP810-500PA), and piezoelectric buzzer (Mallory Sonalert Products Inc., Indianapolis, IN, part number PS-580Q). These components were used for control and alarm tasks such as regulating the air flow rate, measuring flow rate, and sounding the low-flow buzzer. All of the discrete components are readily substitutable with similar products made by multiple manufacturers. We established that the Sonalert buzzer generated a sound of at least 80 dBA (as per CVB-APR-STP-0085) when the flow rate fell below a NIOSH specified threshold (as per CVB-APR-STP-0088).

The housings that enclose the blower, battery, and control components were designed to be airtight when closed with filters and hose attached. In case of the Commercial Design, a Pelican V100 Vault Case (Pelican Products, Torrance, CA) was used with custom made “inserts” for connection to filters and hose via NATO 40mm connections. The Custom housing was designed to be small and lightweight, with integrated connections to the filters and hose. Finally, a hood coupler and a locking ring were designed to connect the hose to the UW facepiece, as well as to other facepieces that use a NATO 40mm threaded connection.

With their respective filters installed, the PanFab Custom and Commercial PAPRs weigh 1.87 kg and 3.36 kg, respectively. Both PanFab PAPRs are worn on the waist using a Skil-Care (Yonkers, NY) PathoShield Gait Belt. This 50 mm wide web belt is heat-sealed (rather than stitched), has a liquid-proof plastic coating covering the vinyl webbing (for easy cleanin) and a Delrin side-release buckle; it is widely available in healthcare settings. The maximum enveloping cuboidal dimensions of the PanFab Custom and Commercial PAPRs (including their respective filters) along lateral, longitudinal, and sagittal axes are 21 cm x 25.8 cm x 12.4 cm and 30.6 cm x 33.6 cm x 25.4 cm respectively. Run time for the PanFab blower units was measured to be approximately 3 hours and 55 minutes and charge time approximately 2 hours and 53 minutes at 0.8 amperes charging current, with a variance on the order of a couple of minutes for the runtime and charge time respectively. This compares to the Ford-3M Limited-Use Public Health Emergency PAPR blower units, which weighs 2.7kg, runs for 4-6 hours and has a charge time of 1.5 hours with a 3 Amp hour battery^40^. Alternative battery packs could easily be added to the PanFab design to increase run time; charge time is primarily a function of the charger. The estimated cost in parts for a single unit of the PanFab Custom PAPR is $284 and for the PanFab Commercial PAPR is $328 (note these prices are not for a finished, commercially distributed product and do not include costs such as labor, nor do they account for any discounts available for larger orders). The Ford-3M PAPR has been reported to sell for $715^41^.

The development of functional PanFab PAPR prototypes took a total of eight months with initial product specification and prototyping completed in three months. Additional manufacturing modifications took an additional two months. The latter set of modifications readied the PAPR’s for large-scale production via injection molding. While prototyping was underway, the design validation and testing procedure was established over a period of five months. Design validation was constrained by the availability of testing resources under pandemic conditions and, together with the dearth of documented rationale and goals behind testing specifications that would have allowed for rapid development of alternative testing setups, was the primary factor slowing completion of the project.

### Testing and validation

PanFab PAPR components underwent a series of rigorous testing and validation steps. As mentioned earlier, many traditional NIOSH tests were not readily available from commercial laboratories due to high demand associated with the COVID-19 pandemic. Moreover, tests are highly prescriptive and not easily set up in an academic research laboratory. A further compromise is that full NIOSH certification would not possible for the PanFab PAPR, regardless of testing procedures, in the absence of documentation that it can be manufactured under established quality-control criteria. Compliance with these manufacturing standards is important, but it is secondary to our goals of developing a functional PAPR design. We therefore established alternate test setups and protocols to replicate several NIOSH tests (**Table 2)**. The ability of the final designs to pass these tests should make traditional and non-traditional manufacturers interested in the PanFab designs confident that finished products are very likely to pass full NIOSH certification; we very strongly encourage formal certification testing prior to use of these designs in a healthcare setting.

#### Filter tests

Two filter cartridges were used in PanFab PAPRs. One was an off-the-shelf HEPA-rated vacuum cleaner filter manufactured by Milwaukee, and the other was a P-100 rated filter designed in collaboration with Custom Filters LLC. Two Milwaukee filter cartridges and one Custom Filters filter were challenged with KCl aerosol at 230 lpm. Filtration efficiency with 300 nm aerosol size was found to be 99.99% and 100.00% for two replicate Milwaukee filters and 99.99% for the Custom Filters filter, thereby exceeding the NIOSH salt aerosol filtration efficiency criteria of 99.97%. Equipment was not available to measure filtration efficiency below 300 nm but it is generally observed that HEPA filtration efficiency is lowest at 300 nm and increases as particle size falls^42^. Results from our testing apparatus also correlate with prior testing done at ICS Laboratories, Inc. (Brunswick, OH) for N95-style respirators; ICS Laboratories, Inc. performs third party testing to NIOSH standards using NIOSH STPs (for more information, visit Cleanmask.org^43^).

In the PAO-based instantaneous filtration test carried out by Custom Filters LLC, two Milwaukee filter cartridges and one Custom Filters cartridge were challenged with 90.56 mg/m^3 PAO aerosols at 85 lpm. Filtration efficiency was 99.18% and 99.58% for the two Milwaukee filter replicates and 99.98% for the Custom Filters filter. While the Milwaukee filter does not pass the NIOSH requirement of efficiency higher than 99.97% for oil-based aerosol, consultation with experts on NIOSH certification and regulations led us to conclude that this would not necessarily preclude use in a healthcare setting, given the low concentration of oil aerosols found in this environment. Oil aerosols are primarily a concern in industrial settings in which PAPRs are also used.

#### Air flow tests

Using the apparatus described in the Methods section, flow rate was calculated as the product of the measured velocity and the cross-sectional area of the anemometer. Flow rate with a 70% blower Pulse Width Modulation (PWM) duty cycle was measured to be close to 240 lpm for both filter types. A positive pressure of 40 pascals was recorded inside the facepiece.

#### Facepiece fit tests

Qualitative fit testing, using Bitrex as the testing agent was performed on one subject, as per RCT-APR-STP-0067. Tests were performed with all configurations of the commercial and custom PAPR designs (i.e. using custom and commercial blower units with commercial and custom filter cartridges). The University of Washington (CLiMB)^33^ facepiece was used in our tests. No Bitrex could be tasted by the subject in any of the test configurations, indicating a successful result.

#### Auditory communication tests

Noise level in the facepiece at the ears was measured at between 58.1 to 59.2 dBA at full battery charge and maximum blower speed, which is lower than the 80 dBA limit set in RCT-APR-STP-0030. Low flow alarm sound level at the ears was found to be between 82.95 and 84.7 dBA at battery charge corresponding to low flow condition of 230 lpm flow rate, which passes the 80dBA requirement in CVB-APR-STP-0085. The ability of PAPR wearers to communicate with other individuals was tested with one individual as per CVB-APR-STP-0089 with a 99.9% performance rating (see the STP for definition of performance rating) for listening and 74% for speaking tasks; both pass the required 70% threshold. Full information regarding PanFab PAPR validation testing performed during pandemic conditions is summarized in **Table 2**.

### End-user feedback

To evaluate factors affecting usability in a clinical setting, we created a clinical feedback survey and distributed it to four participants. Two participants were clinicians, who did not use PAPRs regularly prior to the pandemic, and two were research technical staff for whom PAPR use is a standard part of occupational PPE (**Table 3**).

**Table 3:** Test subject demographic information.

| Subject | Height (cm) | Weight (kg) | BMI | Sex | Regular PAPR Use |
| --- | --- | --- | --- | --- | --- |
| 1 | 160 | 57 | 22.3 | Female | No |
| 2 | 175 | 70 | 22.9 | Male | No |
| 3 | 178 | 100 | 31.6 | Male | Yes |
| 4 | 175 | 136 | 42.9 | Female | Yes |

The PanFab PAPR was tested on three criteria: (1) comparison to current PPE options; (2) sense of security with use in a clinical setting; and (3) comfort. Two additional questions assessed the PAPR facepiece alone. Full survey questions and results are available in **Supplementary Material 3**. Four versions of the PanFab PAPR were assessed using different types of filters and blower units: one with a commercial filter and blower unit (PanFab Commercial Design), one with a custom filter and blower unit (PanFab Custom Design), and two versions with mixed custom and commercial filters and housings. Of all PanFab PAPR versions, the PanFab Custom Design performed most favorably: all four respondents rated the PanFab Custom PAPR superior to current PPE options, with a score of 4 to 5 on a 5 Likert-scale across every survey question **(Table 4)**.

**Table 4:** Clinical feedback survey results. Scored averaged among four users.

| Enclosure | Filter | Compare to available PPE, average user score<sup>1</sup> (n=4) | Sense of security with use, average user score<sup>2</sup> (n=4) | Comfort compared to standard PPE, average user score<sup>3</sup> (n=4) |
| --- | --- | --- | --- | --- |
| Custom | Custom | 4 | 4.75 | 5 |
| Custom | Milwaukee | 3.75 | 4.75 | 4.25 |
| Pelican | Custom | 3.25 | 4.5 | 3.75 |
| Pelican | Milwaukee | 3 | 4 | 3.25 |

Note 1: Score of 1 = PanFab PAPR much worse than current PPE options, 5 = PanFab PAPR much better than current PPE options
Note 2: Score of 1 = Very uncomfortable, 5 = Very comfortable
Note 3: Score of 1 = PanFab PAPR much worse than standard, 5 = PanFab PAPR much better than standard

The three other versions of the designs (with a commercial blower unit, filter, or both) performed favorably, with survey responses consisting of scores ranging from 3-5. Participants experienced more issues with mobility as compared to the fully-custom PAPR, and comments on PAPR versions using commercial parts emphasized the need for better weight redistribution to improve balance. Participant comments across all PAPR design versions focused on possible improvements regarding the sizing and comfort of the PAPR facepiece, which was not a component of the current study. In sum, clinical feedback demonstrated that the PanFab PAPR is a favorable alternative form of PPE in the face of supply shortages in terms of user comfort, mobility, and sense of security with use.

## DISCUSSION

The successful design, production and testing of a PAPR by a volunteer team comprising medical professionals, scientists, student engineers and concerned citizens (PanFab) demonstrates the potential for addressing pandemic-related shortages of relatively complex types of PPE using a rapid and iterative approach to prototyping and design^31^. The process generated near-final PAPR designs with full-time effort by three graduate engineering students, support from a clinical specification and testing team, access to standard academic laboratories and modest financial support. Testing required more time than design and fabrication, as discussed below.

### Design and Results

The PanFab Commercial Design used commercially available components with custom-fabricated modifications while the PanFab Custom Design used additive manufacturing (3D-printing) to create a fully customized, lighter weight, and smaller enclosure. Frequent feedback from clinicians who use PAPRs in a hospital setting strongly influenced design decisions, particularly with respect to PAPR comfort and usability. Key design decisions included determining size and orientation of the motor-blower housing, orientation of the filters, and the method of donning/doffing the motor-blower unit. Feedback from manufacturing experts yielded a design that is amenable to large-scale production by injection molding the motor-blower housing component. PanFab PAPR designs are modular and compatible with several standard commercial PAPR components, including facepieces and filters, allowing for substitution of components in limited supply. The PanFab Custom Design compares favorably to the 3M Ford Limited-Use Public Health Emergency PAPR^13^ with respect to weight and size, although the 3M Ford unit appears to use higher performance batteries.

The safety and functionality of PanFab PAPRs was evaluated using protocols that closely followed NIOSH STP’s and aimed to meet or exceed the functional objectives of those tests. Given the limitations imposed by pandemic conditions, it was necessary to use substitute tests in university laboratories rather than use a NIOSH-specific apparatus at a commercial pre-certification laboratory. PanFab PAPRs passed all of the performed tests, in various combinations of commercial and custom components. User feedback on the PAPRs was obtained from clinicians inexperienced in using PAPRs and technical research staff who routinely use PAPRs in a major Boston-area hospital. The PanFab Custom Design scored favorably as compared to the traditionally manufactured PAPRs (primarily from ILC Dover) available to hospital staff.

### Challenges in design, testing, and regulatory approval

During early specification and prototyping, we faced significant challenges in locating relevant design criteria for PAPRs: there also exists very limited information in the public domain on testing equipment that could be used to validate prototypes based on physical properties and objective engineering criteria. Answers to questions such as minimum time of continuous operation required, relevant material characteristics for facepiece fabrics, and appropriate materials for components in the pathway for inhaled, filtered air were not readily available. We therefore sought out experts with relevant expertise. Substantial time and effort would have been saved had a centralized resource of information been available. To streamline the process for future crises, we have consolidated relevant information collected from US regulatory agencies in supplementary materials. We encourage individuals from other countries to contact us with information relevant to products in their markets and will provide this updated information on PANFAB.ORG.

When it came to evaluating our designs, we found that it was difficult to use NIOSH standard testing procedures since the necessary equipment was not readily available. Informed substitution was made difficult by prescriptive procedures and opaque objectives in terms of fundamental mechanical or physical principles being assessed. Our use of third-party commercial testing labs that test to NIOSH standards was also limited by long lead times and a requirement for multiple samples of each prototype, which would require financial resources beyond those available to our group. Thus, testing equipment and protocols, as opposed to design and fabrication, emerged as the primary challenge in developing the PanFab PAPR.

Despite our best efforts, regulatory hurdles remain to use PanFab PAPRs in a clinical setting. To receive NIOSH certification, a product must be manufactured and submitted by a NIOSH-approved manufacturer with a quality management system in place. Under normal circumstances, this requirement guarantees the safety of products made in volume. However, this restricts the development of new products to NIOSH-approved manufacturers, which has had the effect of creating near-monopolies for some types of PPE. To improve resilience in future emergencies, regulators might consider how to optimally balance the risk of non-traditional PPE against the risk of no protection at all. We propose that consideration be given to rules that allow non-NIOSH certified fabricators to respond to declared healthcare emergencies while still complying with the most critical aspects of functional testing. Under this process, critical “go or no-go” tests would be defined by clearly-described physical principles and corresponding testing protocols that could be performed on generally available laboratory equipment. The modified standards would include an explicit description of the end goal of the test and suggest a range of alternative devices that can be used to measure airflow, filtration efficiency, audibility, user testing etc. Rational substitution of instruments would also be allowed. While we do not advocate for relaxing standards under non-crisis conditions, modifying and streamlining testing procedures for prototype devices would reduce barriers to the entry of new products and promote innovation.

## Conclusions

The current COVID-19 crisis has revealed major weaknesses and points of failure in our health care system and its supply chains, particularly for PPE. This does not come as a surprise. Multiple studies over a 15-year period have decried the absence of innovation in design and provision of respiratory protection for health care and other essential workers^10^. For example, a 2006 report by the Institute of Medicine (IOM) called for urgent research to inform the design and development of new medical masks and respirators;^44^ a 2008 IOM report addressed the design and engineering of more effective PPE;^45^ the 2009 Project BREATHE report laid out a comprehensive action plan for a new generation of respirators;^46^ and a 2019 consensus report from the US National Academies echoed the urgent needs identified^47^. Despite these repeated calls for action and greater innovation, there has been little response from the commercial sector or from government: in the COVID-19 pandemic, most innovation has come from volunteer groups of scientists and clinicians allied with maker communities with access to rapid prototyping and fabrication equipment, technology that is increasingly inexpensive and available to ordinary citizens.^10^ Thus, open source product development (OSPD)^25^ emerges as perhaps the only avenue to mitigating existing weaknesses while increasing product innovation under both normal and crisis conditions.

An OSPD approach is not a panacea and it is not free. As discussed above, it would be very helpful for NIOSH and other regulatory agencies to develop less prescriptive testing procedures for products such as PAPRs. Funding is also essential. The work described here benefited from the generosity of many individuals but all attempts to fund it via competitive applications to foundations or universities were turned down because research into respiratory protection is not considered innovative by conventional academic criteria (funding was, however, temporarily available under relaxed grant guidelines from the US National Cancer Institute under NOT-CA-20-054). This speaks to a larger problem in matching acute healthcare needs to available expertise and necessary resources in academia, not just industry.

The production of a regulated medical product is difficult to achieve in the absence of commercial expertise. This is consistent with previous data showing that OSPD is most effective within the context of private-public partnerships^48^. We are therefore working with an industry partner to develop PanFab PAPRs into commercial products. However, all the PanFab PAPR designs and software described here remain public domain resources and are available under non-restrictive Creative Commons Attribution-Share Alike 4.0 International Public License; the design files, CAD files, and code are available on GitHub. All materials needed for the construction and use of the design are also available in **Supplementary Materials 2 and 4** and through the online repository https://github.com/labsyspharm/PanFab-PAPR-2021. We hope that these materials serve as a resource for further development and innovation.

## Data Availability

All study materials and data are available in Supplementary Materials and through the online repository https://github.com/labsyspharm/PanFab-PAPR-2021.

## ACKNOWLEDGMENTS

We thank Jennifer Delaney (Washington University), Hunter Engineering Company (Bridgeton, MO), Joseph Iaquinto (University of Washington), Ben Linville-Engler (Manufacturing Emergency Response Team), Jinhan Mo (Tsinghua University), the MGB Full Body Protection Working Group, the Ju Li Group (MIT), the Rutledge Lab (MIT), Paul Nederhoed (Paul Nederhoed Photography, LLC), David Krikorian and Meghan Patterson (DFCI), Mike Copponi (DFCI), Brandon Beller (PanFab), Thomas Pouchot (NIOSH), Norman Wen (Emulate Inc), David Turner (DFCI Animal Research Facility), Myron Kassaraba (MIT Technology Licensing Office), and Grant Zimmermann (Harvard Office of Technology Development).

Local citizens and engineers have generously donated their time and resources to PanFab and they are essential to program success. This work was supported by the MIT COVID-19 Emergency Fund, Harvard-MIT Center for Regulatory Science by NIH/NCI grants P30-CA006516, U54-CA225088 (to PKS, NL and DP) and by T32-GM007753 (to DP) and by the Harvard Ludwig Center. AKC is supported by the Hugh Hampton Young Fellowship of MIT.

## AUTHOR CONTRIBUTIONS

PanFab PAPR design, prototyping, and production: A.K., L.A., D.P., F.W., C.D., A.C., J.F., O.M., P.G., C.V., C.H., M.S.S., N.R.L., P.K.S.

PanFab PAPR filtration testing: E.T., J.L.

PanFab PAPR prototype iteration and clinical feedback coordination: N.R.L.

Manuscript writing: A.K., L.A., D.P., P.K.S.

Manuscript review and editing: A.K., L.A., D.P., F.W., C.D., A.C., R.M., J.F., E.T., O.M., P.G., C.V., C.H., H.Y., M.S.S., J.L., S.H.Y., N.R.L., P.K.S.

Greater Boston Pandemic Fabrication Team (PanFab) Consortium Coordination: D.P., H.Y., N.R.L., P.K.S.

## ETHICS APPROVAL AND CONSENT TO PARTICIPATE

The Partners Healthcare Institutional Review Board declared that the study did not require ethical oversight (protocol #2021P000727).

## SUPPLEMENTARY MATERIALS

**Supplementary Material 1:** NIOSH PAPR standard testing procedures.

**Supplementary Material 2:** Full description of PanFab PAPR design methodology and selected prototype components.

**Supplementary Material 3:** Full clinical survey questions and results.

**Supplementary Material 4:** PanFab PAPR design materials and instructions, including: 3D-printing instructions, Arduino code, assembly instructions, bill of materials, CAD design files, custom Arduino shield design, and use instructions.

