## Supplementary Material 1 for "De novo Powered Air-Purifying Respirator Design and Fabrication for Pandemic Response"

UNCONTROLLED COPY FOR INSPECTION AND USE

### DETERMINATION OF RESPIRATOR FIT, QUANTITATIVELY USING CORN OIL AEROSOL, FOR POWERED AIR-PURIFYING RESPIRATORS WITH LOOSE-FITTING RESPIRATORY INLET COVERINGS, STANDARD TESTING PROCEDURE (STP)

#### 1. PURPOSE

This procedure establishes the method by which a generated corn oil aerosol is used for determining if powered, air-purifying respirators (PAPRs) supplied with loose fitting respiratory inlet coverings meet the facepiece-fit requirements at 42 CFR Part 84, Subpart K, Section 84.176(b).

#### 2. GENERAL

This STP describes the Determination of Respirator Fit, Quantitatively Using Corn Oil Aerosol, For Powered Air-Purifying Respirators With Loose-Fitting Respiratory Inlet Coverings test procedure in sufficient detail that a person knowledgeable in the appropriate technical field can select equipment with the necessary resolution, conduct the test, and determine whether or not the evaluated product passes the test.

#### 3. EQUIPMENT/MATERIAL

3.1. TSI Rear Light Scattering Laser Photometer, model 8587A, or equivalent with concentration range 1.0 μg/m$^3$ to >200 mg/m$^3$. See Figure 1.

3.2. NIOSH Dynamic Fit Software. (The software is used to monitor the particle count both inside and outside the respirator, calculate a fit factor for each exercise and the average fit factor for the trial in real time.)

3.3. Aerosol Generator, MSP Model 2045 High Output Aerosol Generator or equivalent - The aerosol generator is required to be capable of maintaining 5 to 100 mg/m$^3$ of corn oil challenge aerosol concentrations with a Mass Median Aerodynamic Diameter (MMAD) of 0.4 to 0.6 μm for the specified test duration in the test chamber. See Figure 2.

3.4. TSI model 8530, DustTrak II Aerosol Monitor, or equivalent - Range 0.001 to 400 mg/m$^3$ (Calibrated to ISO 12103-1, A1 test dust), and resolution ±0.1% of reading or ±0.001 mg/m$^3$, whichever is greater – See Figure 3.

3.5. Corn Oil - 99% Pure. CAS Number 8001-30-7 - Commercial product names are Maise/Maize Oil, Maydol and Mazola Oil.

3.6. Scanning Mobility Particle Sizer (SMPS), TSI model 3936 series - SMPS is composed of the TSI model 3080 Electronic Classifier, TSI model 3775 Condensation Particle Counter and Long Differential Mobility Analyzer (DMA). See Figure 4.

3.7. Environmental test chamber - The chamber shall be designed so that the individual(s) performing fit factor testing are visible at all times while in the chamber. The chamber design must include an entry vestibule designed to allow safe entry and exit from the chamber with minimal disturbance to both the aerosol concentration and the concentration uniformity. An example of a charged corn oil chamber is illustrated in Figure 5.

3.8. Chamber Communications - Electronic audio communications (chamber loudspeaker) are employed to transmit a real-time voice signal from laboratory technicians to test subjects to ensure that test subjects can clearly hear when to start and stop the test exercise regimen or receive safety information.

3.9. Facial Size Measurement Calipers - Calibrated face sizing calipers shall be used to measure the human test subject to the requirements identified in Appendix A. Examples of calipers are sliding measurement calipers: Seritex model GPM 104, 0-200 mm length, or spreading measurement calipers: Seritex model GPM 106, 0 – 300 mm width. These are shown in Figure 6 and Figure 7.

3.10. Facepiece Direct Probes. The sample probes shall be of the shape defined by Liu [AIHAJ (45); 278-283, 1984] and shall not interfere with the fit or function of the respirator. Figure 8 and Figure 9 are photographs of the probes used. Each probe bulkhead fitting is sealed using two rubber washers, one metal washer and one nut.

4. <u>TESTING REQUIREMENTS AND CONDITIONS</u>

4.1. Prior to beginning any testing, confirm that all measuring equipment employed has been calibrated in accordance with the testing laboratory's calibration procedure and schedule. All measuring equipment utilized for this testing must have been calibrated using a method traceable to recognized international standards when available.

4.2. General respiratory inlet covering fit requirements for PAPRs -

4.2.1. The fit test shall be performed using a panel of test subjects of various facial sizes measured in accordance with the NIOSH Bivariate Panel (NIOSH Panel). The measured face length and face width are used to designate the subject's NIOSH Panel cell number, as illustrated in Figure 10.

4.2.1.1. Face Width is the Bizygomatic Breadth measurement (Figure 9), using the spreading measurement calipers.

4.2.1.2. Face Length is the Menton-Sellion measurement (Figure 9), using the sliding measurement calipers.

4.2.2. Any PAPR part which must be removed to perform the respiratory inlet covering fit test shall be replaceable without special tools and without disturbing the facepiece on the wearer's face.

4.2.3. The respirator, including the respiratory inlet covering, shall be adjusted

UNCONTROLLED COPY FOR INSPECTION AND USE

according to the manufacturer's user instructions, prior to entering the chamber; however, upon entry into the test chamber neither the respirator, nor the respiratory inlet covering shall not be re-adjusted.

4.3. Test subject selection

4.3.1. For PAPRs with up to three sizes of loose-fitting respiratory inlet coverings, the test will be conducted employing 18 individual test subjects which represent the NIOSH Panel (Appendix 8.5). See Table 1 for the suggested test subject distribution in relation to the NIOSH Panel (NIOSH is allowing flexibility in the use of subjects from well-populated panel cells. NIOSH will attempt to test using a panel that contains at least one subject from each cell, but when subjects from all cells are unavailable they may be supplanted by adding subjects from more populated cells; however, no more than four subjects from any one cell may contribute to the overall test panel composition for any respirator under evaluation.)

Table 1: Recommended test subject distribution to be used for fit testing in relation to the NIOSH Panel

| NIOSH Panel – Cell Number | Number of Test Subjects |
|---|---|
| 1 | 1 |
| 2 | 1 |
| 3 | 2 |
| 4 | 4 |
| 5 | 1 |
| 6 | 1 |
| 7 | 4 |
| 8 | 2 |
| 9 | 1 |
| 10 | 1 |

4.3.1.1. For PAPRs supplied with respiratory inlet coverings in only one size - a subject failing to achieve a pass in their initial trial is considered to be a face-size failure. (no alternate size to try)

4.3.1.2. For PAPRs supplied with loose-fitting, respiratory inlet coverings in two sizes -

4.3.1.2.1. Subjects from panel cells 1-4 and cell 6 shall be tested first wearing the smaller size inlet covering. If the subject does not achieve a trial pass in the smaller size inlet covering, the subject can be tested again (second trial) wearing the larger size inlet covering.

4.3.1.2.2. Subjects from panel cells 7-10 and cell 5 shall be tested first wearing the larger size inlet covering. If the subject does not achieve a trial pass in the larger size inlet covering, the

UNCONTROLLED COPY FOR INSPECTION AND USE

subject can be tested again (second trial) wearing the smaller size inlet covering.

4.3.1.2.3. A subject failing to achieve a trial pass in either of the sizes available for testing is considered to be a face-size failure.

4.3.1.3. For PAPRs supplied with loose-fitting, respiratory inlet coverings in three sizes -

4.3.1.3.1. Subjects from panel cells 1 and 2 shall be tested wearing the smaller size initially.

4.3.1.3.2. Subjects from panel cells 3-7 shall be tested wearing the regular/medium size initially.

4.3.1.3.3. Subjects from panel cells 8, 9, and 10 shall be tested wearing the larger size initially.

4.3.1.3.4. If a subject does not achieve a trial pass in the first inlet covering size evaluated, the subject will be retested in the next available size (second trial). A subject failing in the smaller size inlet covering, can try the medium size (second trial), and then larger size (third trial) inlet covering. A subject failing to achieve a trial pass in the medium size inlet covering can try the smaller and larger size inlet coverings. A subject failing in the larger size inlet covering can try the medium, then the smaller size inlet covering. A subject failing to achieve a trial pass in any of the sizes available for testing is considered a face-size failure.

4.3.1.3.5. The test administrator may determine whether or not third trials are needed for test completion. For a subject failing to achieve a trial pass in the small or medium size inlet covering, trying the large size may not be necessary since the large size may be obviously too big for the subject. For a subject failing to achieve a trial pass in the large and medium size inlet covering, trying the small size may not be necessary since the small size may be obviously too small for the subject.

5. PROCEDURE

5.1. Chamber Set-up

5.1.1. Turn on air handling unit with sufficient airflow to maintain the proper corn oil concentration.

5.1.2. Turn on vacuum pump for laser photometers

UNCONTROLLED COPY FOR INSPECTION AND USE

UNCONTROLLED COPY FOR INSPECTION AND USE

5.1.3. Turn on mixing fans to 6.1 volts.

5.1.4. Turn on air compressor for corn oil generators to maintain the proper corn oil concentration. Corn Oil Challenge Concentration = 30 to 40 mg/m$^3$.

5.1.5. Turn on laser photometers.

5.1.6. Turn on SMPS and warm up for 15 minutes.

5.1.7. Turn on DustTrak.

5.1.8. Allow 30 minutes for the chamber concentration to stabilize.

5.1.9. Use the DustTrak to monitor the chamber concentration.

5.1.10. Adjust the air pressure at the generators regulator to establish the corn oil concentration of 30 to 40 mg/m$^3$.

5.1.11. Use the SMPS according to the manual to determine the particle size. The correct size should be 0.4 to 0.6 µm with a geometric standard deviation of less than 2.0.

5.2. <u>Conducting the Corn Oil Test</u>

5.2.1. The Users Instructions (UI) provided with the test samples shall be reviewed by all test facility personnel. Test subjects will be taught by the test facility administrator on the areas of manufacturer's size selection, donning, doffing and procedures related to the accessories as specified by the UI.

5.2.2. Test subject training will be conducted by test facility personnel based on the manufacturer's users' instructions. Each test subject shall perform an unassisted donning of the respirator. Self-donning under supervision of the test administrator is permitted to make the appropriate adjustments to the respiratory inlet covering until they are satisfied that they are wearing the respirator in compliance with the manufacturer's users' instructions. Expert donning is not allowed in the conduct of this test.

5.2.3. Subjects will be assigned to a specific photometer and moved to the chamber in groups of four or less based on the number of photometers.

5.2.4. Test subjects entering and leaving the corn oil-charged chamber must enter the vestibule first. Once the outside door is closed, the interior door is opened to allow subjects in the chamber. Once subjects are in the chamber they will be instructed to attach their sample line tubing to their assigned photometer. Chamber concentration is required to be monitored continuously during the entire duration of each individual (face-size trial) Corn Oil test.

5.2.5. Information for each test subject will be recorded in the NIOSH Dynamic Fit software program. Test Administrator will start the software program and relay the information of time to start the test, exercise, and timing of the exercise being performed.

5.2.6. A fit factor test consists of a set of four two-minute standard exercises. During the test, each human subject will perform the following four exercises for two minutes each in the below listed sequence. Subjects should not touch any portion of the respirator during any part of the testing exercises. Test administrator will give verbal commands to stop and start each exercise.

5.2.6.1. Two (2) minutes nodding up and down and turning head side to side.

5.2.6.2. Two (2) minutes callisthenic arm movements.

5.2.6.3. Two (2) minutes running in place.

5.2.6.4. Two (2) minutes pumping with tire pump.

5.2.7. Instruct the subjects to disconnect sample line from the photometer. Exit the chamber using the vestibule room. Inform the subject to return to the ready line and await further instructions for doffing the respirator or leaving the respirator donned. Subjects that are being reviewed for test failure protocol will remain with respirator donned until instructed to doff.

5.2.8. An overall pass/fail statement for each individual will be recorded by the NIOSH Dynamic Fit software and written on the test data sheet as shown in attachment 8.1.

5.2.9. All comments and observations by test subjects, which are voluntary, will be written on the test data sheet.

5.2.10. If a respirator is identified as a failure upon trial termination, test administrator will conduct failure assessment protocol of the respirator in two phases. First phase is to inspect the respirator while it is still donned on the test subject. Second phase is to inspect the respirator when it is doffed. Post-test failure analysis should consist of inspection of the test subjects eye to eye lens positioning, head harness positioning, head harness strap twists, probe loose, missing or on a molded seal or surface causing seal gap or any other case dependent situations. If noted deficiencies are confirmed with the respirator being improperly probed, reassign another like respirator to the test subject and retest. If the respirator has a serviceable probe but continues to fail, log it as a Corn Oil failure. Only inspect the probe assembly if test results are consistently failing or suddenly failing after successful exercise results are indicated. Probe failures such as ripped face blank material or inadequate probe sealing areas are cause for reanalysis of the determined probe entry point.

6. <u>PASS/FAIL CRITERIA</u>

UNCONTROLLED COPY FOR INSPECTION AND USE

6.1. The requirement for passing this test is set forth in 42 CFR Part 84, Subpart K, Section 84.176(b).

6.2. The number of face-size failures will not exceed four.

6.3. If an overall pass is achieved, but three subjects report the same issue about the comfort of the facepiece, the test will be considered a failure.

6.4. Fit factor for loose-fitting PAPR -

6.4.1. For each face-size trial, an overall average fit factor of 500 must be achieved.

6.4.2. A minimum average fit factor of 500 must be achieved during each of the subject exercises which comprise a single face-size trial.

7. RECORDS/TEST SHEETS

7.1. All test data collected will be recorded on the appropriate Determination of Quantitative Corn Oil Fit Test data sheet.

8. ATTACHMENTS

8.1. Example Data Sheet - PAPR Fit Corn Oil Test Data Sheet – Page 1

8.2. Example Data Sheet - PAPR Fit Corn Oil Test Data Sheet – Page 2

8.3. Photographs

8.3.1. Figure 1. Photograph of Laser Photometer

8.3.2. Figure 2. Photograph of Aerosol Generator

8.3.3. Figure 3. Photograph of DustTrak II Aerosol Monitor

8.3.4. Figure 4. Photograph of Scanning Mobility Particle Sizer

8.3.5. Figure 5. Photograph of Charged Test Chamber

8.3.6. Figure 6. Photograph of Sliding Calipers

8.3.7. Figure 7. Photograph of Spreading Calipers

8.3.8. Figure 8. Photograph of Front View of Sample Probe

8.3.9. Figure 9. Photograph of Side View of Sample Probe

8.4. Figure 9. Anthropometric Measurements

8.5. Figure 10. Diagram, NIOSH Panel

UNCONTROLLED COPY FOR INSPECTION AND USE

8.1. Example Data Sheet – Page 1

**National Institute for Occupational Safety and Health**
**Respirator Branch**
**Test Data Sheet**

NIOSH

**Task Number:** TN-XXXXX
**Test:** PAPR Fit Testing (Cornoil)
**Manufacturer:** Company Name
**Item Tested:**
**Minimum Fit Factor:**

**Reference No.:** CFR 84.176(b)
**STP No.:**

| Subject | Face Size | Facepiece | Result | |
|---|---|---|---|---|
| | | | Trial 1 | Trial 2 |

**Overall Result:** ____________________

**Test Operator:** ____________________ **Date:** ____________
**Engineering Technician**

**Comments:**

**Was all equipment verified to be in calibration throughout testing?**

**Test Operator Signature:** ______________________

UNCONTROLLED COPY FOR INFORMATION USE ONLY

UNCONTROLLED COPY FOR INSPECTION USE

8.2. Example Data Sheet – Page 2

Fit Testing Report

| Name | GAGEPak # | | Calibration Due |
|---|---|---|---|
| TSI Photometer | | | |
| TSI Photometer | | | |

8.3. Photographs

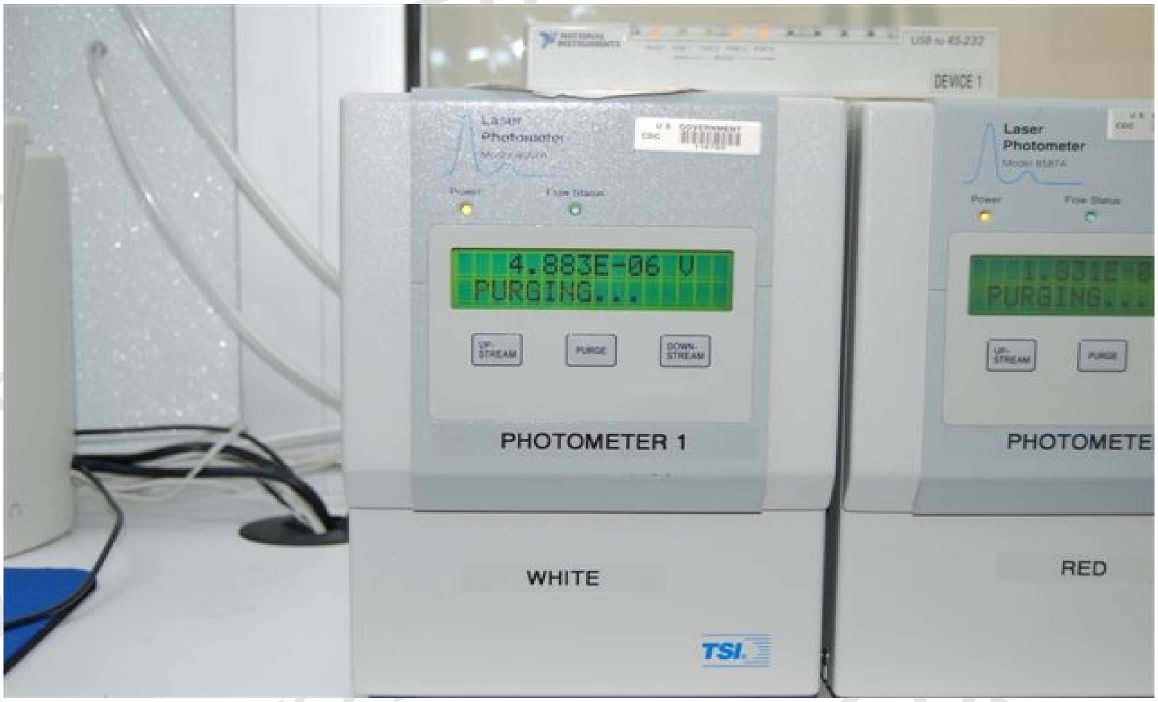

Figure 1: Laser Photometer, Model 8587A

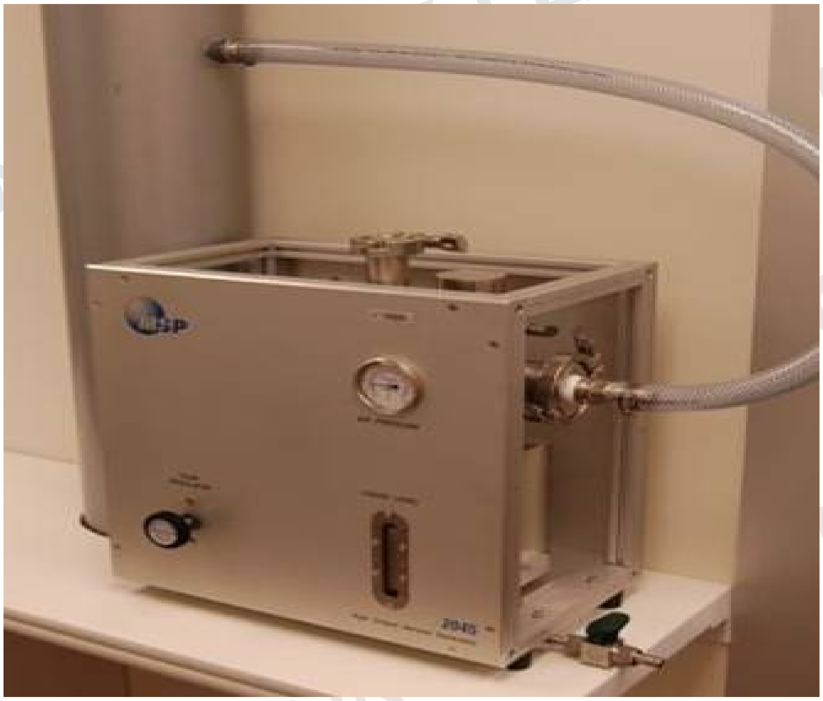

Figure 2: Aerosol Generator

8.3. Photographs

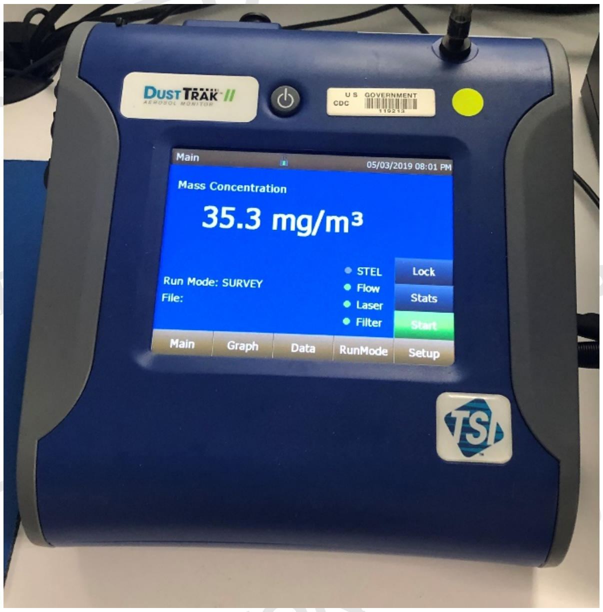

Figure 3: DustTrak II Aerosol Monitor

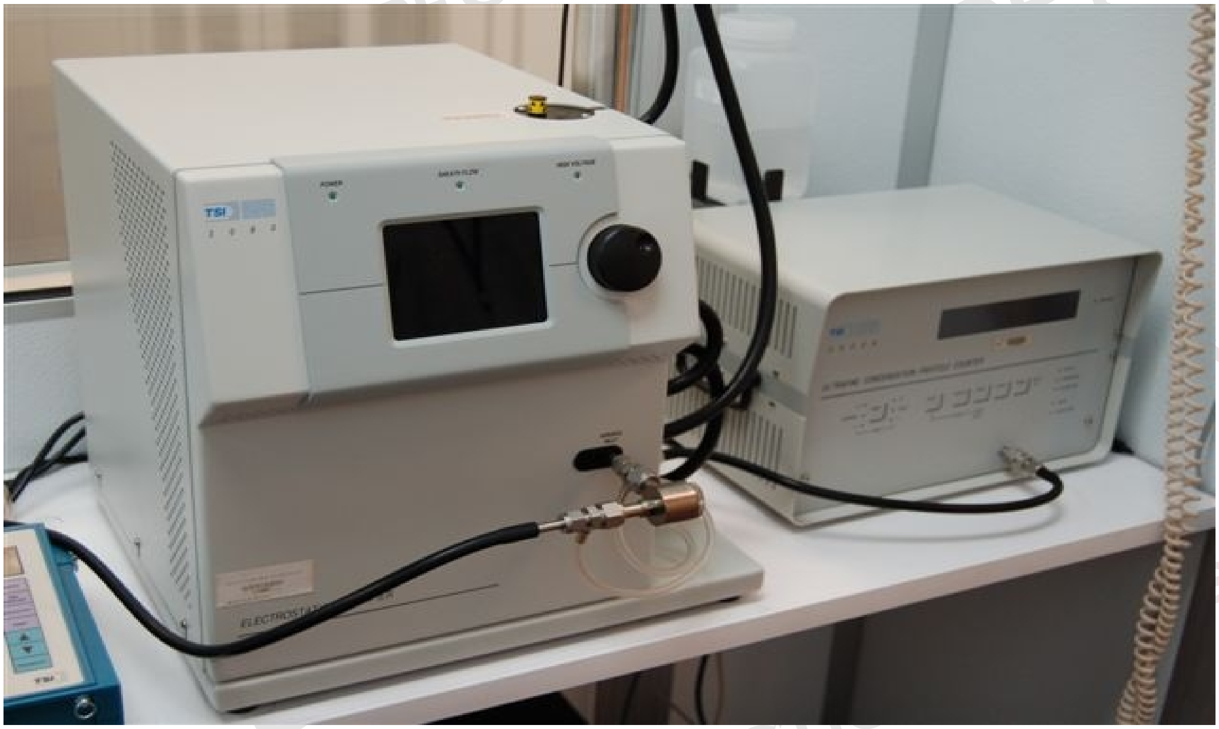

Figure 4: Scanning Mobility Particle Sizer

UNCONTROLLED COPY FOR INSPECTION USE

8.3. Photographs

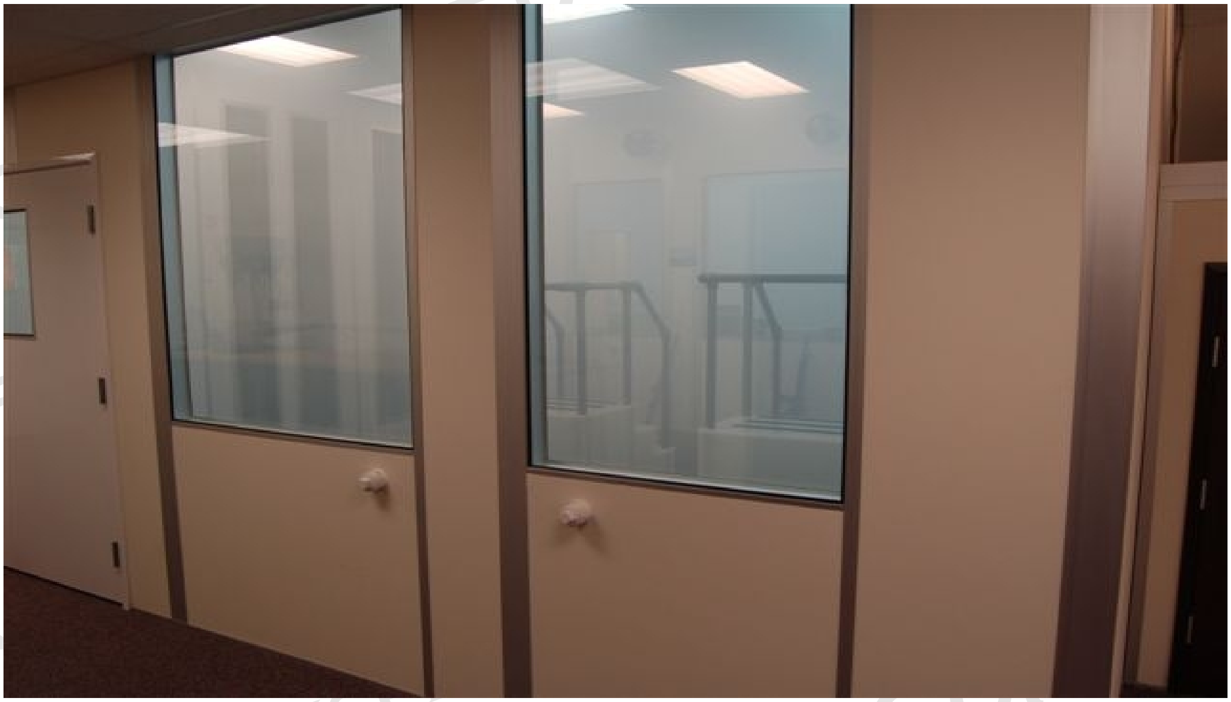

Figure 5: Charged Test Chamber

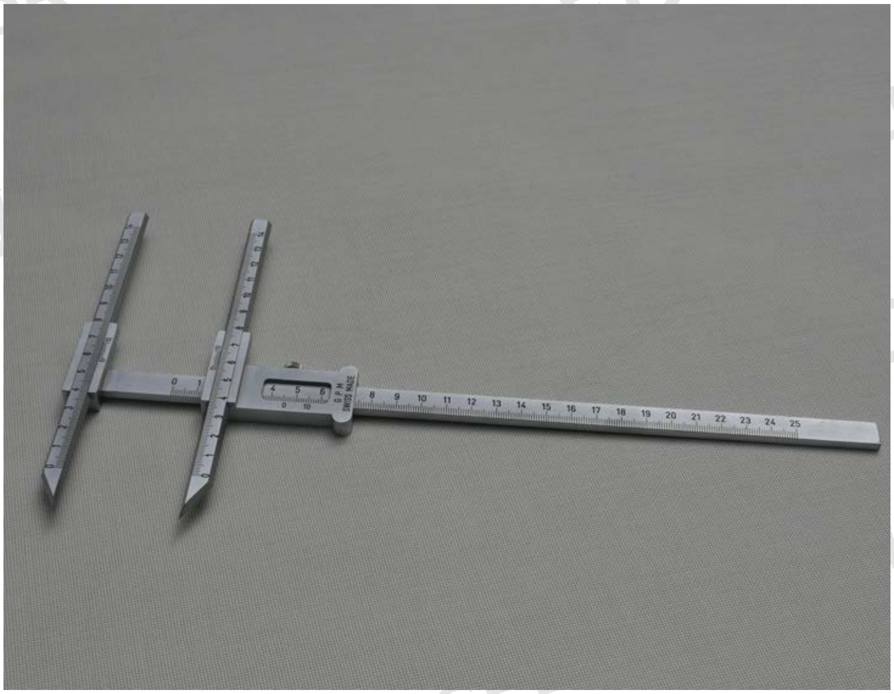

Figure 6: Sliding Calipers

8.3. Photographs

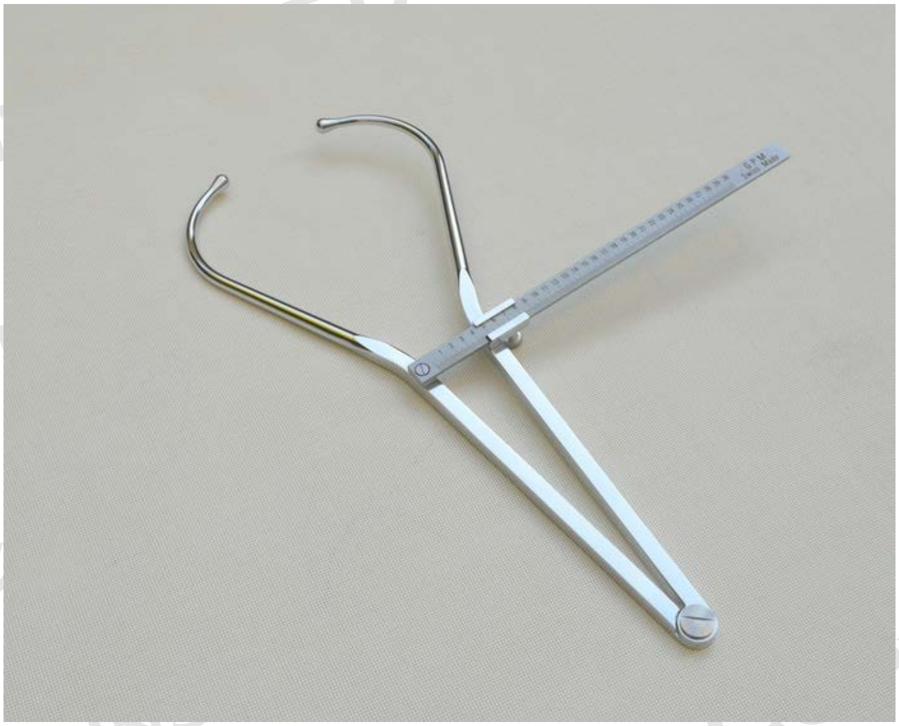

Figure 7: Spreading Calipers

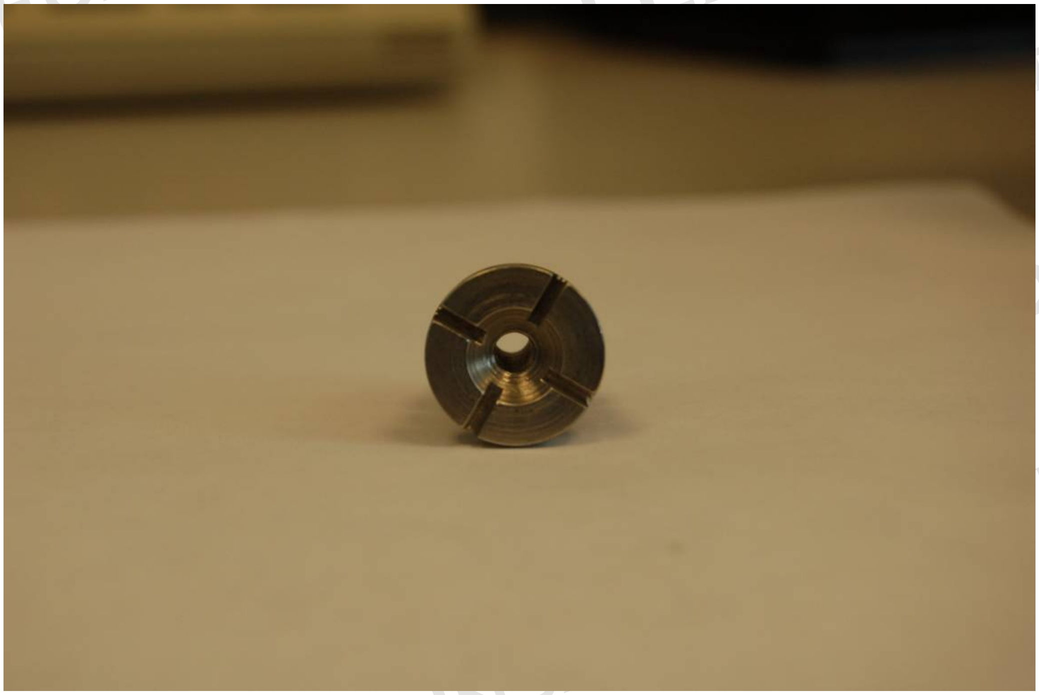

Figure 8: Front View of Sample Probe

UNCONTROLLED COPY FOR INFORMATION USE

8.3 Photographs

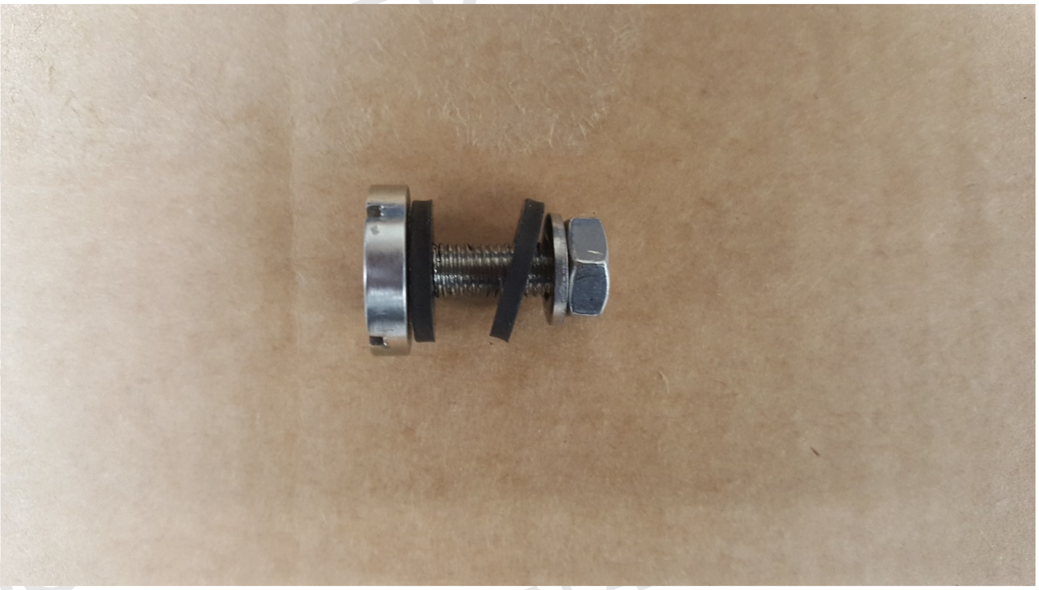

Figure 9. Side View of Sample Probe

UNCONTROLLED COPY FOR INSPECTION AND USE
UNCONTROLLED COPY FOR INSPECTION AND USE
UNCONTROLLED COPY FOR INSPECTION AND USE

8.4. Anthropometric Measurements

| Description | Definition | Diagram |
|---|---|---|
| **Bizygomatic Breadth** | Maximum horizontal breadth of the face as measured with a spreading caliper between the zygomatic arches. | 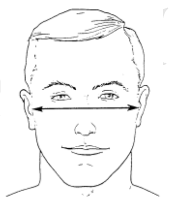 |
| **Menton–Sellion Length** | Distance as measured with a sliding caliper in the midsagittal plane between the menton landmark and the sellion landmark. | 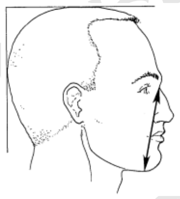 |

Figure 9: Anthropometric Measurements

UNCONTROLLED COPY FOR INSPECTION AND USE

8.5. NIOSH Panel

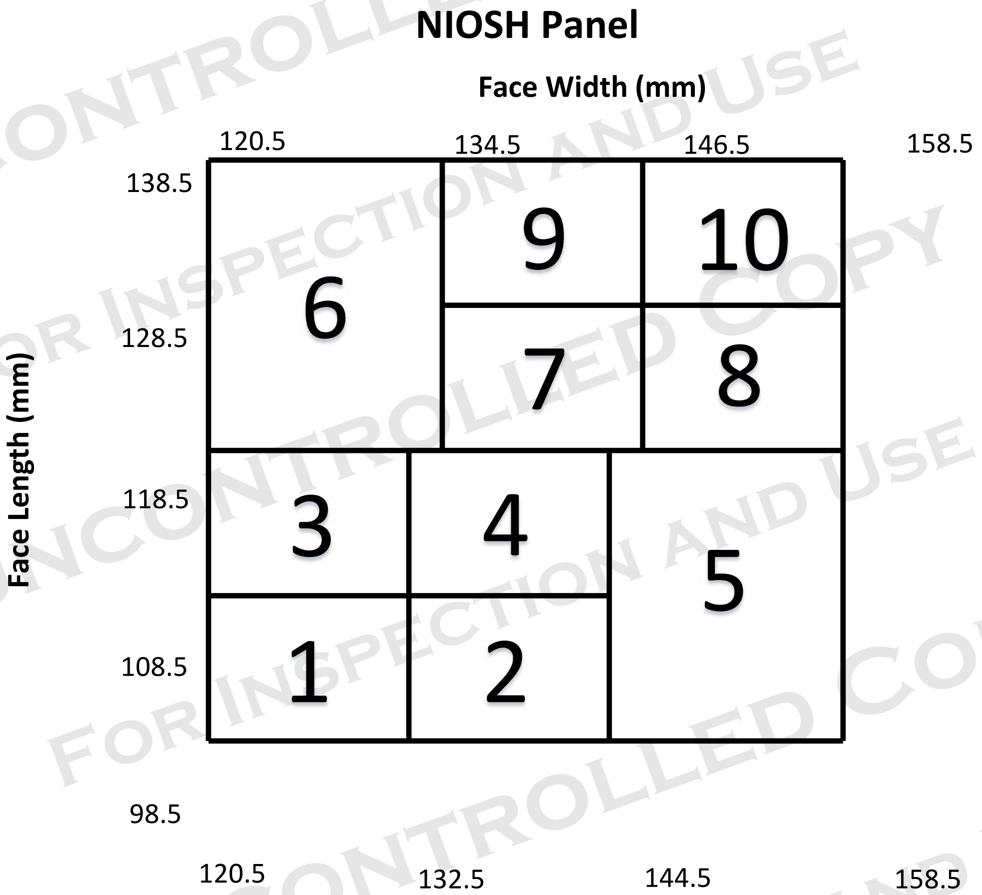

Figure 10: NIOSH Bivariate Panel (NIOSH PANEL)

UNCONTROLLED COPY FOR INSPECTION AND USE

### Revision History

| Revision | Date | Reason for Revision |
|---|---|---|
| 0.0 | 18 March 2020 | Original release |

UNCONTROLLED COPY
FOR INSPECTION AND USE

### DETERMINATION OF PARTICULATE FILTER EFFICIENCY LEVEL AGAINST SOLID PARTICULATES FOR POWERED AIR-PURIFYING RESPIRATORS (PAPRs), SERIES PAPR100-N, STANDARD TESTING PROCEDURE (STP)

#### 1. PURPOSE

This procedure establishes the means for ensuring that the particulate filtering efficiency of PAPR100-N series filters meet the requirements set forth in 42 CFR, Part 84, Subpart K, Section 84.180. These filters or filter cartridges may be integral to respirator construction; mounted individually, or in sets of up to three; used in conjunction with filters, cartridges and canisters for half-mask, full facepieces, hoods, and helmets.

#### 2. GENERAL

This STP describes the test method to be used for the Determination of Particulate Filter Efficiency Level Against Solid Particulates for Powered, Air-purifying Respirators, Series PAPR100-N, test procedure in sufficient detail that a person knowledgeable in the appropriate technical field can conduct the test and determine whether, or not the product passes the test.

#### 3. EQUIPMENT/MATERIALS

3.1. The list of necessary test equipment and materials follows.

3.1.1. TSI Model 8130 Automated Filter Tester or equivalent instrument. Air flow control accuracy is 2% of full scale. Pressure measurement accuracy is 2% of full scale. Penetrations can be measured to 0.001%, efficiencies to 99.999%.

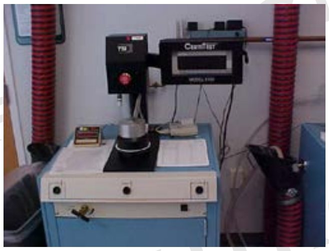

3.1.2. Microbalance accurate to 0.0001 grams (g).

3.1.3. Type A/E glass filters, 102 mm diameter, high efficiency filters with a 1 micrometer pore size.

3.1.4. Timer (accurate to 0.01 percent).

3.1.5. 2% sodium chloride solution in distilled water (NaCl).

UNCONTROLLED COPY FOR INSPECTION AND USE

3.1.6. Temperature and humidity chamber capable of maintaining 38 ±2.5 °C and 85 ± 5% relative humidity.

3.1.7. Respirator filter holder supplied for specific manufacturer type which is compatible with TSI filter tester. NIOSH will not be obligated to use these holders for actual certification testing. All manufacturer test fixtures must be correlated with the NIOSH test method.

3.1.8. Thermal printer (supplied with TSI 8130) or optional data acquisition system.

3.1.9. TSI, Green Line paper, part number 813010. Lot number must be included on each box. Each lot number must include the "Penetration vs. Resistance graph".

4. TESTING REQUIREMENTS AND CONDITIONS

4.1. Prior to beginning any testing, confirm that all measuring equipment employed has been calibrated in accordance with the testing laboratory's calibration procedure and schedule. All measuring equipment utilized for this testing must have been calibrated using a method traceable to recognized international standards when available.

4.1.1. Respirator filters and filter cartridges shall be tested as follows. Filters used in conjunction with PAPR100-Ns, and odd or unusually shaped filters may be tested on a test fixture provided by the applicant.

4.1.2. If a test fixture is supplied by the applicant, the test fixture shall have a serial number or other unique, easily referenced identifier permanently etched, engraved, or affixed.

5. PROCEDURE

5.1. Respirator filters will be challenged by a NaCl aerosol at 25 ±5 °C and a relative humidity of 30 ±10% that has been neutralized to the Boltzmann equilibrium state. The particle size distribution will be a count median diameter of 0.075 ± 0.020 micrometer and a geometric standard deviation not exceeding 1.86. Each respirator filter unit will be challenged with an aerosol concentration not exceeding 200 $mg/m^{3.}$

5.1.1. The NaCl aerosol concentration will be determined on the days that initial penetration testing is performed by the following gravimetric method and calculated as milligrams per cubic meter ($mg/m^3$).

5.1.2. Weigh a 102 mm filter to the nearest 0.1 mg., mount in the gravimetric filter holder, subject it to the generated aerosol at 30 Lpm for 40 minutes and reweigh the filter. Use a timer to monitor the duration of the test. Record the pre- and post-weights, time, and average flow rate on the data sheet and calculate the aerosol concentration in $mg/m^3$ by the following formula:

UNCONTROLLED COPY FOR INSPECTION AND USE

UNCONTROLLED COPY FOR INSPECTION AND USE

$$\text{Concentration (C) in mg/m}^3 = \frac{(W2 - W1)}{(Q / 1000)(T)}$$

Where:

W1 = Initial filter weight in mg
W2 = Final filter weight in mg
Q = Flowrate in liters per minute
T = Test time in minutes

With a flowrate of 30 Lpm for 40 minutes, the above formula simplifies to:

$$C = \frac{W2 - W1}{1.2}$$

5.1.3. Use the following formula to calculate the test duration:

$$T \text{ in minutes} = \frac{(\text{mg load})(1000\ L / m3)}{(C)\ (Q)}$$

Where:

C = Concentration in mg/m3 from 5.1.2.
Q = Flow rate for test in Lpm

5.1.4. The upstream and downstream photometer readings are used for monitoring stability and for calculating a photometer correlation factor (CF). The correlation factor is determined with an empty filter holder and is calculated internally as shown below:

$$CF = \frac{\text{Downstream Photometer Voltage} - \text{Downstream Background Voltage}}{\text{Upstream Photometer Voltage} - \text{Downstream Background Voltage}}$$

The correlation factor is used by the software to express the upstream photometer signal in terms of the downstream photometer signal.

5.1.5. The NaCl particle size distribution shall be verified using “green line” filter discs supplied by TSI with a known penetration range. Graphs of penetration vs. resistance for two sheets and five sheets of stacked filter discs are supplied with each lot of the standard filters, with a central line and upper and lower lines representing the expected penetration range at a given resistance. The test data should fall within an acceptance zone having boundaries defined by the upper and lower curves on the graphs. The standard filter test using both 2 sheets and 5 sheets will be run at least once in each 8 hour test period to verify that the aerosol distribution is within the acceptance zone.

5.2. Respirator filters will be pre-conditioned at 85 ± 5% relative humidity and 38 ±2.5°C for

25 ±1 hours. After conditioning, the filters shall be sealed in a gas tight container and tested within 10 hours.

5.3. Filters will be mounted and sealed on holders to prevent leakage around the filter holder. Single air purifying respirator filters will be tested at a challenge flow rate of 85± 4 Lpm. Filters used as pairs on a respirator are tested using a single filter of the pair at 42.5 ± 2 Lpm challenge flow rate. Filters used in threes are tested using a single filter of the set at 28.3 ± 1 Lpm challenge flow rate.

5.3.1. The challenge flow rate must be checked for stability for at least 30 seconds prior to testing.

5.4. A sample of 20 filter units will be tested against the NaCl aerosol. Three filters will be loaded until the aerosol mass loading levels as shown in the table below are reached and evaluated to determine the method for the remaining 17 filters. This is the mass amount of NaCl aerosol that has contacted the filter.

| Number of Filters In Respirator Configuration | Aerosol Mass Loading Level |
|---|---|
| Single | 200 ± 5 mg. |
| Double | 100 ± 5 mg. |
| Triple | 66.7 ± 5 mg. |

5.4.1. Type 1. If preliminary testing of all three initial test filters consistently results in a straight line (Figure 3), for the remaining 17 filters, record the initial penetration reading.

5.4.2. Type 2. If filter testing of all three initial test filters consistently results in a curve which indicates increased efficiency during the complete run (Figure 3), for the remaining 17 filters, record the initial penetration reading.

5.4.3. Type 3. If filter testing of all three initial test filters consistently results in decreased efficiency over time (Figure 3), load the remaining 17 filters with NaCl to the level specified in the table above and record the maximum penetration reading.

5.4.4. Type 4. If filter testing of all three initial test filters consistently results in increased efficiency, then a decrease in efficiency, and then flattens out during the remainder of the complete run (Figure 3), for the remaining 17 filters, record the maximum penetration reading after reaching and maintaining a flat line for a period of 20 minutes following the decreasing segment in efficiency.

5.4.5. For any other filter type, determine loading at which maximum penetration consistently occurs and test at that loading value for the remaining 17 filters.

5.4.6. If any one of the 20 filters have a penetration greater than 0.030%, further testing of that filter will be terminated. Any filter that exceeds the specified limit shall

UNCONTROLLED COPY FOR INSPECTION AND USE

be remounted and retested to ensure that leakage was not caused by a mounting leak If retesting eliminates the excessive leakage and testing has gone beyond the initial penetration, that sample will be considered an invalid sample, and another tested in its place.

5.5. The penetration of the first three filters will be measured, recorded, and printed at approximately 1-minute intervals during the test period. The highest penetration observed throughout the test of each filter will be recorded as the maximum penetration of that filter.

5.6. Determine and record on the data sheet the maximum filter penetration for each of the 20 filters.

6. PASS/FAIL CRITERIA

6.1. The requirement for passing this test is set forth in 42 CFR, Part 84, Subpart K, Section 84.180.

6.2. The minimum efficiency for each of the 20 filters shall be determined and recorded and shall be equal to or greater than 99.97 %.

6.3. For the sample of 20 filters or filter cartridges to demonstrate acceptable performance, each filter shall meet or exceed the specified minimum efficiency level at the end point of the test.

7. RECORDS/TEST SHEETS

7.1. Record the test data in a format that shall be stored and retrievable.

8. ATTACHMENTS

8.1. Filtration Efficiency versus Time Example Plots

8.2. Example Data Sheet

8.3. Photograph of TSI 8130 CertiTester with chuck open

8.4. Photograph of TSI 8130 CertiTester with the chuck closed

UNCONTROLLED COPY FOR INSPECTION AND USE

8.1. Filtration Efficiency versus Time Example Plots

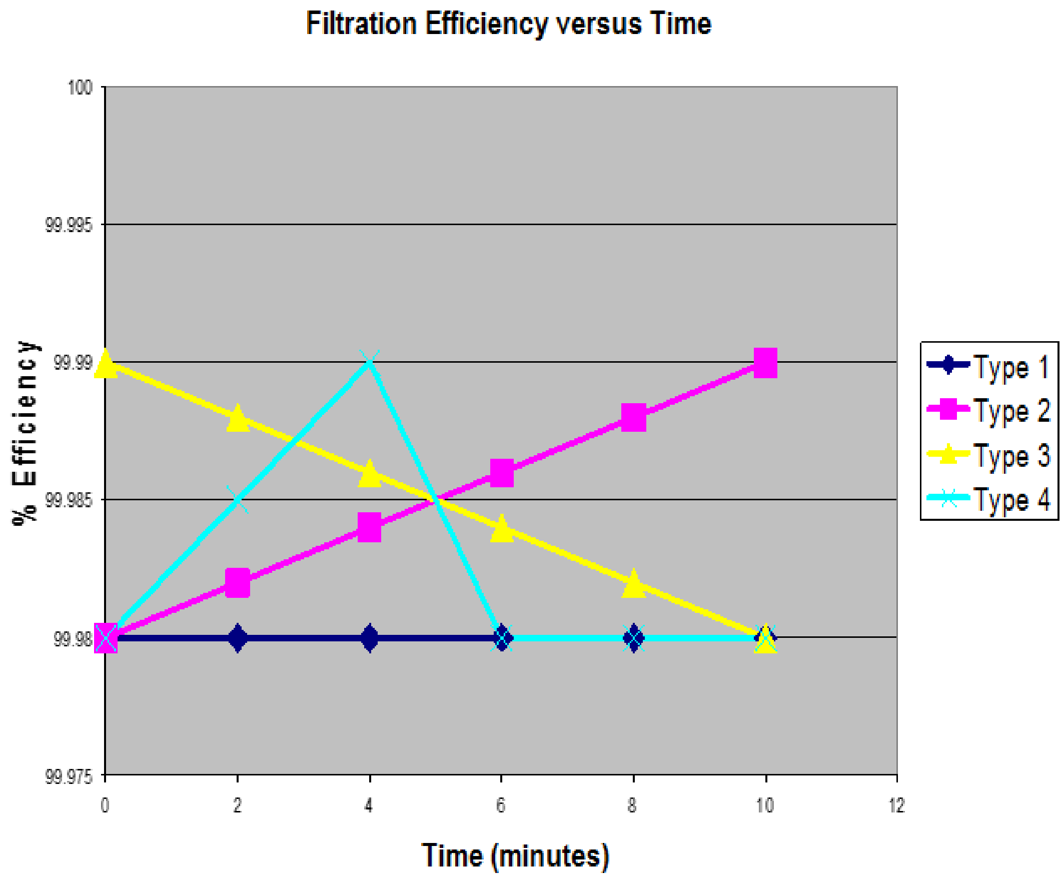

UNCONTROLLED COPY
FOR INSPECTION AND USE
UNCONTROLLED COPY
FOR INSPECTION AND USE

8.2. Example Data Sheet

**National Institute for Occupational Safety and Health**
**Respirator Branch**
**Test Data Sheet**

**Task Number:** TN-XXXXX
**Reference No.:** CFR 84.180
**Test:** Sodium Chloride (NaCl) PAPR100-N
**STP No.:**
**Manufacturer:** Company Name
**Item Tested:**

| Filter | Flow Rate | Initial Filter Resistance | Maximum Allowable Percent Leakage | Initial Percent Leakage | Maximum Percent Leakage | Result |
|---|---|---|---|---|---|---|
| 1 | 85 | 7.9 | 0.03 | .002 | .002 | PASS |
| 2 | 85 | 8.2 | 0.03 | .002 | .002 | PASS |
| 3 | 85 | 8.1 | 0.03 | .001 | .003 | PASS |
| 4 | 85 | 7.8 | 0.03 | .003 | .004 | PASS |
| 5 | 85 | 7.8 | 0.03 | .001 | .003 | PASS |
| 6 | 85 | 8.3 | 0.03 | .002 | .002 | PASS |
| 7 | 85 | 8.2 | 0.03 | .001 | .002 | PASS |
| 8 | 85 | 8.1 | 0.03 | .002 | .003 | PASS |
| 9 | 85 | 8.2 | 0.03 | .001 | .002 | PASS |
| 10 | 85 | 8.2 | 0.03 | .001 | .002 | PASS |
| 11 | 85 | 8.3 | 0.03 | .002 | .003 | PASS |
| 12 | 85 | 7.8 | 0.03 | .001 | .001 | PASS |
| 13 | 85 | 7.9 | 0.03 | .001 | .001 | PASS |
| 14 | 85 | 8.3 | 0.03 | .002 | .002 | PASS |
| 15 | 85 | 8.2 | 0.03 | .001 | .002 | PASS |
| 16 | 85 | 7.9 | 0.03 | .002 | .002 | PASS |
| 17 | 85 | 8.2 | 0.03 | .002 | .003 | PASS |
| 18 | 85 | 7.9 | 0.03 | .001 | .002 | PASS |
| 19 | 85 | 8.2 | 0.03 | .000 | .002 | PASS |
| 20 | 85 | 7.8 | 0.03 | .002 | .003 | PASS |

**Overall Result: PASS**

**Signature:** ___________________________ **Date:**____________________
**Engineering Technician**

UNCONTROLLED COPY FOR INSPECTION USE

UNCONTROLLED COPY FOR INSPECTION AND USE

8.3. Photograph of TSI 8130 with chuck open

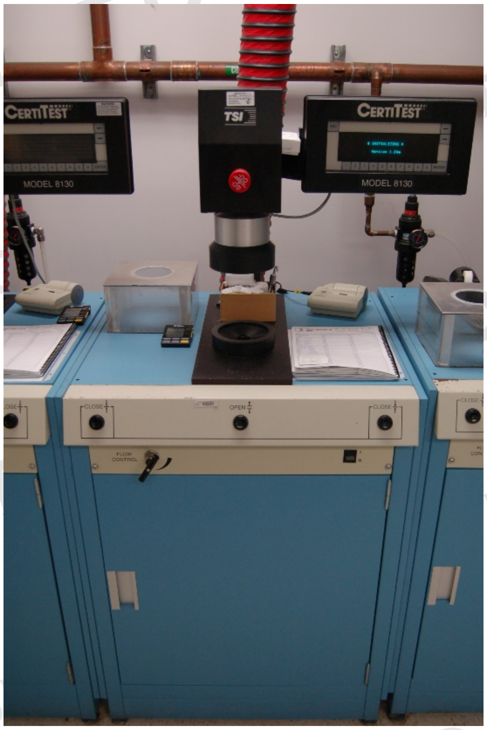

8.4. Photograph of TSI 8130 with chuck closed

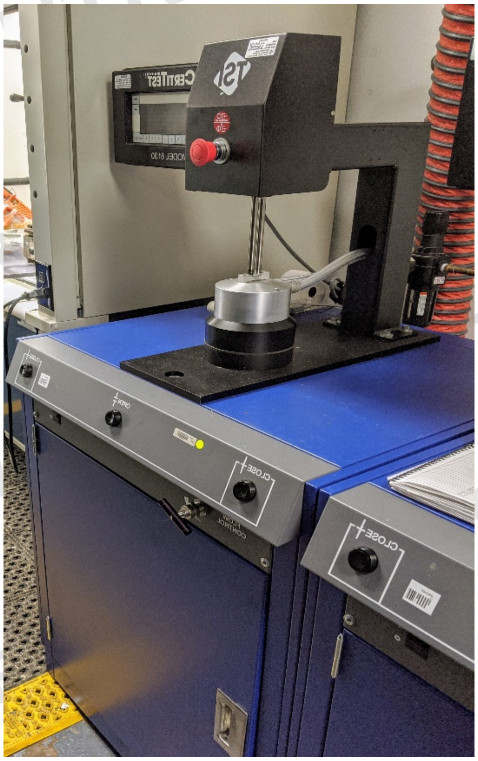

### Revision History

| Revision | Date | Reason for Revision |
|---|---|---|
| 0.0 | 23 March2020 | Original release |

UNCONTROLLED COPY FOR INSPECTION AND USE

UNCONTROLLED COPY FOR INSPECTION AND USE

DETERMINATION OF LOW FLOW WARNING DEVICE SOUND LEVEL ON POWERED AIR PURIFYING RESPIRATOR (PAPR), SERIES PAPR100, STANDARD TESTING PROCEDURE (STP)

1. PURPOSE

This test establishes the method for ensuring that the level of protection provided by the Low Flow Warning Devices Sound Level, series PAPR100, meet the minimum requirements set forth in 42 CFR, Part 84, Subpart K, Section 84.171(j)(6).

2. GENERAL

This STP describes the Determination of Low Flow Warning Device Sound Level on PAPR100s test procedure in sufficient detail that a person knowledgeable in the appropriate technical field can select equipment with the necessary resolution, conduct the test, and determine whether or not the product passes the test.

3. EQUIPMENT/MATERIAL

3.1. The list of necessary test equipment and materials follows:

3.1.1. Noise Dosimeter-- Quest Technologies Noise Pro Series Dosimeter For OSHA use, the dosimeter must have a 5 dB exchange rate, use a 90 dBA criterion level, be set at slow response, and use either an 80 dBA or 90 dBA threshold gate, or a dosimeter that has both capabilities, whichever is appropriate for the evaluation.

3.1.2. Lifesize mannequin.

4. TESTING REQUIREMENTS AND CONDITIONS

4.1. Prior to beginning any testing, confirm that all measuring equipment employed has been calibrated in accordance with the testing laboratory's calibration procedure and schedule. All measuring equipment utilized for this testing must have been calibrated using a method traceable to recognized international standards when available.

4.2. Noise level test must be performed in a location that has a maximum background noise level of no more than 60 dBA.

5. PROCEDURE

5.1. Position the microphones of the Quest NoisePro Dosimeter on each ear of the mannequin. Following the respirator manufacturer's instructions, mount the respirator assembly onto the mannequin.

5.2. Turn PAPR system on and follow the low flow user check, in user manual instructions, to activate the audible alarm.

5.3. Take and record five measurements at the mannequin's ear locations.

5.4. Data Analysis

5.4.1. Average the five readings taken from the left ear.

5.4.2. Average the five readings taken from the right ear.

5.4.3. Average the Left Ear Average and Right Ear Average to get the Overall Average.

6. PASS/FAIL CRITERIA

6.1. The requirement for passing this test is set forth in 42 CFR Part 84, Subpart K, Section 84.171(j)(6).

6.2. If the warning provided is audible only, the minimum sound level must be 80 dBA.

6.3. This test should be done on a minimum of two respirators.

7. RECORDS/TEST SHEETS

7.1. All test data collected will be recorded on the appropriate Determination of Minimum Sound Level for Low Flow Warning Device on PAPR100 Test Data Sheet.

8. ATTACHMENTS

8.1. Example Data Sheet

8.2. Photograph – Mannequin Wearing Sound Meters

UNCONTROLLED COPY FOR INSPECTION AND USE

UNCONTROLLED COPY FOR INSPECTION USE

8.1. Example Data Sheet

**Determination of Minimum Sound Level for Low Flow Warning Device "On"**
**PAPR100 Test Data Sheet**

**Project No.:** ______________________ Date: __________

**Company:** ______________________

**Respirator Type:** ______________________

**Reference:** 42 CFR, Part 84, Subpart K, Section 84.171(j)(6)

**Requirement:** The average sound level at both ears must be greater than 80 dBA.

**Procedure:** The respirator is mounted on a mannequin. Five sound level measurements are taken at each ear and averaged. The results for the left and right ears are then averaged to arrive at an overall test average.

**Results:**
**Background Noise:**__________**dBA**

| Unit # 1: | Left Ear\dBA | Right Ear\dBA | Unit # 2: | Left Ear\dBA | Right Ear\dBA |
|---|---|---|---|---|---|
| 1. | _______ | _______ | | _______ | _______ |
| 2. | _______ | _______ | | _______ | _______ |
| 3. | _______ | _______ | | _______ | _______ |
| 4. | _______ | _______ | | _______ | _______ |
| 5. | _______ | _______ | | _______ | _______ |

**Left Ear Average:** __________ __________

**Right Ear Average:** __________ __________

**Overall Average:** __________ __________

**Comments:**

______________________________________________

______________________________________________

______________________________________________

**Test Engineer:** ______________________ **PASS**________ **FAIL**________

8.2. Photograph of Sound Test Mannequin Wearing Sound Level Meters

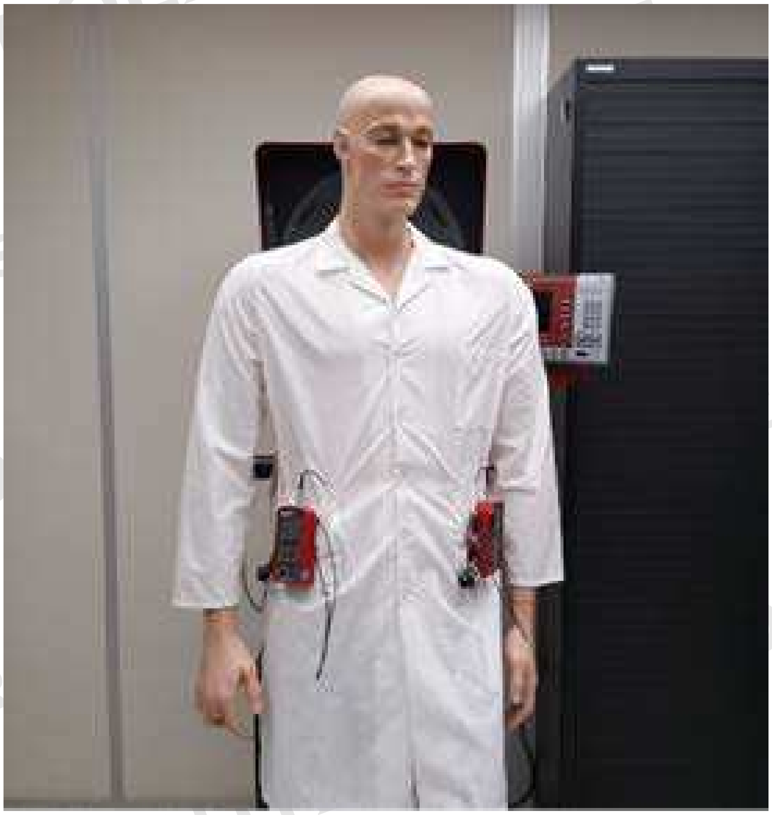

UNCONTROLLED COPY
FOR INSPECTION AND USE
UNCONTROLLED COPY
FOR INSPECTION AND USE
UNCONTROLLED COPY
FOR INSPECTION AND USE

### Revision History

| Revision | Date | Reason for Revision |
| --- | --- | --- |
| 0.0 | 23 March 2020 | Original release |

UNCONTROLLED COPY FOR INSPECTION AND USE
UNCONTROLLED COPY FOR INSPECTION AND USE
UNCONTROLLED COPY FOR INSPECTION AND USE
UNCONTROLLED COPY FOR INSPECTION AND USE

UNCONTROLLED COPY FOR INSPECTION AND USE

### DETERMINATION OF LOW FLOW WARNING DEVICE ACTIVATION FOR POWERED AIR-PURIFYING RESPIRATORS, SERIES PAPR100, STANDARD TESTING PROCEDURE (STP)

#### 1. PURPOSE

This procedure establishes the method for ensuring that the level of protection provided by the low flow warning device requirement for powered air-purifying respirators, series PAPR100, meet the requirements set forth in 42 CFR Part 84, Subpart K, 84.171(j).

#### 2. GENERAL

This procedure describes the Determination of Low Flow Warning Device Activation for Powered Air-Purifying Respirators, Series PAPR100, test procedure in sufficient detail that a person knowledgeable in the appropriate technical field can select equipment with the necessary resolution, conduct the test, and determine whether or not the product passes the test.

#### 3. EQUIPMENT/MATERIAL

3.1. Air-tight chamber – approximately 24 inches by 24 inches by 16 inches with a bolt on door, a 3-inch diameter inlet for accepting breathing tubes and adapter, a 1 inch diameter outlet, and a ¼ inch outlet for a manometer probe.

3.2. Setra Datum 2000 Model 239 digital manometer with an accuracy of ± 0.023 in-H2O or better, or equivalent.

3.3. Teledyne Hastings L-25S Laminar Mass Flow Meter, or equivalent.

3.4. Spencer 075-1/3 Centrifugal Blower vacuum source, or equivalent.

3.5. Anthropometric Headform, in accordance with ISO 16900, size medium, or equivalent.

#### 4. TESTING REQUIREMENTS AND CONDITIONS

4.1. Prior to beginning any testing, confirm that all measuring equipment employed has been calibrated in accordance with the testing laboratory's calibration procedure and schedule. All measuring equipment utilized for this testing must have been calibrated using a method traceable to recognized international standards when available.

4.2. Testing shall be conducted on as-received respirators at 25 ± 2.5 degrees Celsius (°C) and adhere to User's Instructions.

4.3. Three complete respirator systems will be evaluated for low flow warning device activation.

4.4. Determination of Low Flow Warning Device Visibility will be completed for any respirator with a visual warning device. The standard testing procedure is described in CVB-APR-STP-0087.

4.5. Determination of Low Flow Warning Device Sound Level on Series PAPR100 will be completed for any respirator with an audible warning device, where the warning provided is audible only, or other warnings are not readily apparent. The standard testing procedure is described in CVB-APR-STP-0085.

5. PROCEDURE

5.1. Prior to any testing, the respirator shall be evaluated per the user check instructions in the User's Manual to ensure that the low flow warning(s) activate as designed.

5.2. Test setup for low flow warning device activation for continuous flow powered air-purifying respirators

5.2.1. The test chamber outlet is connected to the mass flow meter inlet. The mass flow meter outlet is connected to the vacuum blower inlet. A flow control valve is placed between the mass flow meter and vacuum blower. The vacuum blower vents to atmosphere. Ensure that all pipe lengths are sufficient to maintain laminar flow.

5.2.2. Mount the respirator to the headform in the as-worn configuration following all pertinent User's Instructions.

5.2.3. Connect the headform and respirator to the chamber.

5.2.3.1. On units with breathing tubes, the breathing tube is placed through the chamber inlet port and the blower is placed outside of the chamber. The headform and respirator are placed inside the chamber. Seal the chamber inlet around the breathing tube.

5.2.3.2. On units without breathing tubes, the headform trachea outlet tube is placed through the chamber inlet port and the headform and respirator are placed outside the chamber. Seal the chamber inlet around the headform outlet tube.

5.2.3.2.1. Loose fitting respiratory inlet coverings may be adjusted to capture air-flow that would otherwise exit the respirator.

5.2.4. Connect the digital manometer to the pressure tap on the headform. Pressure is measured at a pitot ring positioned 25 mm inside of the trachea inlet.

5.2.4.1. Ensure the manometer has a reading of zero at ambient conditions, and adjust if necessary.

UNCONTROLLED COPY FOR INSPECTION AND USE

5.2.5. Close the chamber door and ensure the chamber is sealed.

5.2.6. Turn on the PAPR and the vacuum blower. Ensure that no air flow warnings are present on the respirator.

5.2.7. Adjust the flow control valve until manometer has a reading of zero.

5.2.8. Restrict the flow to the respirator. This is done by incrementally adding restriction to the inlet of the respirator, such as attaching small pieces of adhesive tape or a similar flow restricting item.

5.2.8.1. While restricting flow to the respirator, adjust the flow control valve to ensure that the manometer continuously reads zero.

5.2.9. Add increased flow restriction to the respirator inlet until the low flow warning device activates. Record the maximum airflow which activates the low flow warning device.

5.2.10. Turn off the PAPR100 and vacuum blower.

#### 6. PASS/FAIL CRITERIA

6.1. The criterion for passing this test is set forth in 42 CFR Part 84, Subpart K, Section 81.171(j).

6.1.1. The low flow warning must actively and readily indicate when flow inside the respiratory inlet covering falls below the minimum required air flow. The minimum air flow shall be 115 LPM for tight-fitting PAPR100 and 170 LPM for loose-fitting PAPR100.

6.1.2. Any warning must be detectable by the wearer without any intervention by the wearer.

6.1.3. Warning devices must be configured so that they may not be de-energized while the blower is energized.

6.1.4. Any warnings which require different reactions by the wearer must be distinguishable from one another.

#### 7. RECORDS/TEST SHEETS

7.1. Record the test data in a format that shall be stored and retrievable.

UNCONTROLLED COPY FOR INSPECTION AND USE

### Revision History

| Revision | Date | Reason for Revision |
|---|---|---|
| 0.0 | 23 March 2020 | Original Release |

UNCONTROLLED COPY FOR INSPECTION AND USE

UNCONTROLLED COPY FOR INSPECTION AND USE

DETERMINATION OF COMMUNICATION PERFORMANCE TEST FOR SPEECH CONVEYANCE AND INTELLIGIBILITY OF POWERED AIR-PURIFYING RESPIRATOR (PAPR) SERIES PAPR100 STANDARD TESTING PROCEDURE

1. PURPOSE

1.1. This test establishes the method for ensuring that the level of speech conveyance and intelligibility provided by the Communication Performance Test on Powered Air-Purifying Respirator (PAPR) Series PAPR100 meet requirements set forth in 42 CFR Part 84, Subpart K, Section 84.181.

1.2. The purpose of this test is to quantify the performance of a respirator in transmitting intelligible speech of a human test subject. This is accomplished by determining a listener's ability to comprehend single-syllable words spoken by a subject wearing the respirator.

2. GENERAL

2.1. This STP describes the Communication Performance Tests for Speech Conveyance and Intelligibility of a PAPR100 in sufficient detail that a person knowledgeable in the appropriate technical field can select equipment with the necessary resolution, conduct the test, and determine whether or not the product passes the test.

2.2. The Communication Performance Test shall be performed using the Modified Rhyme Test (MRT) to evaluate a listener's ability to comprehend single-syllable words from a speaker. Both listeners and speakers shall be in combinations of masked and unmasked postures during the MRT to provide an indication of speech transmission and intelligibility.

2.3. This test is considered a human factors test that requires participation of 8 test subjects to quantify the overall performance rating of a PAPR100 system.

2.4. Two (2) Test Administrators are necessary to properly conduct this test.

3. EQUIPMENT/MATERIALS

3.1. Laboratory space that permits an unobstructed 10 +1/-0 ft distance between test speaker and the listener group when facing one another. Refer to Figure 1. for the subject and equipment positioning for the Modified Rhyme Test setup.

3.2. Noise Plug, Precision Pink Noise Test Generator (GTC Industries) or equivalent for producing pink noise in the frequency range of 20.0 Hz to 50.0 kHz. Accuracy: 3dB per octave rolloff from 20 Hz to 20 KHz. Pink noise is defined as an audio test signal that contains all the frequencies in the audio spectrum with equal energy at each octave. Pink noise contains less energy at the higher audio frequencies than at the lower ones - See Figure 1.

3.3. A stereo amplifier used to transmit and amplify the signal for the pink noise background.

3.4. The two loudspeakers positioned midway between the test speaker and listeners. Details of the exact positioning of the speaker equipment are further defined in Section 5.3.2.

3.5. Two Type 2 digital sound level meters (Sper Scientific, LTD, Model 840029 – See Figure 3.) with an "A" weighting decibel scale of 30 to 130 dB or equivalent. One sound level meter will be positioned in front of the Test Speaker and the second sound level meter will be positioned at head level beside listener L2- see Figure 1.

3.6. Sper Scientific, LTD Acoustical calibrator (model 840031) or equivalent is used to calibrate the sound level meters – See Figure 4.

3.7. Twelve word lists from the Modified Rhyme Test (MRT). These twelve word lists are separated onto individual sheets to be used by the test speakers during the test – See Appendix A.

3.8. Test listeners' multiple-choice answer pages on laptops or equivalent. Modified Rhyme Test listener responses are recorded and scored using laptops that have been programmed for this application; otherwise, the manual method using multiple-choice answer sheets can be used – See Example Data Sheet 8.1.

3.9. Sound and/or audiovisual recording device for recording speech.

3.10. <u>Human Test Subjects</u>

3.10.1. Test Administrators shall have successfully completed the CDC/ATSDR Scientific Ethics Training, the DHHS/NIH Human Participant Protections Education for Research Teams, or equivalent course.

3.10.2. At least eight (8) volunteer test subjects are required for this test.

#### 4. <u>TESTING REQUIREMENTS AND CONDITIONS</u>

4.1. Prior to beginning any testing, confirm that all measuring equipment being used has been calibrated in accordance with the testing laboratory's calibration procedure and schedule. All measuring equipment utilized for this testing must have been calibrated using a method traceable to recognized international standards when available.

4.2. Administrator 1 will monitor the test speakers' speech and the actions of the listener group to make sure they are responding to the test speaker.

UNCONTROLLED COPY FOR INSPECTION AND USE

4.3. Administrator 2 will monitor the dBA level of the test speaker and give verbal or nonverbal feedback as to loudness of his/her speech and record the dBA level on the data sheet – see Example Data Sheet 8.2.

4.4. The eight test subjects will be divided into two groups, a listener group comprised of three subjects (at least one female and one male) and a speaker group comprised of five subjects (at least one female and one male).

4.5. All subjects shall be fluent in English. In addition, these subjects shall have no obvious or strong regional or foreign accents.

4.6. The eight test subjects shall be trained in the donning and use of the respirator per manufacturer's instructions by the test administrator.

4.7. Each subject shall be sized and fitted for the respirator per manufacturer's instructions. Where an individual is qualified to wear multiple sizes of the respirator, the subject shall select the respirator size that provides the most comfortable fit.

4.8. The subjects shall have normal hearing.

4.9. The test subjects shall not have any facial hair or conditions that may cause interference with the seal of the respirator facepiece.

4.10. Test Equipment Set-Up.

4.10.1. Room selected to perform the MRT shall be of ample size to comfortably house all the test equipment indicated and the personnel associated with the test. It shall be free of external noise interference.

4.10.2. The loudspeakers shall be positioned opposite each other at a height of approximately 29.5 inches from the floor and 9 +0.5/-0 ft from the center of the sound field between the test speaker and listeners. The laptops will be placed directly in front of the listeners at a convenient distance away – See Figure 1.

5. <u>PROCEDURE</u>

5.1. Fill in the Pretest data on the data sheet – See Example Data Sheet 8.1.

5.2. Record the background noise of the room, assuring no external noise, at the center listener's head position.

5.3. Training of the Test Speakers.

5.3.1. Without the listeners in the room, one subject at a time will be seated in the test speaker's position and given an MRT word list – See Appendix A. A sound level meter will be positioned in front of the test speaker to assess the volume of their voice as the word list is read. The sound level meter shall be set to display "A"-weighted sound levels.

UNCONTROLLED COPY FOR INSPECTION AND USE

UNCONTROLLED COPY
FOR INSPECTION AND USE

5.3.2. Test administrator 1 shall be seated beside the test speaker to monitor the reading of the word list. The administrator shall instruct the test speaker to read, without placing any unusual emphasis on any stimulus word, at a rate of approximately one phrase every 6 seconds, using the introductory phrase "The word is (list word)". In addition, the test speaker shall be directed to communicate each word without using any visual gestures, such as hand signals, and without repeating any of the list words.

5.3.3. Test administrator 2 shall be seated in the center test listener's position and will instruct the test speaker to begin reading the word list at a voice level of 75 to 85 dBA.

5.3.4. During the reading of the word list, test administrator 2 will monitor the "A"-weighted output volume of the speaker and provide feedback as to the loudness of the speaker's performance throughout the reading of the word list. Test administrator 1 will provide feedback to the test speaker regarding the pronunciation of the list words and the rate of performance. Additional training will be done if warranted per the judgment of the test administrators. These procedures will be completed without the use of a respirator and without any background noise above ambient conditions.

5.3.5. Additionally, training with background noise of 60 ± 2 dBA (consisting of pink noise), will also be completed according to the above procedures.

5.3.6. This will be repeated for all subjects in the speaker group.

5.4. Training of the Test Listeners.

5.4.1. The three test listeners are seated facing a single test speaker at a distance of 10 ft. The listeners are seated next to one another with approximately one foot between them. Each listener will be given a laptop with a list of multiple-choice words for recording his or her responses.

5.4.2. An MRT word list will be provided to a trained test speaker. The test speaker will be instructed to communicate the word list to the test listeners as was done during speaker training.

5.4.3. As a group, listeners will be instructed to listen attentively as the test speaker reads 50 words to them each with the introductory phrase "The word is (list word)." The test listeners shall be directed to select, or make a best guess of the word, that was perceived to be spoken from the six possible response words provided to them on the laptops. Test listeners are instructed to provide a "thumbs-up" hand signal to the speaker as a cue to say the next phrase.

5.4.4. During the reading of the word list, administrator 2 will monitor the A-weighted output volume of the speaker at the listening position and provide feedback as to the loudness throughout the reading of the word list. In addition, test administrator 1 will provide the test speaker feedback regarding the rate of performance. Administrator 1 will also monitor the actions of the listener group and provide additional instructions as needed. Test administrators will determine if additional training is required. These procedures will be completed without the use of a respirator and without any added background noise (pink noise) above ambient conditions.

5.4.5. Additionally, training with added background noise of 60 ± 2 dBA (consisting of pink noise), will also be conducted according to the procedures above.

5.5. Conducting the Test.

5.5.1. The noise generator shall be turned on and set to produce 60 ±2 dBA of pink noise, as measured by the sound level meter at the center test listener's head position without listeners present. This value shall be recorded on the Respirator Communication Performance Test Data Collection Sheet.

5.5.2. The three test listeners shall be seated in the listening position with a laptop located conveniently in front of them.

5.5.3. A test speaker will then be positioned in the speaking position and present one complete MRT word list to the listening panel. A different test speaker shall then be used to present the next MRT trial. Test speakers will continue to rotate among the speaker test panel until all trials have been completed.

5.5.4. Data will be obtained without the respirator (unmasked) and with the respirator (masked) worn and operated per the manufacturer's instructions by both speakers and listeners. All conditions shall be randomly assigned, and a different word list shall be used for each test. A test matrix of the MRT conditions is provided in Table 2 to assist the test administrator in establishing the sequence of testing.

5.5.5. Background noise levels shall be monitored at the center test listener's head position and recorded, on the Respirator Communication Performance Test Data Collection Sheet, at the beginning, middle, and end of each MRT session by administrator 2.

5.5.6. The test speakers shall be monitored and recorded during the test to determine if the test speakers conform to the word list specified for each trial by test administrator 1. Test administrator 1 shall also make note of any improperly pronounced or misspoken words by the test speakers.

5.5.7. A total of 10 MRT trials shall be performed; 5 unmasked and 5 masked. This will result in a total of 15 MRT scores (five per listener) for the unmasked condition and 15 scores for the masked condition.

5.6. Data Analysis.

UNCONTROLLED COPY FOR INSPECTION AND USE

5.6.1. Use the Respirator Communication Performance Test Data Collection Sheets to assist with the analysis of the 10 MRT scores for each listener.

5.6.2. The number of correct responses shall be adjusted for chance or guessing made possible by the multiple-choice answers using the equation:

$$\text{Adjusted Score} = \text{Number of Correct Listener Responses} - \frac{\text{Number of Wrong Listener Responses}}{5}$$

5.6.3. Determine the Number of Words Spoken Correctly by the speakers from either the Test Administrators notes or if necessary, by listening to the Audio recorder tapes.

5.6.4. Listener performance on the MRT shall be scored in terms of the percentage of words correctly identified using the equation for both the masked and unmasked posture:

% Correct = (Adjusted Score/Number of Words Spoken Correctly) * 100

5.6.5. Average Unmasked and Average Masked % Correct Scores shall be calculated for each individual listener.

5.6.6. Each individual listener's Average Masked % Correct Score shall be divided by their Average Unmasked % Correct Score to calculate a Performance Rating using the equation:

5.6.7. Performance Rating (%) = $\left(\frac{\text{Average Masked \% Correct Score}}{\text{Average Unmasked \% Correct Score}}\right) \times 100$

5.6.8. The performance rating of all listeners shall then be averaged to determine the Overall Performance Rating of the PAPR100 using the following Equation:

$$\text{Overall Performance Rating (\%)} = \frac{\text{Performance Rating (\%)}_{L1} + \text{Performance Rating (\%)}_{L2} + \text{Performance Rating (\%)}_{L3}}{3}$$

6. PASS/FAIL CRITERIA

6.1. The requirement for passing this test is set forth in 42 CFR, Part 84, Subpart K, Section 84.181.

6.2. A candidate PAPR100 must obtain an Overall Performance Rating greater than or equal to 70% to meet the Communication requirement.

7. RECORDS/TEST SHEETS

UNCONTROLLED COPY FOR INSPECTION AND USE

7.1. All test data shall be recorded on the Respirator Communication Performance Test Data Collection Sheets.

8. ATTACHMENTS

8.1. Example Data Sheet - Communication Performance Test Listener Data Sheet

8.2. Example Data Sheet - Communication Performance Test Data Collection Sheet

8.3. Table 1. MTR Trial Matrix

8.4. Figures

8.4.1. Figure 1. Modified Rhyme Test Setup

8.4.2. Figure 2. Photograph of Noise Plug, Precision Pink Noise Generator (GTC Industries)

8.4.3. Figure 3. Photograph of Sper Scientific LTD digital sound level meter, Model 840029

8.4.4. Figure 4. Photograph of Acoustical calibrator (Sper Scientific model 840031)

8.5. Appendix A. Communication Performance Word List Test Sheets

UNCONTROLLED COPY FOR INSPECTION AND USE

#### 8.1. Example Data Sheet – Communication Performance Test Listener Data Sheet

NIOSH Application Number ______________________ Date: __________

Respirator Manufacture: ______________________ ; MRT/Sheet# __________

Respirator Type: __________ ; Speakers Respirator Number, if worn __________ (Indicate if unworn)

Listener's Respirator (circle one): Unworn Worn If Worn, Listeners Respirator Number __________

Listener #:______ Listening Position:_ Speaker #:______ MRT Word List#:________

| | | | | | | | | | | | | | | | |
|---|---|---|---|---|---|---|---|---|---|---|---|---|---|---|---|
| 1 | but | bug | bus | 14 | map | mat | math | 27 | wed | fed | bed | 40 | cake | came | Cave |
| | buff | bun | buck | | mad | mass | man | | led | shed | red | | cane | case | cape |
| 2 | kin | kid | kick | 15 | hop | cop | shop | 28 | sane | sake | safe | 41 | fang | bang | hang |
| | king | kit | kill | | mop | pop | top | | save | same | sale | | sang | gang | rang |
| 3 | peak | peach | peas | 16 | sack | sad | sap | 29 | pit | pin | pig | 42 | law | saw | paw |
| | peal | peace | peat | | sag | sat | sass | | pill | pick | pip | | jaw | raw | thaw |
| 4 | dig | wig | big | 17 | say | pay | may | 30 | heel | peel | keel | 43 | rake | rate | ray |
| | fig | pig | rig | | gay | way | day | | feel | eel | reel | | raze | race | rave |
| 5 | fold | sold | gold | 18 | heath | heave | heap | 31 | toil | boil | foil | 44 | dip | dim | din |
| | hold | cold | told | | heat | heal | hear | | coil | oil | soil | | dill | did | dig |
| 6 | kick | lick | sick | 19 | tame | came | fame | 32 | fig | fizz | fit | 45 | tear | teal | teak |
| | tick | wick | pick | | same | name | game | | fib | fin | fill | | team | tease | teach |
| 7 | path | pack | pass | 20 | page | pane | pace | 33 | mark | bark | dark | 46 | tin | fin | sin |
| | pat | pad | pan | | pave | pale | pay | | lark | hark | park | | win | pin | din |
| 8 | beat | beak | beach | 21 | dust | gust | must | 34 | bash | bat | ban | 47 | seethe | seek | seen |
| | beam | bean | bead | | bust | just | rust | | back | bath | bad | | seed | seep | seem |
| 9 | pot | hot | lot | 22 | pun | puff | pup | 35 | will | hill | kill | 48 | run | bun | fun |
| | not | tot | got | | pub | pus | puck | | bill | fill | till | | sun | nun | gun |
| 10 | fit | hit | bit | 23 | then | den | ten | 36 | pale | sale | bale | 49 | neat | beat | seat |
| | sit | kit | wit | | pen | hen | men | | gale | male | tale | | meat | feat | heat |
| 11 | sup | sub | sud | 24 | cuss | cud | cup | 37 | duck | dud | dung | 50 | lip | hip | dip |
| | sum | sun | sung | | cut | cub | cuff | | dun | dug | dub | | sip | rip | tip |
| 12 | dent | tent | rent | 25 | hook | shook | book | 38 | sit | sip | sill | | **Score** | | |
| | went | sent | bent | | took | cook | look | | sick | sin | sing | | | | |
| 13 | best | west | nest | 26 | late | lake | lay | 39 | tack | tan | tab | | | | |
| | vest | test | rest | | lame | lane | lace | | tang | tam | tap | | | | |

#### 8.2. Example Data Sheet – Communication Performance Test Data Collection Sheet

**TN # :** ________________ **MRT Trial # :** _______________ **Speaker ID # :** ____________

Word List # : _______ Please Circle: Masked Unmasked

**Number of words correctly spoken by the MRT Speaker:** __________

**Listener #1 ID:** __________________

1. % of Words Answered Correctly = __________________
2. Adjusted Score = ______________________

**Listener #2 ID:** __________________

1. % of Words Answered Correctly = __________________
2. Adjusted Score = ______________________

**Listener #3 ID:** __________________

1. % of Words Answered Correctly = __________________
2. Adjusted Score = ______________________

Background Noise (w/o pink noise): ___________

Pink Noise Before Session: ___________

Pink Noise Middle of Session: ___________

Pink Noise After Session: ____________

UNCONTROLLED COPY FOR REFERENCE USE

8.3. Table 2. MRT Trial Matrix

| Trial # | Speaker | With or Without Mask | Word list |
|---|---|---|---|
| 1 | 1 | No mask | 11A |
| 2 | 2 | No mask | 3A |
| 3 | 3 | Masked | 5A |
| 4 | 4 | Masked | 7A |
| 5 | 5 | No mask | 4A |
| 6 | 2 | Masked | 12A |
| 7 | 4 | No mask | 5A |
| 8 | 1 | Masked | 1A |
| 9 | 5 | Masked | 8A |
| 10 | 3 | No mask | 6A |

8.4. Diagram 1. Modified Rhyme Test Setup

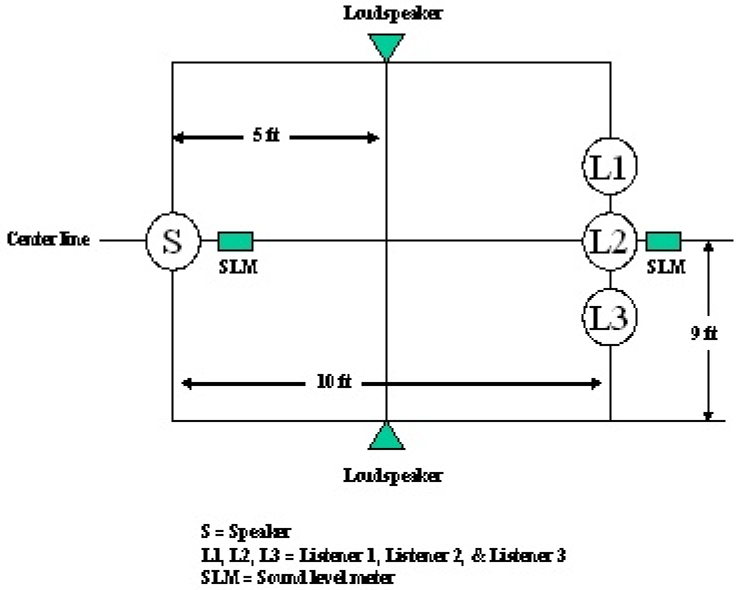

UNCONTROLLED COPY
FOR INSPECTION AND USE

8.5. Figure 1. Noise Plug, Precision Pink Noise Generator (GTC Industries)

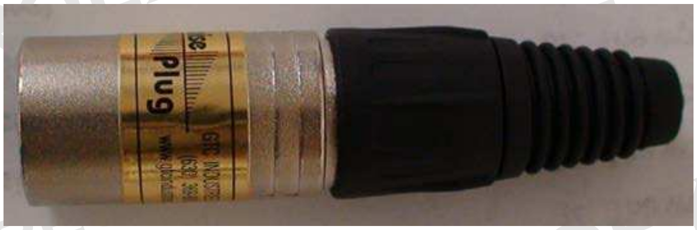

8.6. Figure 2. Sper Scientific LTD digital sound level meter, Model 840029

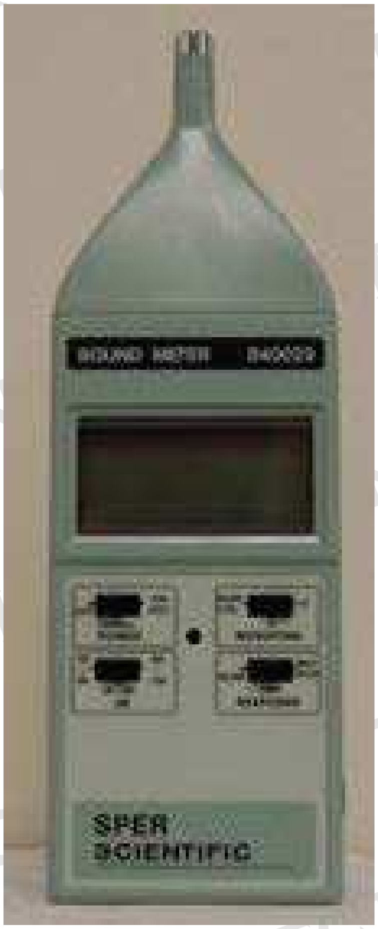

UNCONTROLLED COPY FOR INSPECTION AND USE

8.7. Figure 3. Acoustical calibrator (Sper Scientific model 840031)

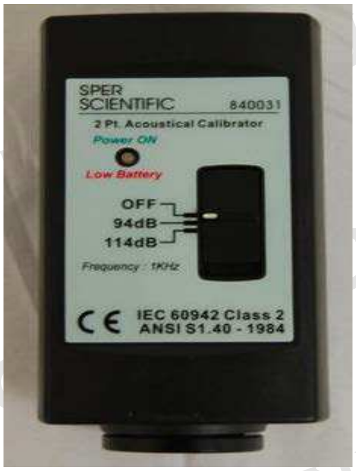

UNCONTROLLED COPY FOR INSPECTION AND USE

8.8. Appendix A. Communication Performance Word List Test Sheets

UNCONTROLLED COPY
FOR INSPECTION AND USE

### Appendix A

### Modified Rhyme Test Word Lists

UNCONTROLLED COPY FOR INSPECTION AND USE

**List 1A**

1. The word is **but**
2. The word is **kit**
3. The word is **peak**
4. The word is **pig**
5. The word is **cold**
6. The word is **sick**
7. The word is **pat**
8. The word is **beat**
9. The word is **hot**
10. The word is **fit**
11. The word is **sung**
12. The word is **sent**
13. The word is **rest**
14. The word is **mat**
15. The word is **top**
16. The word is **sack**
17. The word is **day**
18. The word is **heal**
19. The word is **name**
20. The word is **pay**
21. The word is **must**
22. The word is **pun**
23. The word is **hen**
24. The word is **cud**
25. The word is **book**
26. The word is **late**
27. The word is **led**
28. The word is **same**
29. The word is **pin**
30. The word is **feel**
31. The word is **soil**
32. The word is **fizz**
33. The word is **park**
34. The word is **bash**
35. The word is **till**
36. The word is **male**
37. The word is **dud**
38. The word is **sin**
39. The word is **tack**
40. The word is **case**
41. The word is **bang**
42. The word is **thaw**
43. The word is **ray**
44. The word is **dig**
45. The word is **team**
46. The word is **win**
47. The word is **seek**
48. The word is **bun**
49. The word is **feat**
50. The word is **sip**

**List 2A**

1. The word is **bus**
2. The word is **kick**
3. The word is **peas**
4. The word is **fig**
5. The word is **hold**
6. The word is **lick**
7. The word is **pass**
8. The word is **beach**
9. The word is **pot**
10. The word is **bit**
11. The word is **sub**
12. The word is **went**
13. The word is **west**
14. The word is **map**
15. The word is **cop**
16. The word is **sap**
17. The word is **pay**
18. The word is **heath**
19. The word is **tame**
20. The word is **pale**
21. The word is **gust**
22. The word is **pus**
23. The word is **then**
24. The word is **cuss**
25. The word is **cook**
26. The word is **lace**
27. The word is **bed**
28. The word is **save**
29. The word is **pit**
30. The word is **reel**
31. The word is **boil**
32. The word is **fib**
33. The word is **bark**
34. The word is **bad**
35. The word is **fill**
36. The word is **gale**
37. The word is **duck**
38. The word is **sit**
39. The word is **tap**
40. The word is **cane**
41. The word is **sang**
42. The word is **saw**
43. The word is **race**
44. The word is **dim**
45. The word is **teak**
46. The word is **din**
47. The word is **seethe**
48. The word is **run**
49. The word is **meat**
50. The word is **dip**

UNCONTROLLED COPY FOR INSPECTION AND USE

**List 3A**

1. The word is **bug**
2. The word is **kin**
3. The word is **peach**
4. The word is **rig**
5. The word is **gold**
6. The word is **tick**
7. The word is **pan**
8. The word is **beak**
9. The word is **got**
10. The word is **hit**
11. The word is **sup**
12. The word is **rent**
13. The word is **best**
14. The word is **math**
15. The word is **hop**
16. The word is **sad**
17. The word is **gay**
18. The word is **heap**
19. The word is **fame**
20. The word is **page**
21. The word is **bust**
22. The word is **puff**
23. The word is **ten**
24. The word is **cup**
25. The word is **took**
26. The word is **lake**
27. The word is **shed**
28. The word is **safe**
29. The word is **pig**
30. The word is **eel**
31. The word is **toil**
32. The word is **fill**
33. The word is **lark**
34. The word is **bath**
35. The word is **will**
36. The word is **tale**
37. The word is **dung**
38. The word is **sill**
39. The word is **tang**
40. The word is **cave**
41. The word is **rang**
42. The word is **jaw**
43. The word is **raze**
44. The word is **dip**
45. The word is **teal**
46. The word is **pin**
47. The word is **seem**
48. The word is **fun**
49. The word is **heat**
50. The word is **rip**

UNCONTROLLED COPY FOR INSPECTION AND USE

**List 4A**

1. The word is **bun**
2. The word is **kill**
3. The word is **peace**
4. The word is **big**
5. The word is **told**
6. The word is **kick**
7. The word is **pad**
8. The word is **bean**
9. The word is **lot**
10. The word is **kit**
11. The word is **sum**
12. The word is **bent**
13. The word is **vest**
14. The word is **man**
15. The word is **mop**
16. The word is **sat**
17. The word is **say**
18. The word is **hear**
19. The word is **game**
20. The word is **pave**
21. The word is **dust**
22. The word is **puck**
23. The word is **men**
24. The word is **cuff**
25. The word is **hook**
26. The word is **lane**
27. The word is **fed**
28. The word is **sale**
29. The word is **pip**
30. The word is **peel**
31. The word is **coil**
32. The word is **fit**
33. The word is **mark**
34. The word is **bat**
35. The word is **bill**
36. The word is **bale**
37. The word is **dub**
38. The word is **sing**
39. The word is **tam**
40. The word is **cape**
41. The word is **hang**
42. The word is **law**
43. The word is **rake**
44. The word is **dill**
45. The word is **tease**
46. The word is **fin**
47. The word is **seen**
48. The word is **gun**
49. The word is **seat**
50. The word is **hip**

UNCONTROLLED COPY FOR INSPECTION AND USE

UNCONTROLLED COPY FOR INSPECTION AND USE

**List 5A**

1. The word is **buck**
2. The word is **king**
3. The word is **peat**
4. The word is **dig**
5. The word is **fold**
6. The word is **wick**
7. The word is **pack**
8. The word is **bead**
9. The word is **not**
10. The word is **wit**
11. The word is **sun**
12. The word is **tent**
13. The word is **test**
14. The word is **mad**
15. The word is **pop**
16. The word is **sass**
17. The word is **way**
18. The word is **heat**
19. The word is **same**
20. The word is **pane**
21. The word is **just**
22. The word is **pup**
23. The word is **pen**
24. The word is **cut**
25. The word is **shook**
26. The word is **lay**
27. The word is **red**
28. The word is **sane**
29. The word is **pill**
30. The word is **keel**
31. The word is **oil**
32. The word is **fig**
33. The word is **hark**
34. The word is **ban**
35. The word is **hill**
36. The word is **pale**
37. The word is **dun**
38. The word is **sick**
39. The word is **tab**
40. The word is **cake**
41. The word is **fang**
42. The word is **raw**
43. The word is **rate**
44. The word is **did**
45. The word is **teach**
46. The word is **sin**
47. The word is **seed**
48. The word is **sun**
49. The word is **neat**
50. The word is **tip**

**List 6A**

1. The word is **buff**
2. The word is **kid**
3. The word is **peal**
4. The word is **wig**
5. The word is **sold**
6. The word is **pick**
7. The word is **path**
8. The word is **beam**
9. The word is **tot**
10. The word is **sit**
11. The word is **sud**
12. The word is **dent**
13. The word is **nest**
14. The word is **mass**
15. The word is **shop**
16. The word is **sag**
17. The word is **may**
18. The word is **heave**
19. The word is **came**
20. The word is **pace**
21. The word is **rust**
22. The word is **pub**
23. The word is **den**
24. The word is **cub**
25. The word is **look**
26. The word is **lame**
27. The word is **wed**
28. The word is **sake**
29. The word is **pick**
30. The word is **heel**
31. The word is **foil**
32. The word is **fin**
33. The word is **dark**
34. The word is **back**
35. The word is **kill**
36. The word is **sale**
37. The word is **dug**
38. The word is **sip**
39. The word is **tan**
40. The word is **came**
41. The word is **gang**
42. The word is **paw**
43. The word is **rave**
44. The word is **din**
45. The word is **tear**
46. The word is **tin**
47. The word is **seep**
48. The word is **nun**
49. The word is **beat**
50. The word is **lip**

UNCONTROLLED COPY FOR INSPECTION AND USE

UNCONTROLLED COPY FOR INSPECTION AND USE

**List 7A**

1. The word is **lick**
2. The word is **beat**
3. The word is **puff**
4. The word is **cook**
5. The word is **tip**
6. The word is **rave**
7. The word is **hang**
8. The word is **till**
9. The word is **math**
10. The word is **sale**
11. The word is **same**
12. The word is **peal**
13. The word is **kit**
14. The word is **sat**
15. The word is **sin**
16. The word is **gold**
17. The word is **buff**
18. The word is **lay**
19. The word is **nun**
20. The word is **must**
21. The word is **pad**
22. The word is **din**
23. The word is **sit**
24. The word is **win**
25. The word is **teak**
26. The word is **dent**
27. The word is **sub**
28. The word is **led**
29. The word is **tot**
30. The word is **dub**
31. The word is **pip**
32. The word is **seen**
33. The word is **way**
34. The word is **west**
35. The word is **pace**
36. The word is **bat**
37. The word is **mop**
38. The word is **big**
39. The word is **tab**
40. The word is **case**
41. The word is **name**
42. The word is **soil**
43. The word is **fin**
44. The word is **cuff**
45. The word is **heal**
46. The word is **hark**
47. The word is **heat**
48. The word is **then**
49. The word is **law**
50. The word is **bean**

**List 8A**

1. The word is **wick**
2. The word is **neat**
3. The word is **puck**
4. The word is **took**
5. The word is **rip**
6. The word is **ray**
7. The word is **sang**
8. The word is **will**
9. The word is **man**
10. The word is **gale**
11. The word is **safe**
12. The word is **peas**
13. The word is **kid**
14. The word is **sass**
15. The word is **sick**
16. The word is **hold**
17. The word is **but**
18. The word is **lane**
19. The word is **bun**
20. The word is **just**
21. The word is **pan**
22. The word is **dig**
23. The word is **bit**
24. The word is **pin**
25. The word is **tease**
26. The word is **sent**
27. The word is **sup**
28. The word is **red**
29. The word is **not**
30. The word is **dung**
31. The word is **pig**
32. The word is **seem**
33. The word is **day**
34. The word is **rest**
35. The word is **page**
36. The word is **bash**
37. The word is **shop**
38. The word is **fig**
39. The word is **tam**
40. The word is **cane**
41. The word is **same**
42. The word is **toil**
43. The word is **fill**
44. The word is **cuss**
45. The word is **feel**
46. The word is **bark**
47. The word is **heath**
48. The word is **ten**
49. The word is **thaw**
50. The word is **bead**

UNCONTROLLED COPY FOR INSPECTION AND USE

UNCONTROLLED COPY FOR INSPECTION AND USE

**List 9A**

1. The word is **pick**
2. The word is **meat**
3. The word is **pub**
4. The word is **look**
5. The word is **sip**
6. The word is **rake**
7. The word is **fang**
8. The word is **bill**
9. The word is **mat**
10. The word is **male**
11. The word is **sane**
12. The word is **peach**
13. The word is **kill**
14. The word is **sack**
15. The word is **sill**
16. The word is **cold**
17. The word is **bug**
18. The word is **lace**
19. The word is **sun**
20. The word is **gust**
21. The word is **pass**
22. The word is **dim**
23. The word is **fit**
24. The word is **fin**
25. The word is **teach**
26. The word is **went**
27. The word is **sum**
28. The word is **wed**
29. The word is **hot**
30. The word is **dud**
31. The word is **pill**
32. The word is **seethe**
33. The word is **pay**
34. The word is **vest**
35. The word is **pace**
36. The word is **back**
37. The word is **top**
38. The word is **rig**
39. The word is **tap**
40. The word is **cave**
41. The word is **game**
42. The word is **foil**
43. The word is **fit**
44. The word is **cup**
45. The word is **eel**
46. The word is **park**
47. The word is **heap**
48. The word is **pen**
49. The word is **paw**
50. The word is **beam**

**List 10A**

1. The word is **kick**
2. The word is **feat**
3. The word is **pup**
4. The word is **book**
5. The word is **lip**
6. The word is **rate**
7. The word is **bang**
8. The word is **fill**
9. The word is **mass**
10. The word is **tale**
11. The word is **sale**
12. The word is **peak**
13. The word is **king**
14. The word is **sag**
15. The word is **sip**
16. The word is **told**
17. The word is **bun**
18. The word is **lake**
19. The word is **gun**
20. The word is **bust**
21. The word is **pat**
22. The word is **did**
23. The word is **kit**
24. The word is **tin**
25. The word is **tear**
26. The word is **bent**
27. The word is **sun**
28. The word is **shed**
29. The word is **pot**
30. The word is **duck**
31. The word is **pin**
32. The word is **seed**
33. The word is **gay**
34. The word is **test**
35. The word is **pave**
36. The word is **bath**
37. The word is **pop**
38. The word is **pig**
39. The word is **tan**
40. The word is **cape**
41. The word is **came**
42. The word is **boil**
43. The word is **fib**
44. The word is **cud**
45. The word is **keel**
46. The word is **lark**
47. The word is **heave**
48. The word is **den**
49. The word is **saw**
50. The word is **beat**

UNCONTROLLED COPY FOR INSPECTION AND USE

**List 11A**

1. The word is **tick**
2. The word is **heat**
3. The word is **pus**
4. The word is **hook**
5. The word is **dip**
6. The word is **raze**
7. The word is **gang**
8. The word is **hill**
9. The word is **mad**
10. The word is **bale**
11. The word is **save**
12. The word is **peace**
13. The word is **kin**
14. The word is **sad**
15. The word is **sit**
16. The word is **fold**
17. The word is **bus**
18. The word is **late**
19. The word is **run**
20. The word is **rust**
21. The word is **pack**
22. The word is **dill**
23. The word is **wit**
24. The word is **din**
25. The word is **teal**
26. The word is **rent**
27. The word is **sung**
28. The word is **bed**
29. The word is **lot**
30. The word is **dug**
31. The word is **pick**
32. The word is **seek**
33. The word is **say**
34. The word is **best**
35. The word is **pay**
36. The word is **ban**
37. The word is **hop**
38. The word is **dig**
39. The word is **tang**
40. The word is **cake**
41. The word is **fame**
42. The word is **coil**
43. The word is **fig**
44. The word is **cut**
45. The word is **peel**
46. The word is **mark**
47. The word is **heal**
48. The word is **men**
49. The word is **jaw**
50. The word is **beach**

UNCONTROLLED COPY FOR INSPECTION AND USE

**List 12A**

1. The word is **sick**
2. The word is **seat**
3. The word is **pun**
4. The word is s**hook**
5. The word is **hip**
6. The word is **race**
7. The word is **rang**
8. The word is **kill**
9. The word is **map**
10. The word is **pale**
11. The word is **sake**
12. The word is **peat**
13. The word is **kick**
14. The word is **sap**
15. The word is **sing**
16. The word is **sold**
17. The word is **buck**
18. The word is **lame**
19. The word is **fun**
20. The word is **dust**
21. The word is **path**
22. The word is **dip**
23. The word is **hit**
24. The word is **sin**
25. The word is **team**
26. The word is **tent**
27. The word is **sud**
28. The word is **fed**
29. The word is **got**
30. The word is **dun**
31. The word is **pit**
32. The word is **seep**
33. The word is **may**
34. The word is **nest**
35. The word is **pane**
36. The word is **bad**
37. The word is **cop**
38. The word is **wig**
39. The word is **tack**
40. The word is **came**
41. The word is **tame**
42. The word is **oil**
43. The word is **fizz**
44. The word is **cub**
45. The word is **reel**
46. The word is **dark**
47. The word is **hear**
48. The word is **hen**
49. The word is **raw**
50. The word is **beak**

UNCONTROLLED COPY FOR INSPECTION AND USE

### Revision History

| Revision | Date | Reason for Revision |
|---|---|---|
| 0.0 | 21 March 2020 | Original release |

UNCONTROLLED COPY
FOR INSPECTION AND USE

UNCONTROLLED COPY

DETERMINATION OF AIR FLOW FOR
POWERED AIR-PURIFYING RESPIRATORS
STANDARD TESTING PROCEDURE (STP)

1. PURPOSE

This test establishes the procedure for ensuring that the level of protection provided by the air flow test requirements on powered air-purifying respirator submitted for Approval, Extension of Approval, or examined during Certified Product Audits, meet the minimum certification standards set forth in 42 CFR, Part 84, Subpart G, Section 84.63(a)(c)(d), and Subpart KK, Section 84.1157(a); Volume 60, Number 110, June 8, 1995.

2. GENERAL

This procedure describes the Determination of Air Flow For Powered Air-Purifying Respirators test in sufficient detail that a person in the appropriate technical field can conduct the test and determine whether or not the product passes the test.

3. EQUIPMENT/MATERIALS

3.1. The list of necessary test equipment and materials is as follows:

3.1.1. Air tight chamber - approximately 24 inches by 24 inches by 16 inches with a hinged door, a 3 inch diameter inlet for accepting breathing tubes and adapters, a one inch diameter outlet, and a 1/4 inch outlet for a manometer probe.

3.1.2. Setra electronic manometer.

3.1.3. Tubing and connectors.

3.1.4. Dry test meter - 10 cubic feet per revolution.

3.1.5. Vacuum source - Spencer turbo compressor Model 075-1/3.

3.1.6. Digital stopwatch.

4. TESTING REQUIREMENTS AND CONDITIONS

4.1. Prior to beginning any testing, all measuring equipment to be used must have been calibrated in accordance with the manufacturer's calibration procedure and schedule. At a minimum, all measuring equipment utilized for this testing must have been calibrated within the preceding 12 months using a method traceable to the National Institute of Standards and Technology (NIST).

4.2. Normal laboratory safety practices must be observed. This includes safety precautions described in the current ALOSH Facility Laboratory Safety Manual.

4.2.1. Safety glasses, lab coats, and hard-toe shoes must be worn at all times.

4.2.2. Work benches must be maintained free of clutter and non-essential test equipment.

4.2.3. When handling any glass laboratory equipment, lab technicians and personnel must wear special gloves which protect against lacerations or punctures.

5. PROCEDURE

Note: Reference Section 3 for equipment, model numbers and manufacturers. For calibration purposes use those described in the manufacturer's operation and maintenance manuals.

5.1. Set up the equipment as show in Figure 1.

5.2. Connect the respirator to the chamber. On units with breathing tubes, the blower is placed outside of the chamber and the breathing tube is attached to an adapter identical to the manufacturer’s connector on the facepiece, helmet, or hood. On units where the blower is in the helmet, an adapter is attached to the helmet and inserted through the inlet of the chamber and sealed.

5.3. Close the door of the chamber.

5.4. Check the electric manometer for zero, adjust to zero if necessary.

5.5. Turn on the PAPR and the vacuum pump.

5.6. Attach the tubing to the electric manometer and adjust to zero using the valve on the vacuum pump.

5.7. Check all connections for leaks.

5.8. If a leak is detected, reseal and readjust the vacuum.

5.9. Time 1 minute on a stopwatch and count the number of CFM on the dry test meter. 1 revolution equals 10 CFM.

UNCONTROLLED COPY

5.10. Calculate the airflow of the respirator using equation #1 (Listed in 6.3).

5.11. Disconnect manometer tubing.

5.12. Turn off the PAPR and vacuum pump.

6. PASS/FAIL CRITERIA

6.1. The criterion for passing this test is set forth in 42 CFR, Part 84, Subpart G, Section 84.63(a)(c)(d), and Subpart KK, Section 84.1157(a); Volume 60, Number 110, June 8, 1995.

6.2. This test establishes the standard procedure for ensuring that:

84.63. Test requirements; general.

(a) Each respirator and respirator component shall when tested by the applicant and by the Institute, meet the applicable requirements set forth in subparts H through L of this part.

(c) In addition to the minimum requirements set forth in subparts H through L of this part, the Institute reserves the right to require, as a further condition of approval, any additional requirements deemed necessary to establish the quality, effectiveness, and safety of any respirator used as protection against hazardous atmospheres.

(d) Where it is determined after receipt of an application that additional requirements will be required for approval, the Institute will notify the applicant in writing of these additional requirements, and necessary examinations, inspections, or tests, stating generally the reasons for such requirements, examinations, inspections, or tests.

84.1157. Chemical cartridge respirators with particulate filters; performance requirements; general. Chemical cartridge respirators with particulate filters and the individual components of each such device shall, as appropriate, meet the following minimum requirements for performance and protection:

(a) Breathing resistance test. (1) Resistance to airflow will be measured in the facepiece, mouthpiece, hood, or helmet of a chemical cartridge respirator mounted on a test fixture with air flowing at a continuous rate of 85 liters per minute, both before and after each test conducted in accordance with paragraphs (d) through (f) of this section

UNCONTROLLED COPY

(2) The maximum allowable resistance requirements for chemical cartridge respirators are as follows:

Maximum Resistance
[mm. water-column height]

| Type of chemical cartridge respirator | Inhalation | | Exhalation |
|---|---|---|---|
| | | Final[1] | |
| For gases, vapors, or gases and vapors, and dusts, fumes, and mists | 50 | 70 | 20 |
| For gases, vapors, or gases and vapors, and mists of paints, lacquers, and enamels | 50 | 70 | 20 |

[1]Measured at end of service life specified in Table 11 in subpart L of this part.

6.3. Equation #1: Airflow (LPM) = $\frac{(28.32\ \text{LPM}) * (\text{number of CFM})}{1\ \text{min}}$

7. RECORDS/TEST SHEETS

7.1. Test data collected shall be recorded on the DETERMINATION OF AIR FLOW FOR POWERED AIR PURIFYING RESPIRATORS test data sheet.

7.2. All videotapes and photographs of the actual test being performed, or of the tested equipment shall be maintained in the task file as part of the permanent record.

7.3. All equipment failing any portion of this test will be handled as follows:

7.3.1. If the failure occurs on a new certification application, or extension of approval application, send a test report to the RCT Leader and prepare the hardware for return to the manufacturer.

7.3.2. If the failure occurs on hardware examined under an Off-the-Shelf Audit the hardware will be examined by a technician and the RCT Leader for cause. All equipment failing any portion of this test may be sent to the manufacturer for examination and then returned to NIOSH. However, the hardware tested shall be held at the testing laboratory until authorized for release by the RCT Leader, or his designee, following the standard operating procedures outlined in Procedure for Scheduling, and Processing Post-Certification Product Audits, RB-SOP-0005-00.

UNCONTROLLED COPY

National Institute for Occupational Safety and Health
Respirator Branch
Test Data Sheet

NIOSH

Task Number:
Test:
Manufacturer:
Item Tested:

Reference No.:
STP No.:

Mask Type:

| AIR FLOW | | | |
|---|---|---|---|
| Sample | Minimum Allowed (Lpm) | Actual (Lpm) | Result |
| Initial<br>Final | | | |
| Overall Result: | | | |

Signature:

Date: ____________

Engineering Technician

UNCONTROLLED COPY

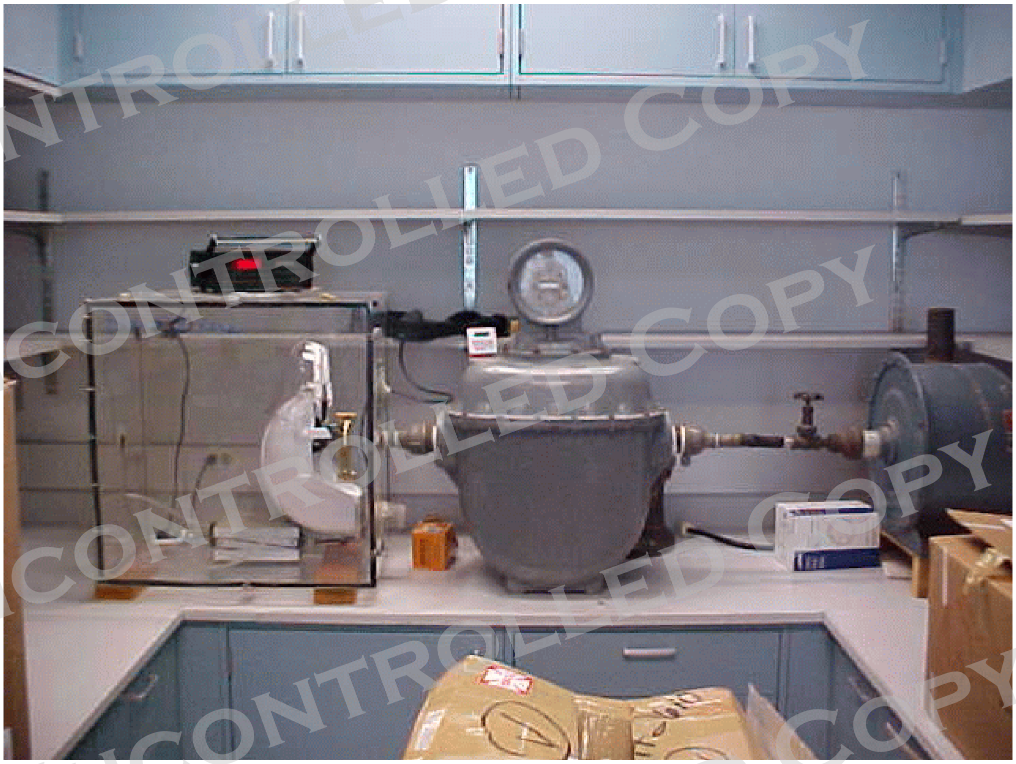

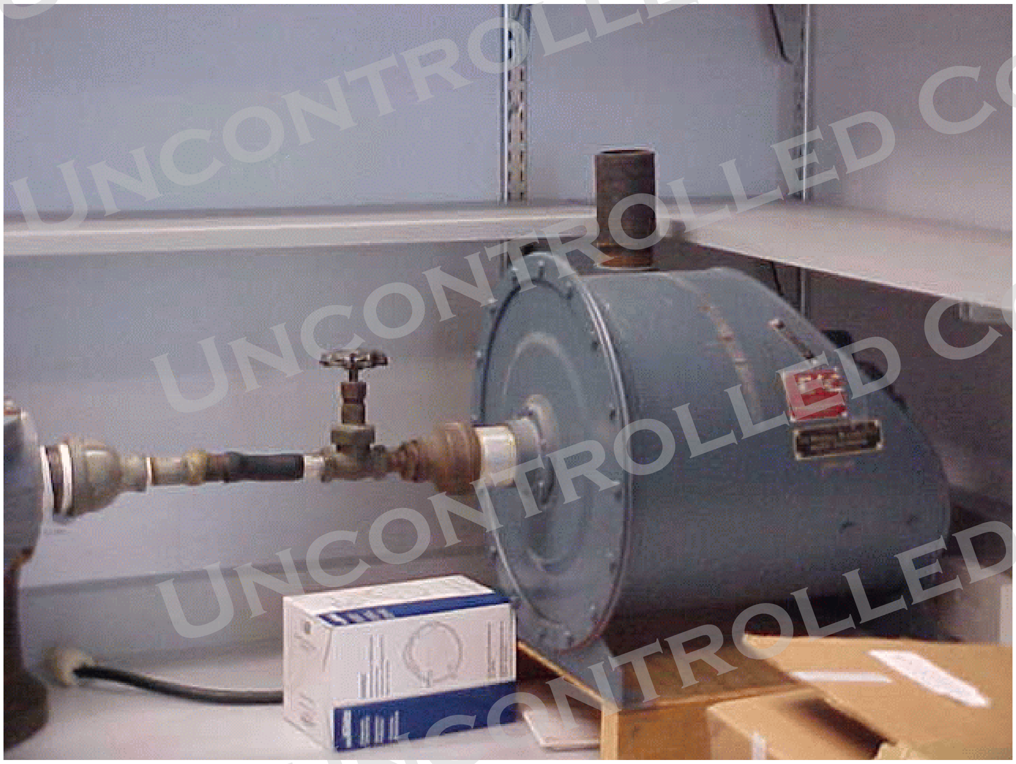

UNCONTROLLED COPY

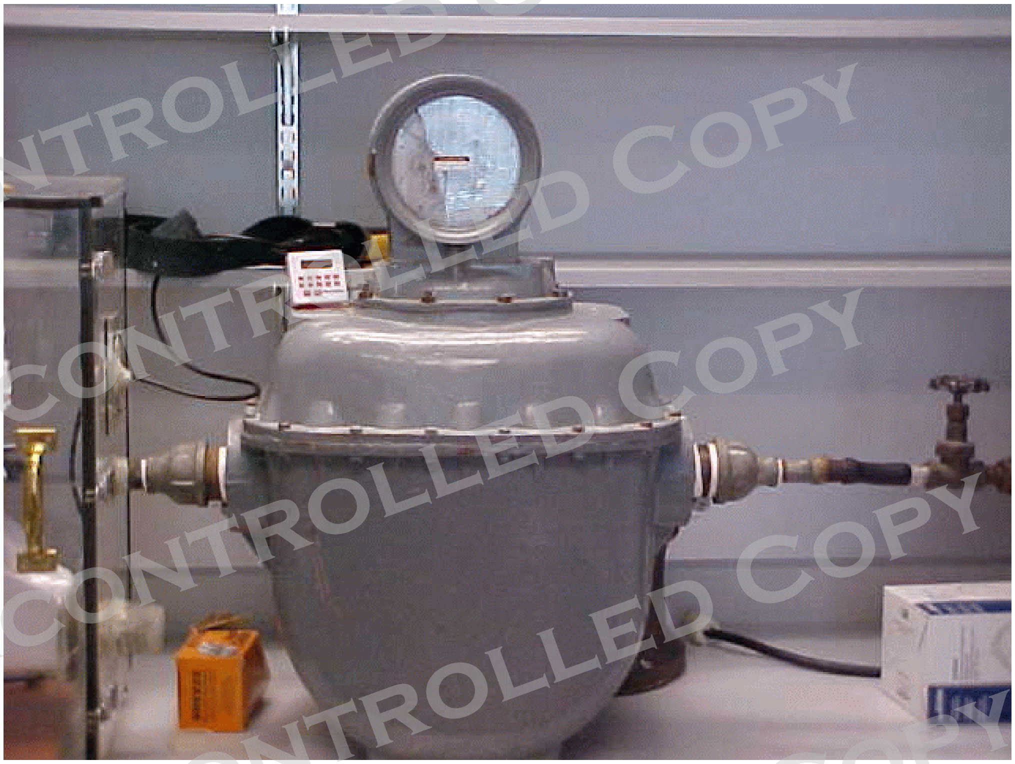

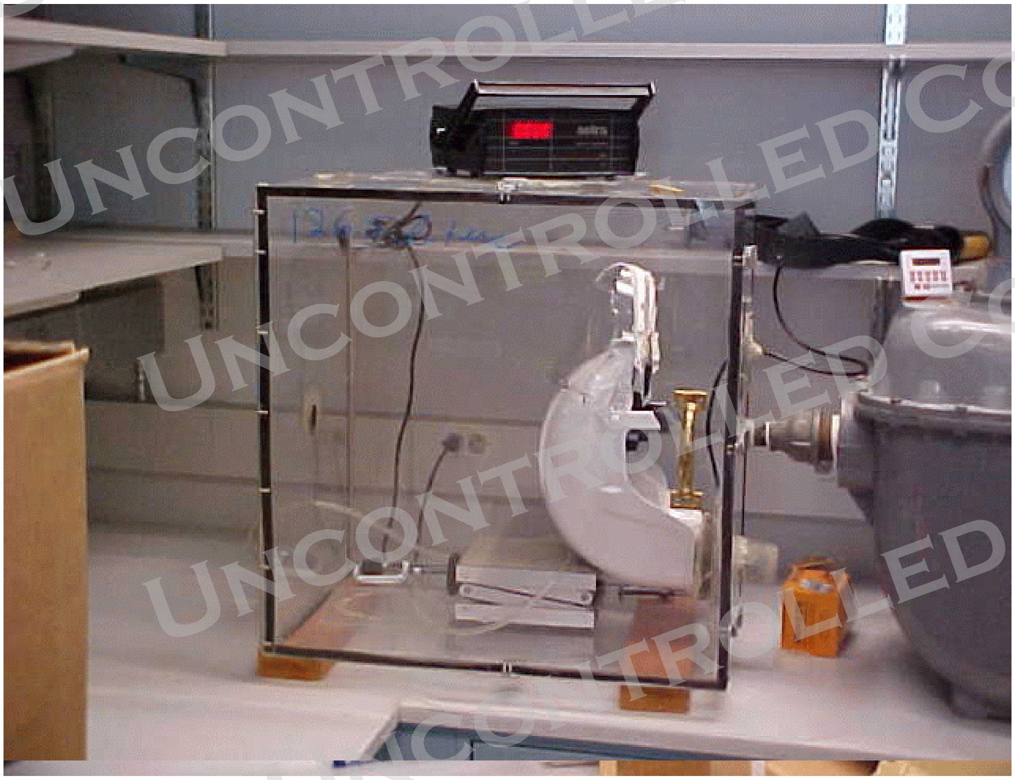

UNCONTROLLED COPY

### Revision History

| Revision | Date | Reason for Revision |
|---|---|---|
| 1.0 | 12 July 2001 | Historic document |
| 1.1 | 6 June 2005 | Update header and format to reflect lab move from Morgantown, WV<br>No changes to method |

UNCONTROLLED COPY

UNCONTROLLED COPY

### DETERMINATION OF NOISE LEVEL TEST, POWERED AIR-PURIFYING RESPIRATOR WITH HOODS OR HELMETS STANDARD TESTING PROCEDURE (STP)

#### 1. PURPOSE

This document establishes the procedure for ensuring that operational noise levels of powered air-purifying respirators with hoods or helmets, as submitted for Approval, Extension of Approval, or examined during Certified Product Audits, do not exceed the established maximum certification requirements as provided for by 42 CFR Part 84, Subpart G, Section 84.63(a)(c)(d), Subpart L, Section 84.202, and Subpart KK, Section 84.1139; Volume 60, Number 110, June 8, 1995.

#### 2. GENERAL

This STP describes the Determination of Noise Level Test, Powered Air-Purifying Respirators with Hoods or Helmets test in sufficient detail that a person knowledgeable in the appropriate technical field can select equipment with the necessary resolution, conduct the test, and determine whether or not the product passes the test.

#### 3. EQUIPMENT/MATERIALS

3.1. The list of necessary test equipment and materials follows.

3.1.1. Completely assembled, powered air-purifying hood or helmet respirator in the configuration as worn by the user with fully charged battery and new air purifying elements

3.1.2. Life-size mannequin

3.1.3. Precision, fast-response sound level meter with built in A-weighting network, capable of averaging measurements over selected time intervals of at least up to 30 seconds. Sound level meter must accommodate two microphone inputs.

#### 4. TESTING REQUIREMENTS AND CONDITIONS

4.1. Prior to beginning any testing, confirm that all measuring equipment employed has been calibrated in accordance with the testing laboratory's calibration procedure and schedule. All measuring equipment utilized for this testing must have been calibrated using a method traceable to recognized international standards when available.

4.2. A background noise level of no greater than 60 dB shall be established and maintained in the location where the procedure is performed.

5. PROCEDURE

5.1 Prior to the use of the test subjects, a noise level screening test will be performed on the complete respirator assembly affixed to a mannequin. The purpose of the screening is to prevent exposing subjects to noise levels which may exceed 85 dBA.

5.1.1. Position the microphones of the sound level meter on each ear of the mannequin.

5.1.2. Using the sound level meter, verify the background noise requirement per section 4.2.

5.1.1. Following the respirator manufacturer's instructions, mount the respirator assembly onto the mannequin.

5.2. Each sample measurement should be averaged over 30 seconds. Once the dBA noise level of the mannequin setup has been determined to be below the 85 dBA safety limit, test subject testing may begin.

5.3 The evaluation is made on three test subjects.

5.4. It is recommended that both males and females be employed as test subjects, and that a wide variation in body size and shape of subjects be sought.

5.5. The test subjects will be allowed to wear ear-insert type hearing protectors, which do not interfere with the positioning of the microphones, if they desire. A choice of protectors will be provided for this purpose.

5.6. Two readings are taken on each subject at both ears and the results averaged.

5.7. Record the results.

6. PASS/FAIL CRITERIA

6.1. The criterion for passing this test is set forth in 42 CFR Part 84, Subpart G, Section 84.63(a)(c)(d), Subpart L, Section 84.202 and Subpart KK, Section 84.1139; Volume 60, Number 110, June 8, 1995.

6.2. This test establishes the standard procedure for ensuring that:

84.63 Test requirements; general.

(a) Each respirator and respirator component shall when tested by the applicant and by the Institute, meet the applicable requirements set forth in subparts H through L of this part.

(c) In addition to the minimum requirements set forth in subparts H through L of this part, the Institute reserves the right to require, as a further condition of approval, any

UNCONTROLLED COPY

additional requirements deemed necessary to establish the quality, effectiveness, and safety of any respirator used as protection against hazardous atmospheres.

(d) Where it is determined after receipt of an application that additional requirements will be required for approval, the Institute will notify the applicant in writing of these additional requirements, and necessary examinations, inspections, or tests, stating generally the reasons for such requirements, examinations, inspections, or tests.

84.202 Air velocity and noise levels; hoods and helmets; minimum requirements.

Noise levels generated by the respirator will be measured inside the hood or helmet at maximum airflow obtainable and shall not exceed 80 dBA.

84.1139 Air velocity and noise levels; hoods and helmets; minimum requirements.

Noise levels generated by the respirator will be measured inside the hood or helmet at maximum airflow obtainable and shall not exceed 80 dBA.

7. RECORDS\TEST SHEETS

7.1. Record test data in a format that shall be stored and retrievable. Data is to be reported as shown in the attached example data sheet.

8. ATTACHMENTS

8.1. Example Test Data Sheet

UNCONTROLLED COPY

Attachment 8.1. Example Test Data Sheet

**National Institute for Occupational Safety and Health**
**Respirator Branch**
**Test Data Sheet**

NIOSH

**Task Number:**
**Test:** Sound Level Test-Direct dB
**Manufacturer:**
**Item Tested:**

**Reference No.:** CFR 84.1139; 84.202
**STP No.:** 30.1

| Subject | Trial 1 (dba) | | Trial 2 (dba) | | Average (dba) | | Maximum Allowable (dba) | Result |
|---|---|---|---|---|---|---|---|---|
| | Left Ear | Right Ear | Left Ear | Right Ear | Left Ear | Right Ear | | |
| Manikin | | | | | | | 85 | |
| 1 | | | | | | | 80 | |
| 2 | | | | | | | 80 | |
| 3 | | | | | | | 80 | |

**Overall Result:**

**Signature** Date:_____

Engineering Technician

---

Task Number:
Test: Sound Level Test-Direct dB
Manufacturer: Item Tested:

Reference No.: CFR 84.1139; 84.202
STP No.: 30.1

Comments:
Testing was done using the (respirator part numbers). All test subjects were medically cleared for testing.
Was all equipment verified to be in calibration throughout all testing? O Yes O No

**Signature** Date:_____

Engineering Technician

UNCONTROLLED COPY

### Revision History

| Revision | Date | Reason for Revision |
| --- | --- | --- |
| 1.0 | 7 March 2002 | Historic document |
| 1.1 | 14 June 2005 | Update header and format to reflect lab move from Morgantown, WV No changes to method |
| 2.0 | 28 May 2019 | The document is updated to current style and content standards. There is no change to the test set up or method, but the specified sound measurement instrument has been updated. The ability to collect an average measurement expressed in dBA over the specified 30-second interval eliminates the need to convert dose to dBA. |

UNCONTROLLED COPY

UNCONTROLLED COPY

### DETERMINATION OF PARTICULATE FILTER PENETRATION TEST POWERED AIR-PURIFYING RESPIRATOR FILTERS STANDARD TESTING PROCEDURE (STP)

#### l. PURPOSE

This test establishes the procedure for ensuring that the level of protection provided by powered air-purifying respirator filters submitted for Approval, Extension of Approval, or examined during Certified Product Audits, meet the filter penetration requirements set forth in 42 CFR, Part 84, Subpart G, Section 84.63(a)(c)(d) and Subpart KK, Section 84.1151(a)(c), except that the flow requirements of Section 84.1151(a)(c) are not used. In their place, flow requirements specified in Section 84.1156(c)(2) are applied. This is done in order that PAPR filters are tested, to the extent possible, at the minimum required air flow rate of the PAPR.

#### 2. GENERAL

This STP describes the Determination of Particulate Filter Penetration Test, Powered Air-purifying Respirator Filters test in sufficient detail that a person knowledgeable in the appropriate technical field can select equipment with the necessary resolution, conduct the test, and determine whether or not the product passes the test.

#### 3. EQUIPMENT/MATERIALS

3.1. The list of necessary test equipment and materials follows:

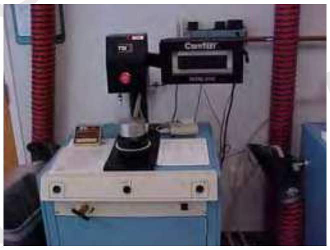

3.1.1. TSI Model 8130 Automated Filter Tester or equivalent instrument. Air flow control accuracy is 2% of full scale. Pressure measurement accuracy is 2% of full scale. Penetrations can be measured to 0.001%, efficiencies to 99.999%.

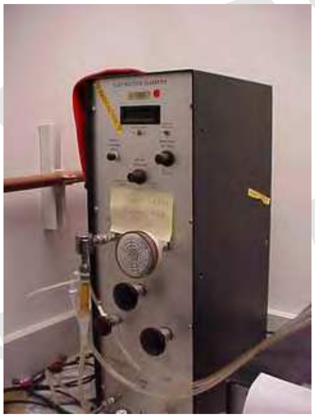

3.1.2. Particle sizing instrument (such as TSI Model 3936 Scanning Mobility Particle Size Spectrometer or equivalent) that is capable of determining submicrometer particles according to count median diameter (CMD).

.

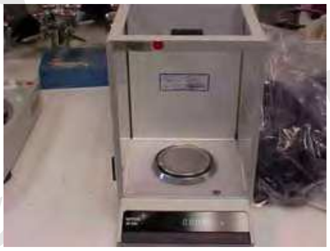

3.1.3. Microbalance accurate to 0.0001 grams (g).

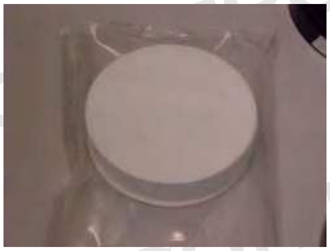

3.1.4. Gelman 102 mm diameter, type A/E glass filters or equivalent high efficiency filters with a 1 micrometer pore size.

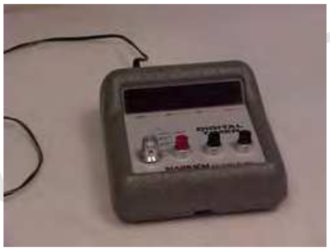

3.1.5. Timer (accurate to 0.01 second).

UNCONTROLLED COPY

3.1.6. Dioctyl phthalate ((DOP, di(2-ethylhexyl)phthalate)) min. 98%.

3.1.7. Respirator filter holder supplied for specific manufacturer type which is compatible with TSI filter tester. NIOSH will not be obligated to use these holders for actual certification testing. All manufacturer test fixtures must be correlated with the NIOSH test method (see Work Instruction WI- 1611).

3.1.8 Thermal printer (supplied) or optional data acquisition system.

3.2. Refer to the following Work Instructions for further information on performing this test:
TEB-RCT-APR-WI-1011 – Laboratory Safety Procedures for Particulate Tests for Powered Air Purifying Respirators
TEB-RCT-APR-WI-1111 – Calibration Procedures for Particulate Test for Powered Respirators
TEB-RCT-APR-WI-1211 – Start–Up and Shut–Down Procedures for Particulate Test for Powered Air Purifying Respirators
TEB-RCT-APR-WI-1411 – Reporting Results for Particulate Test for Powered Air Purifying Respirators
TEB-RCT-APR-WI-1511 – Checking System Performance for Particulate Test for Powered Air Purifying Respirators
TEB-RCT-APR-WI-1611 – Correlating Manufacturer – Supplied Test Fixtures for Particulate Test for Powered Air Purifying Respirators

4. TESTING REQUIREMENTS AND CONDITIONS

4.1. Prior to beginning any testing, all measuring equipment to be used must have been calibrated in accordance with the testing laboratory's calibration procedure and schedule. All measuring equipment utilized for this testing must have been calibrated using a method traceable to the National Institute of Standards and Technology (NIST) when available.

4.2. Any laboratory using this procedure to supply certification test data as a contractor to NIOSH will be subject to the provisions of the NIOSH Supplier Qualification Program (SQP). This program is based on the tenets of *ISO/IEC 17025, the NIOSH Manual of Analytical Methods* and other NIOSH guidelines. An initial complete quality system audit and follow on audits are requirements of the program. Additional details of the Program and its requirements can be obtained directly from the Institute.*
***Note** 4.2 does not apply to Pretest data from applicants as required under 42 CFR 84.64.

4.3. Precision and accuracy (P&A) must be determined for each instrument in accordance with laboratory procedures and NIOSH/NPPTL guidance. Sound practice requires, under *NIOSH Manual of Analytical Methods*, demonstrating a tolerance range of expected data performance of a plus or minus 25% of a 95% confidence interval of the stated standard requirement. NIOSH/NPPTL P&A tolerance can be higher but not lower.

UNCONTROLLED COPY

4.4. The precision and accuracy of this method is monitored by the validation method which is incorporated in the automated filter tester procedure. This procedure is performed on a daily basis when testing is performed. This procedure is designed to test many aspects of the method, for proper photometer and general system operation. The validation technique uses "green line" filter media discs, 6 inch diameter, HE 1071 grade, H & V brand, P/N 813010, with a known penetration range, which are tested at least once in each 8-hour test period (see 5.2.5).

4.4.1. Two sheets of unused filter media are stacked together and the penetration, flow rate and pressure drop are measured to evaluate the higher range of penetration values. Five unused sheets are stacked together to evaluate the lower range of penetration values.

4.4.2. The analysis of these readings over the long term was used to examine the precision and accuracy of this test method. The table below summarizes the data.

| | Two Sheets | Five Sheets |
|---|---|---|
| Mean | 2.459% | 0.011% |
| Std. Dev. | 0.157 | 0.001 |
| Range | 2.04 – 2.97% | 0.008 – 0.017% |
| N | 56 | 56 |

4.5. Normal laboratory safety practices must be observed. Please refer to Material Safety Data Sheets and the current NIOSH Pittsburgh Health and Safety Program for the proper protection and care in handling, storing, and disposing of the chemicals used in this procedure.

4.6. Dioctyl phthalate is considered a low hazard material with a recommended exposure limit (REL) of 5 $mg/m^3$ with a short-term exposure limit of 10 $mg/m^3$. It may cause mild skin or eye irritation. Carcinogenic effects: Classified as a proven animal carcinogen with unknown relevance to humans by ACGIH; classified as a suspect carcinogen by NTP; not listed by IARC. Local exhaust ventilation is used for the potential sources of DOP from the TSI 8310 filter tester. Safety eyewear and a lab coat should be worn. Splash goggles, protective clothing, boots and gloves should be worn in case of a large spill. Discharge, treatment or disposal may be subject to national, state or local laws.

5. PROCEDURE

Note: Reference Section 3. for equipment, model numbers and manufacturers. For calibration purposes use those described in the manufacturers' operation and maintenance manuals.

5.1. Respirator filters will be challenged by a neat cold–nebulized DOP aerosol at 25 ±5˚C that has been neutralized to the Boltzmann equilibrium state. The particle size distribution will be a count median diameter of 0.185 ± 0.020 micrometer and a geometric standard deviation not exceeding 1.6. Each respirator filter unit will be challenged with an aerosol concentration of 100 ± 10 $mg/m^3$.

UNCONTROLLED COPY

5.1.1. The DOP aerosol concentration will be determined daily by the following gravimetric method and calculated as milligrams per cubic meter ($mg/m^3$).

5.1.2. Weigh a Gelman 102 mm filter to the nearest 0.1 mg., mount in the gravimetric filter holder, subject it to the generated aerosol at 30 Lpm for 40 minutes , and reweigh the filter. Use a timer to monitor the duration of the test. Record the pre- and post-weights, time, and average flow rate on the data sheet and calculate the aerosol concentration in $mg/m^3$ by the following formula:

$$\text{Concentration in } mg/m^3 = \frac{W2 - W1}{(Q / 1000)\,(T)}$$

Where:
W1 = Initial filter weight in mgs.
W2 = Final filter weight in mgs.
Q = Flowrate in liters per minute
T = Elapsed time in minutes

With a flowrate of 30 Lpm for 40 minutes, the above formula simplifies to:

$$C = \frac{W2 - W1}{1.2}$$

5.1.3. The upstream and downstream photometer readings are used for monitoring stability and for calculating a photometer correlation factor (CF). The correlation factor is determined with an empty filter holder and is calculated internally as shown below:

$$CF = \frac{\text{Downstream Photometer Voltage} - \text{Downstream Background Voltage}}{\text{Upstream Photometer Voltage} - \text{Downstream Background Voltage}}$$

The correlation factor is used by the software to express the upstream photometer signal in terms of the downstream photometer signal. Follow Work Instruction WI- 1511 for determining, monitoring and recording the CF.

5.1.4. The DOP particle size distribution shall be verified using “green line” filter discs supplied by TSI with a known penetration range. Graphs of penetration vs. resistance for two sheets and five sheets of stacked filter discs are supplied with each lot of the standard filters, with a central line and upper and lower lines representing the expected penetration range at a given resistance. The test data should fall within an acceptance zone having boundaries defined by the upper and lower curves on the graphs. Follow the procedure in Work Instruction WI-1505. The standard filter test using both 2 sheets and 5 sheets will be run at least once in each 8 hour test period to verify that the aerosol distribution is within the acceptance zone.

5.1.5. If the instantaneous filter penetration is not within the acceptance zone for any sample, abort testing and check the aerosol particle size with the Scanning

UNCONTROLLED COPY

Mobility Particle Size (SMPS) Spectrometer.

5.2. The DOP particle size will be monitored at least once every three months (quarterly) with the SMPS spectrometer to ensure the particle size distribution count median diameter remains in the range of 0.185 ± 0.020 micrometer with a geometric standard deviation of not more than 1.6.

5.3. Filters shall be tested as follows:

5.3.1. The filter, including the filter holders and gaskets, shall be tested for particle penetration. When the filtering element is not separable from the cartridge or canister, the complete component shall be tested.

5.3.2. When filters are not separable from the respirator body, any exhalation valves shall be sealed to ensure that any leakage due to an exhalation valve is not included in the filter penetration measurement.

5.3.3. Filters not separable from cartridges, canisters, respirators, and odd or unusually shaped filters may be tested on a headform assembly or an assembly provided by manufacturer. Note: NIOSH is not obligated to use the headform assembly or any assembly provided by the manufacturer for certification testing.

5.4. Filters shall be mounted and sealed on holders to prevent leakage around the filter holder. PAPRs are normally designed to use from one to four filters. Filters shall be tested using a single filter regardless of the number of filters used on the unit. Adjustment is made to the flow rate of the test by dividing the specified flow rate for the test, which is based on the minimum required air flow of 115 lpm for tight fitting PAPRs and 170 lpm for loose fitting PAPRs, by the number of filters used. The table below shows the test flow rate depending on the type of PAPR and the number of filters employed. The highest obtainable air flow for the automated filter testers is typically 96 lpm and this flow rate will be used to test single PAPR filters.

| | PAPR FILTER TEST - FLOW RATE (LPM) | |
|---|---|---|
| NUMBER OF FILTERS | TIGHT FITTING FACEPIECE | LOOSE FITTING FACEPIECE |
| 1 | 96 ± 5 | 96 ± 5 |
| 2 | 57 ± 3 | 85 ± 4 |
| 3 | 38 ± 2 | 57 ± 3 |
| 4 | 29 ± 2 | 43 ± 2 |

5.4.1. The challenge flow rate must be checked for stability for at least 30 seconds prior to testing.

5.4.2. If using a TSI 8130 tester, the tester rise time shall be set at 10 seconds, the tester sample time shall be set at 10 seconds, and the tester purge time shall be set at 9 seconds.

5.5. A total of 3 filters shall be tested against the DOP liquid aerosol. Each filter shall be instantaneously loaded and evaluated.

UNCONTROLLED COPY

5.5.1. Any filter that exceeds the specified limit shall be remounted and retested to ensure that leakage was not caused by a mounting leak. If retesting eliminates the leakage, that filter shall be considered an invalid sample and another filter shall be tested in its place.

5.6. The penetration of the 3 filters shall be measured and recorded.

6. PASS/FAIL CRITERIA

6.1. The legal basis for passing this test is set forth in 42 CFR, Part 84, Subpart G, Section 84.63(a)(c)(d) and Subpart KK, Section 84.1151(a)(c); except that the flow requirements of Section 84.1151(a)(c) are not used. In their place, flow requirements specified in Section 84.1156(c)(2) are applied to the extent possible.

6.2. The total leakage for the connector and filter shall not exceed 0.03 percent of the ambient DOP concentration for any test sample.

7. RECORDS/TEST SHEETS

7.1. Record the test data in a format that shall be stored and retrievable.

8. ATTACHMENTS

8.1. Data Sheet

8.2. Test Setup

UNCONTROLLED COPY

8.1. Data Sheet

**National Institute for Occupational Safety and Health**
**Respirator Branch**
**Test Data Sheet**

NIOSH

**Task Number:** **Reference No.:**
**Test:** **STP No.:**
**Manufacturer:**
**Item Tested:**

| Filter | Flow Rate | Maximum Allowable Percent Leakage | Actual Percent Leakage | Result |
|---|---|---|---|---|

**Overall Result:**

**Signature:** **Date:** ______________

**Engineering Technician**

UNCONTROLLED COPY

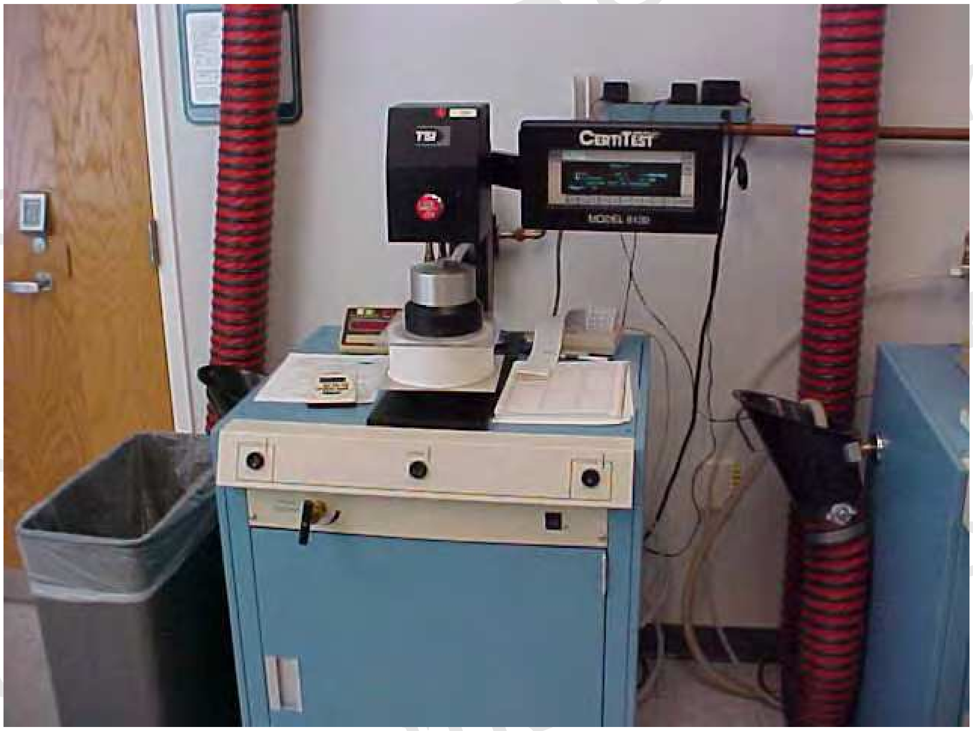

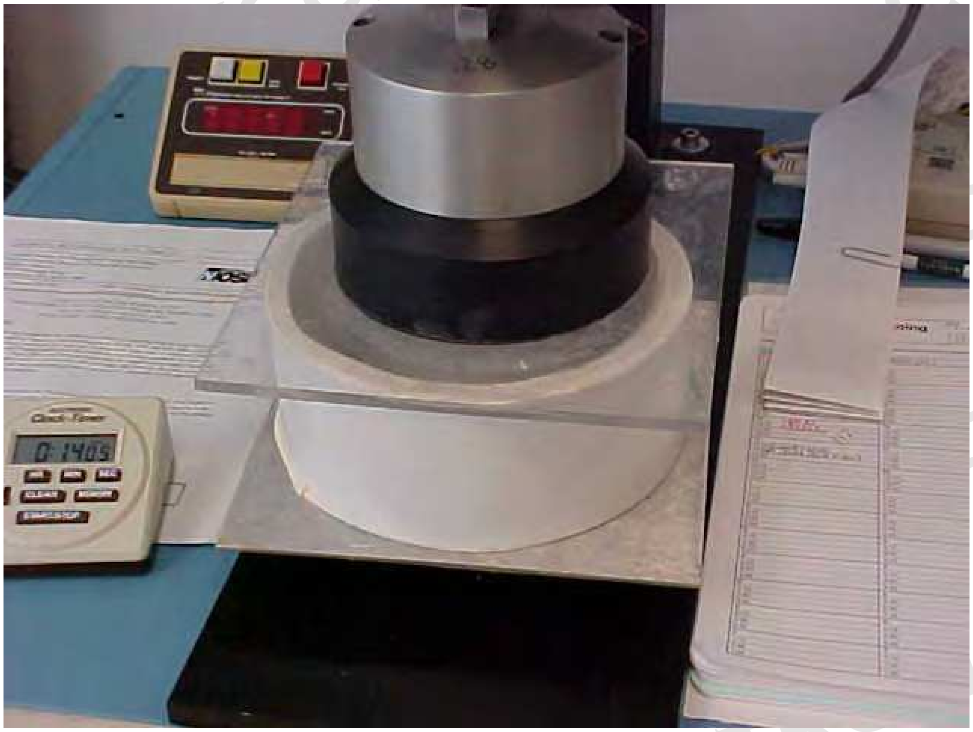

8.2. Test Setup

UNCONTROLLED COPY

### Revision History

| Revision | Date | Reason for Revision | |
|---|---|---|---|
| 1.0 | 7 March 2004 | Historic document | |
| 1.1 | 1 June 2005 | Update header and format to reflect lab move from Morgantown, WV No changes to method | |
| 2.0 | 14 April 2008 | Section | Change |
| | | 3.2. | List of Work Instructions added |
| | | 4. | Requirements and data for precision and accuracy added |
| | | 5. | Editorial changes to clarify procedures |
| | | 5.1.2. | Example calculation for challenge concentration added |
| | | 5.4. | Clarifications to test flow in light of the maximum total test flow of 96 lpm. A table of appropriate flow values is added. |
| | | All | Editorial changes to improve clarity throughout |

UNCONTROLLED COPY
