## Supplementary Material 2 for "De novo Powered Air-Purifying Respirator Design and Fabrication for Pandemic Response"

**PanFab PAPR Design Methods and Design**

Peter Sorger, PhD, 0000-0002-3364-1838

### METHODS

This section describes the methodology pursued in designing the PanFab Custom PAPR and PanFab Commercial PAPR filters and blower units.

Filter selection

NIOSH regulations designate several classes of PAPRs depending on filtration requirements. A key distinction between classes is whether the respirator is approved for use with oil-based aerosols (P type filters, tested for filtration efficiency with dioctyl phthalate (DOP)) or only aqueous aerosols (N type filters, tested for filtration efficiency with NaCl-containing droplets). Commercial PAPR filter cartridges are commonly of the P100 type as they provide protection against both oil and salt-based aqueous aerosols. Another, less commonly used, class of PAPR filter is the N100, which only needs to demonstrate its effectiveness against salt-based aqueous aerosols and not oil. Here, "100" after the type designation refers to filters demonstrating filtration of 99.97% of tested particles[1].

The pressure drop across a filter at the required airflow is an additional consideration that affects blower selection and power consumption. The pressure drop should be as low as possible for a given filtration efficiency, so as to permit the use of a less powerful blower, and consequently a smaller battery, making the system lighter and able to operate longer between charging[2].

© President and Fellows of Harvard College and The Massachusetts Institute of Technology, 2020.

In selecting the PanFab Commercial Filter Cartridge, our goal was to identify and test commercially available products whose supply would not be impacted by shortages during pandemic conditions, and which would be easily adaptable to various blower units. The PanFab Custom Filter Cartridge was designed to have a small overall volume, be lightweight, and have integrated connection features to allow for compatibility with various blower units.

Blower Unit Design

The PAPR blower units consist of a blower, battery, buzzer to warn of low flow rate, control systems, and a housing to enclose these components. The PanFab commercial and custom units were designed to be comfortable to wear for extended periods of time, to be easy to don and doff, and to not hamper the range of motion required in clinical tasks. Additionally, the materials for the external surfaces of the unit were selected so as to be compatible with sterilization with alcohol based wipes. Below we outline the selection criteria used for each of the blower unit components.

The PanFab blowers were selected to produce enough static pressure as to overcome the pressure drop incurred from passing air through the filters and the facepiece at the required flow rate of 170 liters per minute[3]. The commercial and custom battery that powers the blower unit were selected to match the voltage rating of the blower and have sufficient discharge current. We also ensured the batteries had the charge capacity necessary to continuously power the blower for one hour or longer at the required flow rate, a time selected based on the recommendations from HCWs at a major academic medical center. Finally, we ensured that the blower and battery combinations met the NIOSH specification for a noise level at the user's ears below 80 dBA[3].

The blower unit control system was designed to measure the air flow rate through the PAPR and to sound an alarm at a sound level above 80 dBA at the ears (as per NIOSH STP

© President and Fellows of Harvard College and The Massachusetts Institute of Technology, 2020.

CVB-APR-STP-008) to alert the wearer of a low flow condition. The control system was also designed so as to allow for modulating flow rate as needed by the wearer.

The blower unit housings were selected to couple and decouple with the filters and hose by hand without requiring specialized tools, and to be conducive to frequent opening and closing for operations including battery charging, flow rate adjustment, and troubleshooting. Since the air inside the housing has been filtered and there is negative pressure inside the housing, it is essential for the connections of the housing with the filters and the hose, as well the interface between the separable sections of the housing, to be airtight.

Facepiece selection

We used a loose-fitting facepiece, known as the VHA ADAPT PAPR Hood, developed by the The Center for Limb Loss and MoBility (CLiMB) at the University of Washington[4]. This low cost facepiece was developed as part of COVID-19 response efforts and was chosen for prototype testing as commercial facepieces were not readily available.

### RESULTING DESIGNS

This section details the designs that resulted for both the PanFab Custom PAPR and the PanFab Commercial PAPR filter and blower units.

PanFab filter cartridges

*PanFab Commercial Filter Cartridge*

We investigated commercial High Efficiency Particulate Air (HEPA) filters commonly used in vacuum cleaners as possible candidates. HEPA filters are rated as 99.97% efficient at

© President and Fellows of Harvard College and The Massachusetts Institute of Technology, 2020.

300um particle size[5]. The cost and availability of the Milwaukee Tool (Brookfield, WI)[6] in commercial stores was particularly well suited to the crisis-setting PanFab PAPR design.

In order to connect the Milwaukee filter to the housings, an adapter was designed, which could be either 3D printed or injection molded. This adapter would be sealed to the filter outlet on one side using sealants or adhesives such as cyanoacrylate or epoxies, and on the other side would connect to the housing using male NATO 40mm threads. Filter covers were also designed for these filter cartridges, that protect the filter sheet from damage. These covers are also designed to be printed or molded.

*PanFab Custom Filter Cartridge*

We also designed a custom filter cartridge in collaboration with Custom Filters LLC (Aurora, IL) to decrease the weight and size of these devices as compared to the Milwaukee filters, making them more comfortable and easier to use for prolonged periods of time in healthcare settings. These cartridges do not need a separate filter cover, since the cartridge body is designed to envelop the filter media. Additionally, Custom Filters tests every filter cartridges' instantaneous filtration efficiency with PAO aerosol, ensuring the integrity of all filters. This filter also connects to the housing through integrated male NATO 40mm threads.

PanFab blower units

We designed the PanFab Commercial and Custom blower units to have the same internal components, with different housings enclosing the components. NIOSH guidelines specify that airflow into a loose-fitting facepiece must exceed 170 liters/minute (lpm) to ensure maintenance of positive pressure at all activity levels [7]. We chose to design our PAPR to provide 230 lpm,

© President and Fellows of Harvard College and The Massachusetts Institute of Technology, 2020.

similar to some of the commercial PAPR's. In selecting a blower, we determined that compact centrifugal blowers (compared to axial fans) could provide the necessary static pressures needed to overcome the pressure drop created by the filter at the required flow rate, as well as to overcome losses over the flow path to the facepiece. The pressure drop across the filters at the required flow rate was determined as part of the filtration efficiency tests. The blower selected (Delta Electronics, Neihu, Taiwan, Part Number BFB1012HD-04D4L) has a maximum static pressure of 40.6 mm of $H_2O$ and a maximum flow rate of 18.3 cubic feet per meter (cfm). Additionally, the noise level generated by running the PAPR should not exceed 80 dBA per NIOSH guidelines and should allow for sufficient communication between users[8]. The blower selected has a maximum noise level rated at 50.5dBA at a distance of 1 m. In order to meet the requirements of the blower, a 12V NiMH battery pack (Tenergy, Fremont, CA, Amazon Standard Identification Number: B077Y9HNTF) was selected. The battery has a charge capacity of 2000mA, which provides almost 4 hours of continuous operation, at the maximum flow rate, which is higher than the 1hour requirement. The battery connects to the switch through a manual disconnect, which can be disconnected to connect to a Tenergy battery charger (Amazon Standard Identification Number: B08J4H39JV).

The control system of the PAPR consists of an Arduino Uno R3 (Arduino LLC, Boston, MA) board, a linear potentiometer (Precision Electronics Corporation, North York, ON, Canada, part number RV4NAYSD103A) which controls the duty cycle, and consequently the speed of the blower, a printed circuit board (PCB)-mounted, I2C protocol operated differential pressure sensor (Sensirion AG, Staefa, Switzerland, part number SDP810-500PA) which estimates the flow rate, a piezoelectric buzzer (Mallory Sonalert Products Inc., Indianapolis, IN, part number PS-580Q) which warns of low flow rate, and a manual SPST switch (purchased from McMaster-

© President and Fellows of Harvard College and The Massachusetts Institute of Technology, 2020.

Carr, Elmhurst, IL, part number 8002K114) to turn on and off the device. A custom shield was designed and printed at OSH Park (Portland, OR), which assembles with the Arduino, and solders to the various components. An Arduino Protoshield can also be used in place of the custom printed shield.

NIOSH requires PAPRs to have a low flow rate alarm that is activated when airflow into the facepiece falls below the threshold of 170 lpm, which in our design is activated below 230 lpm. In the PanFab PAPR design, the differential pressure sensor detects pressure differential across a venturi (provided by Hunter Engineering Company, Bridgeton, MO), connected to the outlet of the blower, via a 3D printed blower adapter and a 1” OD Silicone tube (purchased from McMaster-Carr, part number 3038K29). Ports of the venturi are connected to the corresponding ones on the differential pressure sensor through a 7/32” OD Silicone tube (purchased from McMaster-Carr, part number 3038K12). It should be noted that the blower adapter connecting the blower outlet to the 1” Silicone tube was designed specifically for the blower selected; a different blower would require a blower adapter design compliant with its outlet geometry.

In order to calibrate the threshold differential pressure corresponding to the threshold flow rate of 230 lpm, the outlet of the venturi was first connected to the Vernier anemometer. The differential pressure sensor was connected to an Arduino Uno R3 board through a custom printed shield, compatible with the Arduino board. The differential pressure across the venturi is read on the Arduino Serial Monitor User Interface, which is plotted against the air speed as the blower speed is increased. The minimum flow rate of 230 lpm is now correlated to the corresponding differential pressure. Once this calibration is done, the differential pressure value obtained is used in microcontroller logic to sound a piezoelectric buzzer, when the differential pressure falls below this value (and the corresponding flow rate) for 10 consecutive seconds. The

© President and Fellows of Harvard College and The Massachusetts Institute of Technology, 2020.

plot of flow rate vs. differential pressure measured, with observed datapoints is shown in **Figure 1** below. The cutoff pressure corresponding to the cutoff flow rate of 230 lpm is marked, which was programmed in the Arduino.

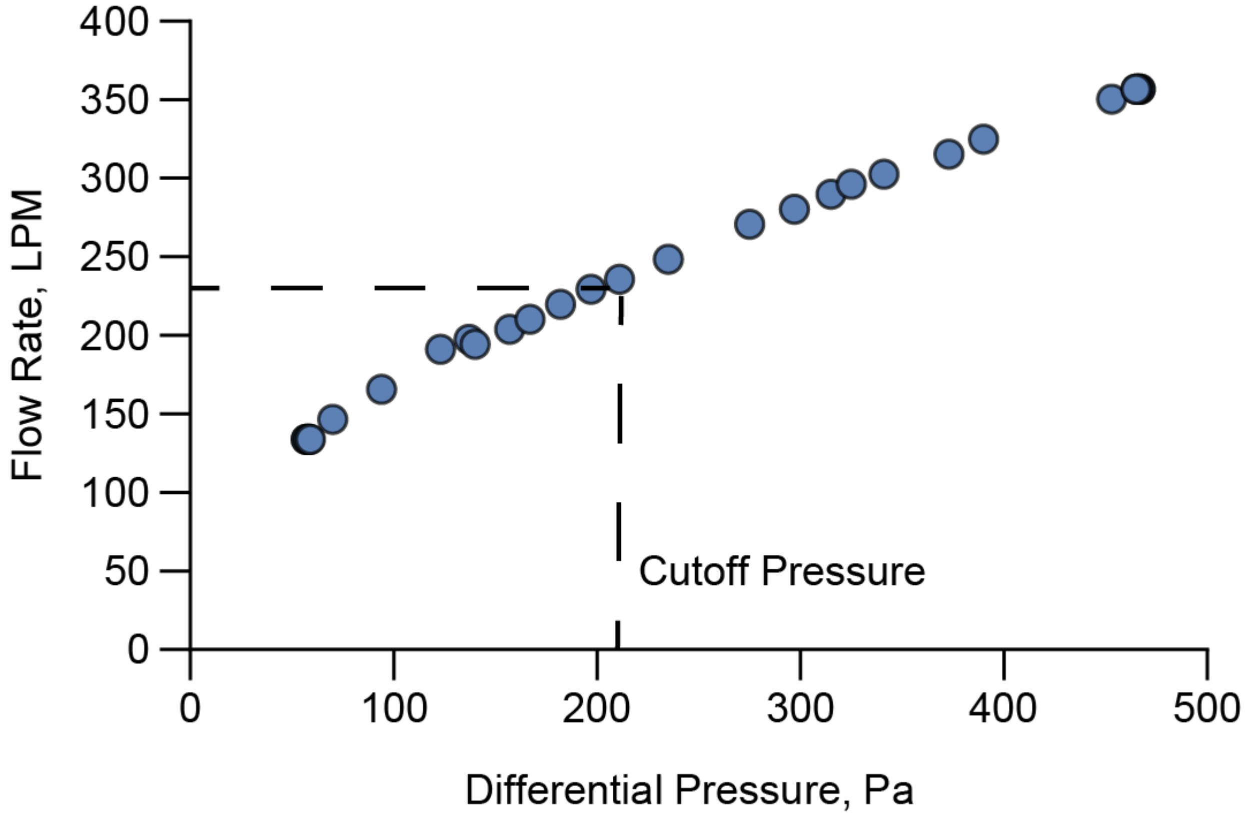


**Figure 1:** Calibration of flow rate through the venturi to the pressure differential across the device, allowing identification of cutoff pressure corresponding to the cutoff flow rate of 230 LPM. Below this cutoff pressure, the piezoelectric buzzer is programmed to sound, alerting the wearer of low flow.

The sound level of the piezoelectric buzzer needs to be loud enough so as to sufficiently alert the wearer of a low flow condition. NIOSH requires the alarm to have a sound level above 80dBA at the ears. We chose a buzzer with a higher sound level since the buzzer is placed inside the housing. Unobstructed, the buzzer chosen is specified as producing 100 dB at a distance of 100 cm.

© President and Fellows of Harvard College and The Massachusetts Institute of Technology, 2020.

In order to maximize the sound level at the ears, the piezoelectric buzzer, installed inside the housing, with filters, hose, and facepiece connected, was programmed to sound at various frequencies. The frequency producing the maximum sound level at the ears was selected, which was done for both housing types, given the different sound attenuating properties expected from the two housings. For the custom housing that will be discussed later, the frequency obtained was 500 Hz, while for the commercial housing, it was 1500 Hz.

In order to simplify the control electronic design and code, a compensatory logic that would automatically increase the flow rate in the event of flow obstruction was considered but not adopted. Based on clinical feedback, very few scenarios were known to exist in clinical procedures where flow obstruction through a PAPR would be of concern. Nevertheless, the user can increase or decrease the flow rate to a comfortable level using the potentiometer, as long as it is above the threshold of 230 lpm.

All the components inside the blower unit is shown in **Figure 2A** and the circuit diagram of the control system is shown in **Figure 2B**.

© President and Fellows of Harvard College and The Massachusetts Institute of Technology, 2020.

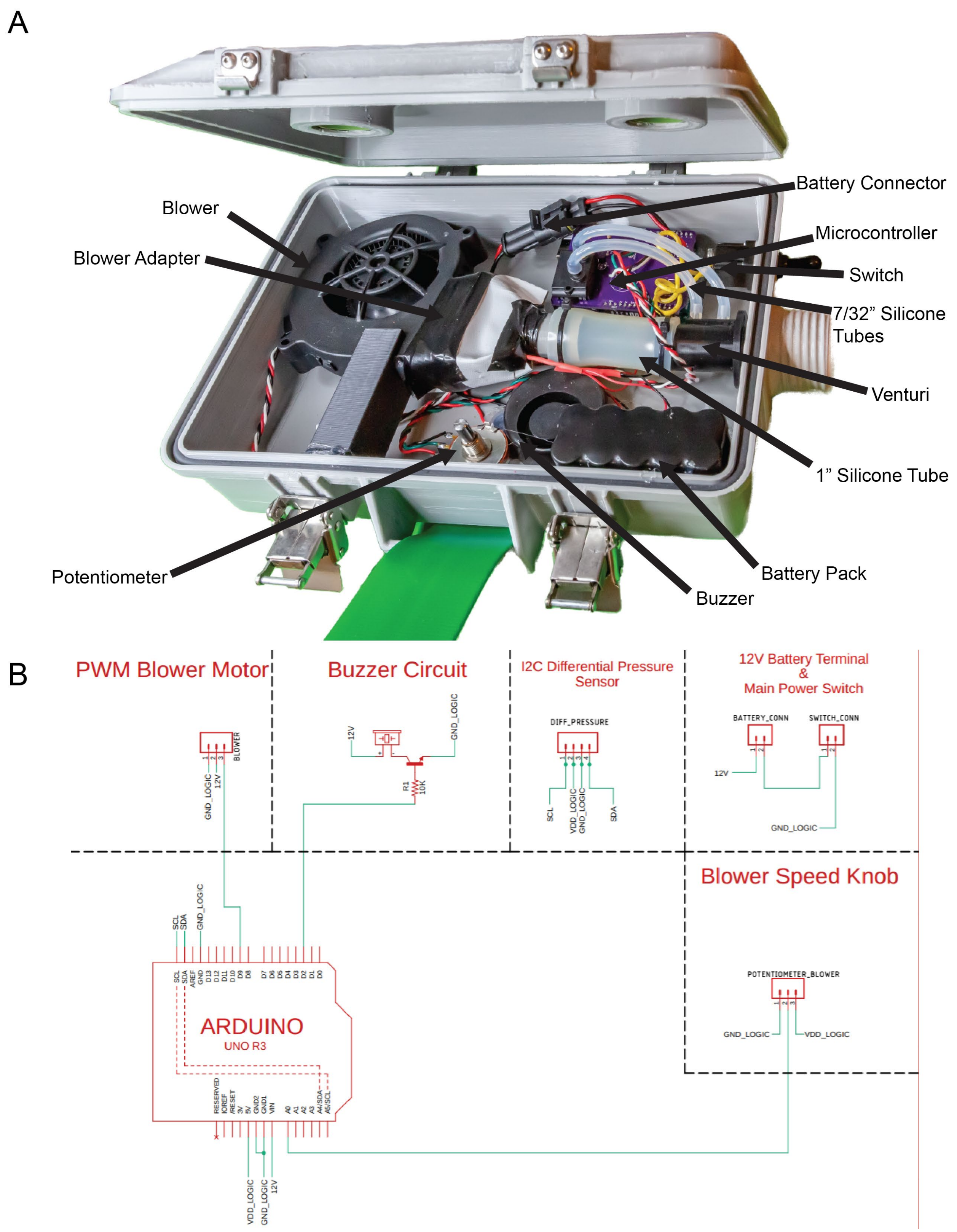


© President and Fellows of Harvard College and The Massachusetts Institute of Technology, 2020.

**Figure 2:** PanFab blower unit. **A)** Components inside the PAPR blower unit. **B)** Circuit diagram of the blower unit components.

The housing for the components of the blower units requires an airtight seal and needs to be compatible with the filter cartridge connectors and the facepiece hose connector. We designed two housings, the PanFab Commercial Design blower unit housing uses a commercially available container, modified to connect to the filter cartridges and the facepiece hose, and the PanFab Custom Design blower unit housing is fabricated entirely using 3D-printing or injection molding methods. Both housings are compatible with both of the PanFab-designed filter cartridges, as well as ILC Dover (Frederica, DE) filter cartridges via 40mm NATO threaded connections.

*PanFab Custom blower units housing*

The blower, battery, and all the electronics were enclosed in a custom housing made of PET-G designed for 3D printing and also optimized for injection molding. 3D printing is advantageous in this setting because it is widely available and compatible with low-volume production; injection molding requires a relatively expensive mold but has the advantage of lower unit cost and rapid production for higher volume production. The housing was printed using Fused Deposition Modeling (FDM) printing, on a RailCore II 300ZLT 3D printer. Layer height of 300 micron was used with 15% rectilinear infill and with 4 vertical shells. Integrated industry-standard female NATO 40 mm threaded connections enable attachment of the two filter cartridges and a male NATO 40 mm threaded connection enables attachment of a thermoplastic polyurethane hose to the housing while maintaining an airtight seal. Custom adapters were designed to attach hose to the NATO threaded outlet on one end and to the facepiece on the other

© President and Fellows of Harvard College and The Massachusetts Institute of Technology, 2020.

end. The hose adapters are threaded into the hose to form a tight, interference fit. All NATO 40mm female connection features incorporate a gasket seat, which accommodates an ethylene propylene diene monomer (EPDM) 1/4” cam-and-groove gasket (purchased from McMaster-Carr, part number 5647K62). **Figure 3** below shows the connection of the hose to the housing outlet, through the hose adapter.

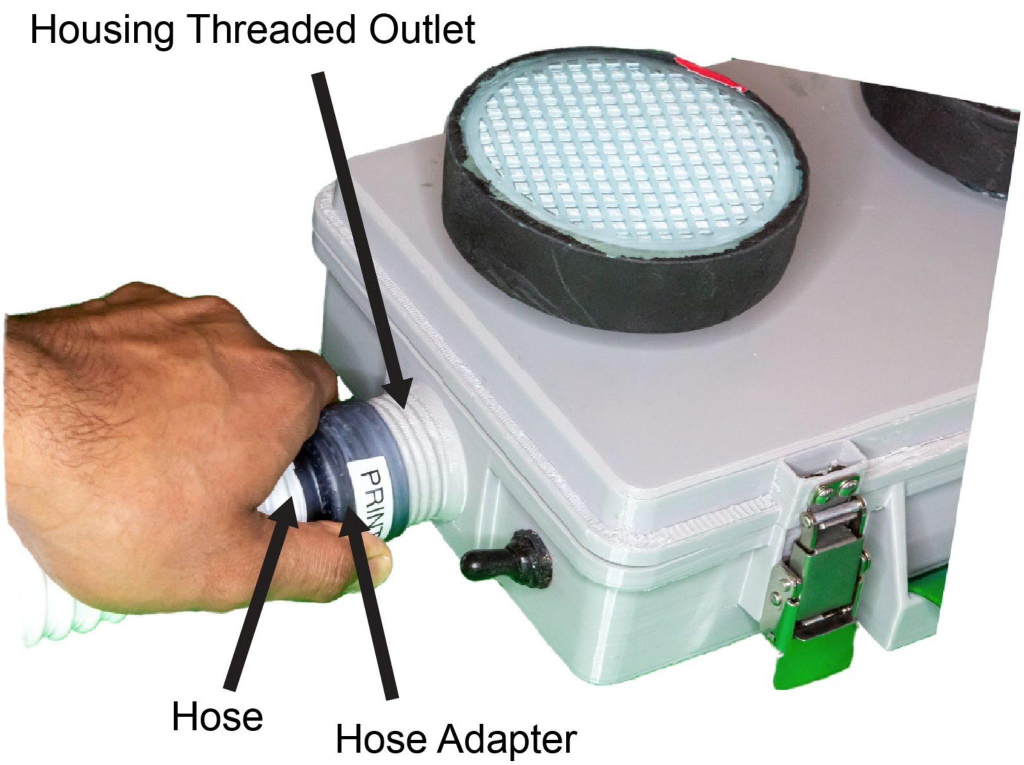


**Figure 3:** Hose connection to PanFab blower unit outlet.

The housing is made of two detachable parts- a “lid” that closes over a “bin” and is locked with 4 draw latches (purchased from McMaster-Carr, part number 1794A55). The bin has a groove feature along its perimeter to accommodate a 1/8” silicone O-ring. The lid has a tongue profile that fits inside the groove and compresses the O-ring, providing sealing. In our prototypes, we used O-ring cord stock (purchased from McMaster-Carr, part number 5229T51), which was cut to size and glued at ends with silicone-based adhesive. For production parts, O-

© President and Fellows of Harvard College and The Massachusetts Institute of Technology, 2020.

rings of the required diameter (according to total perimeter of the groove, as per CAD models in **Supplementary Material 4**) can be molded. **Figures 4A** and **4B** below shows the parts of the housing and the sealing features respectively.

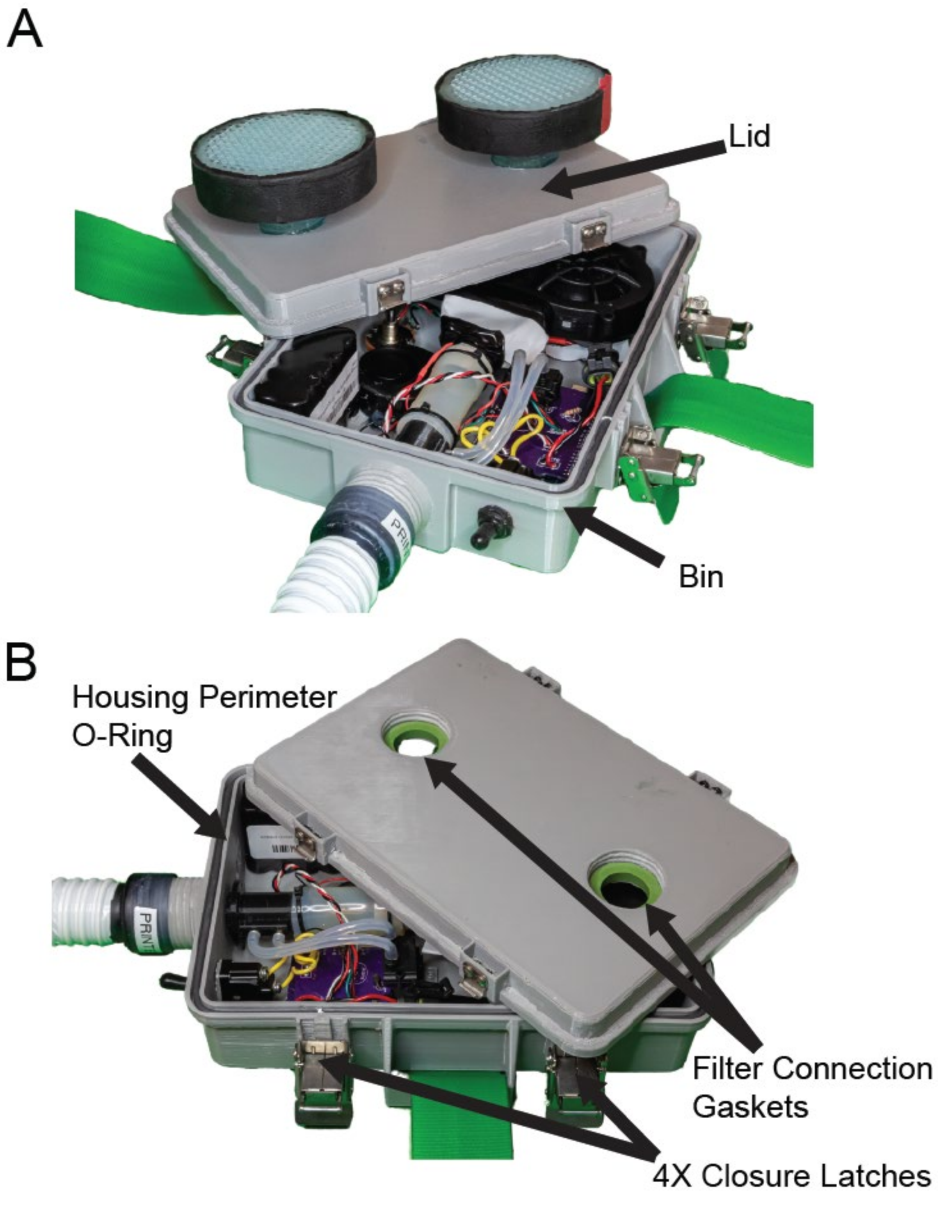


**Figure 4:** PanFab blower unit. **A)** Custom blower unit housing and **B)** sealing features.

The housing is printed with a hole to accommodate a MIL-spec power switch which connects/disconnects the battery pack to the rest of the circuit. Sealing between the switch and

© President and Fellows of Harvard College and The Massachusetts Institute of Technology, 2020.

the housing is achieved by screwing a switch cover (purchased from McMaster-Carr, part number 70205K4) over the switch toggle. The base of the cover forms a seal with the housing surface upon tightening, which is kept from loosening by applying Loctite Threadlocker Red 271 (Loctite, Hartford, CT) on the threads during the cover installation.

A waist belt, designated as PathoShield Gait Belt, purchased from Skil-Care (Yonkers, NY) is run through two integrated loops in the bin part of the housing, which allows donning the blower unit on the lower back position. The belt is specifically manufactured to be amenable to sterilization.

CAD files for the housing are available in **Supplementary Material 4.**

*PanFab Commercial blower unit housing*

Since some groups may not have ready access to sufficiently large 3D printers, we have also adapted a commercially molded plastic case, the Pelican V100 Vault Small Pistol Case (Pelican products, Torrance, CA). Four holes of different sizes are made in this case, two of which accommodate and seal with two filter connection inserts, one accommodates and seals with a hose insert, and one with the power switch. These custom designed inserts are also compatible with both 3D printing and injection molding. These inserts enable connecting the housing to the filter and hose through the NATO 40mm threaded connectors, as in the case of the custom housing. The switch is also installed and sealed in a similar way. The inserts would be plastic welded onto the Pelican case, however in our prototypes, we used cyanoacrylate to seal the inserts to the case. The Pelican case comes with a pre-installed perimeter gasket, which provides the required sealing. Finally, the as-bought case also has an equalization valve, which in

© President and Fellows of Harvard College and The Massachusetts Institute of Technology, 2020.

its normal use prevents buildup of moisture inside the case, which is sealed off first using cyanoacrylate, and then hot melt glue over it, for its use as a PAPR.

Two loops of the Skil-Care belts are separately printed or molded and then welded (glued with cyanoacrylate in the prototype) to the case.

Facepiece integration

We used our prototypes with the loose-fitting UW VHA ADAPT PAPR Hood. The facepiece has a circular hole at the back of the head, to allow for connection with the hose. The hose adapter on the facepiece side was screwed over a hollow hood coupler, which passes through the hole in the facepiece. A tight connection between the facepiece material and the hood coupler was achieved by engaging a locking ring, which has a locking feature and locks with the hood coupler upon twisting, pressing the facepiece between them. **Figure 5** below shows this connection. The PAPR designs discussed in this paper are also compatible with other facepieces with the NATO 40 mm threaded connection. Furthermore, the design of the hood coupler can be modified as needed, to make it compatible with any other facepiece.

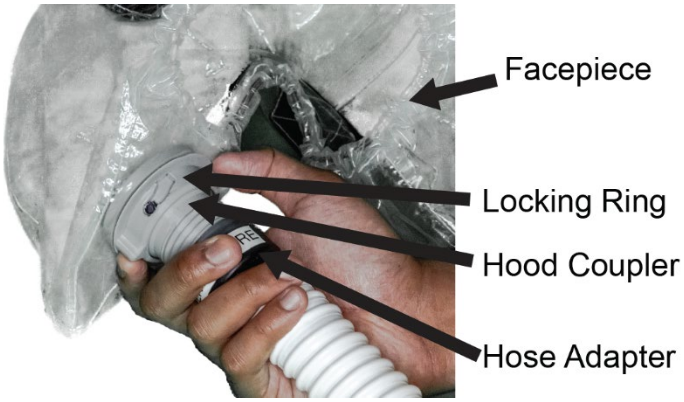


**Figure 5:** Connection of the hose with facepiece, through hose adapter, hood coupler, and locking ring.

© President and Fellows of Harvard College and The Massachusetts Institute of Technology, 2020.


© President and Fellows of Harvard College and The Massachusetts Institute of Technology, 2020.
