## Supplementary Material 4 for "De novo Powered Air-Purifying Respirator Design and Fabrication for Pandemic Response": 3D Printing Instructions.pdf

**3D Printing Instructions for PanFab PAPR Parts**

Sherry H. Yu: 0000-0002-1432-9128
Nicole R. LeBoeuf, MD, MPH, 0000-0002-8264-834X

Peter Sorger, PhD, 0000-0002-3364-1838

### INTRODUCTION

The PanFab PAPR unit was designed in response to the personal protective equipment (PPE) shortages caused by the COVID-19 pandemic. As such, viable tested prototypes were printed in low-cost, readily available materials such as Polyethylene terephthalate – glycol (PETG) polyester polymer on a high-speed fused deposition modeling (FDM) printer. PETG was chosen as the prime material candidate based on preliminary testing for layer adhesion, total part strength, impact resistance, ease of printing, and viability of post-processing. MSLA Resin printing was used to fabricate smaller parts with tighter tolerances. The primary resin used was SirayaTech Blu resin (polyurethane – acrylate copolymer).

While all the prototypes for PanFab Custom and Commercial PAPR's used in design validation testing were 3D printed, all the parts have been optimized for injection molding for high volume production.

This document presents printing settings that were used by the PanFab team in prototyping the Custom PAPR and Commercial PAPR designs. Note that the settings and material should be optimized for the specific printer used in printing the parts.

### PANFAB CUSTOM DESIGN PAPR HOUSING

The PanFab PAPR Custom Design housing has the following components:

- Lid, has integrated female NATO 40 mm threaded connecter for filters
- Bin, which attaches to it all the components to power and control the blower

<u>Bin with Gasket Recess</u>

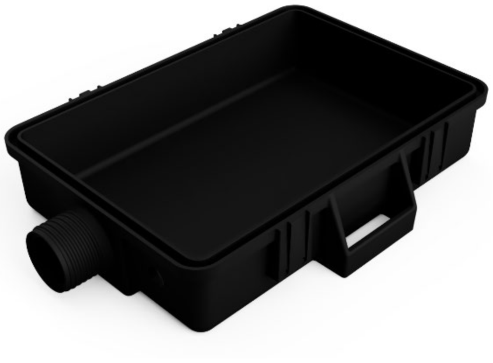

The housing parts and Milwaukee Filter Cartridge Cover were printed on a RailCore II 300ZLT FDM 3D printer. The "bin" and "lid" parts must be nominally air-tight, so all air handled by the device must first pass through filters. The printed part can maintain function even without dense infill, but top and bottom layers must be increased, and slight over extrusion (or top layer "ironing" is recommended). Layer height of 300 microns was used with 15% rectilinear infill. Increase vertical shells to 4 for increased part strength. This part is designed to minimize supports, but still requires support on the gasket seal overhang, and threaded outlet. The threads on the outlet *will* need to be manually finished, and this can be accomplished with a cylindrical file (or m3

© President and Fellows of Harvard College and The Massachusetts Institute of Technology, 2020.

threaded rod, if necessary). Additional air tightness can be provided with a conformal coating of resin or other sealant. Print in default orientation with the interior of the part facing upwards.

Lid with Tongue Feature
Settings similar to "Bottom with Gasket Recess." Print with the surface visible in the figure to the right on the build plate (larger opening on threaded holes toward build plate)

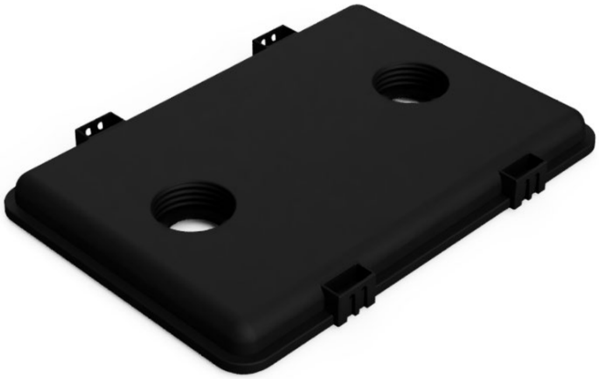

**COMMERCIAL MILWAUKEE FILTER CARTRIDGE COVER**

This part covers and protects the filter media of the Milwaukee commercial filter cartridge from damage. Print this part with flat surface on build plate, 100% infill, two walls. This is not a structural part, and therefore can be printed much thinner than either the bottom with gasket recess or the lid.

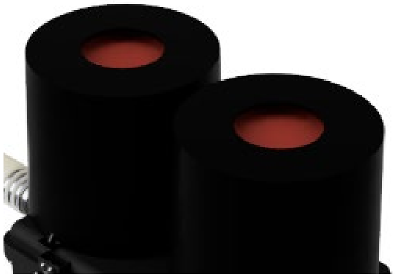

**ALL OTHER PARTS**

In addition to the parts discussed, there are several smaller parts such as the hose adapter, hood coupler, locking ring, filter inserts, hose insert, blower adapter, blower mounts, and belt loops. Print these in resin for optimal air tightness. Resin used was SirayaTech Blu, on Anycubic LCD 3D printers (Photon S and Mono X). The resin must be optimized for the printer being used. Parts can also be printed in FDM, but this was not tested by our team for NIOSH-equivalent tests.

© President and Fellows of Harvard College and The Massachusetts Institute of Technology, 2020.
