## Supplementary Material 4 for "De novo Powered Air-Purifying Respirator Design and Fabrication for Pandemic Response": sdp3x_default_address.ino

```
#include <Wire.h>

#include <sdpsensor.h>

// SDP3x on the default I2C address of 0x21:
SDP3XSensor sdp;

void setup() {
  Wire.begin();
  Serial.begin(9600);
  delay(1000); // let serial console settle

  int ret = sdp.init();
  if (ret == 0) {
    Serial.print("init(): success\n");
  } else {
    Serial.print("init(): failed, ret = ");
    Serial.println(ret);
    while (true) {
      delay(1000);
    }
  }
}

void loop() {
  int ret = sdp.readSample();
  if (ret == 0) {
    Serial.print("Differential pressure: ");
    Serial.print(sdp.getDifferentialPressure());
    Serial.print("Pa | ");

    Serial.print("Temp: ");
    Serial.print(sdp.getTemperature());
    Serial.print("C\n");
  } else {
    Serial.print("Error in readSample(), ret = ");
    Serial.println(ret);
  }

  delay(500);
}
```
