## Supplementary Material 4 for "De novo Powered Air-Purifying Respirator Design and Fabrication for Pandemic Response": sdp3x_manual_address.ino

```
#include <Wire.h>

#include <sdpsensor.h>

// To configure the I2C address manually, choose one of the three calls below;
// make sure you comment out the other two, otherwise the sketch won't build!

SDP3XSensor sdp(SDP3XSensor::SDP3X_I2C_ADDR_21); // Address 0x21
// SDP3XSensor sdp(SDP3XSensor::SDP3X_I2C_ADDR_22); // Address 0x22
// SDP3XSensor sdp(SDP3XSensor::SDP3X_I2C_ADDR_23); // Address 0x23

void setup() {
  Wire.begin();
  Serial.begin(9600);
  delay(1000); // let serial console settle
  do {
    int ret = sdp.init();
    if (ret == 0) {
      Serial.print("init(): success\n");
      break;
    } else {
      Serial.print("init(): failed, ret = ");
      Serial.println(ret);
      delay(1000);
    }
  } while(true);
}

void loop() {
  int ret = sdp.readSample();
  if (ret == 0) {
    Serial.print("Differential pressure: ");
    Serial.print(sdp.getDifferentialPressure());
    Serial.print("Pa | ");

    Serial.print("Temp: ");
    Serial.print(sdp.getTemperature());
    Serial.print("C\n");
  } else {
    Serial.print("Error in readSample(), ret = ");
    Serial.println(ret);
  }

  delay(500);
}
```
