## Supplementary Material 4 for "De novo Powered Air-Purifying Respirator Design and Fabrication for Pandemic Response": i2chelper.cpp

```
/*
 *  Copyright (c) 2015-2018, Sensirion AG <>
 *  All rights reserved.
 *
 *  Redistribution and use in source and binary forms, with or without
 *  modification, are permitted provided that the following conditions are met:
 *      * Redistributions of source code must retain the above copyright
 *        notice, this list of conditions and the following disclaimer.
 *      * Redistributions in binary form must reproduce the above copyright
 *        notice, this list of conditions and the following disclaimer in the
 *        documentation and/or other materials provided with the distribution.
 *      * Neither the name of the Sensirion AG nor the names of its
 *        contributors may be used to endorse or promote products derived
 *        from this software without specific prior written permission.
 *
 *  THIS SOFTWARE IS PROVIDED BY THE COPYRIGHT HOLDERS AND CONTRIBUTORS "AS IS"
 *  AND ANY EXPRESS OR IMPLIED WARRANTIES, INCLUDING, BUT NOT LIMITED TO, THE
 *  IMPLIED WARRANTIES OF MERCHANTABILITY AND FITNESS FOR A PARTICULAR PURPOSE
 *  ARE DISCLAIMED. IN NO EVENT SHALL <COPYRIGHT HOLDER> BE LIABLE FOR ANY
 *  DIRECT, INDIRECT, INCIDENTAL, SPECIAL, EXEMPLARY, OR CONSEQUENTIAL DAMAGES
 *  (INCLUDING, BUT NOT LIMITED TO, PROCUREMENT OF SUBSTITUTE GOODS OR SERVICES;
 *  LOSS OF USE, DATA, OR PROFITS; OR BUSINESS INTERRUPTION) HOWEVER CAUSED AND
 *  ON ANY THEORY OF LIABILITY, WHETHER IN CONTRACT, STRICT LIABILITY, OR TORT
 *  (INCLUDING NEGLIGENCE OR OTHERWISE) ARISING IN ANY WAY OUT OF THE USE OF
 *  THIS SOFTWARE, EVEN IF ADVISED OF THE POSSIBILITY OF SUCH DAMAGE.
*/

#include <Arduino.h>
#include <Wire.h>
#include "i2chelper.h"

int8_t I2CHelper::i2c_read(uint8_t addr, uint8_t* data, uint16_t count)
{
    Wire.requestFrom(addr, count);
    if (Wire.available() != count) {
        return -1;
    }
    for (int i = 0; i < count; ++i) {
        data[i] = Wire.read();
    }
    return 0;
}

int8_t I2CHelper::i2c_write(uint8_t addr, const uint8_t* data, uint16_t count, bool appendCrc)
{
    Wire.beginTransmission(addr);
    for (int i = 0; i < count; ++i) {
        if (Wire.write(data[i]) != 1) {
            return 1;
        }
    }
    if (appendCrc) {
      uint8_t crc = crc8(data, count);
      if (Wire.write(crc) != 1) {
          return 2;
      }
    }

    if (Wire.endTransmission() != 0) {
        return 3;
    }
    return 0;
}

uint8_t I2CHelper::crc8(const uint8_t* data, uint8_t len)
{
  // adapted from SHT21 sample code from http://www.sensirion.com/en/products/humidity-temperature/download-center/

  uint8_t crc = 0xff;
  uint8_t byteCtr;
  for (byteCtr = 0; byteCtr < len; ++byteCtr) {
    crc ^= (data[byteCtr]);
    for (uint8_t bit = 8; bit > 0; --bit) {
      if (crc & 0x80) {
        crc = (crc << 1) ^ 0x31;
      } else {
        crc = (crc << 1);
      }
    }
  }
  return crc;
}
```
