## Supplementary Material 4 for "De novo Powered Air-Purifying Respirator Design and Fabrication for Pandemic Response": README.md

# Arduino library for Sensirion SDP3x and SDP8xx series

This is a library for Sensirion SDP3x and SDP8xx sensors. Both 500Pa
and 125Pa sensor types are supported.

## Installation

1. Download the latest ZIP version from the [github releases page](https://github.com/Sensirion/arduino-sdp/releases).
2. Open Arduino IDE, and select `Sketch > Include Library > Add .ZIP Library`, and select the ZIP file you downloaded in step 2
3. Try out one of the examples from `File > Examples > arduino-sdp` (replace `arduino-sdp` with `arduino-sdp-<version>` if necessary)

## Getting started

To get started, have a look at the examples from `File > Examples > arduino-sdp` (replace `arduino-sdp` with `arduino-sdp-<version>` if necessary):

- For SDP3x on its default I2C address 0x21, use example `sdp3x_default_address`
- For SDP3x on I2C address 0x22 or 0x23, use example  `sdp3x_manual_address`
- For the SDP8xx series, use example `sdp8xx`

### Initializing for SDP3x on I2C address 0x21

I2C address is the default for the SDP3x series. The `sdp_generic` sample
will do that by default, without requiring any changes.

### Initializing for SDP3x on an I2C address other than 0x21

To use an address other than 0x21, you can pass an argument when setting up
the sensor. In the example, comment out line 6, and uncomment line 10, like so:

```c++
// SDP3XSensor sdp;

// If your SDP3x is not using the default I2C address of 0x21, uncomment the
// line below:
SDP3XSensor sdp(SDP3X_I2C_ADDR_22);
```
Alternatively, you can use `SDP3X_I2C_ADDR_23` instead of `SDP3X_I2C_ADDR_22`

### Initializing for SDP8xx

To use it with an SDP8xx series sensor, comment out line 6, and uncomment
line 14, like so:

```c++
// SDP3XSensor sdp;

// If your SDP3x is not using the default I2C address of 0x21, uncomment the
// line below:
// SDP3XSensor sdp(SDP3X_I2C_ADDR_22);


// If you're using an SDP8xx, uncomment the line below instead:
SDP8XXSensor sdp;
```


### API

There are just four functions to interface with the sensor:


#### int init()
Use this to initialize the sensor. It will return 0 if everything went fine, and a non-zero value otherwise. Success when calling init() means that the sensor is connected correctly, and responds to SDPXXX I2C commands.

##### Usage
```c++
int ret = sdp.init();
if (ret == 0) {
    Serial.print("init(): success\n");
} else {
    Serial.print("init(): failed, ret = ");
    Serial.println(ret);
}
```

#### int readSample()
Use this to read a new data sample - differential pressure and temperature - from the sensor. It will return 0 on success, and a non-zero value otherwise.

##### Usage
```c++
int ret = sdp.readSample();
if (ret == 0) {
   // Success; go on to read out differential pressure and temperature
} else {
    Serial.print("Error in readSample(), ret = ");
    Serial.println(ret);
}
```
#### float getDifferentialPressure()
Use this to read the differential pressure value in Pascal (Pa) read from the sensor in the last `readSample()` call. Note that `readSample()` must be called before calling `getDifferentialPressure()`, otherwise the the value will be outdated at best, and invalid at worst.

##### Usage
```c++
float dp = sdp.getDifferentialPressure();
```

#### float getTemperature()
Use this to read the temperature in degrees celcius value read from the sensor in the last `readSample()` call. Note that `readSample()` must be called before calling `getTemperature()`, otherwise the the value will be outdated at best, and invalid at worst.

##### Usage
```c++
float temp = sdp.getTemperature();
```
