## Supplementary Material 4 for "De novo Powered Air-Purifying Respirator Design and Fabrication for Pandemic Response": sdpsensor.cpp

```
/*
 *  Copyright (c) 2018, Sensirion AG <>
 *  All rights reserved.
 *
 *  Redistribution and use in source and binary forms, with or without
 *  modification, are permitted provided that the following conditions are met:
 *      * Redistributions of source code must retain the above copyright
 *        notice, this list of conditions and the following disclaimer.
 *      * Redistributions in binary form must reproduce the above copyright
 *        notice, this list of conditions and the following disclaimer in the
 *        documentation and/or other materials provided with the distribution.
 *      * Neither the name of the Sensirion AG nor the names of its
 *        contributors may be used to endorse or promote products derived
 *        from this software without specific prior written permission.
 *
 *  THIS SOFTWARE IS PROVIDED BY THE COPYRIGHT HOLDERS AND CONTRIBUTORS "AS IS"
 *  AND ANY EXPRESS OR IMPLIED WARRANTIES, INCLUDING, BUT NOT LIMITED TO, THE
 *  IMPLIED WARRANTIES OF MERCHANTABILITY AND FITNESS FOR A PARTICULAR PURPOSE
 *  ARE DISCLAIMED. IN NO EVENT SHALL <COPYRIGHT HOLDER> BE LIABLE FOR ANY
 *  DIRECT, INDIRECT, INCIDENTAL, SPECIAL, EXEMPLARY, OR CONSEQUENTIAL DAMAGES
 *  (INCLUDING, BUT NOT LIMITED TO, PROCUREMENT OF SUBSTITUTE GOODS OR SERVICES;
 *  LOSS OF USE, DATA, OR PROFITS; OR BUSINESS INTERRUPTION) HOWEVER CAUSED AND
 *  ON ANY THEORY OF LIABILITY, WHETHER IN CONTRACT, STRICT LIABILITY, OR TORT
 *  (INCLUDING NEGLIGENCE OR OTHERWISE) ARISING IN ANY WAY OUT OF THE USE OF
 *  THIS SOFTWARE, EVEN IF ADVISED OF THE POSSIBILITY OF SUCH DAMAGE.
*/

#include <Arduino.h>

#include "sdpsensor.h"
#include "i2chelper.h"

int SDPSensor::init()
{
  // try to read product id
  const uint8_t CMD_LEN = 2;
  uint8_t cmd0[CMD_LEN] = { 0x36, 0x7C };
  uint8_t cmd1[CMD_LEN] = { 0xE1, 0x02 };

  const uint8_t DATA_LEN = 18;
  uint8_t data[DATA_LEN] = { 0 };

  uint8_t ret = I2CHelper::i2c_write(mI2CAddress, cmd0, CMD_LEN);
  if (ret != 0) {
    return 1;
  }
  ret = I2CHelper::i2c_write(mI2CAddress, cmd1, CMD_LEN);
  if (ret != 0) {
    return 2;
  }
  ret = I2CHelper::i2c_read(mI2CAddress, data, DATA_LEN);
  if (ret != 0) {
    return 3;
  }

  // at this point, we don't really care about the data just yet, but
  // we may use that in the future. Either way, the sensor responds, and
  return 0;
}

int SDPSensor::readSample()
{
  const uint8_t CMD_LEN = 2;
  uint8_t cmd[CMD_LEN] = { 0x36, 0x2F };

  const uint8_t DATA_LEN = 9;
  uint8_t data[DATA_LEN] = { 0 };

  if (I2CHelper::i2c_write(mI2CAddress, cmd, CMD_LEN) != 0) {
    return 1;
  }

  delay(100); // theoretically 45ms

  if (I2CHelper::i2c_read(mI2CAddress, data, DATA_LEN) != 0) {
    return 2;
  }

  // TODO: check CRC

  int16_t dp_raw   = (int16_t)data[0] << 8 | data[1];
  int16_t temp_raw = (int16_t)data[3] << 8 | data[4];
  int16_t dp_scale  = (int16_t)data[6] << 8 | data[7];

  mDifferentialPressure = dp_raw / (float)dp_scale;
  mTemperature = temp_raw / 200.0;

  return 0;
}

float SDPSensor::getDifferentialPressure() const
{
  return mDifferentialPressure;
}

float SDPSensor::getTemperature() const
{
  return mTemperature;
}
```
