## Supplementary Material 4 for "De novo Powered Air-Purifying Respirator Design and Fabrication for Pandemic Response": sdpsensor.h

```
/*
 *  Copyright (c) 2018, Sensirion AG <>
 *  All rights reserved.
 *
 *  Redistribution and use in source and binary forms, with or without
 *  modification, are permitted provided that the following conditions are met:
 *      * Redistributions of source code must retain the above copyright
 *        notice, this list of conditions and the following disclaimer.
 *      * Redistributions in binary form must reproduce the above copyright
 *        notice, this list of conditions and the following disclaimer in the
 *        documentation and/or other materials provided with the distribution.
 *      * Neither the name of the Sensirion AG nor the names of its
 *        contributors may be used to endorse or promote products derived
 *        from this software without specific prior written permission.
 *
 *  THIS SOFTWARE IS PROVIDED BY THE COPYRIGHT HOLDERS AND CONTRIBUTORS "AS IS"
 *  AND ANY EXPRESS OR IMPLIED WARRANTIES, INCLUDING, BUT NOT LIMITED TO, THE
 *  IMPLIED WARRANTIES OF MERCHANTABILITY AND FITNESS FOR A PARTICULAR PURPOSE
 *  ARE DISCLAIMED. IN NO EVENT SHALL <COPYRIGHT HOLDER> BE LIABLE FOR ANY
 *  DIRECT, INDIRECT, INCIDENTAL, SPECIAL, EXEMPLARY, OR CONSEQUENTIAL DAMAGES
 *  (INCLUDING, BUT NOT LIMITED TO, PROCUREMENT OF SUBSTITUTE GOODS OR SERVICES;
 *  LOSS OF USE, DATA, OR PROFITS; OR BUSINESS INTERRUPTION) HOWEVER CAUSED AND
 *  ON ANY THEORY OF LIABILITY, WHETHER IN CONTRACT, STRICT LIABILITY, OR TORT
 *  (INCLUDING NEGLIGENCE OR OTHERWISE) ARISING IN ANY WAY OUT OF THE USE OF
 *  THIS SOFTWARE, EVEN IF ADVISED OF THE POSSIBILITY OF SUCH DAMAGE.
*/

#ifndef SDPSENSOR_H
#define SDPSENSOR_H

class SDPSensor
{
public:
  const static int SDP3X_I2C_ADDR_21   = 0x21;
  const static int SDP3X_I2C_ADDR_22   = 0x22;
  const static int SDP3X_I2C_ADDR_23   = 0x23;
  const static int SDP3X_I2C_ADDR_DEFAULT = SDP3X_I2C_ADDR_21;

  const static int SDP8XX_I2C_ADDR_DEFAULT = 0x25;

  SDPSensor(uint8_t i2cAddr) : mI2CAddress(i2cAddr) {}

  /**
   * initialize the sensor
   * @return 0 on sucess, error code otherwise
   */
  int init();

  /**
   * read sensor data from sensor
   * @return 0 on success, error code otherwise
   */
  int readSample();

  /**
   * Returns the last differential pressure value read - does NOT trigger a new measurement
   * @return last differential pressure value read
   */
  float getDifferentialPressure() const;

  /**
   * Returns the last temperature value read - does NOT trigger a new measurement
   * @return last temperature value read
   */
  float getTemperature()  const;

private:
  uint8_t mI2CAddress;

  float mDifferentialPressure;
  float mTemperature;
};

class SDP3XSensor : public SDPSensor
{
public:
  SDP3XSensor() : SDPSensor(SDPSensor::SDP3X_I2C_ADDR_DEFAULT) {}
  SDP3XSensor(uint8_t i2cAddr) : SDPSensor(i2cAddr) {}
};

class SDP8XXSensor : public SDPSensor
{
public:
  SDP8XXSensor() : SDPSensor(SDPSensor::SDP8XX_I2C_ADDR_DEFAULT) {}
};

#endif /* SDPSENSOR_H */
```
