## Supplementary Material 4 for "De novo Powered Air-Purifying Respirator Design and Fabrication for Pandemic Response": BlowerPWM.ino

```
#include <SoftPWM.h>

// Set up Arduino pin 9 as PWM channel 0:
// (For examples on how to set this see:
//  File > Examples > arduino-softpwm > SoftPWM_example )
SOFTPWM_DEFINE_CHANNEL(0, DDRB, PORTB, PORTB1);  //Arduino pin 9

/* Here you make an instance of desired channel counts you want
   with the default 256 PWM levels (0 ~ 255). */
SOFTPWM_DEFINE_OBJECT(1);

/* If you want to use the SoftPWM object outside where it's defined,
   add the following line to the file. */
SOFTPWM_DEFINE_EXTERN_OBJECT(1);

// set analog input pin
int analogInputPin = A0;

void setup() {
  Serial.begin(9600);

  // begin with 100hz pwm frequency
  Palatis::SoftPWM.begin(100);
}

void loop() {

  long inputVal = analogRead(analogInputPin);

  // CONVERSION:
  // - analogRead() returns a value from 0..1023
  // - SoftPWM takes values from 0..255
  // - blower reaches max speed at 90%
  // - probably want to have a minimum blower speed too: minPWM
  // So, convert inputVal range from 0..1023 to minPWM..230
  const long minPWM = 64; // set min to be 25% of 255
  const long maxPWM = 230; // set max to be 90% of 255
  long PWMLevel = minPWM + (inputVal * (maxPWM-minPWM)) / (1023);

  // set pin 9 (pwm channel 0) to new PWM level
  Palatis::SoftPWM.set(0, PWMLevel);
  Serial.println(PWMLevel); // debugging

  delay(10); // no need to respond to user input faster than ~100Hz
}
```
