## Supplementary Material 4 for "De novo Powered Air-Purifying Respirator Design and Fabrication for Pandemic Response": SoftPWM_example.ino

```
/*
  Arduino-SoftPWM: a software PWM library for Arduino
  Copyright 2016, Victor Tseng <>

  All rights reserved.

  Redistribution and use in source and binary forms, with or without
  modification, are permitted provided that the following conditions
  are met:

  1. Redistributions of source code must retain the above copyright
     notice, this list of conditions and the following disclaimer.

  2. Redistributions in binary form must reproduce the above copyright
     notice, this list of conditions and the following disclaimer in the
     documentation and/or other materials provided with the distribution.

  3. Neither the name of the copyright holder nor the names of its
     contributors may be used to endorse or promote products derived
     from this software without specific prior written permission.

  THIS SOFTWARE IS PROVIDED BY THE COPYRIGHT HOLDERS AND CONTRIBUTORS "AS
  IS" AND ANY EXPRESS OR IMPLIED WARRANTIES, INCLUDING, BUT NOT LIMITED TO,
  THE IMPLIED WARRANTIES OF MERCHANTABILITY AND FITNESS FOR A PARTICULAR
  PURPOSE ARE DISCLAIMED. IN NO EVENT SHALL THE COPYRIGHT HOLDER OR
  CONTRIBUTORS BE LIABLE FOR ANY DIRECT, INDIRECT, INCIDENTAL, SPECIAL,
  EXEMPLARY, OR CONSEQUENTIAL DAMAGES (INCLUDING, BUT NOT LIMITED TO,
  PROCUREMENT OF SUBSTITUTE GOODS OR SERVICES; LOSS OF USE, DATA, OR PROFITS;
  OR BUSINESS INTERRUPTION) HOWEVER CAUSED AND ON ANY THEORY OF LIABILITY,
  WHETHER IN CONTRACT, STRICT LIABILITY, OR TORT (INCLUDING NEGLIGENCE OR
  OTHERWISE) ARISING IN ANY WAY OUT OF THE USE OF THIS SOFTWARE, EVEN IF
  ADVISED OF THE POSSIBILITY OF SUCH DAMAGE.
*/

#include <SoftPWM.h>

/* pins_arduino.h defines the pin-port/bit mapping as PROGMEM so
   you have to read them with pgm_read_xxx(). That's generally okay
   for ordinary use, but really bad when you're writing super fast
   codes because the compiler doesn't treat them as constants and
   cannot optimize them away with sbi/cbi instructions.

   Therefore we have to tell the compiler the PORT and BIT here.
   Hope someday we can find a way to workaround this.

   Check the manual of your MCU for port/bit mapping.

   The following example demonstrates setting channels for all pins
   on the ATmega328P or ATmega168 used on Arduino Uno, Pro Mini,
   Nano and other boards. */
SOFTPWM_DEFINE_CHANNEL(0, DDRD, PORTD, PORTD0);  //Arduino pin 0
SOFTPWM_DEFINE_CHANNEL(1, DDRD, PORTD, PORTD1);  //Arduino pin 1
SOFTPWM_DEFINE_CHANNEL(2, DDRD, PORTD, PORTD2);  //Arduino pin 2
SOFTPWM_DEFINE_CHANNEL(3, DDRD, PORTD, PORTD3);  //Arduino pin 3
SOFTPWM_DEFINE_CHANNEL(4, DDRD, PORTD, PORTD4);  //Arduino pin 4
SOFTPWM_DEFINE_CHANNEL(5, DDRD, PORTD, PORTD5);  //Arduino pin 5
SOFTPWM_DEFINE_CHANNEL(6, DDRD, PORTD, PORTD6);  //Arduino pin 6
SOFTPWM_DEFINE_CHANNEL(7, DDRD, PORTD, PORTD7);  //Arduino pin 7
SOFTPWM_DEFINE_CHANNEL(8, DDRB, PORTB, PORTB0);  //Arduino pin 8
SOFTPWM_DEFINE_CHANNEL(9, DDRB, PORTB, PORTB1);  //Arduino pin 9
SOFTPWM_DEFINE_CHANNEL(10, DDRB, PORTB, PORTB2);  //Arduino pin 10
SOFTPWM_DEFINE_CHANNEL(11, DDRB, PORTB, PORTB3);  //Arduino pin 11
SOFTPWM_DEFINE_CHANNEL(12, DDRB, PORTB, PORTB4);  //Arduino pin 12
SOFTPWM_DEFINE_CHANNEL(13, DDRB, PORTB, PORTB5);  //Arduino pin 13
SOFTPWM_DEFINE_CHANNEL(14, DDRC, PORTC, PORTC0);  //Arduino pin A0
SOFTPWM_DEFINE_CHANNEL(15, DDRC, PORTC, PORTC1);  //Arduino pin A1
SOFTPWM_DEFINE_CHANNEL(16, DDRC, PORTC, PORTC2);  //Arduino pin A2
SOFTPWM_DEFINE_CHANNEL(17, DDRC, PORTC, PORTC3);  //Arduino pin A3
SOFTPWM_DEFINE_CHANNEL(18, DDRC, PORTC, PORTC4);  //Arduino pin A4
SOFTPWM_DEFINE_CHANNEL(19, DDRC, PORTC, PORTC5);  //Arduino pin A5


/* Or you may want inverted outputs: */
/*
  SOFTPWM_DEFINE_CHANNEL_INVERT(0, DDRD, PORTD, PORTD0);  //Arduino pin 0
  SOFTPWM_DEFINE_CHANNEL_INVERT(1, DDRD, PORTD, PORTD1);  //Arduino pin 1
  SOFTPWM_DEFINE_CHANNEL_INVERT(2, DDRD, PORTD, PORTD2);  //Arduino pin 2
  SOFTPWM_DEFINE_CHANNEL_INVERT(3, DDRD, PORTD, PORTD3);  //Arduino pin 3
  SOFTPWM_DEFINE_CHANNEL_INVERT(4, DDRD, PORTD, PORTD4);  //Arduino pin 4
  SOFTPWM_DEFINE_CHANNEL_INVERT(5, DDRD, PORTD, PORTD5);  //Arduino pin 5
  SOFTPWM_DEFINE_CHANNEL_INVERT(6, DDRD, PORTD, PORTD6);  //Arduino pin 6
  SOFTPWM_DEFINE_CHANNEL_INVERT(7, DDRD, PORTD, PORTD7);  //Arduino pin 7
  SOFTPWM_DEFINE_CHANNEL_INVERT(8, DDRB, PORTB, PORTB0);  //Arduino pin 8
  SOFTPWM_DEFINE_CHANNEL_INVERT(9, DDRB, PORTB, PORTB2);  //Arduino pin 9
  SOFTPWM_DEFINE_CHANNEL_INVERT(10, DDRB, PORTB, PORTB2);  //Arduino pin 10
  SOFTPWM_DEFINE_CHANNEL_INVERT(11, DDRB, PORTB, PORTB3);  //Arduino pin 11
  SOFTPWM_DEFINE_CHANNEL_INVERT(12, DDRB, PORTB, PORTB4);  //Arduino pin 12
  SOFTPWM_DEFINE_CHANNEL_INVERT(13, DDRB, PORTB, PORTB5);  //Arduino pin 13
  SOFTPWM_DEFINE_CHANNEL_INVERT(14, DDRC, PORTC, PORTC0);  //Arduino pin A0
  SOFTPWM_DEFINE_CHANNEL_INVERT(15, DDRC, PORTC, PORTC1);  //Arduino pin A1
  SOFTPWM_DEFINE_CHANNEL_INVERT(16, DDRC, PORTC, PORTC2);  //Arduino pin A2
  SOFTPWM_DEFINE_CHANNEL_INVERT(17, DDRC, PORTC, PORTC3);  //Arduino pin A3
  SOFTPWM_DEFINE_CHANNEL_INVERT(18, DDRC, PORTC, PORTC4);  //Arduino pin A4
  SOFTPWM_DEFINE_CHANNEL_INVERT(19, DDRC, PORTC, PORTC5);  //Arduino pin A5
*/

/* Here you make an instance of desired channel counts you want
   with the default 256 PWM levels (0 ~ 255). */
//SOFTPWM_DEFINE_OBJECT(20);

/* Or you can make one with only 100 PWM levels (0 ~ 99).
   By using less PWM levels, you may be able to use higher
   pwm frequencies. */
SOFTPWM_DEFINE_OBJECT_WITH_PWM_LEVELS(20, 100);

/* If you want to use the SoftPWM object outside where it's defined,
   add the following line to the file. */
//SOFTPWM_DEFINE_EXTERN_OBJECT(16);
SOFTPWM_DEFINE_EXTERN_OBJECT_WITH_PWM_LEVELS(20, 100);

void setup() {
  Serial.begin(19200);

  // begin with 60hz pwm frequency
  Palatis::SoftPWM.begin(60);

  // print interrupt load for diagnostic purposes
  Palatis::SoftPWM.printInterruptLoad();
}

static volatile uint8_t v = 0;
void loop() {
  for (uint8_t i = 0; i < Palatis::SoftPWM.size(); ++i) {
    Serial.print(micros());
    Serial.print(" loop(): ");
    Serial.println(i);

    unsigned long const WAIT = 1000000 / Palatis::SoftPWM.PWMlevels() / 2;
    unsigned long nextMicros = 0;
    for (int v = 0; v < Palatis::SoftPWM.PWMlevels() - 1; ++v) {
      while (micros() < nextMicros);
      nextMicros = micros() + WAIT;
      Palatis::SoftPWM.set(i, v);
    }
    for (int v = Palatis::SoftPWM.PWMlevels() - 1; v >= 0; --v) {
      while (micros() < nextMicros);
      nextMicros = micros() + WAIT;
      Palatis::SoftPWM.set(i, v);
    }
  }
}
```
