## Supplementary Material 4 for "De novo Powered Air-Purifying Respirator Design and Fabrication for Pandemic Response": README.md

arduino-softpwm
===============

Software PWM library for [Arduino](https://arduino.cc/), and other compatible AVR boards.

AVR microcontrollers provide hardware PWM on some pins but if you need PWM on other pins then it must be implemented in software. This library provides easy and efficient software PWM on any pin. Each channel can be set to a different PWM duty cycle.

This library is licensed under [BSD3](https://opensource.org/licenses/BSD-3-Clause)

#### Installation
- Download the most recent version here: https://github.com/Palatis/arduino-softpwm/archive/master.zip
- Using Arduino IDE 1.0.x:
  - Sketch > Import Library... > Add Library... > select the downloaded file > Open
- Using Arduino IDE 1.5+:
  - Sketch > Include Library > Add ZIP Library... > select the downloaded file > Open


#### Usage
See **File > Examples > arduino-softpwm > SoftPWM_example** for demonstration of library usage.

Everything in this library except for preprocessor defines are wrapped in `namespace Palatis {}`, so if your compiler blames you about symbol not found, try add `Palatis::` before that thing. For example, `Palatis::SoftPWM.size()`.

Alternatively, you can add `using namespace Palatis;` at the top of the source file to import everything under the `Palatis` namespace, this might be convenient if you're using my other libraries.

`#define SOFTPWM_OUTPUT_DELAY` - Add this line above `#include <SoftPWM.h>` for a 1 PWM clock cycle delay between outputs to prevent large in-rush currents.

`SOFTPWM_DEFINE_CHANNEL(CHANNEL, PMODE, PORT, BIT)` - Configure a pin for software PWM use. Consult the datasheet for your microcontroller for the appropriate `PORT` and `BIT` values for the physical pin. This information is shown in the pinout diagram, for example: [ATmega328P datasheet](http://www.atmel.com/Images/Atmel-8271-8-bit-AVR-Microcontroller-ATmega48A-48PA-88A-88PA-168A-168PA-328-328P_datasheet_Summary.pdf) **Figure 1-1** found on page 3. If you want to determine the Arduino pin assigned to the physical pin http://www.pighixxx.com/test/pinoutspg/boards/ provides this information for the most popular Arduino boards or you can look at the pins_arduino.h file in the **variant** folder used by your board.
- Parameter: **CHANNEL** - The channel number is used as an ID for the pin.
- Parameter: **PMODE** - DDRx register of the pin's port. Append the port letter to `DDR`. For example: The [Arduino UNO diagram](http://www.pighixxx.com/test/portfolio-items/uno/?portfolioID=314) shows that Arduino pin 13 is **PB5** which means that the port is **B** so you should use the value `DDRB` for that pin.
- Parameter: **PORT** - The port of the pin. For example: Arduino pin 13 is **PB5** so you should use the value `PORTB` for that pin.
- Parameter: **BIT** - The bit of the pin. For example: Arduino pin 13 is **PB5** so you should use the value `PORTB5` for that pin.

`SOFTPWM_DEFINE_CHANNEL_INVERT( CHANNEL, PMODE, PORT, BIT )` - Depending on your application you may prefer to invert the output. See `SOFTPWM_DEFINE_CHANNEL()` for description of parameters.

`SOFTPWM_DEFINE_OBJECT(CHANNEL_CNT)` - Define the softPWM object with the default 256 PWM levels.
- Parameter: **CHANNEL_CNT** - The number of channels that are defined.

`SOFTPWM_DEFINE_OBJECT_WITH_PWM_LEVELS(CHANNEL_CNT, PWM_LEVELS)` - Define the softPWM object with the specified number of PWM levels.
- Parameter: **CHANNEL_CNT** - The number of channels that are defined.
- Parameter: **PWM_LEVELS** - The number of PWM levels. Using less PWM levels may allow a higher PWM frequency. The maximum value is 256.

`SOFTPWM_DEFINE_EXTERN_OBJECT(CHANNEL_CNT)` - Add this if you want to use the SoftPWM object outside where it's defined. See `SOFTPWM_DEFINE_OBJECT()` for description of the parameter.

`SOFTPWM_DEFINE_EXTERN_OBJECT_WITH_PWM_LEVELS(CHANNEL_CNT, PWM_LEVELS)` - Add this if you want to use the SoftPWM object outside where it's defined. See `SOFTPWM_DEFINE_OBJECT_WITH_PWM_LEVELS()` for description of parameters.

`SoftPWM.begin(hertz)` - Initialize SoftPWM.
- Parameter: **hertz** - The PWM frequency. Setting the value too high will cause incorrect operation, too low will cause a visible flicker.
  - Type: long

`SoftPWM.printInterruptLoad()` - Prints diagnostic information to the serial monitor. This can be used to find the optimal PWM frequency by setting different PWM frequency values in begin() and then checking the resulting interrupt load. Calling this function will momentarily turn off the PWM on all channels.

`SoftPWM.set(channel_idx, value)` - Set the PWM level of the given channel.
- Parameter: **channel_idx** - The channel to set.
  - Type: int
- Parameter: **value** - The PWM level to set.
  - Type: byte

`SoftPWM.size()`
- Returns: Number of channels defined.
  - Type: size_t

`SoftPWM.PWMlevels()`
- Returns: The number of PWM levels.
  - Type: unsigned int

`SoftPWM.allOff()` - Set the PWM value of all channels to 0.


#### Troubleshooting
- LEDs have a visible flicker, especially noticeable when the LED is moving relative to the viewer.
  - The PWM frequency set in `SoftPWM.begin()` is too low. Use `SoftPWM.printInterruptLoad()` to determine the optimum PWM frequency. You may be able to achieve a higher PWM frequency by setting less PWM levels with `SOFTPWM_DEFINE_OBJECT_WITH_PWM_LEVELS()` or `SOFTPWM_DEFINE_EXTERN_OBJECT_WITH_PWM_LEVELS()`.
- Erratic PWM operation
  - The interrupt load is too high. Use `SoftPWM.printInterruptLoad()` to determine the interrupt load. You can decrease the interrupt load by setting less PWM levels with `SOFTPWM_DEFINE_OBJECT_WITH_PWM_LEVELS()` or `SOFTPWM_DEFINE_EXTERN_OBJECT_WITH_PWM_LEVELS()` or setting the PWM frequency lower in `SoftPWM.begin()`.
- LED brightness changes between low brightness PWM values are larger than at brighter PWM values.
  - This is caused by the way LEDs work and is not caused by a problem with the library. If possible, use more PWM levels or never allow the LED to get dimmer than the level below which the difference between PWM levels is too distinct.
