## Supplementary Material 4 for "De novo Powered Air-Purifying Respirator Design and Fabrication for Pandemic Response": SoftPWM.h

```
/*
  Arduino-SoftPWM: a software PWM library for Arduino
  Copyright 2016, Victor Tseng <>

  All rights reserved.

  Redistribution and use in source and binary forms, with or without
  modification, are permitted provided that the following conditions
  are met:

  1. Redistributions of source code must retain the above copyright
     notice, this list of conditions and the following disclaimer.

  2. Redistributions in binary form must reproduce the above copyright
     notice, this list of conditions and the following disclaimer in the
     documentation and/or other materials provided with the distribution.

  3. Neither the name of the copyright holder nor the names of its
     contributors may be used to endorse or promote products derived
     from this software without specific prior written permission.

  THIS SOFTWARE IS PROVIDED BY THE COPYRIGHT HOLDERS AND CONTRIBUTORS "AS
  IS" AND ANY EXPRESS OR IMPLIED WARRANTIES, INCLUDING, BUT NOT LIMITED TO,
  THE IMPLIED WARRANTIES OF MERCHANTABILITY AND FITNESS FOR A PARTICULAR
  PURPOSE ARE DISCLAIMED. IN NO EVENT SHALL THE COPYRIGHT HOLDER OR
  CONTRIBUTORS BE LIABLE FOR ANY DIRECT, INDIRECT, INCIDENTAL, SPECIAL,
  EXEMPLARY, OR CONSEQUENTIAL DAMAGES (INCLUDING, BUT NOT LIMITED TO,
  PROCUREMENT OF SUBSTITUTE GOODS OR SERVICES; LOSS OF USE, DATA, OR PROFITS;
  OR BUSINESS INTERRUPTION) HOWEVER CAUSED AND ON ANY THEORY OF LIABILITY,
  WHETHER IN CONTRACT, STRICT LIABILITY, OR TORT (INCLUDING NEGLIGENCE OR
  OTHERWISE) ARISING IN ANY WAY OUT OF THE USE OF THIS SOFTWARE, EVEN IF
  ADVISED OF THE POSSIBILITY OF SUCH DAMAGE.
*/

#ifndef _SOFTPWM_H_
#define _SOFTPWM_H_

#include <Arduino.h>

// helper macros
#define SOFTPWM_DEFINE_PINMODE(CHANNEL, PMODE, PORT, BIT) \
  namespace Palatis { \
    template <> \
    inline void pinModeStatic<CHANNEL>(uint8_t const mode) { \
      if (mode == INPUT) { \
        bitClear(PMODE, BIT); bitClear(PORT, BIT); \
      } \
      else if (mode == INPUT_PULLUP) { \
        bitClear(PMODE, BIT); bitSet(PORT, BIT); \
      } \
      else { \
        bitSet(PMODE, BIT); \
      } \
    } \
  }

#define SOFTPWM_DEFINE_CHANNEL(CHANNEL, PMODE, PORT, BIT) \
  namespace Palatis { \
    template <> \
    inline void bitWriteStatic<CHANNEL>(bool const value) { \
      bitWrite( PORT, BIT, value ); \
    } \
  } \
  SOFTPWM_DEFINE_PINMODE(CHANNEL, PMODE, PORT, BIT)

#define SOFTPWM_DEFINE_CHANNEL_INVERT(CHANNEL, PMODE, PORT, BIT) \
  namespace Palatis { \
    template <> \
    inline void bitWriteStatic<CHANNEL>(bool const value) { \
      bitWrite(PORT, BIT, !value); \
    } \
  } \
  SOFTPWM_DEFINE_PINMODE(CHANNEL, PMODE, PORT, BIT)

#if defined(__AVR_ATtiny24__) || defined(__AVR_ATtiny44__) || defined(__AVR_ATtiny84__)
#define SOFTPWM_DEFINE_OBJECT_WITH_PWM_LEVELS(CHANNEL_CNT, PWM_LEVELS) \
  namespace Palatis { \
    CSoftPWM<CHANNEL_CNT, PWM_LEVELS> SoftPWM; \
  } \
  ISR(TIM1_COMPA_vect) { \
    interrupts(); \
    Palatis::SoftPWM.update(); \
  }
#else
	#if defined (__AVR_ATmega8__) || defined (__AVR_ATmega8A__)
		#define TIMSK1	TIMSK
	#endif
#define SOFTPWM_DEFINE_OBJECT_WITH_PWM_LEVELS(CHANNEL_CNT, PWM_LEVELS) \
  namespace Palatis { \
    CSoftPWM<CHANNEL_CNT, PWM_LEVELS> SoftPWM; \
  } \
  ISR(TIMER1_COMPA_vect) { \
    interrupts(); \
	Palatis::SoftPWM.update(); \
  }
#endif

#define SOFTPWM_DEFINE_OBJECT(CHANNEL_CNT) \
  SOFTPWM_DEFINE_OBJECT_WITH_PWM_LEVELS(CHANNEL_CNT, 0)

#define SOFTPWM_DEFINE_EXTERN_OBJECT_WITH_PWM_LEVELS(CHANNEL_CNT, PWM_LEVELS) \
  namespace Palatis { \
    extern CSoftPWM<CHANNEL_CNT, PWM_LEVELS> SoftPWM; \
  }

#define SOFTPWM_DEFINE_EXTERN_OBJECT(CHANNEL_CNT) \
  SOFTPWM_DEFINE_EXTERN_OBJECT_WITH_PWM_LEVELS(CHANNEL_CNT, 0)

// here comes the magic @o@
namespace Palatis {

template <int channel> inline void bitWriteStatic(bool value) {}
template <int channel> inline void pinModeStatic(uint8_t mode) {}

template <int channel>
struct bitWriteStaticExpander {
  void operator() (bool value) const {
    bitWriteStatic<channel>(value);
    bitWriteStaticExpander < channel - 1 > ()(value);
  }

  void operator() (uint8_t const &count, uint8_t const * const &channels) const {
#ifdef SOFTPWM_OUTPUT_DELAY
    bitWriteStatic<channel>((count + channel) < channels[channel]);
#else
    bitWriteStatic<channel>(count < channels[channel]);
#endif
    bitWriteStaticExpander < channel - 1 > ()(count, channels);
  }
};

template <>
struct bitWriteStaticExpander < -1 > {
  void operator() (bool) const {}
  void operator() (uint8_t const &, uint8_t const * const &) const {}
};

template <int channel>
struct pinModeStaticExpander {
  void operator() (uint8_t const mode) const
  {
    pinModeStatic<channel>(mode);
    pinModeStaticExpander < channel - 1 > ()(mode);
  }
};

template <>
struct pinModeStaticExpander < -1 > {
  void operator() (uint8_t const mode) const {}
};

template <unsigned int num_channels, unsigned int num_PWM_levels>
class CSoftPWM {
  public:
    void begin(const unsigned long hertz) {
      allOff();  //this prevents inverted channels from momentarily going LOW
      asm volatile ("/************ pinModeStaticExpander begin ************/");
      const uint8_t oldSREG = SREG;
      noInterrupts();
      pinModeStaticExpander < num_channels - 1 > ()( OUTPUT );
      SREG = oldSREG;
      asm volatile ("/************ pinModeStaticExpander end ************/");

      /* the setup of timer1 is stolen from ShiftPWM :-P
         http://www.elcojacobs.com/shiftpwm/ */
      asm volatile ("/************ timer setup begin ************/");
      TCCR1A = 0b00000000;
      TCCR1B = 0b00001001;
      OCR1A = (F_CPU - hertz * PWMlevels() / 2) / (hertz * PWMlevels());
      bitSet(TIMSK1, OCIE1A);
      asm volatile ("/************ timer setup end ************/");

      _count = 0;
    }

    void set(const int channel_idx, const uint8_t value) {
      _channels[channel_idx] = value;
    }

    size_t size() const {
      return num_channels;
    }

    unsigned int PWMlevels() const {
      return num_PWM_levels ? num_PWM_levels : 256;
    }

    void allOff() {
      asm volatile ("/********** CSoftPWM::allOff() begin **********/");
      const uint8_t oldSREG = SREG;
      noInterrupts();
      for (int i = 0; i < num_channels; ++i)
        _channels[i] = 0;
      bitWriteStaticExpander < num_channels - 1 > ()(false);
      SREG = oldSREG;
      asm volatile ("/********** CSoftPWM::allOff() end **********/");
    }

    /* This function cannot be private because the ISR uses it, and I have
       no idea about how to make friends with ISR. :-( */
    void update() __attribute__((always_inline)) {
      asm volatile ("/********** CSoftPWM::update() begin **********/");
      const uint8_t count = _count;
      bitWriteStaticExpander < num_channels - 1 > ()(count, _channels);
      ++_count;
      if (_count == PWMlevels())
        _count = 0;
      asm volatile ("/********** CSoftPWM::update() end **********/");
    }
		
    #if defined(HAVE_HWSERIAL0)
      /* this function is stolen from ShiftPWM :-P
         http://www.elcojacobs.com/shiftpwm/ */
      void printInterruptLoad() {
        unsigned long time1, time2;

        bitSet(TIMSK1, OCIE1A); // enable interrupt
        time1 = micros();
        delayMicroseconds(5000);
        time1 = micros() - time1;

        bitClear(TIMSK1, OCIE1A); // disable interrupt
        time2 = micros();
        delayMicroseconds(5000);
        time2 = micros() - time2;

        const double load = static_cast<double>(time1 - time2) / time1;
        const double interrupt_frequency = static_cast<double>(F_CPU) / (OCR1A + 1);
        const double cycles_per_interrupt = load * F_CPU / interrupt_frequency;

        Serial.println(F("SoftPWM::printInterruptLoad():"));
        Serial.print(F("  Load of interrupt: "));
        Serial.println(load, 10);
        Serial.print(F("  Clock cycles per interrupt: "));
        Serial.println( cycles_per_interrupt );
        Serial.print(F("  Interrupt frequency: "));
        Serial.print(interrupt_frequency);
        Serial.println(F(" Hz"));
        Serial.print(F("  PWM frequency: "));
        Serial.print(interrupt_frequency / PWMlevels());
        Serial.println(F(" Hz"));
        Serial.print(F("  PWM levels: "));
        Serial.println(PWMlevels());

        bitSet(TIMSK1, OCIE1A);  // enable interrupt again
      }
    #endif

  private:
    uint8_t _channels[num_channels];
    uint8_t _count;
};

} // namespace Palatis
#endif // #ifndef _SOFTPWM_H_
```
