## Supplementary Material 4 for "De novo Powered Air-Purifying Respirator Design and Fabrication for Pandemic Response": MasterCode_Custom Design.ino

```
 #include <Wire.h>

#include <SoftPWM.h>

#include <sdpsensor.h>

SDP8XXSensor sdp;

//Stores the minimum interval before activating the timer, in milliseconds
unsigned long MinAlarm = 10 * 1000; // * 60 seconds/min

//stores the amount of time since the last alarm-qualifying event
unsigned long startTime = 0;

//stores the current time regardless.
unsigned long currTime = 0;

//Stores the frequency of the buzzer tone in Hertz.
unsigned int BuzzTone = 500;


//Everything from here until the start of void setup is for PWM control
// Set up Arduino pin 9 as PWM channel 0:
// (For examples on how to set this see:
//  File > Examples > arduino-softpwm > SoftPWM_example )
SOFTPWM_DEFINE_CHANNEL(0, DDRB, PORTB, PORTB1);  //Arduino pin 9

/* Here you make an instance of desired channel counts you want
   with the default 256 PWM levels (0 ~ 255). */
SOFTPWM_DEFINE_OBJECT(1);

/* If you want to use the SoftPWM object outside where it's defined,
   add the following line to the file. */
SOFTPWM_DEFINE_EXTERN_OBJECT(1);

// set analog input pin
int analogInputPin = A0;
int digitalOutputPin = 2; 

void setup() {
  //the part below is for the diff. pressure sensor, until the while statement
  Wire.begin();
  Serial.begin(9600);
  delay(1000); // let serial console settle
  do {
    int ret = sdp.init();
    if (ret == 0) {
      Serial.print("init(): success\n");
      break;
    } else {
      Serial.print("init(): failed, ret = ");
      Serial.println(ret);
      delay(1000);
    }
  } while(true);

  //the part below, until the beginning of void loop is for the PWM control
  // begin with 100hz pwm frequency
  Palatis::SoftPWM.begin(100);
}

void loop() {
  //the part below, until the 'delay(10)' statement is for the PWM control
  long inputVal = analogRead(analogInputPin);

  // CONVERSION:
  // - analogRead() returns a value from 0..1023
  // - SoftPWM takes values from 0..255
  // - blower reaches max speed at 90%
  // - probably want to have a minimum blower speed too: minPWM
  // So, convert inputVal range from 0..1023 to minPWM..230
  const long minPWM = 64; // set min to be 25% of 255
  const long maxPWM = 230; // set max to be 90% of 255
  long PWMLevel = minPWM + (inputVal * (maxPWM-minPWM)) / (1023);

  currTime = millis();

  // set pin 9 (pwm channel 0) to new PWM level
  Palatis::SoftPWM.set(0, PWMLevel);
  Serial.println(PWMLevel); // debugging

  delay(10); // no need to respond to user input faster than ~100Hz

  //the part below, until the end is for the diff. pressure sensor
  int ret = sdp.readSample();
  if (ret == 0) {
    Serial.print("Differential pressure: ");
    Serial.print(sdp.getDifferentialPressure());
    Serial.print("Pa | ");

    Serial.print("Temp: ");
    Serial.print(sdp.getTemperature());
    Serial.print("C\n");
    if (sdp.getDifferentialPressure()<205) // this is for the low flow alarm
    {

      Serial.print("Low Alarm Activated: ");
      Serial.print((currTime - startTime)/1000);
      Serial.println(" sec.");
      if((currTime-startTime) >= MinAlarm)
      {
    tone(digitalOutputPin,BuzzTone);
      }
    }
    else
    {
      startTime = currTime;
    noTone(digitalOutputPin);
    }  
  }
  else {
    Serial.print("Error in readSample(), ret = ");
    Serial.println(ret);
  }
  //delay(10);
}
```
