## Supplementary Material 4 for "De novo Powered Air-Purifying Respirator Design and Fabrication for Pandemic Response": README.txt

Creative Commons Attribution-ShareAlike 4.0 International Public License

By exercising the Licensed Rights (defined below), You accept and agree to be bound by the terms and conditions of this Creative Commons Attribution-ShareAlike 4.0 International Public License ("Public License"). To the extent this Public License may be interpreted as a contract, You are granted the Licensed Rights in consideration of Your acceptance of these terms and conditions, and the Licensor grants You such rights in consideration of benefits the Licensor receives from making the Licensed Material available under these terms and conditions.
Section 1 – Definitions.
a.	Adapted Material means material subject to Copyright and Similar Rights that is derived from or based upon the Licensed Material and in which the Licensed Material is translated, altered, arranged, transformed, or otherwise modified in a manner requiring permission under the Copyright and Similar Rights held by the Licensor. For purposes of this Public License, where the Licensed Material is a musical work, performance, or sound recording, Adapted Material is always produced where the Licensed Material is synched in timed relation with a moving image.
b.	Adapter's License means the license You apply to Your Copyright and Similar Rights in Your contributions to Adapted Material in accordance with the terms and conditions of this Public License.
c.	BY-SA Compatible License means a license listed at creativecommons.org/compatiblelicenses, approved by Creative Commons as essentially the equivalent of this Public License.
d.	Copyright and Similar Rights means copyright and/or similar rights closely related to copyright including, without limitation, performance, broadcast, sound recording, and Sui Generis Database Rights, without regard to how the rights are labeled or categorized. For purposes of this Public License, the rights specified in Section 2(b)(1)-(2) are not Copyright and Similar Rights.
e.	Effective Technological Measures means those measures that, in the absence of proper authority, may not be circumvented under laws fulfilling obligations under Article 11 of the WIPO Copyright Treaty adopted on December 20, 1996, and/or similar international agreements.
f.	Exceptions and Limitations means fair use, fair dealing, and/or any other exception or limitation to Copyright and Similar Rights that applies to Your use of the Licensed Material.
g.	License Elements means the license attributes listed in the name of a Creative Commons Public License. The License Elements of this Public License are Attribution and ShareAlike.
h.	Licensed Material means the artistic or literary work, database, or other material to which the Licensor applied this Public License.
i.	Licensed Rights means the rights granted to You subject to the terms and conditions of this Public License, which are limited to all Copyright and Similar Rights that apply to Your use of the Licensed Material and that the Licensor has authority to license.
j.	Licensor means the individual(s) or entity(ies) granting rights under this Public License.
k.	Share means to provide material to the public by any means or process that requires permission under the Licensed Rights, such as reproduction, public display, public performance, distribution, dissemination, communication, or importation, and to make material available to the public including in ways that members of the public may access the material from a place and at a time individually chosen by them.
l.	Sui Generis Database Rights means rights other than copyright resulting from Directive 96/9/EC of the European Parliament and of the Council of 11 March 1996 on the legal protection of databases, as amended and/or succeeded, as well as other essentially equivalent rights anywhere in the world.
m.	You means the individual or entity exercising the Licensed Rights under this Public License. Your has a corresponding meaning.
Section 2 – Scope.
a.	License grant. 
1.	Subject to the terms and conditions of this Public License, the Licensor hereby grants You a worldwide, royalty-free, non-sublicensable, non-exclusive, irrevocable license to exercise the Licensed Rights in the Licensed Material to: 
A.	reproduce and Share the Licensed Material, in whole or in part; and
B.	produce, reproduce, and Share Adapted Material.
2.	Exceptions and Limitations. For the avoidance of doubt, where Exceptions and Limitations apply to Your use, this Public License does not apply, and You do not need to comply with its terms and conditions.
3.	Term. The term of this Public License is specified in Section 6(a).
4.	Media and formats; technical modifications allowed. The Licensor authorizes You to exercise the Licensed Rights in all media and formats whether now known or hereafter created, and to make technical modifications necessary to do so. The Licensor waives and/or agrees not to assert any right or authority to forbid You from making technical modifications necessary to exercise the Licensed Rights, including technical modifications necessary to circumvent Effective Technological Measures. For purposes of this Public License, simply making modifications authorized by this Section 2(a)(4) never produces Adapted Material.
5.	Downstream recipients. 
A.	Offer from the Licensor – Licensed Material. Every recipient of the Licensed Material automatically receives an offer from the Licensor to exercise the Licensed Rights under the terms and conditions of this Public License.
B.	Additional offer from the Licensor – Adapted Material. Every recipient of Adapted Material from You automatically receives an offer from the Licensor to exercise the Licensed Rights in the Adapted Material under the conditions of the Adapter’s License You apply.
C.	No downstream restrictions. You may not offer or impose any additional or different terms or conditions on, or apply any Effective Technological Measures to, the Licensed Material if doing so restricts exercise of the Licensed Rights by any recipient of the Licensed Material.
6.	No endorsement. Nothing in this Public License constitutes or may be construed as permission to assert or imply that You are, or that Your use of the Licensed Material is, connected with, or sponsored, endorsed, or granted official status by, the Licensor or others designated to receive attribution as provided in Section 3(a)(1)(A)(i).
b.	Other rights.
1.	Moral rights, such as the right of integrity, are not licensed under this Public License, nor are publicity, privacy, and/or other similar personality rights; however, to the extent possible, the Licensor waives and/or agrees not to assert any such rights held by the Licensor to the limited extent necessary to allow You to exercise the Licensed Rights, but not otherwise.
2.	Patent and trademark rights are not licensed under this Public License.
3.	To the extent possible, the Licensor waives any right to collect royalties from You for the exercise of the Licensed Rights, whether directly or through a collecting society under any voluntary or waivable statutory or compulsory licensing scheme. In all other cases the Licensor expressly reserves any right to collect such royalties.
Section 3 – License Conditions.
Your exercise of the Licensed Rights is expressly made subject to the following conditions.
a.	Attribution.
1.	If You Share the Licensed Material (including in modified form), You must:
A.	retain the following if it is supplied by the Licensor with the Licensed Material: 
i.	identification of the creator(s) of the Licensed Material and any others designated to receive attribution, in any reasonable manner requested by the Licensor (including by pseudonym if designated);
ii.	a copyright notice;
iii.	a notice that refers to this Public License; 
iv.	a notice that refers to the disclaimer of warranties;
v.	a URI or hyperlink to the Licensed Material to the extent reasonably practicable;
B.	indicate if You modified the Licensed Material and retain an indication of any previous modifications; and
C.	indicate the Licensed Material is licensed under this Public License, and include the text of, or the URI or hyperlink to, this Public License.
2.	You may satisfy the conditions in Section 3(a)(1) in any reasonable manner based on the medium, means, and context in which You Share the Licensed Material. For example, it may be reasonable to satisfy the conditions by providing a URI or hyperlink to a resource that includes the required information.
3.	If requested by the Licensor, You must remove any of the information required by Section 3(a)(1)(A) to the extent reasonably practicable.
b.	ShareAlike. 
In addition to the conditions in Section 3(a), if You Share Adapted Material You produce, the following conditions also apply.
1.	The Adapter’s License You apply must be a Creative Commons license with the same License Elements, this version or later, or a BY-SA Compatible License.
2.	You must include the text of, or the URI or hyperlink to, the Adapter's License You apply. You may satisfy this condition in any reasonable manner based on the medium, means, and context in which You Share Adapted Material.
3.	You may not offer or impose any additional or different terms or conditions on, or apply any Effective Technological Measures to, Adapted Material that restrict exercise of the rights granted under the Adapter's License You apply.
Section 4 – Sui Generis Database Rights.
Where the Licensed Rights include Sui Generis Database Rights that apply to Your use of the Licensed Material:
a.	for the avoidance of doubt, Section 2(a)(1) grants You the right to extract, reuse, reproduce, and Share all or a substantial portion of the contents of the database;
b.	if You include all or a substantial portion of the database contents in a database in which You have Sui Generis Database Rights, then the database in which You have Sui Generis Database Rights (but not its individual contents) is Adapted Material, including for purposes of Section 3(b); and
c.	You must comply with the conditions in Section 3(a) if You Share all or a substantial portion of the contents of the database.
For the avoidance of doubt, this Section 4 supplements and does not replace Your obligations under this Public License where the Licensed Rights include other Copyright and Similar Rights. 
Section 5 – Disclaimer of Warranties and Limitation of Liability.
a.	Unless otherwise separately undertaken by the Licensor, to the extent possible, the Licensor offers the Licensed Material as-is and as-available, and makes no representations or warranties of any kind concerning the Licensed Material, whether express, implied, statutory, or other. This includes, without limitation, warranties of title, merchantability, fitness for a particular purpose, non-infringement, absence of latent or other defects, accuracy, or the presence or absence of errors, whether or not known or discoverable. Where disclaimers of warranties are not allowed in full or in part, this disclaimer may not apply to You.
b.	To the extent possible, in no event will the Licensor be liable to You on any legal theory (including, without limitation, negligence) or otherwise for any direct, special, indirect, incidental, consequential, punitive, exemplary, or other losses, costs, expenses, or damages arising out of this Public License or use of the Licensed Material, even if the Licensor has been advised of the possibility of such losses, costs, expenses, or damages. Where a limitation of liability is not allowed in full or in part, this limitation may not apply to You.
c.	The disclaimer of warranties and limitation of liability provided above shall be interpreted in a manner that, to the extent possible, most closely approximates an absolute disclaimer and waiver of all liability.
Section 6 – Term and Termination.
a.	This Public License applies for the term of the Copyright and Similar Rights licensed here. However, if You fail to comply with this Public License, then Your rights under this Public License terminate automatically.
b.	Where Your right to use the Licensed Material has terminated under Section 6(a), it reinstates:
1.	automatically as of the date the violation is cured, provided it is cured within 30 days of Your discovery of the violation; or
2.	upon express reinstatement by the Licensor.
For the avoidance of doubt, this Section 6(b) does not affect any right the Licensor may have to seek remedies for Your violations of this Public License.
c.	For the avoidance of doubt, the Licensor may also offer the Licensed Material under separate terms or conditions or stop distributing the Licensed Material at any time; however, doing so will not terminate this Public License.
d.	Sections 1, 5, 6, 7, and 8 survive termination of this Public License.
Section 7 – Other Terms and Conditions.
a.	The Licensor shall not be bound by any additional or different terms or conditions communicated by You unless expressly agreed.
b.	Any arrangements, understandings, or agreements regarding the Licensed Material not stated herein are separate from and independent of the terms and conditions of this Public License.
Section 8 – Interpretation.
a.	For the avoidance of doubt, this Public License does not, and shall not be interpreted to, reduce, limit, restrict, or impose conditions on any use of the Licensed Material that could lawfully be made without permission under this Public License.
b.	To the extent possible, if any provision of this Public License is deemed unenforceable, it shall be automatically reformed to the minimum extent necessary to make it enforceable. If the provision cannot be reformed, it shall be severed from this Public License without affecting the enforceability of the remaining terms and conditions.
c.	No term or condition of this Public License will be waived and no failure to comply consented to unless expressly agreed to by the Licensor.
d.	Nothing in this Public License constitutes or may be interpreted as a limitation upon, or waiver of, any privileges and immunities that apply to the Licensor or You, including from the legal processes of any jurisdiction or authority.


© President and Fellows of Harvard College and The Massachusetts Institute of Technology, 2020.
