## Supplementary Material 4 for "De novo Powered Air-Purifying Respirator Design and Fabrication for Pandemic Response": Assembly Instructions- PanFab Custom Blower Unit.pdf

**PanFab Custom PAPR Blower Unit Assembly Instructions**

Peter Sorger, PhD, 0000-0002-3364-1838

### INTRODUCTION

This document details the process of assembling the blower unit for the PanFab Custom PAPR.

In assembling the PanFab Custom blower unit, electronics were integrated within the 3D printed housing, without any need to modify the housing itself, as was the case in the PanFab Commercial blower unit.

All parts mentioned here are listed in the Bill of Materials in **Supplementary Material 4.**

### ELECTRICAL SETUP

#### Electrical Assembly

To electrically assemble the components, the diagram described by **Figure 1** was ultimately followed**.** A custom printed circuit board (PCB) was developed for ease of assembly, as shown in **Figure 2**. The board will ultimately slot into the top of a standard Arduino Uno R3 (Arduino LLC, Boston, MA) to provide the power and data connections to various peripherals.

© President and Fellows of Harvard College and The Massachusetts Institute of Technology, 2020.

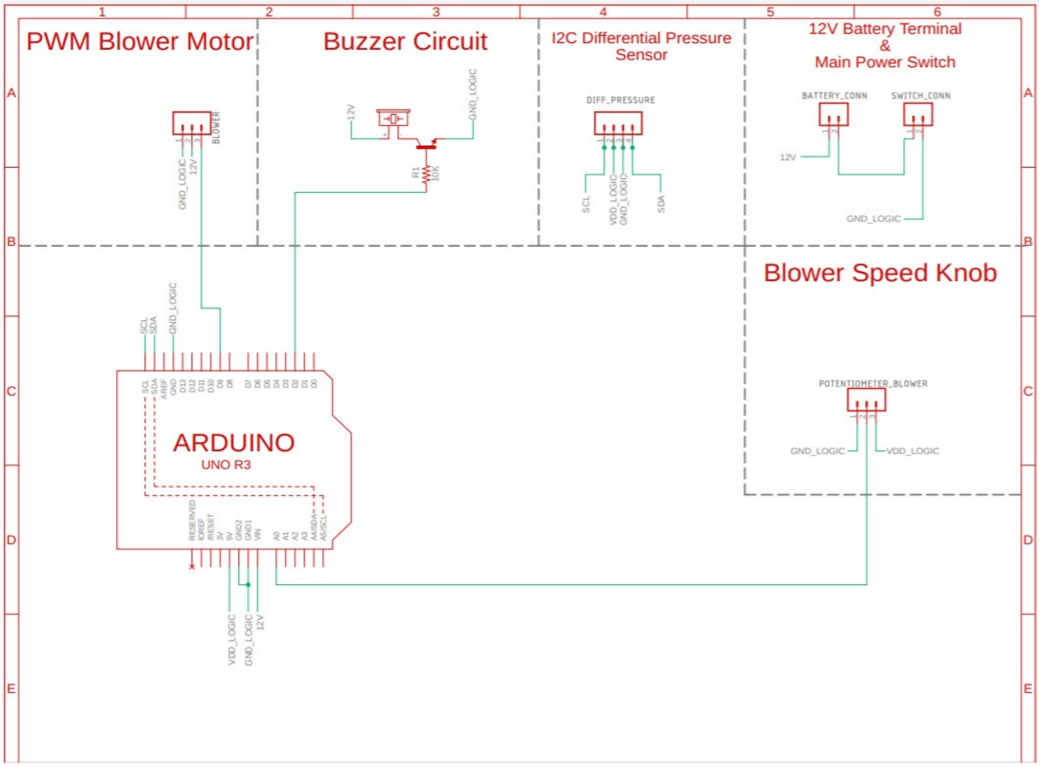


**Figure 1:** An electrical schematic describing the connections in the prototype.

© President and Fellows of Harvard College and The Massachusetts Institute of Technology, 2020.

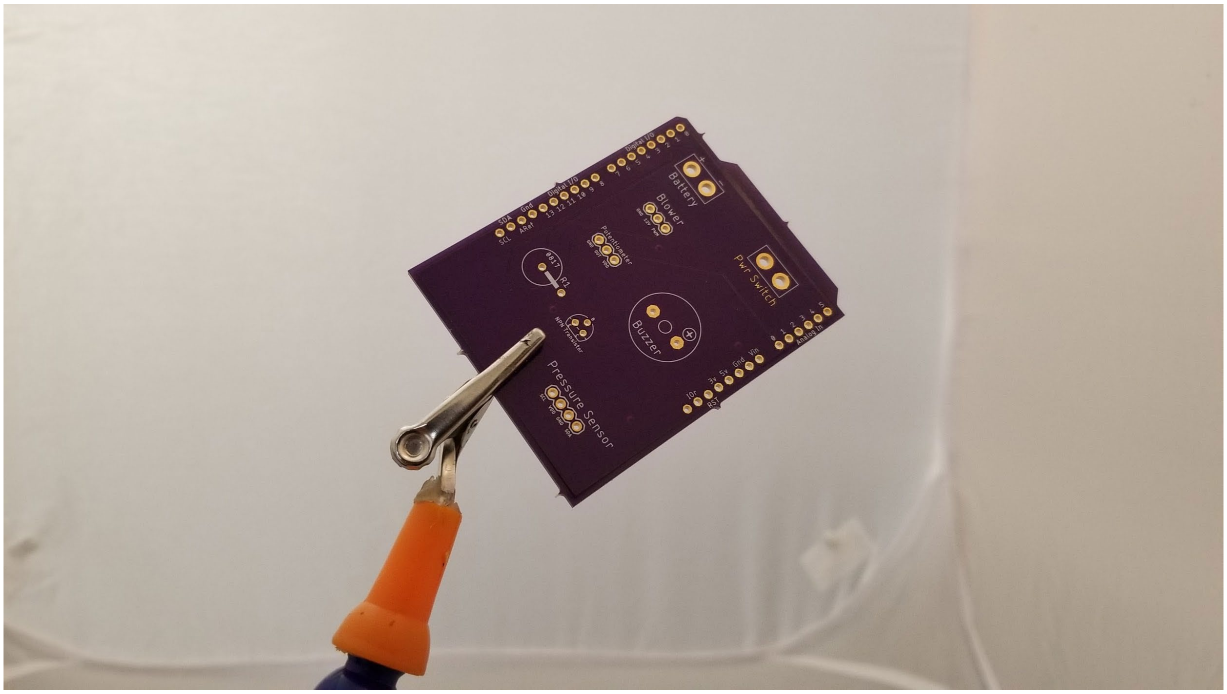


**Figure 2:** A photo of the custom PCB used to connect various electrical components in the prototype.

Male header pins are first inserted into the female headers of the Arduino Uno, as shown in **Figure 3.** From there, the custom PCB is inserted on top of the Arduino Uno to align the male header pins then soldered **(Figure 4).** The white superficial text (to help with component placement) should be facing up. The resulting PCB can then be separated from the Arduino Uno with the male header pins soldered on **(Figure 5)**. A 220-ohm through-hole resistor and a TO-92-3 2N3904BU BJT transistor are then soldered directly to the top of the PCB, as shown in **Figure 6.** The base, collector, and emitter of the transistor match up with the markings on the PCB.

© President and Fellows of Harvard College and The Massachusetts Institute of Technology, 2020.

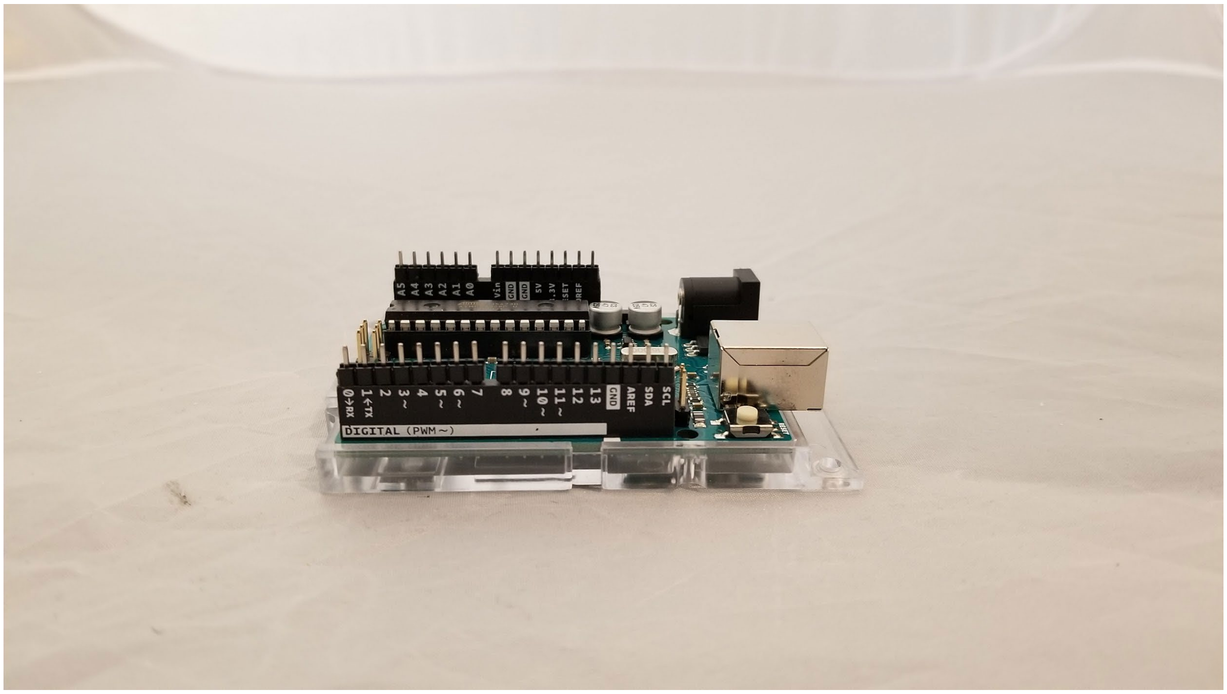


**Figure 3:** Male header pins are inserted into the IO ports of the Arduino Uno.

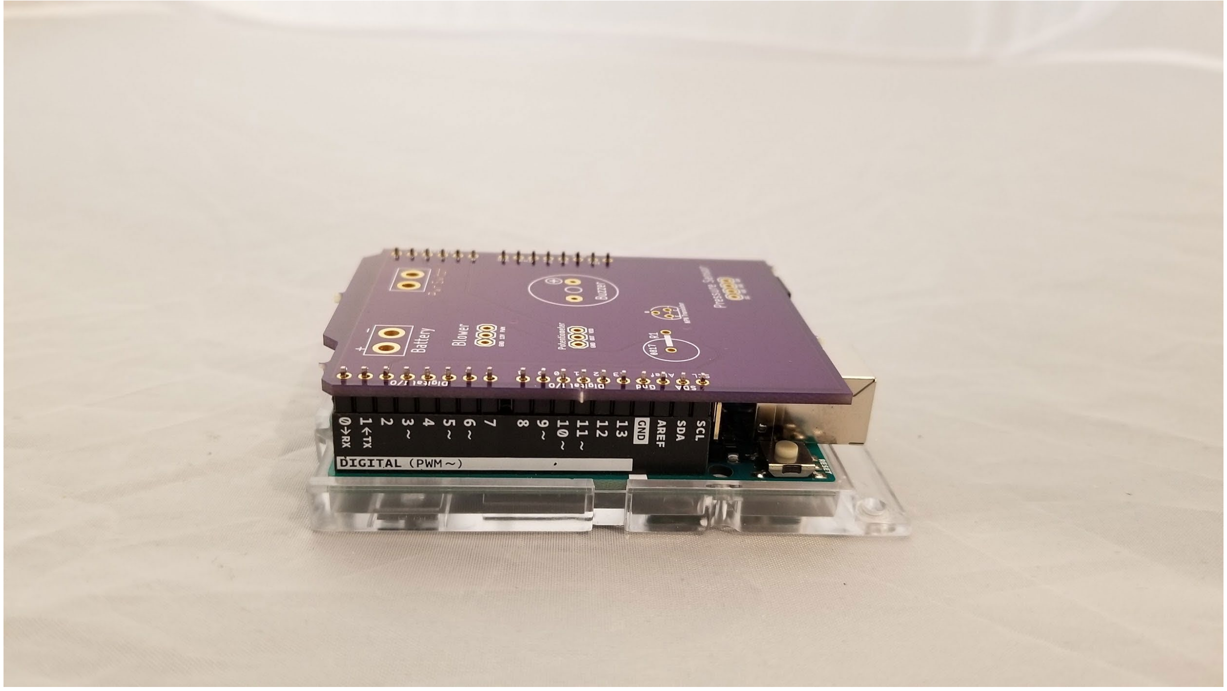


**Figure 4:** The custom PCB is placed on top of the male header pins and the Arduino Uno.

© President and Fellows of Harvard College and The Massachusetts Institute of Technology, 2020.

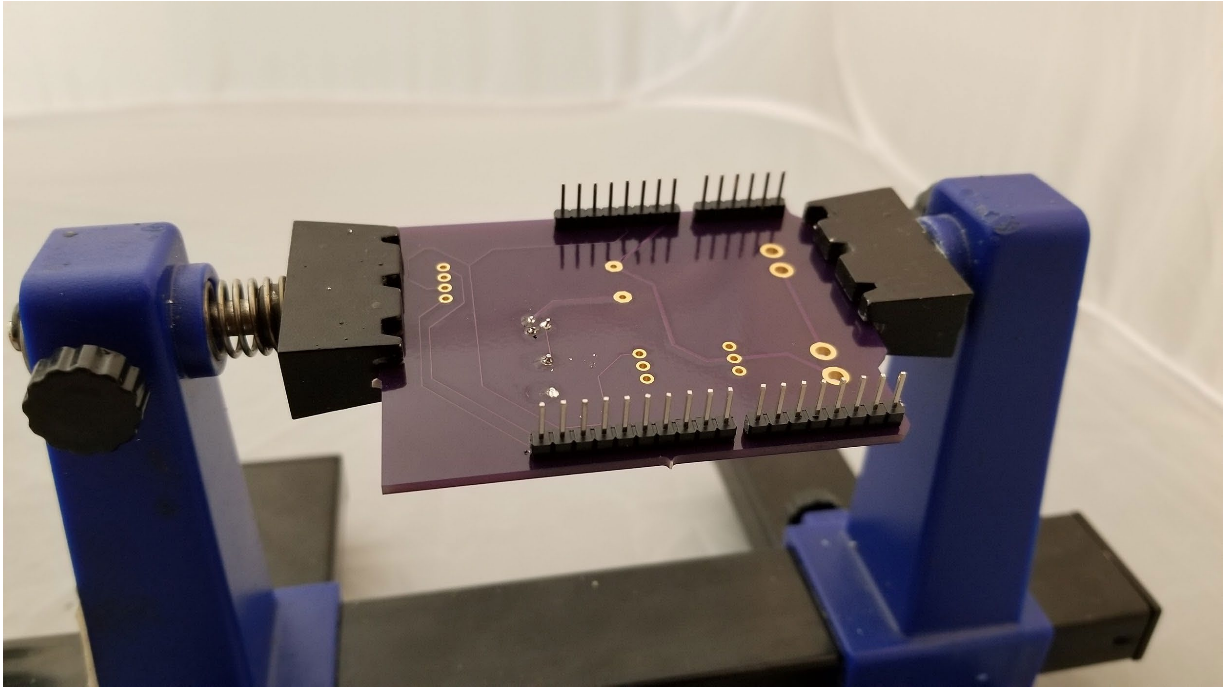

**Figure 5:** The male header pins successfully soldered to the custom PCB.

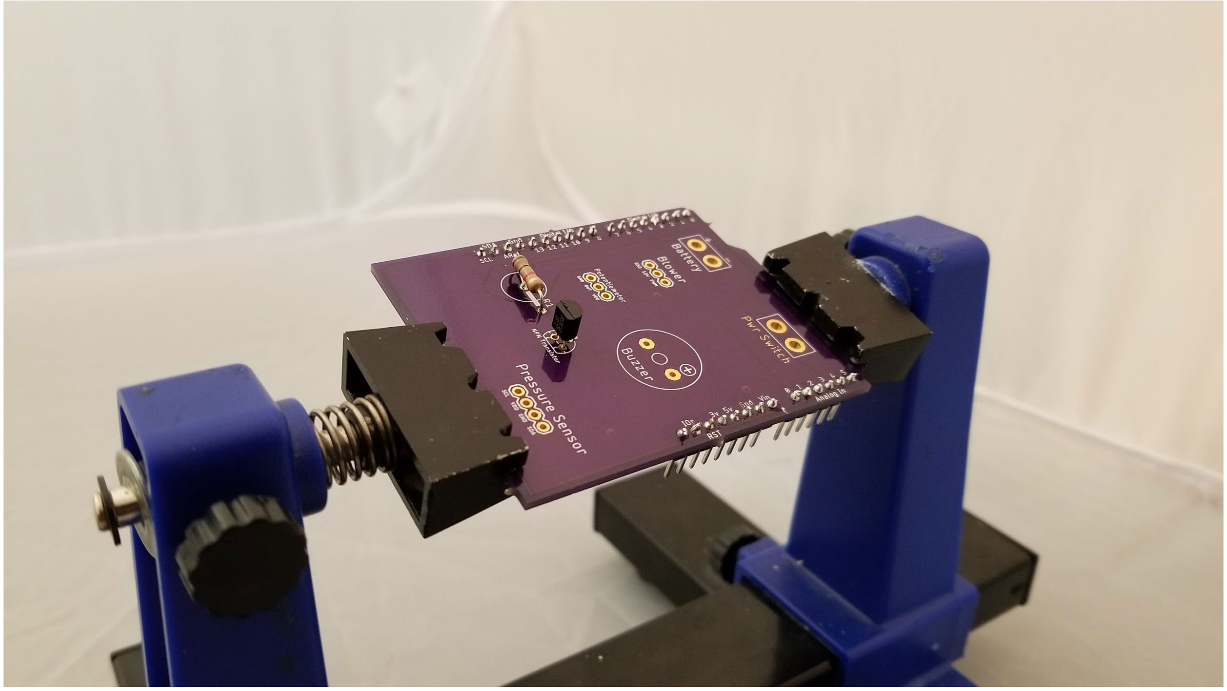


**Figure 6:** The resistor and BJT successfully soldered to the custom PCB.

© President and Fellows of Harvard College and The Massachusetts Institute of Technology, 2020.

Next, the male battery terminal connector with the visible metal pins was obtained and ultimately soldered to the portion of the PCB labelled *Battery*. Care was taken to make sure that the red wire is soldered to the terminal closer to the “+” marking, and the black wire is soldered to the terminal closest to the “-” marking **(Figure 7)**. The accompanying female battery terminal was then soldered to the terminals of the NiMH battery and secured with heat-shrink tubing **(Figure 8).** Additionally, two 12-gauge wires are soldered to the terminals labelled *Pwr Switch***.** To improve wire management, the wires were twisted in a standard power drill to create a twisted pair **(Figure 9)**.

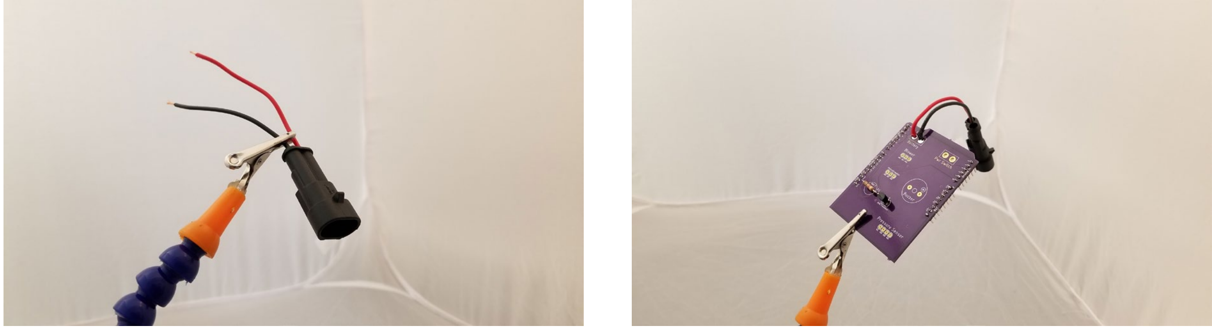

**Figure 7:** The male battery connector before and after being soldered to the PCB.

© President and Fellows of Harvard College and The Massachusetts Institute of Technology, 2020.

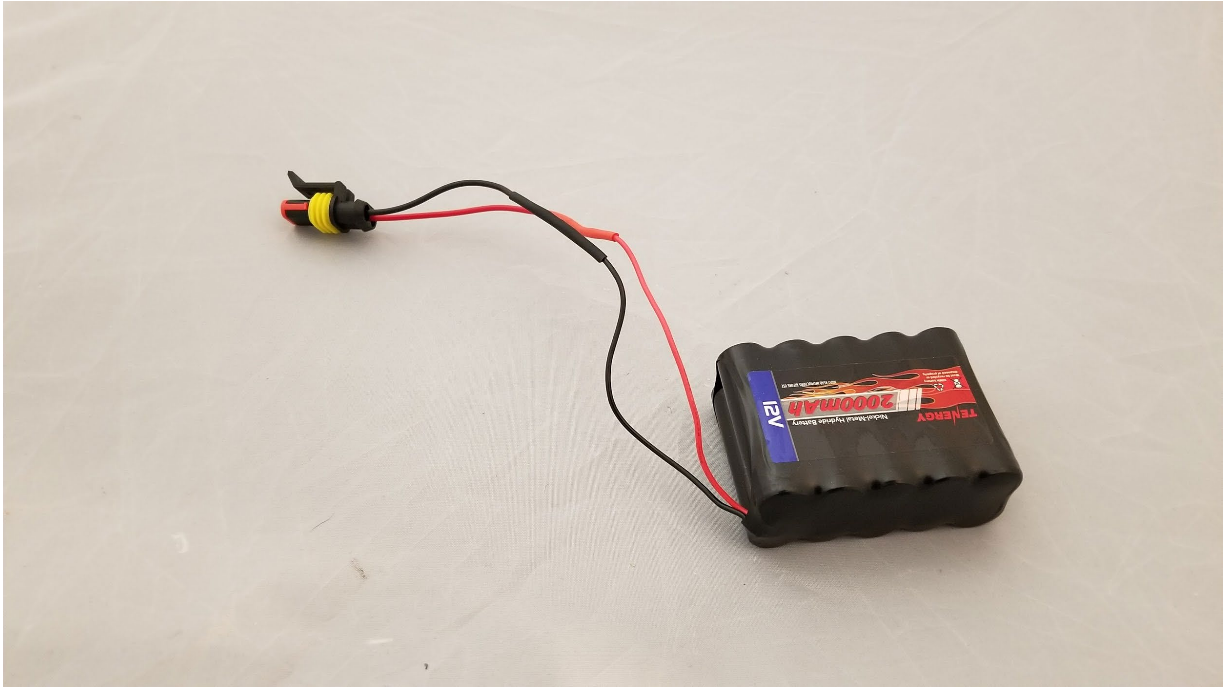


**Figure 8:** The female battery connector after being soldered to the battery.

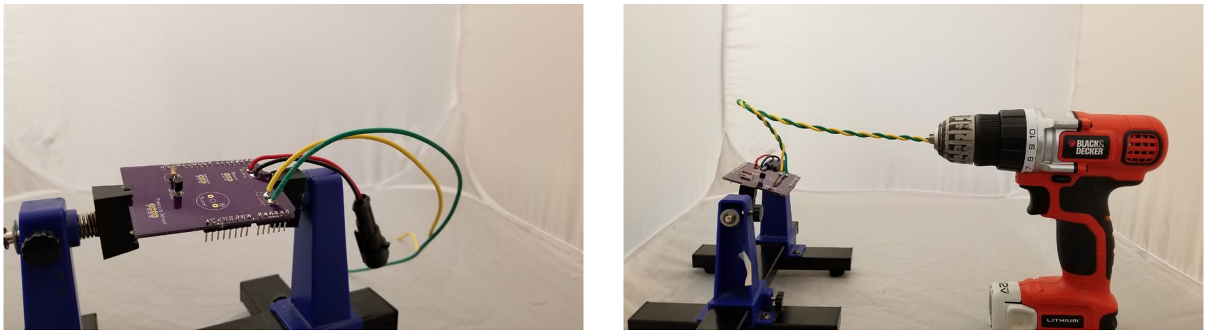


**Figure 9:** The power switch wires after being soldered to the PCB (left) and after being wound into a twisted pair by a power drill (right)

From here, the peripheral setup began. The blower motor wires were twisted in a power drill to simplify cable management. Three 12-inch 22-gauge wires were soldered to the rotary potentiometer **(Figure 10)**. From there, the wires from the blower were soldered to the board directly, with the black wire being in the *GND* position, the red wire being in the *12V* position,

© President and Fellows of Harvard College and The Massachusetts Institute of Technology, 2020.

and the white wire being in the *PWM* position. The wires from the potentiometer are also soldered to the board, with the middle wire soldered to terminal labelled *Out* on the board. The other two wires can be soldered interchangeably. From there, the buzzer was then soldered directly to the board, with the positive terminal being soldered to the red wire, and the negative terminal soldered to the black wire. The final results thus far resembled **Figure 11**.

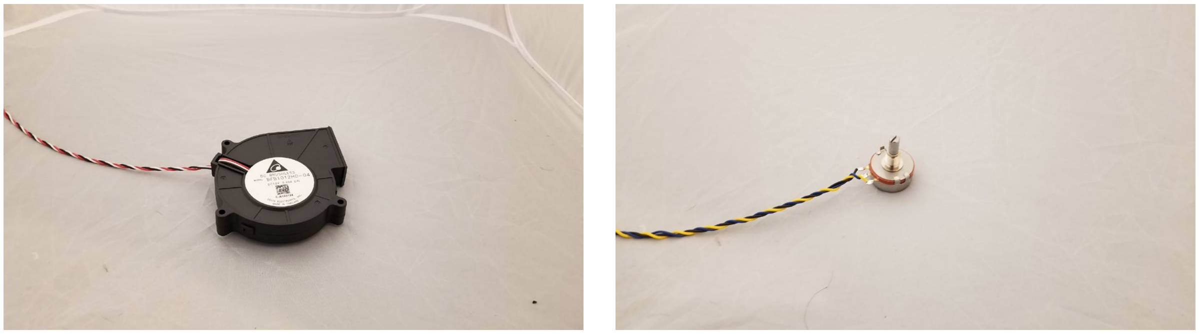

**Figure 10:** The blower motor after the wires were wound into a twisted pair for cable management purposes (left). The rotary potentiometer with wires soldered and twisted (right)

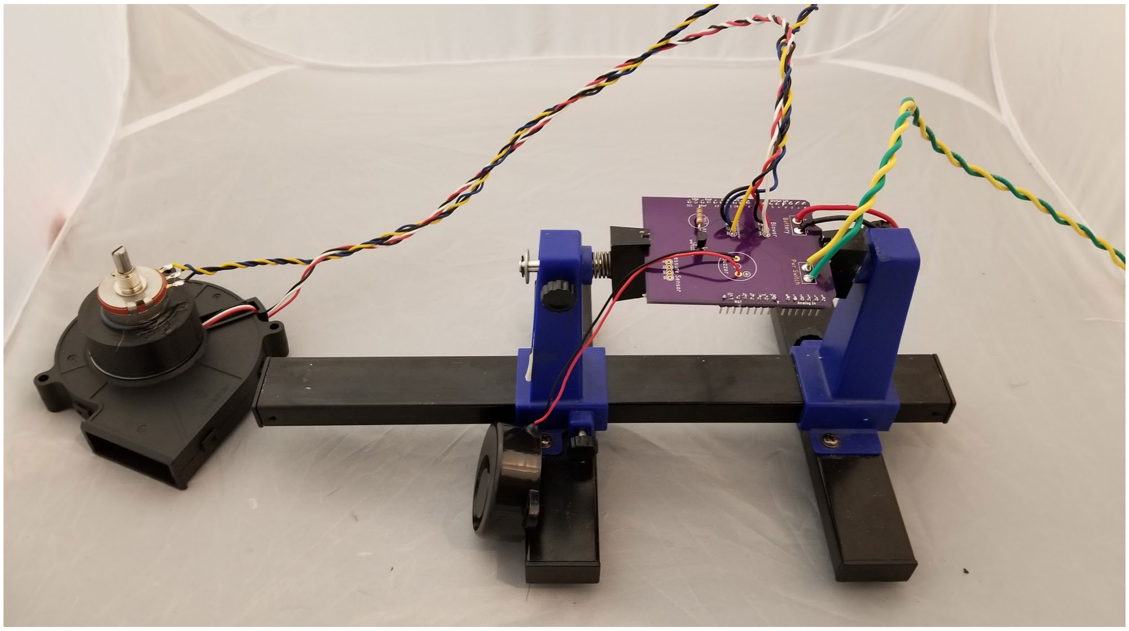

**Figure 11:** The PCB with various peripherals installed.

© President and Fellows of Harvard College and The Massachusetts Institute of Technology, 2020.

The bottom tabs are cut off from the pressure sensor, as shown in **Figure 12**. From there, the four leads are soldered to the board. The inlet and outlet ports should be facing the transistor previously soldered to the board, as shown in **Figure 13.** The PCB is then plugged directly into the Arduino Uno **(Figure 14)**. The two power switch wires are then screwed into the screw terminals on the power switch, as shown in **Figure 15.**

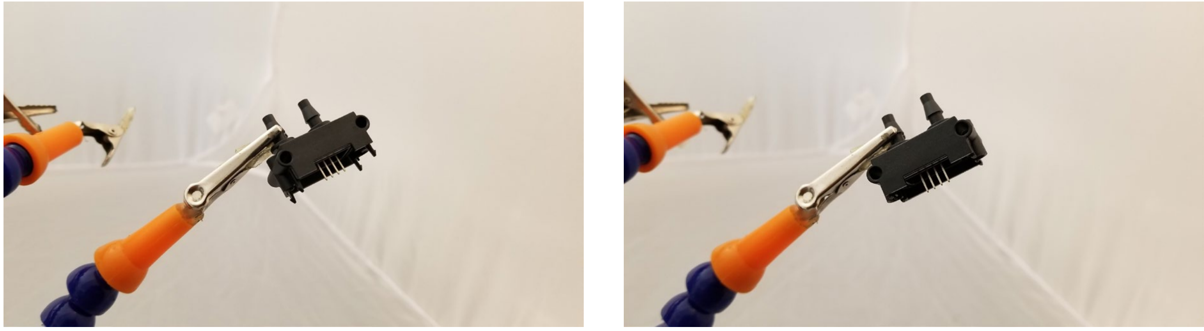

**Figure 12:** The differential pressure sensor before the tabs on the bottom are removed (left) and after (right).

© President and Fellows of Harvard College and The Massachusetts Institute of Technology, 2020.

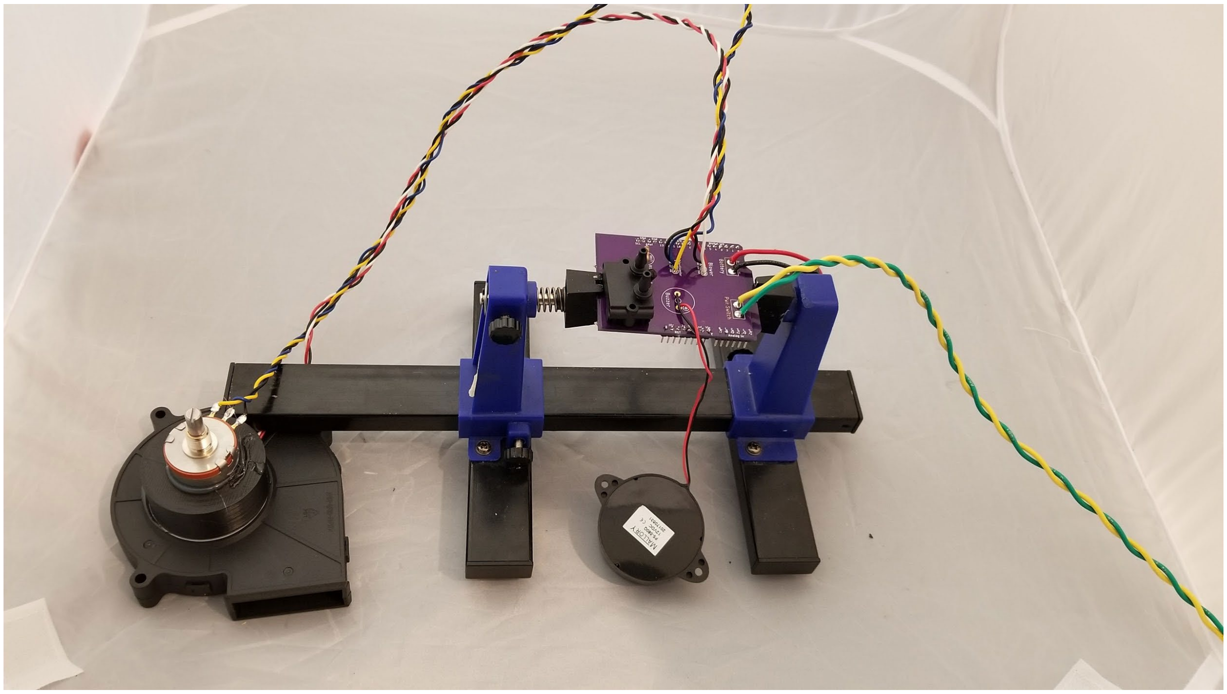

**Figure 13:** The PCB with the pressure sensor installed, before plugging into the Arduino Uno.

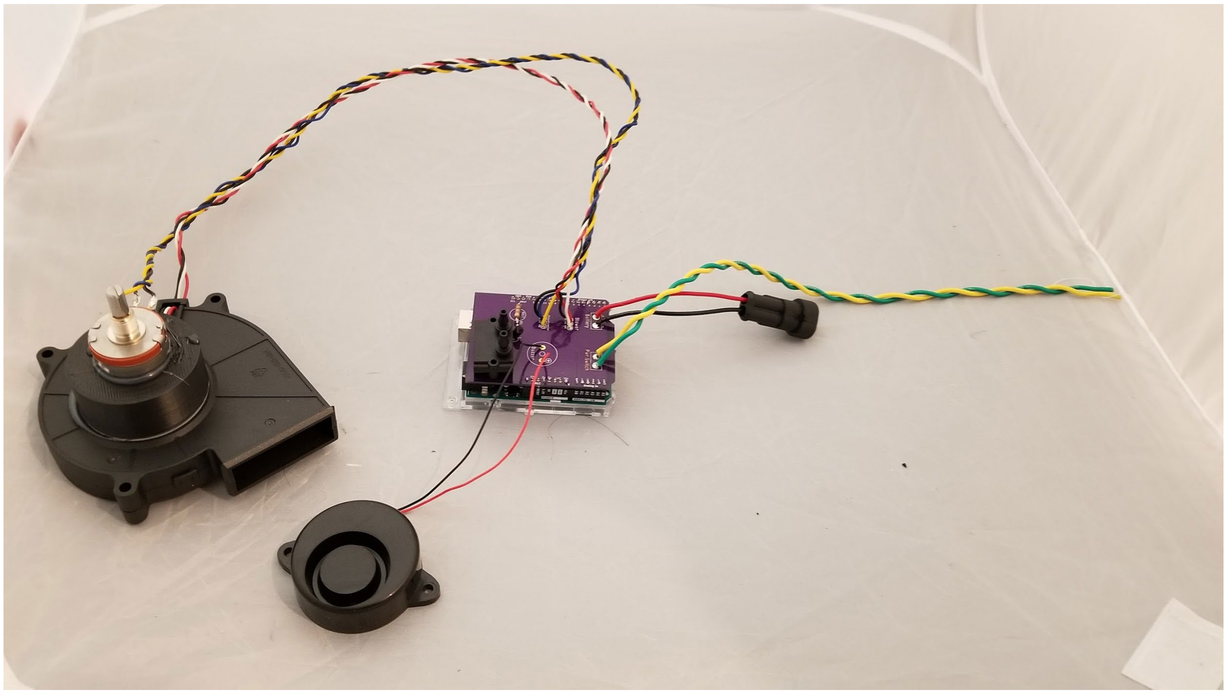

**Figure 14:** The PCB installed to the Arduino Uno. Connection to the battery and power switch not shown.

© President and Fellows of Harvard College and The Massachusetts Institute of Technology, 2020.

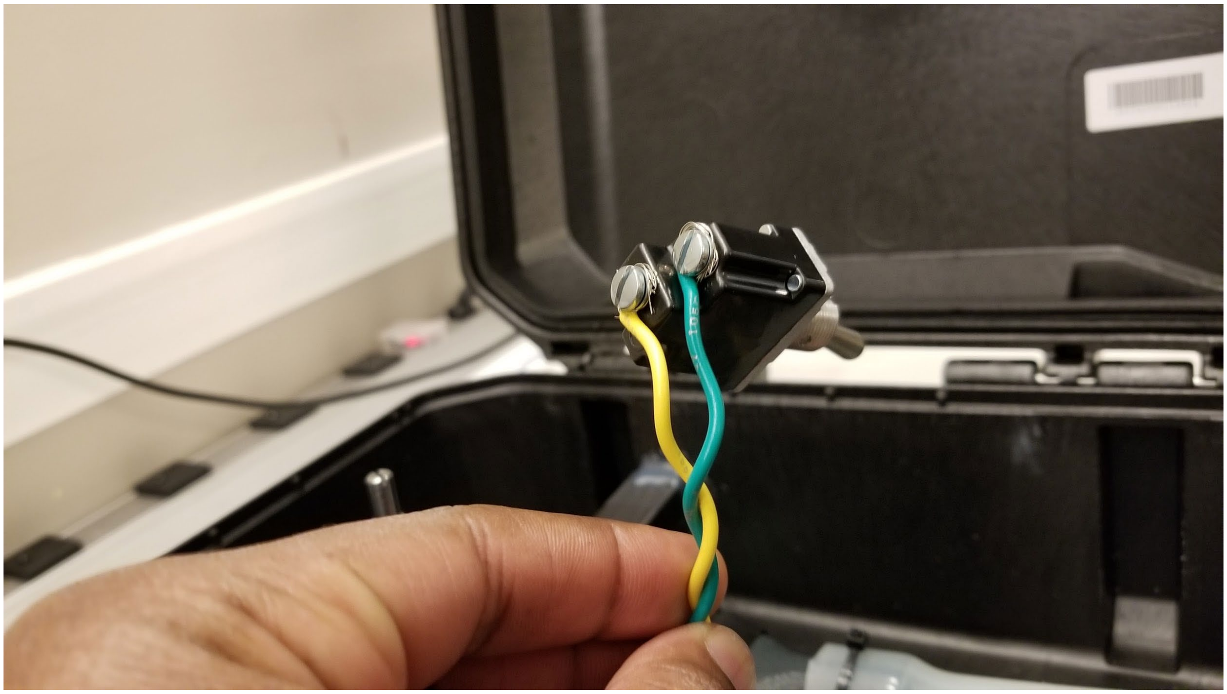

**Figure 15:** The two switch wires installed to the screw terminals.

Additionally, a custom mounting bracket was designed to keep the large blower in-line with the downstream path. The bracket was secured to the case and blower with hot-melt adhesive **Figure 16**. A 1-inch diameter piece of silicone tubing was used to connect the 3D-printed blower adapter, as shown in **Figure 17** in gray, to the black venturi used to measure outlet gas flow rate. Electrical tape was used to secure the 3D-printed blower adapter to the blower motor, and zip-ties to connect the silicone tubing to the blower adapter and venturi. The smaller diameter silicone tubing is connected from the black venturi ports to the differential pressure sensor ports- the pressure sensor port closest to the transistor on the PCB is connected to the port closest to the outlet on the venturi. The switch is placed through the smaller hole on the case, then a switch cover with sealing nut is screwed onto the body and further secured with Loctite Threadlocker

© President and Fellows of Harvard College and The Massachusetts Institute of Technology, 2020.

Red 271 (Loctite, Hartford, CT) **(Figure 18)**. After the electrical connections were made, it was all mechanically fixed to the case with hot-melt glue, as ultimately shown in **Figure 19.**

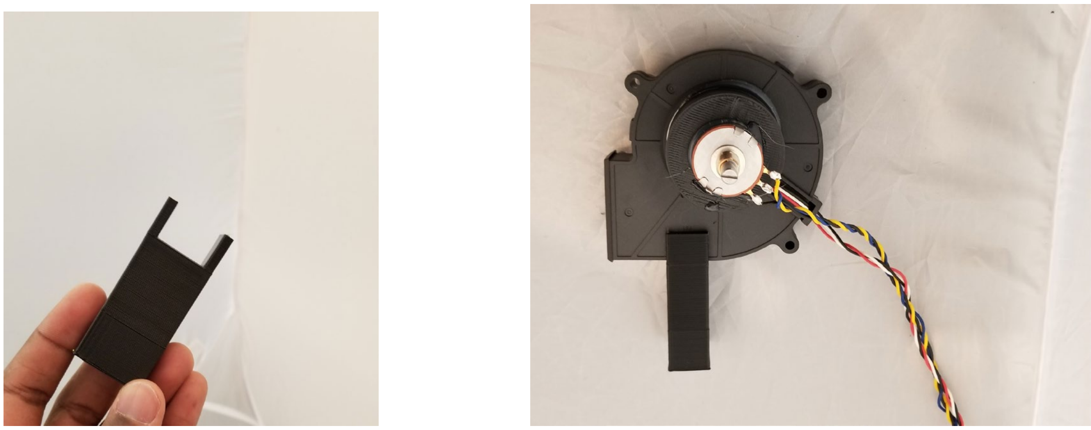

**Figure 16:** The custom blower mounting bracket piece as affixed to the blower motor.

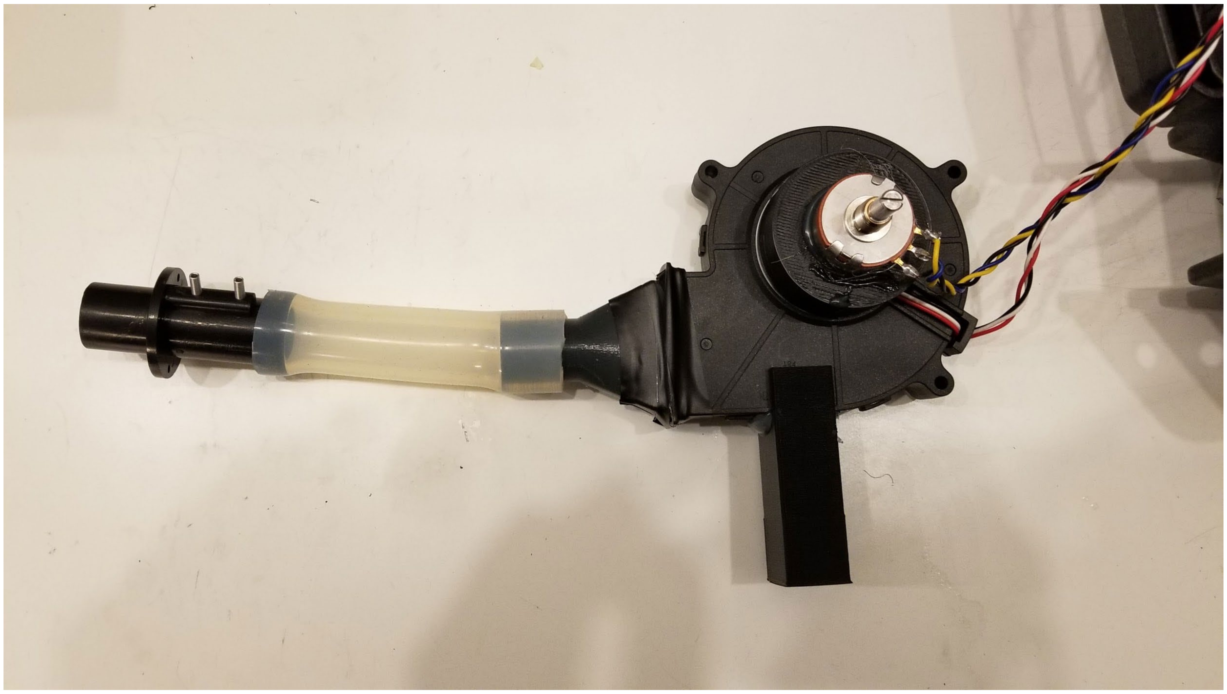

**Figure 17:** The attached silicone tubing with the venturi and 3D-printed blower adapter.

© President and Fellows of Harvard College and The Massachusetts Institute of Technology, 2020.

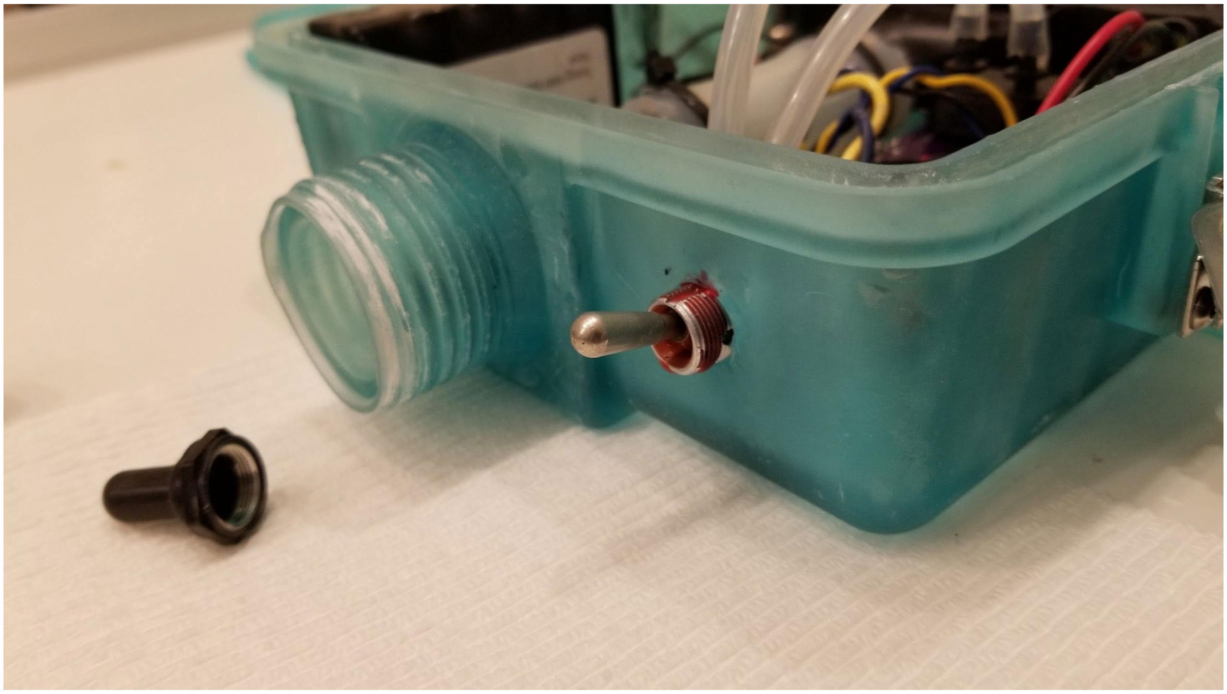

**Figure 18:** The switch with the Threadlocker applied, just before the switch cover with sealing nut is screwed on (shown on the left of the picture).

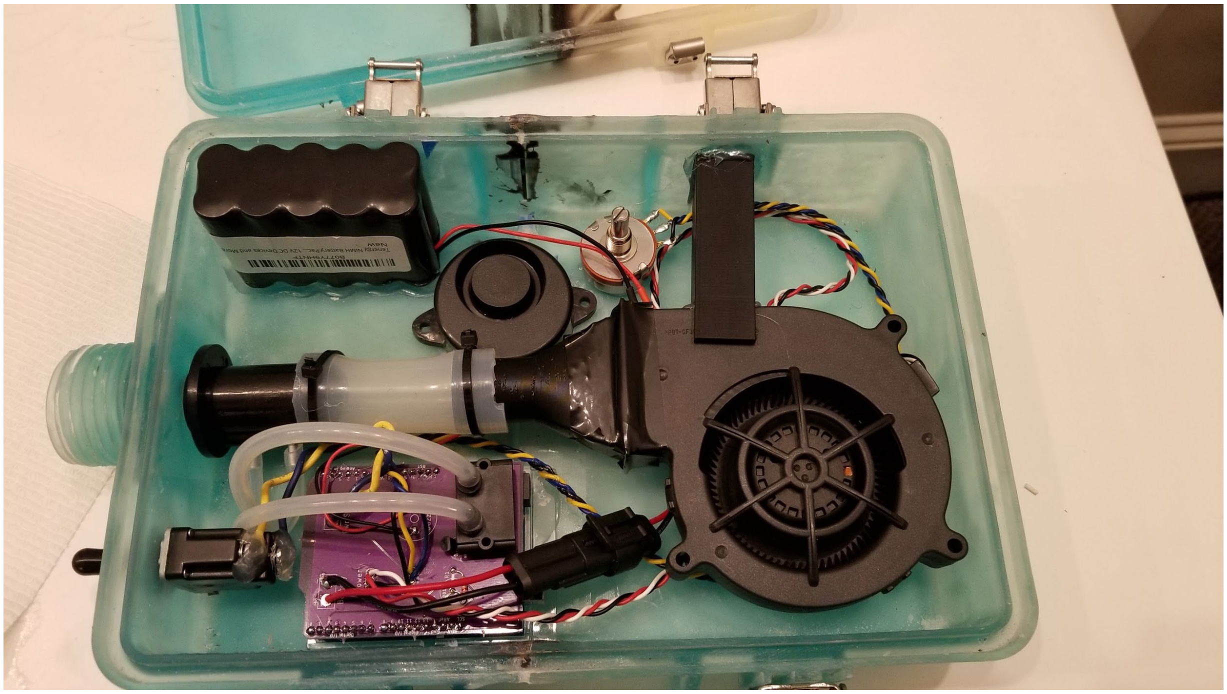

**Figure 19:** A photo of the finished connections of the prototype.

© President and Fellows of Harvard College and The Massachusetts Institute of Technology, 2020.

Programming

To complete the setup, the Arduino Uno controlling the electronics for the prototype had to be programmed with proper firmware. A standard USB cable was connected to the prototype and a computer running current Arduino software. The prototype board was enumerated as a serial port on the computer. From there - the relevant firmware, located on a shared Git-style repository, and on **Supplementary Material 4** was used.

**OPTIONAL DESIGN- ARDUINO PROTOSHIELD**

Earlier versions of this design used an Arduino Protoshield **(Figure 20)** to connect the components instead of a custom PCB. The electrical configuration of said components, however, was identical**.** Using general-purpose 22-gauge wire for the signal connections and 18-gauge wire for the NiMH battery connections and switch, the schematic described in **Figure 1** was used to electrically connect the prototype together. This Protoshield electrically connected to the Arduino Uno to provide functionality.

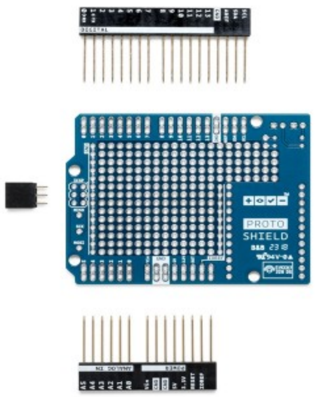

**Figure 20:** The original Arduino Protoshield used in earlier versions of the prototype.

© President and Fellows of Harvard College and The Massachusetts Institute of Technology, 2020.
