## Supplementary Material 4 for "De novo Powered Air-Purifying Respirator Design and Fabrication for Pandemic Response": Custom Design.pdf

**Bill of Materials for PanFab Custom PAPR Design**

Peter Sorger, PhD, 0000-0002-3364-1838

Following are the parts used in the PanFab Custom PAPR Design. The estimated costs assume a total production run of 2000 units.

| S. No. | Item | Qty. | Supplier/Method | Part No. | Estimated cost ($) |
|---|---|---|---|---|---|
| 1 | Housing Bin | 1 | ABS Injection Molding | N/A | 9.60 (Tooling Cost = 10,720) |
| 2 | Housing Lid | 1 | ABS Injection Molding | N/A | 9.70 (Tooling Cost = 19,589) |
| 3 | Blower, Centrifugal | 1 | Delta Electronics | BFB1012HD-04D4L | 36 |
| 4 | Battery pack, 12V NiMH | 1 | Tenergy | Amazon Standard Identification Number: B077Y9HNTF | 23 |
| 5 | Controller | 1 | Arduino | R3 | 23 |
| 6 | PCB Shield | 1 | OSH Park | N/A | 9.50 |
| 7 | Differential Pressure Sensor | 1 | Sensirion | SDP810-500PA | 19 |
| 8 | Buzzer | 1 | Mallory Sonalert Products | PS-580Q | 5 |
| 9 | Potentiometer | 1 | Bourns Inc. | 93R1A-R22-A12L | 3 |
| 10 | Transistor | 1 | ON Semiconductor | 2N3904BU | 0.20 |
| 11 | Resistor | 1 | Vishay BC Semiconductor | PR02000201001JR500 | 0.32 |
| 12 | Electrical connector | 1 | TE Connectivity AMP Connectors | 1-2834184-3 | 1.40 |
| 13 | Venturi | 1 | BPE Inc. | 178-71-2 | 0.54 |

© President and Fellows of Harvard College and The Massachusetts Institute of Technology, 2020.

| | | | | | |
|---|---|---|---|---|---|
| 14 | Venturi Ports | 2 | Car-Anth Manufacturing | 16-1204-2 | 0.68 (Tooling Cost = 375) |
| 15 | Blower silicone tube | 1 | McMaster-Carr | 3038K29 | N/A |
| 16 | Venturi silicone tube | 2 | McMaster-Carr | 3038K12 | N/A |
| 17 | Blower adapter | 1 | ABS 3D printing | N/A | 6 |
| 18 | Housing Gasket | 1 | Apple Rubber | N/A | 3.20 (Tooling Cost = 2560) |
| 19 | Housing Latches | 4 | McMaster-Carr | 1794A55 | 15.20 |
| 20 | Gaskets for threaded connections | 4 | McMaster-Carr | 5647K62 | 3 |
| 21 | Latch screws | 16 | McMaster-Carr | 98164A441 | 1.55 |
| 22 | Latch nuts | 16 | McMaster-Carr | 90730A007 | 0.65 |
| 23 | Switch | 1 | McMaster-Carr | 8002K114 | 28 |
| 24 | Switch Cover | 1 | McMaster-Carr | 70205K4 | 4.40 |
| 25 | Waist strap | 1 | Skil-Care | PathoShield Gait Belt | 11 |
| 26 | Filter | 2 | Custom Filters | N/A | 19 (Tooling Cost = 26020) |
| 27 | Hose adapter | 2 | ABS Injection molding | N/A | 6.60 (Tooling Cost = 9997) |
| 28 | Hose | 1 | Flexaust | Flex-Tube PU-IH, PN: 33800125000 | 8.50 |
| 29 | Hood coupler | 1 | ABS Injection molding | N/A | 3.30 (Tooling Cost = 7456) |
| 30 | Locking ring | 1 | ABS Injection molding | N/A | 2.70 (Tooling Cost = 5156) |
| 31 | Hood | 1 | University of Washington | VHA ADAPT PAPR Hood | 30 |
| **Total** | | | | | **284.04** |

Note: Several costs are unofficial quotes

© President and Fellows of Harvard College and The Massachusetts Institute of Technology, 2020.
