## Supplementary Material 4 for "De novo Powered Air-Purifying Respirator Design and Fabrication for Pandemic Response": Labels, warnings, legal.pdf

### Labels and Warnings

### Appropriate Use Criteria

This supplementary Powered Air Purifying Respirator (PAPR) was created as an emergency action in effort to protect people by providing backup Personal Protective Equipment (PPE) options if the standard PPE has become unavailable. This device has not gone through the same regulatory approval process as standard PPE but has gone through a special verification process expedited strictly for the response to the COVID-19 pandemic. The use of this supplementary PAPR should always come secondary to existing PPE equipment, standards, and protocol options if available. The decision to implement this device should be made with careful consideration and under the consultation of the corresponding institution's occupational health and infection control departments.

The information included in this document provides a best-effort protocol to minimize the risk of viral transmission during assembly and delivery, as well as produce PAPRs whose quality is as consistent as possible.

### LEGAL DISCLAIMER

THIS PAPR DESIGN AND SPECIFICATIONS (THE "**DESIGN**") IS PROVIDED "AS IS" AND PANFAB IS NOT RESPONSIBLE FOR ENSURING THAT ONE'S USE OF THE DESIGN WILL BE CLINICALLY SOUND, WITHOUT ERROR, OR OTHERWISE SUCCESSFUL. THE DESIGN HAS NOT BEEN TESTED OR APPROVED FOR USE IN A HEALTHCARE SETTING AND THE DESIGN IS NOT APPROVED BY ANY REGULATORY BODY. IF YOU CHOOSE TO USE THE DESIGN, YOU ASSUME ALL

© President and Fellows of Harvard College and The Massachusetts Institute of Technology, 2020."

RISK AND BEAR ALL RESPONSIBILITY AND LIABILITY, INCLUDING BUT NOT LIMITED TO, THE RESPONSIBILITY FOR ANY FEDERAL, STATE OR OTHER APPLICABLE REGULATORY REQUIREMENTS THAT APPLY TO THE MANUFACTURE, DISTRIBUTION, AND USE OF PAPRS.  PANFAB SPECIFICALLY DISCLAIMS ALL WARRANTIES, EXPRESS, IMPLIED OR STATUTORY, INCLUDING IMPLIED WARRANTIES OF MERCHANTABILITY, FITNESS FOR A PARTICULAR PURPOSE, AND NONINFRINGEMENT.

© President and Fellows of Harvard College and The Massachusetts Institute of Technology, 2020."
