## Supplementary Material 4 for "De novo Powered Air-Purifying Respirator Design and Fabrication for Pandemic Response": PanFab PAPR User Instructions.pdf

### INTRODUCTION

PanFab has developed two PAPR designs for use by healthcare workers amidst the COVID-19 pandemic. One PAPR, the PanFab Custom PAPR, uses a custom housing and filters designed and engineered for this particular project. The other PAPR, the PanFab Commercial PAPR uses a modified commercial Pelican case as the housing and commercially available Milwaukee filters. Most other parts in the two PAPR's are the same. The instructions here, while described only for the PanFab Custom PAPR, apply to the Commercial PAPR as well, unless otherwise noted. Also note that either blower unit can be used with either filter (custom filter or commercial Milwaukee filter).

### OVERVIEW

This section introduces the PAPR construction and operation.

Parts in the PAPR's

The PAPR's consist of the following parts/assemblies:

- Housing (with housing gasket and waist belt)
    - Custom housing consists of two parts- a bin and a lid
    - Custom housing has separately assembled latches)
- 2 x Filters
- 2 x Filter covers (for Milwaukee HEPA Filter)
- Blower

© President and Fellows of Harvard College and The Massachusetts Institute of Technology, 2020.

- Blower Adapter
- 1” Silicone tube (to connect blower adapter to venturi)
- Venturi (with ports)
- 7/32” Silicone tube (to connect venturi ports with differential pressure sensor)
- Switch
- Battery Pack
- Battery Connector
- Potentiometer
- Buzzer
- Microcontroller (with soldered electronic components)
- Hose (integrated with threaded hose adapters on both ends)
- 4 x Face gaskets (2 for filter inlet and 2 for hose adapters)
- Hood Coupler
- Locking Ring
- Hood
- Head Strap
- Battery charger

<u>PAPR Construction</u>

**Figure 1** below shows the custom PAPR with all the parts labelled.

© President and Fellows of Harvard College and The Massachusetts Institute of Technology, 2020.

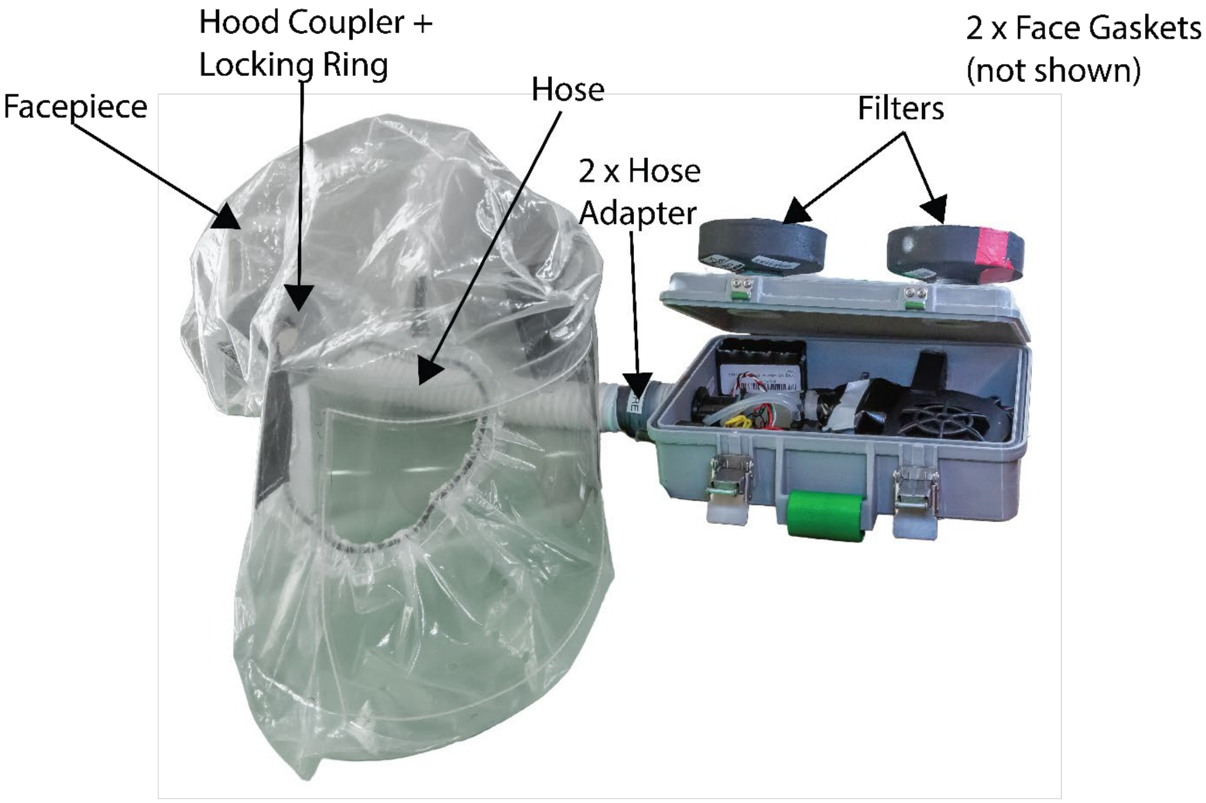


**Figure 1A:** Overview of PanFab Custom PAPR components

© President and Fellows of Harvard College and The Massachusetts Institute of Technology, 2020.

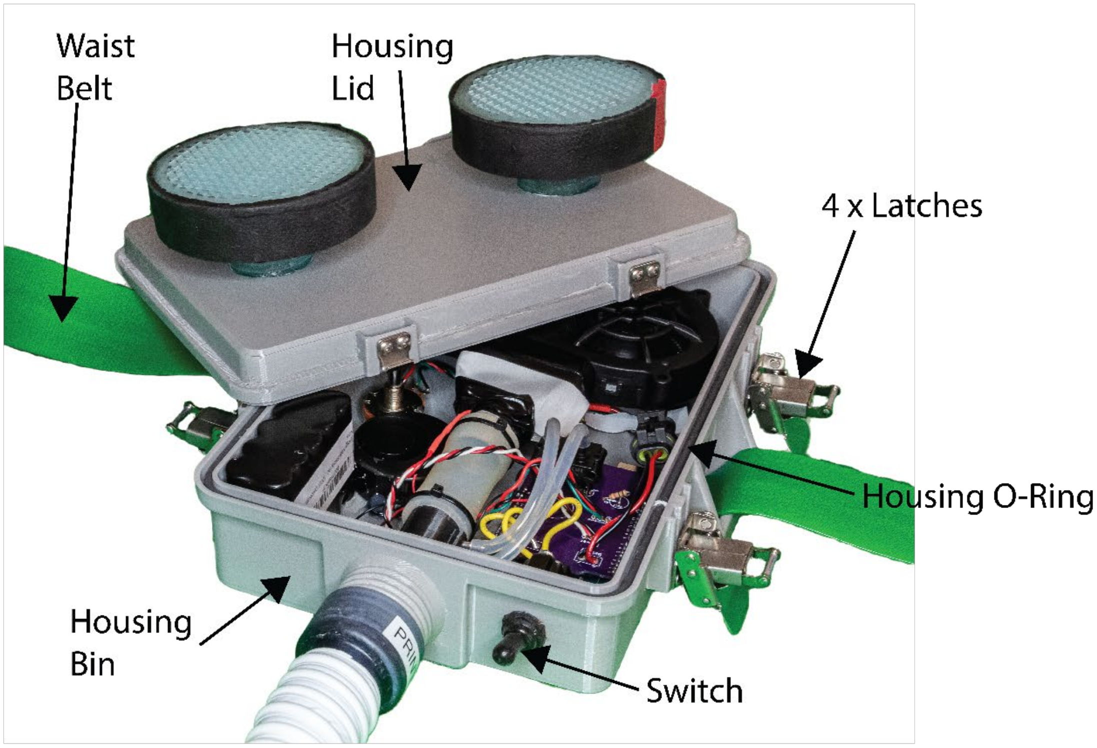


**Figure 1B:** PanFab Custom PAPR Blower Unit External Components

© President and Fellows of Harvard College and The Massachusetts Institute of Technology, 2020.

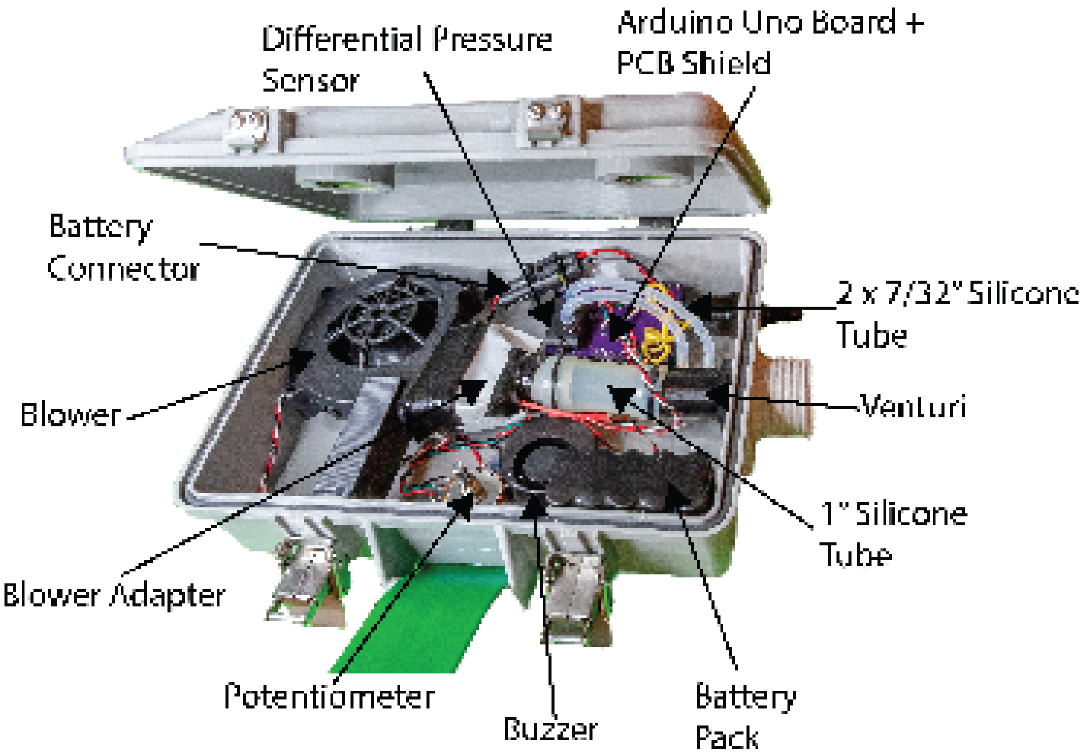


**Figure 1C:** PanFab Custom PAPR Blower Unit Internal Components

The housing “lid” and “bin” are closed together and locked using latches. A housing seal in the perimeter groove on the bin for custom housing, and on the lid for Pelican housing, seals the housings. In the case of Pelican housing, only two latches are present and the housing has a “clamshell” design. The two filters are screwed into the threaded holes on the housing lid for the custom blower unit, and on the side opposite to the lid on the Pelican housing. The face gaskets in these holes provide sealing. A waist belt assembled with the housings enables donning the PAPR.

The hose has two hose adapters permanently installed on either end- one connects to the blower unit outlet and the other connects to the hood inlet. The hose is connected to the blower unit and

© President and Fellows of Harvard College and The Massachusetts Institute of Technology, 2020.

hood (at hood coupler) by screwing the parts together. Again, face gaskets ensure sealing at these joints. The locking ring forms a tight connection between the hood coupler and the hood. The hood is worn over the head and is secured to the head using a tight fitting head strap, joined by Velcro strips.

A switch on the blower unit turns on/off the system. A microcontroller (Arduino Uno board connected to a proto-shield) forms the control system of the PAPR's. The blower, battery, potentiometer, buzzer, switch, and venturi are all connected to the microcontroller. The battery connector connects the battery to the switch. When the connector is closed and the switch is turned on, the microcontroller and blower are turned on. The blower sucks in outside air through the filters, cleaning it. The clean air is then pushed through to the hood for breathing. The air then escapes out through holes in the hood. The blower adapter allows for connecting blower outlet to venturi inlet through a 1" OD silicone tube. The silicone tube provides compliance in blower-venturi alignment. The silicone tube is joined to the venture. At the exit of venture, the air flows to the hose through blower unit outlet.

The venturi has two ports which are connected to a differential pressure sensor mounted on the microcontroller, through 7/32" silicone tubes. The differential pressure sensor is used to estimate the flow rate. When the flow rate drops below 230 LPM for 10 consecutive seconds, the buzzer goes off to alert the wearer of low flow rate. The potentiometer is used to vary the flow rate as per user comfort.

**USE PROCEDURE**

This section describes the steps involved in using the PAPR.

© President and Fellows of Harvard College and The Massachusetts Institute of Technology, 2020.

1. Open the housing by opening the latches. On the custom case, the latches are opened by pressing on the safety catches and then lifting the lower tab. On the Pelican case, the latches are opening by pressing the release button in the middle and lifting the tabs. Ensure that the housing gasket is sitting in its groove and is not falling out of it. Also ensure that the battery connector is connected.
2. Place the housing lid over the housing bin, ensuring none of the wires stick outside the blower unit, and close down the latches. See **figure 2** below.

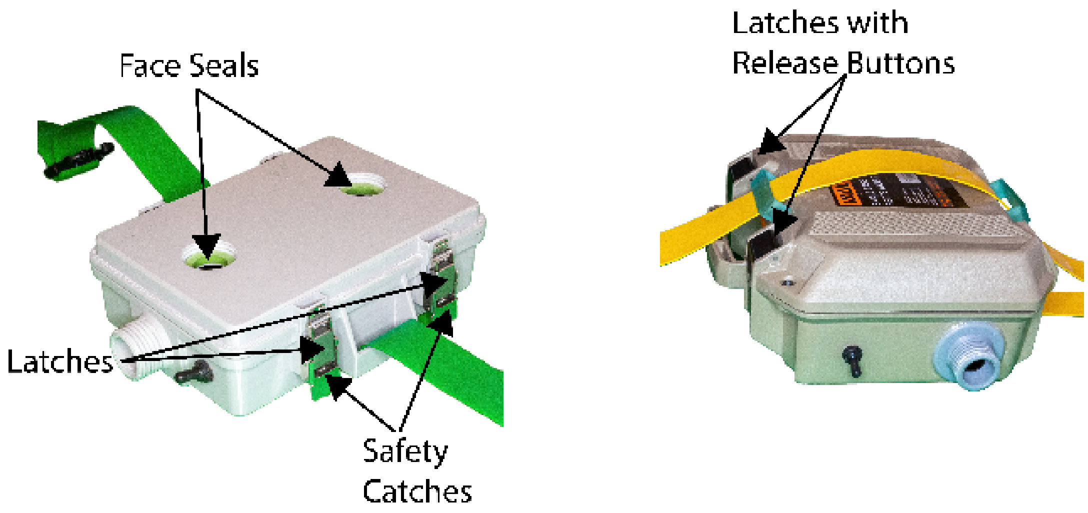


**Figure 2:** Housing Lid closure

3. Check that the face seals are present in the internal threaded holes on the housing and are not raised from their gasket seats (gasket seats are flat surfaces at the bottom of the

© President and Fellows of Harvard College and The Massachusetts Institute of Technology, 2020.

internal threads where the face seals sit). If the seals are raised, push down on them until they make uniform contact with the gasket seats.

4. Filter installation.
    a. Custom filters: Screw in the two filters onto the lid of custom housing, as shown in **Figure 3**. For Pelican housing, the filter inlet holes are on the face opposite to the lid. Hand-tighten the filters until there is significant resistance to further turning. Do not over-torque the filters.

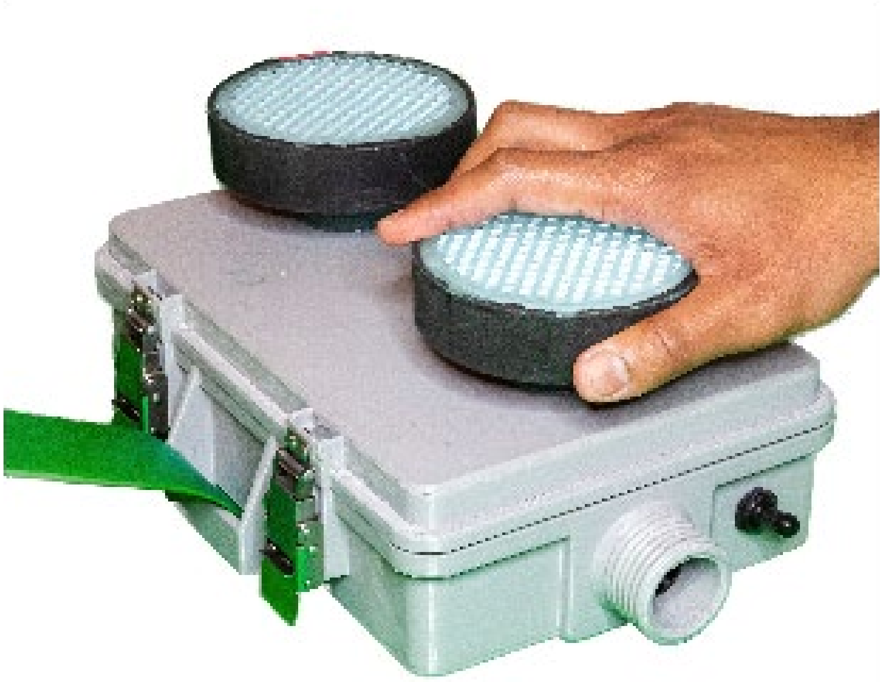

**Figure 3:** Custom Filter Installation

    b. Milwaukee filters: While installing the Milwaukee filters, always use the bottom end cap of the filter to tighten; see **Figure 4** below. Screw in the two filters into

© President and Fellows of Harvard College and The Massachusetts Institute of Technology, 2020.

the threaded holes. Hand-tighten the filters until there is significant resistance to further turning. Do not over-torque the filters.

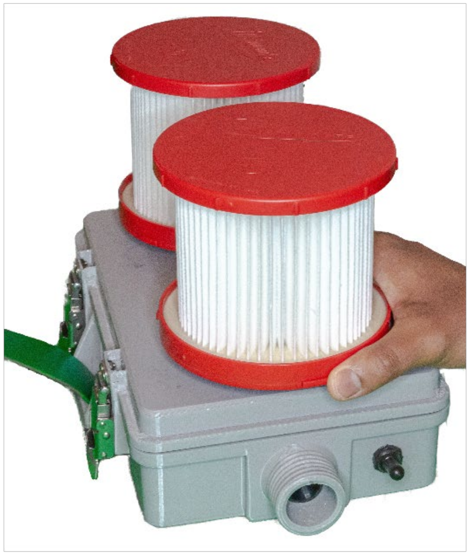

**Figure 4:** Commercial Filter Installation

Slide the two filter covers over the filters and rotate them about their axes so that the two tabs on each cover makes contact with the top surface of the housing. Use a strip of double-sided mounting tape for each cover tab and press down the cover on the housing to make a good bond. Again, ensure that the mounting tape under the tabs make good surface contact with the housing surface. Also be careful in positioning the covers over the two filters so that there is enough room for the two covers; it is advised to put on the mounting tape after sliding both covers over, to ensure compliance. **Figure 5** below shows the Custom PAPR blower unit with Milwaukee filters and filter covers installed.

© President and Fellows of Harvard College and The Massachusetts Institute of Technology, 2020.

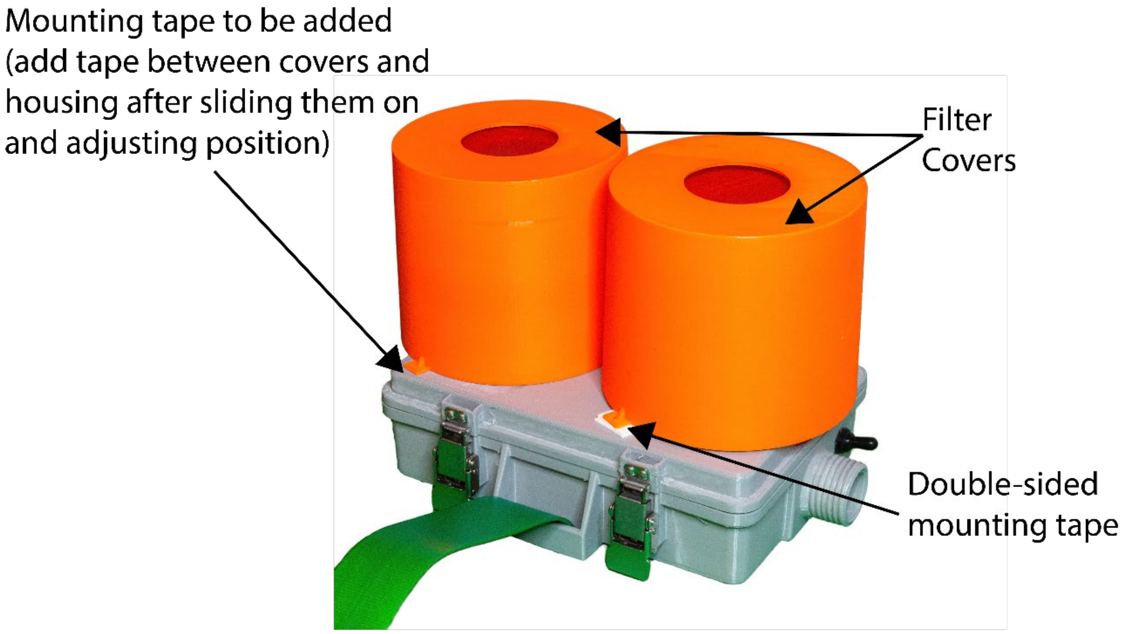


**Figure 5:** Commercial Filter Cover installation

5. Once the face seals are inspected on each hose adapter as per step 3 above, screw the hose onto the outlet of the blower unit, as shown in **Figure 6**. Hand-tighten the hose adapter until there is significant resistance to further turning. Do not over-torque the hose adapter. Always apply torque on the adapter, and not on the hose. Ensure the correct hose is used for a given PAPR blower unit.

© President and Fellows of Harvard College and The Massachusetts Institute of Technology, 2020.

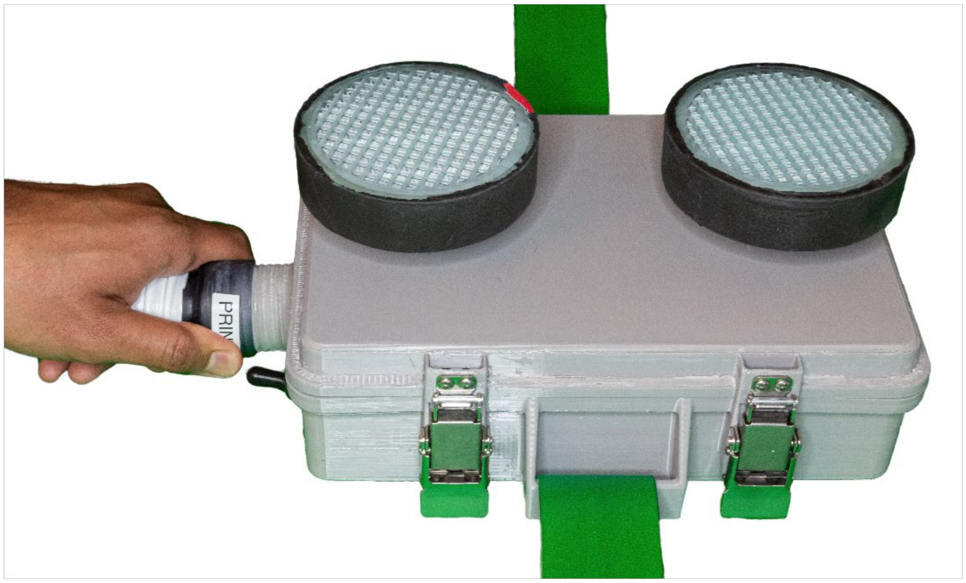

**Figure 6:** Hose connection

6. Pass the hood coupler through the circular hole at back of the hood in such a way that the threaded part faces out of the hood. Make sure the two small cylindrical tabs pass through the hole. See **Figure 7** below.

© President and Fellows of Harvard College and The Massachusetts Institute of Technology, 2020.

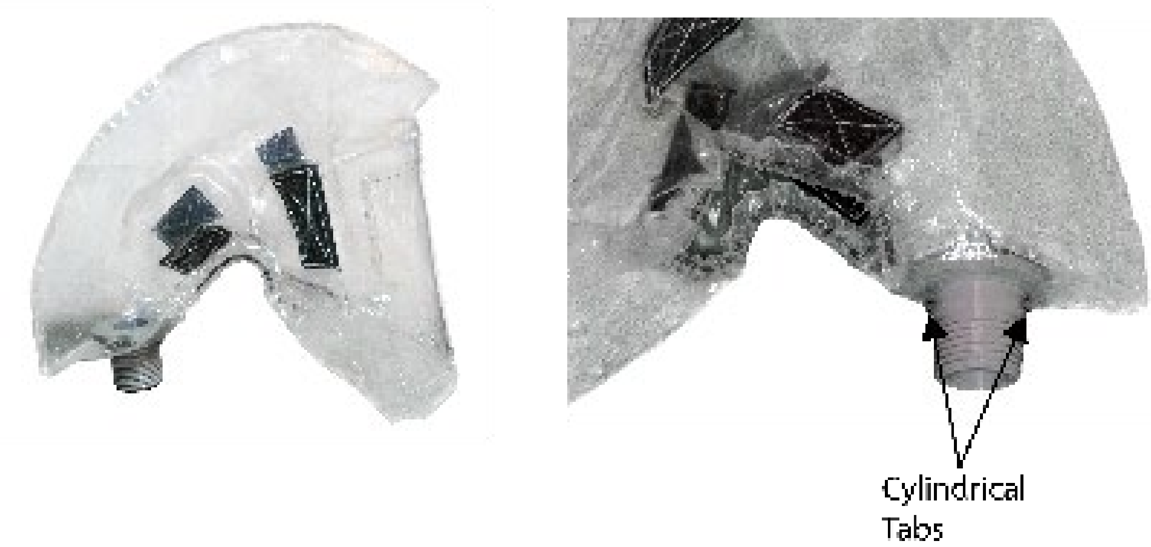


**Figure 7:** Hood Coupler and hood connection

7. Slide the locking ring over the hood coupler. The tabs on the hood coupler should slide into the slot on the locking ring. However, do not turn the locking ring to lock it. The assembly at this step will look like **Figure 8** below.

© President and Fellows of Harvard College and The Massachusetts Institute of Technology, 2020.

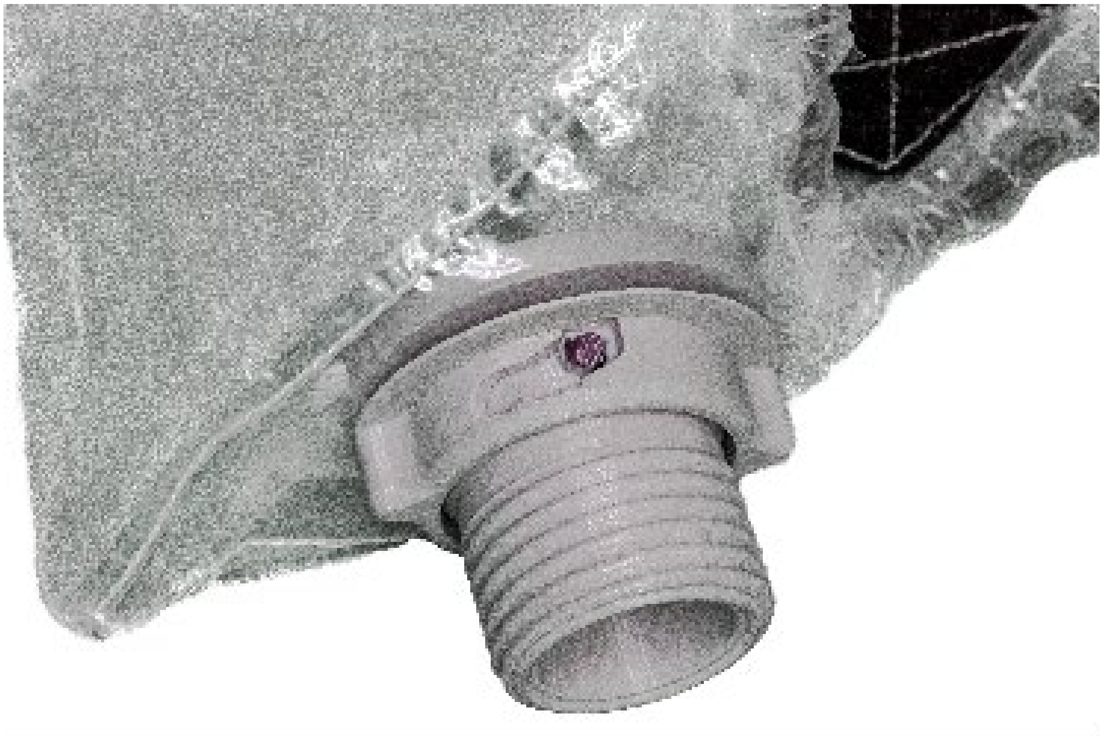

**Figure 8:** Hood Coupler and Locking Ring connection

8. With the locking ring still “unlocked”, screw the hood coupler into the other end of the hose (into the hose adapter), as shown in **Figure 9** below. Hand-tighten the hood coupler until there is significant resistance to further turning. Do not over-torque the hood coupler. Do not apply torque on the hose.

© President and Fellows of Harvard College and The Massachusetts Institute of Technology, 2020.

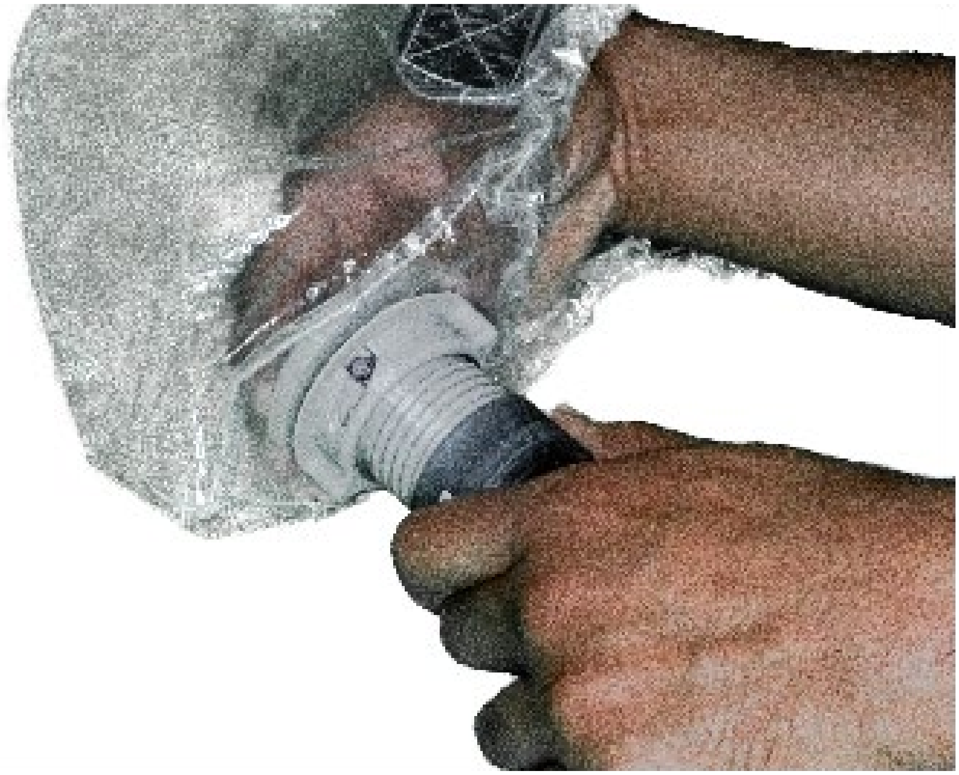

**Figure 9:** Hood Coupler and Hose Adapter Connection

9. Orient the hood such that the “face” of the hood faces away from the filter inlet. i.e. the blower unit is worn on lower back such that the filters face away from the body backward and the hood faces forward.
10. Once the hood is oriented as per step 9, lock the locking ring onto the hood coupler by turning it clockwise. Ensure to lock it only when the cylindrical tabs on the hood coupler are out of the hood, as per step 6. The hood coupler with locking ring locked will be as shown in **Figure 10**.

© President and Fellows of Harvard College and The Massachusetts Institute of Technology, 2020.

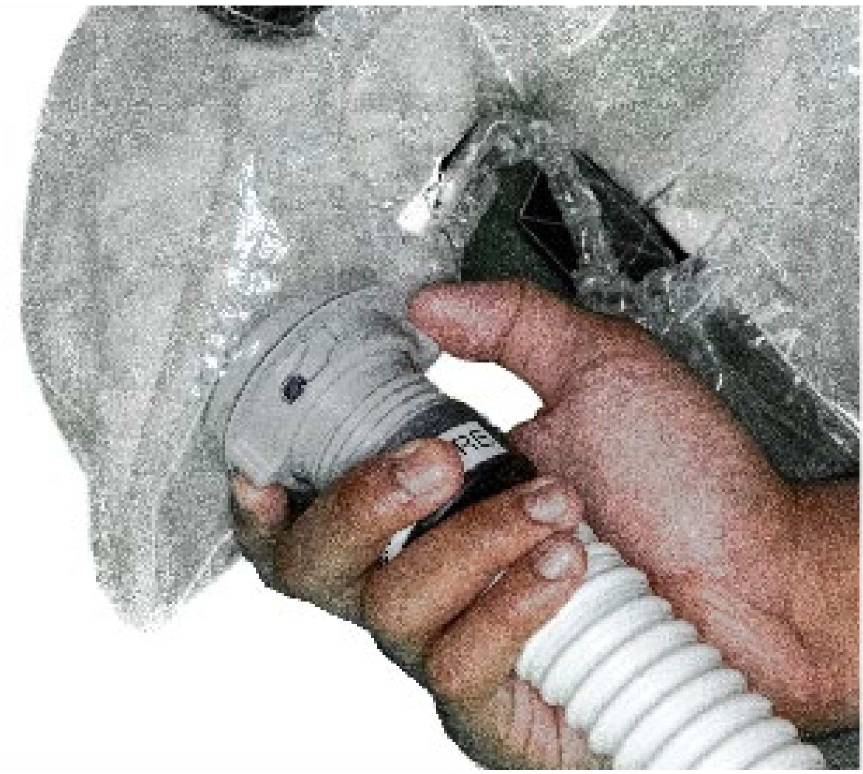

**Figure 10:** Locking Ring locked on Hood Coupler

11. To don the PAPR, first set the blower unit on a waist-high table, with filters facing down. Then, carefully lift the blower unit using the belt straps towards lower back. Close the buckle of the belt and adjust the tightness as required. Tuck in the excess strap length into the tightened strap. **Figure 11** shows the PAPR donning.

© President and Fellows of Harvard College and The Massachusetts Institute of Technology, 2020.

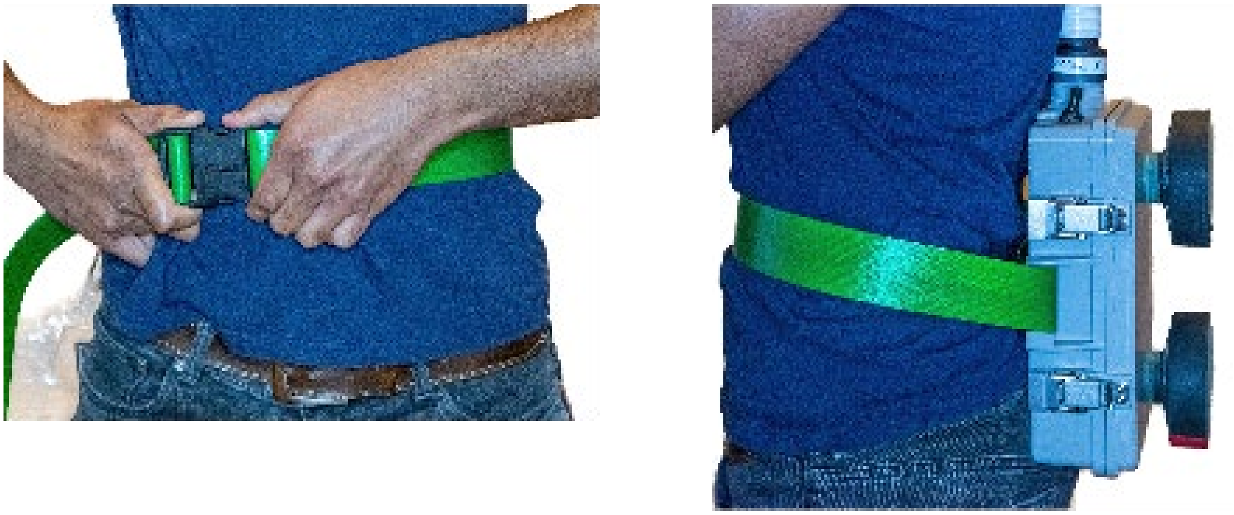

**Figure 11:** PAPR Blower Unit Donning

12. Put on the head strap. There are 7 velcro strips on the head strap in total. 3 of them are used in tightening the head strap on the head, while the other 4 connect to corresponding strips in the hood. The tightening (T) strips and the strips connecting to hood are labelled as shown in **Figure 12** below. The 4 strips connecting to hood are labelled as the position on the head they are located at.

© President and Fellows of Harvard College and The Massachusetts Institute of Technology, 2020.

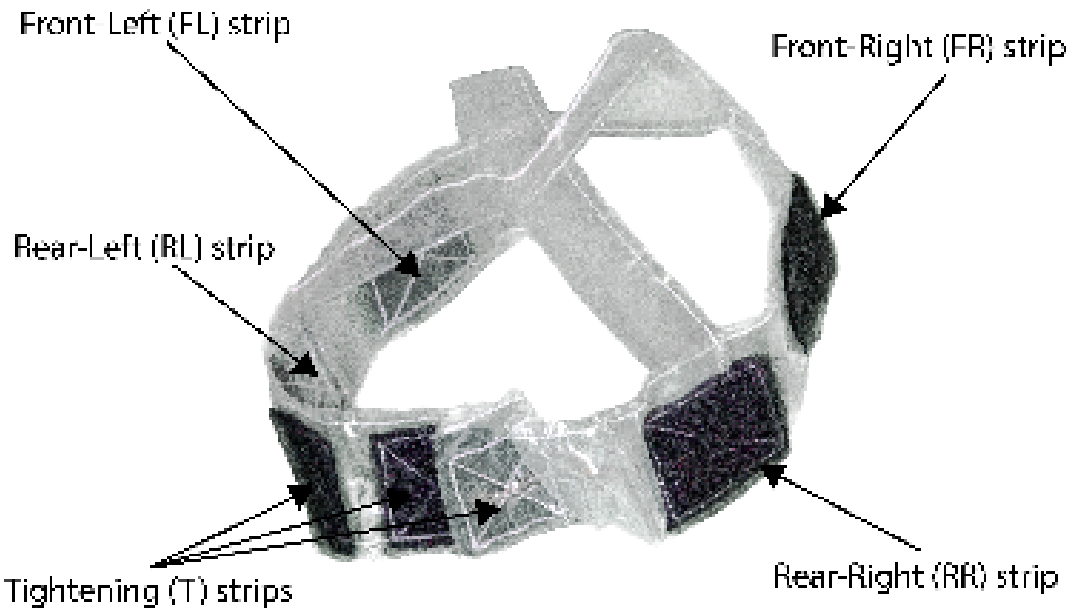


**Figure 12:** Velcro strips on head strap

Ensure the Velcro strips are facing outward, as shown in the **Figure 12** above. Un-couple the tightening strips and place the head strap on the head, with two hands on the rear part of the head strap (the rear part has the 3 T strips). Couple the T strips after having tightened the head strap around the head to required tightness, as shown in **Figure 13**. The coupled T-strips should be at the rear of the head, as per figure below. The tightness should be enough so that the hood and the head strap don't fall off.

© President and Fellows of Harvard College and The Massachusetts Institute of Technology, 2020.

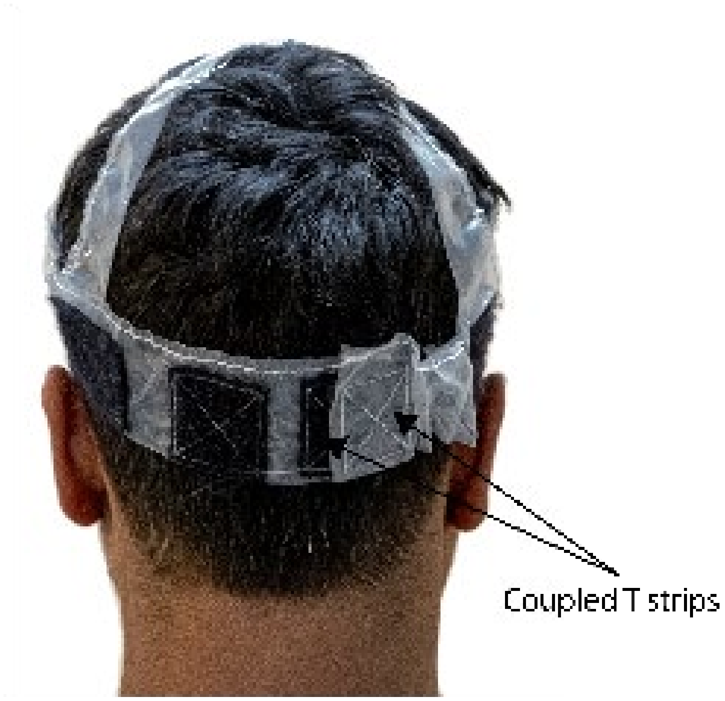


**Figure 13:** Head strap donning

13. Put on the hood. There are 4 velcro strips on the inside of the hood that correspond to FR, FL, RR, RL strips on the head strap. The 4 strips on the hood are labelled in **Figure 14** below.

© President and Fellows of Harvard College and The Massachusetts Institute of Technology, 2020.

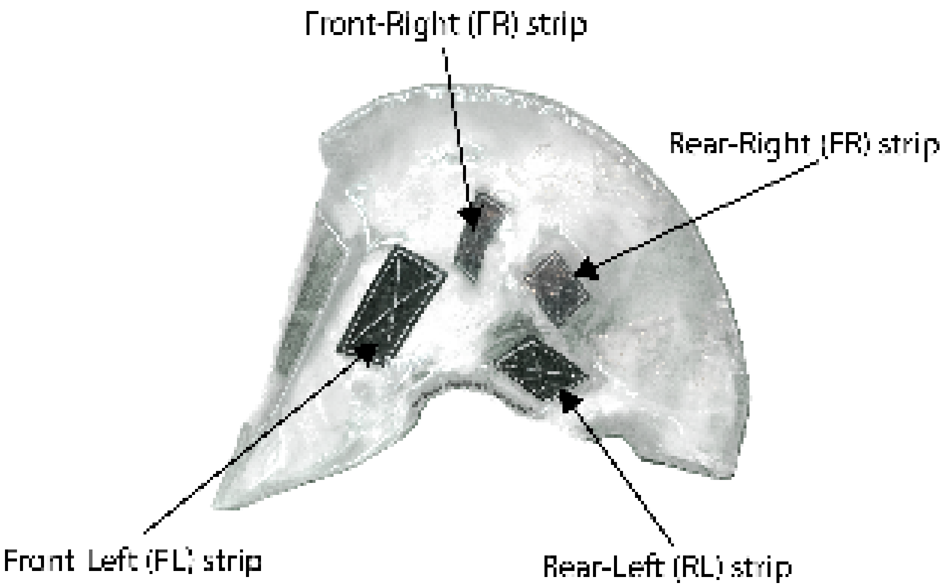


**Figure 14:** Velcro strips on the hood

With two hands, locate RR and RL strips on the hood and connect them to corresponding strips on the head strap. Press down tightly on the Velcro to ensure the connection is secure. Then, locate FR and FL strips on the hood with two hands and connect them to corresponding strips on the head strap. Once the strips are securely connected, pull down the hood so that the lower part of the elastic cord (in the hood) rests below the lower jaw and the part of the elastic cord just below the Velcro strips rests above the ears. Ensure tight fit everywhere and adjust for comfort. After donning the hood, the system should look on the wearer as per **Figure 15** below.

© President and Fellows of Harvard College and The Massachusetts Institute of Technology, 2020.

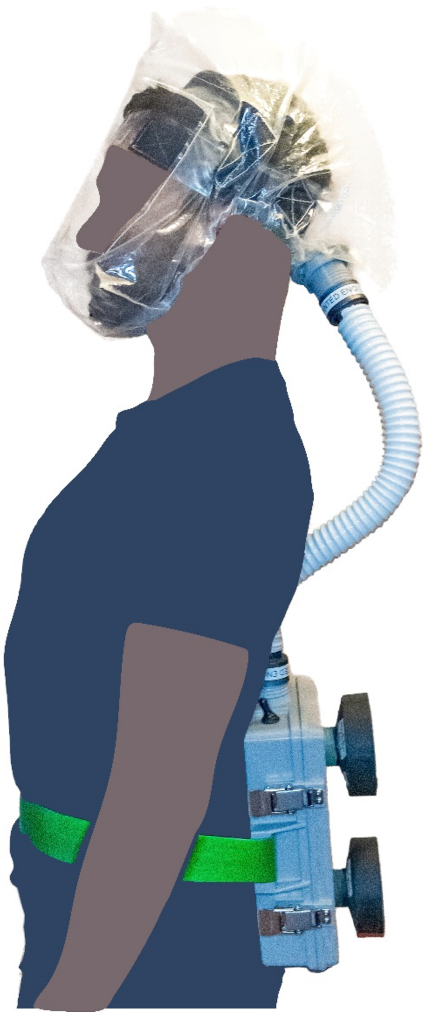

**Figure 15:** Fully donned PAPR

14. To turn on the PAPR, reach behind and turn on the switch, as shown in **Figure 16**.

© President and Fellows of Harvard College and The Massachusetts Institute of Technology, 2020.

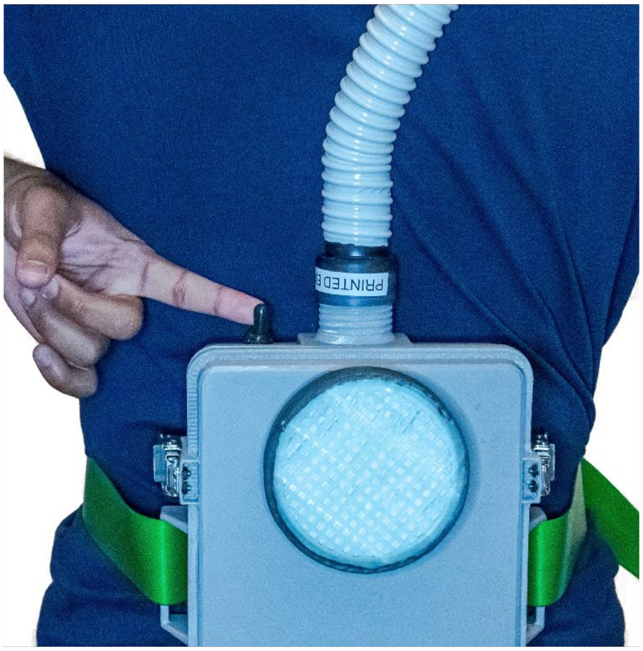

**Figure 16:** Turning on/off the PAPR

15. Doffing the PAPR is carried out by reversing the steps above.
16. The potentiometer knob (see **Figure 1**) can be turned clockwise to increase flow rate or counter-clockwise to reduce the flow rate. Adjust the flow rate to a comfortable level. Note: too little of a flow rate will make the buzzer go off.
17. Always wait 10 seconds after turning on the PAPR to go into a hazardous environment. If the buzzer goes off after 10 seconds of turning on the PAPR, either the flow rate needs to be increased, the filter needs to be replaced (due to excessive clogging), or the battery needs to be recharged. Note that the operational time on full charge, with maximum flow rate (potentiometer turned all the way), is nearly 4 hours, after which the battery would need to be recharged.

© President and Fellows of Harvard College and The Massachusetts Institute of Technology, 2020.

18. The battery is recharged using a charger provided. The battery can be charged either at 0.9A or 1.8A. Higher charging current will charge the battery faster, but may cause heating in the battery. It is advised to charge the battery at 0.9A.

    To charge the battery, open the lid as per step 1. Then, disconnect the battery to the switch by de-coupling the battery connector. Be careful so as to not knock over other parts during this disconnect process. Once disconnected, couple the half of the connecter connected to the battery, to the mating connector half on the charger. Set the charging voltage to either 0.9A or 1.8A. Plug in the other end of the charger to a power outlet. When charging, the red LED on the charger will turn on. When the battery is fully charged, the green LED on the charger will turn on. When charging is complete (which takes a little less than 3 hours at 0.9A), disconnect the connector to the battery and reconnect the battery to the switch. See **Figure 17**.

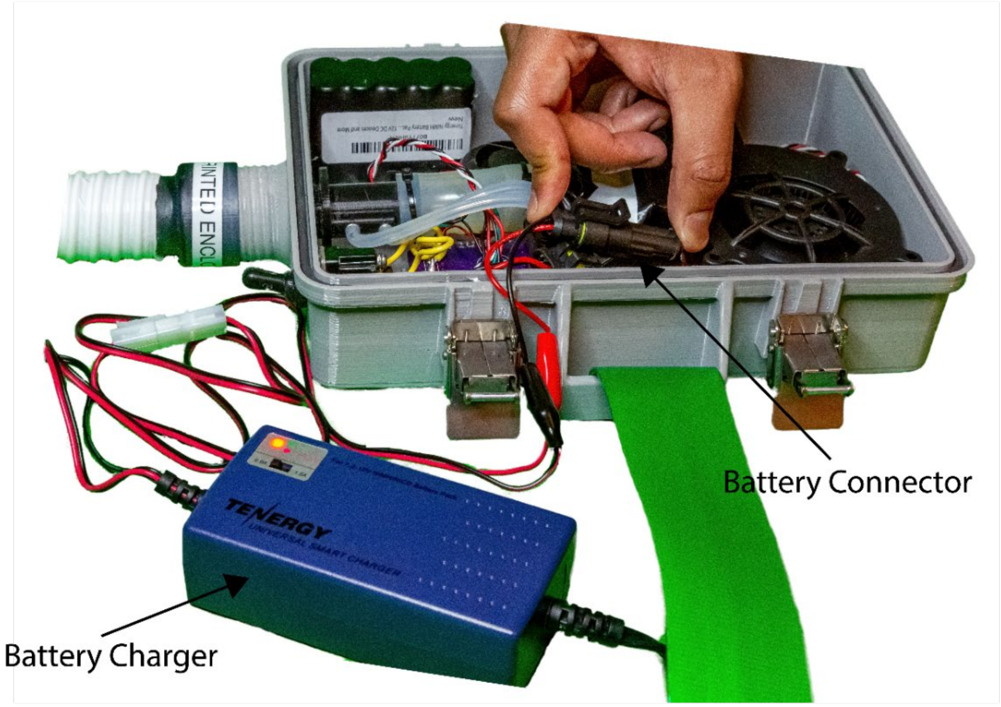


**Figure 17:** Connection of battery pack to battery charger

© President and Fellows of Harvard College and The Massachusetts Institute of Technology, 2020.
