## Supplementary figures and images for "De novo Powered Air-Purifying Respirator Design and Fabrication for Pandemic Response"

### Shield_Schematic.pdf

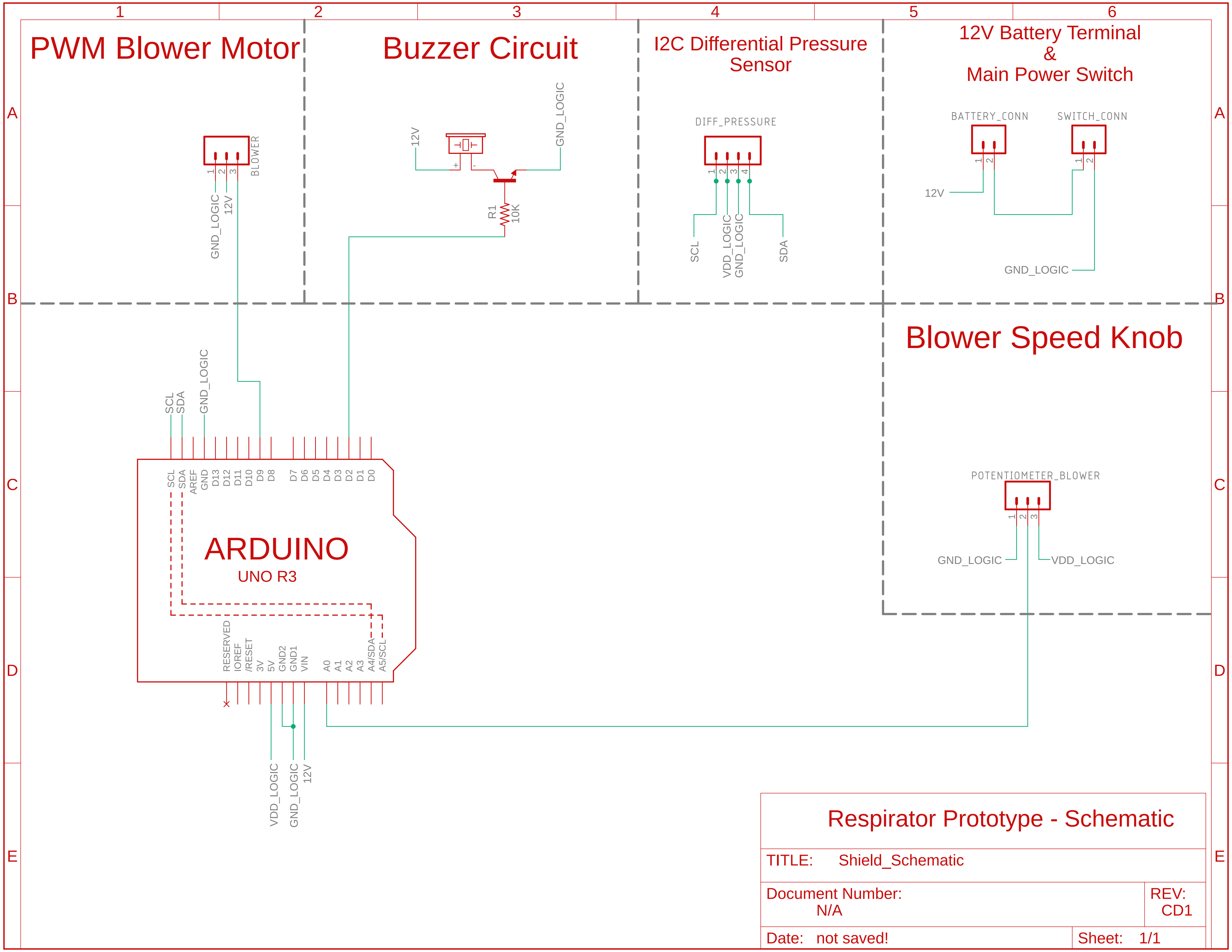


12/2/20 3:22 PM /Users/genone2/Documents/EAGLE/projects/Panfab_PAPR/Shield_Schematic.sch (Sheet: 1/1)

### Shield_Schematic_NoInfo.pdf

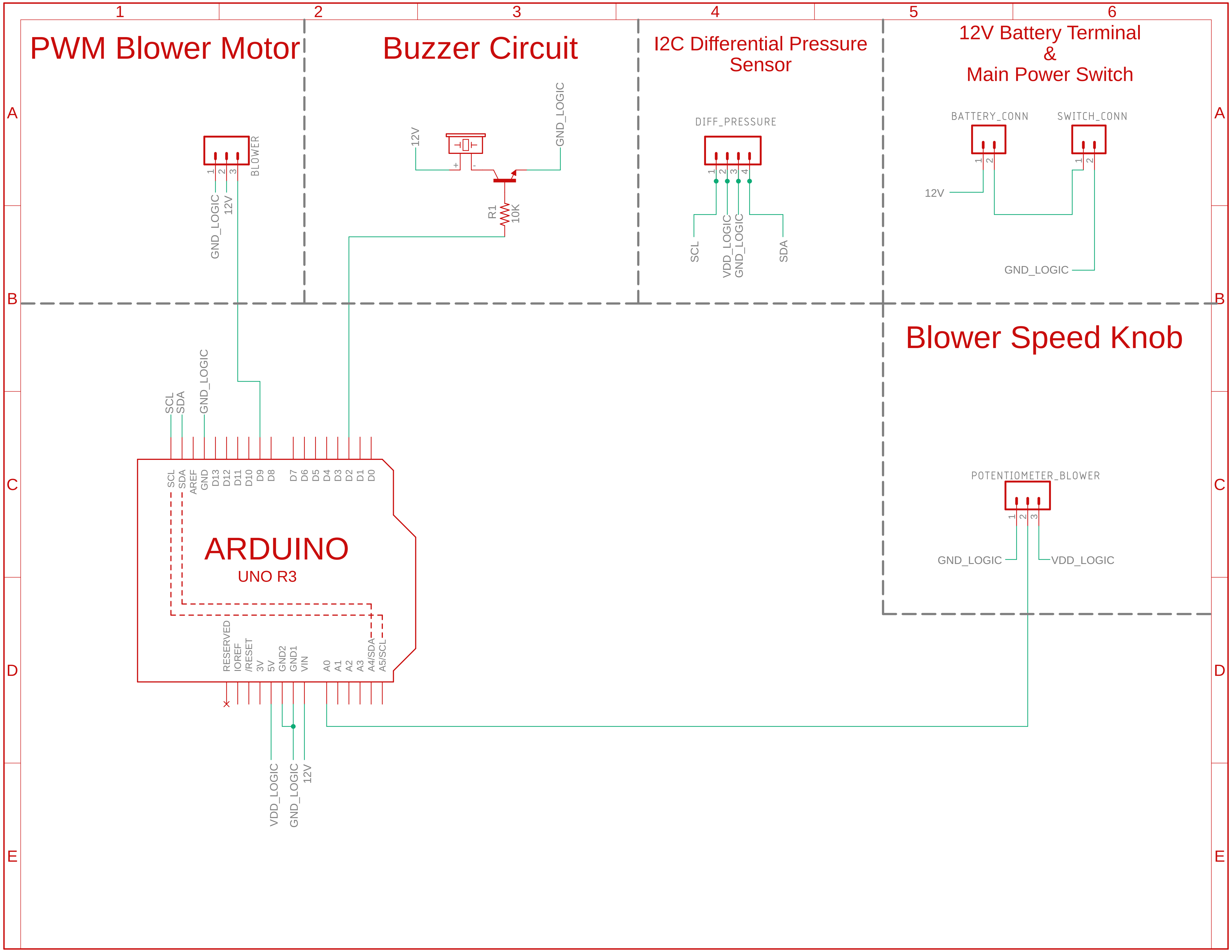


12/2/20 3:26 PM /Users/genone2/Documents/EAGLE/projects/Panfab_PAPR/Shield_Schematic.sch (Sheet: 1/1)
